# Sexual orientation inequalities in self-harm and suicidality in England and Wales – A national population-based study

**DOI:** 10.64898/2026.02.12.26346182

**Authors:** Hannah Bunk, Daniel Ayoubkhani, Vahé Nafilyan, Laia Becares, Vasa Curcin, Amal R. Khanolkar, Emma Sharland

## Abstract

**Background:** Sexual minority (SM) individuals have worse mental health than heterosexual peers. However, there is no total population-based and national-level evidence on differences in risk of self-harm and suicide by sexual orientation. This study provides the first national population-based estimates in England and Wales.

**Methods:** Using 2021 Census data linked with hospital records and death registrations, we analysed sexual orientation (SO) differences in: (i) at least one hospital inpatient admission/emergency attendance for intentional self-harm, and (ii) death by suicide. We calculated age-standardised rates per 100,000 people by SO between March 2021 and December 2023, and stratified by sociodemographic, geographical, socioeconomic and health-related variables. We calculated rate ratios for lesbian/gay/bisexual/other SO (LGB+) groups compared with heterosexuals to estimate sexual identity disparities.

**Findings:** Our study population included 28.7 million people (mean age 48.1 years, 53.7% female, 84.2% White) aged ≥16 years who self-reported their SO in Census 2021 and linked to an NHS number. LGB+ individuals had 2.52 (95% CI 2.48-2.56) times higher risk for self-harm and 2.17 (95% CI 1.98-2.37) times higher risk for suicide than heterosexual people. Relative risk of self-harm was highest for LGB+ females, younger adults, and Black individuals. Relative risk of suicide was highest for LGB+ females, older adults, and Black individuals.

**Interpretation:** This study demonstrates stark inequalities in risk of self-harm and suicide by sexual orientation, consistent across multiple sociodemographic factors. These findings are important for informing government prevention programs and further mental health research.

**Funding:** There was no external funding for this study.

**Research in context:** *Evidence before the study:* The substantial evidence on higher risk for self-harm and suicide in sexual minority groups in the UK (and wider Europe) is impacted by regional samples, younger populations, or surveys limited by smaller numbers precluding analyses by key sociodemographic factors (like sex, ethnic group, socioeconomic indicators, faith, housing situations and geographical indicators) or combining all sexual minority groups together. To date, no study has used total population-based data to examine sexual orientation inequalities in self-harm and suicide and in relation to a range of sociodemographic factors.

*Added value of this study:* To our knowledge, this is the first study in the UK to provide national population-based estimates of intentional self-harm and suicide by sexual orientation, including intersectional analyses across age, sex, ethnic group, and socioeconomic position. This study used a unique linkage between the census, hospital inpatient data, emergency care records and death registrations from across England and Wales, with a study population of 28.7 million people aged ≥16 years who self-reported their sexual orientation in Census 2021 and linked to a National Health Service (NHS) number.

*Implications of all the available evidence:* This research provides national population-level evidence of substantial increased risk for self-harm and suicide among sexual minority individuals, compared with heterosexual individuals. This study also identified key groups of individuals at an increased risk of self-harm and suicide. These findings are important for informing government prevention programs and further research supporting the mental health of sexual minority groups.

## Introduction

Evidence suggests that people who self-identify as lesbian, gay, bisexual and other sexual minority identities (LGB+) have significantly higher risk for self-harm and suicidality compared with heterosexual peers^1–5^. This has led to the World Health Organisation (WHO) identifying LGB+ populations as being at substantial risk of suicide^6^. However, evidence is limited and mostly sourced from birth cohorts and selected population-based surveys^7–15^. Whilst these studies highlight inequalities experienced by sexual minority individuals, they often focus on subgroups of the population, for example adolescents and young adults, or have smaller sample sizes. This limits potential subgroup analyses and omits key population groups, such as older individuals and those with intersecting identities.

The study of suicidal death in minority groups requires larger numbers and linkage with national data. Routine self-harm and mortality statistics are not published by sexual or ethnic identity, precluding our understanding of these acute outcomes across minoritised groups. This is despite greater number of individuals identifying with a sexual minority identity (SMI) in recent years (4.4% of 16- to 24-year-olds identified as lesbian, gay or bisexual in 2018, increasing to 10.4% in 2023^16^) and there is overwhelming evidence of poorer mental health in these groups^5,7–13^.

There is increasing interest in self-harm and suicidality in multiple minoritised individuals, i.e., individuals with SMI who also identify with ethnic and/or faith minority identities. These individuals may be differentially impacted by potential adverse forms of discrimination and stigma associated with individual and multiple intersecting identities, which can compound their risk for self-harm and suicide^17,18^. However, smaller UK studies are limited in their ability to disaggregate to this level of detail. To date, there is a paucity of national population-based estimates for self-harm and suicide by sexual orientation (SO) in England and Wales.

The 2021 Census for England and Wales included a voluntary question on sexual orientation for the first time, asked to all respondents aged ≥16 years. Of the 92.5% of people who answered the SO question, 96.6% of people identified as straight or heterosexual and 3.4% of people identified with an LGB+ orientation^19^. Using a unique linkage between the census, hospital inpatient data, emergency care records and death registrations from across England and Wales, this is the first study in the UK (and wider Europe) to examine differences in self-harm and suicide by SO using total population-based data. Further, we examined whether SO differences in self-harm and suicide varied by sociodemographic, geographical, socioeconomic and health-related variables.

## Methods

### Data sources and study population

We conducted a retrospective cohort study of respondents to the 2021 Census of England and Wales linked to death registrations (for deaths registered between 1 January 2009 and 10 February 2025) and administrative health records including: Hospital Episode Statistics (HES) Admitted Patient Care (APC) from the National Health Service (NHS) England, Emergency Care Data Set (ECDS) from NHS England, Patient Episode Database for Wales (PEDW) Admitted Patient Care (APC) from Digital Health and Care Wales (DHCW) and Emergency Department Data Set (EDDS) from DHCW (Supplementary Figure 1). The Census 2021 data were linked to NHS numbers via the NHS Personal Demographics Service (PDS) 2019, with a linkage rate of 94.6% (Supplementary Table 1).

The study population included all people (N=28,659,465 individuals) aged ≥16 years who were enumerated in Census 2021, were usual residents living in England and Wales at the time of the Census, self-reported their sexual orientation (i.e., proxy replies were excluded), and linked to the PDS. We weighted the study population for PDS linkage bias using post-stratification by sexual orientation, age group, sex, region and ethnic group^20^.

Individuals were followed from 21 March 2021 (Census Day) until either the end of the study period (31 December 2023) or death, whichever occurred first.

### Exposure variables

The main exposure of interest was SO ascertained by the question: ‘Which of the following best describes your sexual orientation?’. Respondents could choose from ‘straight or heterosexual’ (hereafter heterosexual), ‘gay or lesbian’, ‘bisexual’ and ‘other sexual orientation’ (LGB+). For the analysis we created two exposure variables: (i) a binary indicator comparing heterosexual and LGB+, and (ii) a categorical indicator distinguishing the three LGB+ subgroups (heterosexual vs. gay or lesbian, bisexual, or other).

### Outcome measures

We examined two binary outcomes: intentional self-harm and death by suicide. Intentional self-harm was defined as any hospital inpatient admission or emergency attendance during the study period. To define intentional self-harm, within the HES and PEDW datasets we used International Classification of Diseases, Tenth Revision (ICD-10) codes for intentional self-harm (X60-X84), for ECDS we used the chief complaint code 248062006 “Self-injurious behaviour (finding)” and injury intent code 276853009 “Self-inflicted injury (disorder)”, and for the EDDS dataset we used attendance group 13 for “Deliberate Self-Harm”.

For death by suicide, we identified all deaths occurring during the study period and registered by 10 February 2025. The additional one year of registrations was to mitigate the impact of registration delays. Suicide was defined using ICD-10 codes corresponding to intentional self-harm (X60-X84) and injury/poisoning of undetermined intent (Y10-Y34) (see Supplementary Tables 2 and 3)^21^.

### Covariates

This included a range of demographic, socio-economic, geographical and health-related characteristics available from the 2021 Census: sex, age group, ethnic group, region/country of residence, rural/urban classification, country of birth, English language proficiency, religion, self-reported general health, self-reported disability status, highest level of educational attainment, National Statistics Socio-Economic Status (NS-SEC), area-level socioeconomic indicators (Index of Multiple Deprivation (IMD) and Welsh Index of Multiple Deprivation (WIMD)), economic activity, household tenure, household size, and residence type (complete description in Supplementary Table 4). IMD/WIMD was categorised into deciles, with deciles 1 and 10 comprising people living in the 10% most deprived and least deprived areas, respectively. For breakdowns by highest level of educational attainment, we restricted the study population to those aged ≥25 years because education level is generally more stable in this age group.

### Statistical analyses

Descriptive statistics, including frequencies of the study population characteristics, were calculated and stratified across all covariates and for both outcomes. We estimated age-standardised rates per 100,000 people, standardised to the age distribution of the combined self-identified LGB+ group in Census 2021 using direct standardisation^22^. Next, we calculated ratios of age-standardised rates (rate ratios, RRs), to provide estimates of relative increased risk of self-harm or suicide for the LGB+ groups compared with their heterosexual counterparts. 95% confidence intervals (CIs) for the RRs were estimated by simulation. For each of 100,000 iterations, an age-standardised rate for each group was drawn from a normal distribution, with mean equal to the actual age-standardised rate and standard deviation equal to the standard error of the age-standardised rate. Following this, 100,000 RRs were calculated and the 2.5th and 97.5th percentiles defined the lower and upper limits of the 95% CI, respectively.

All statistics were estimated for both the combined LGB+ and LGB+ subgroups of the SO variables where possible, or only the combined LGB+ group for breakdowns with small sample sizes (N < 10 or self-harm/suicide events < 3). All calculations were based on estimates rounded to the nearest multiple of five for disclosure control reasons.

### Sensitivity analyses

We conducted several sensitivity analyses. First, we re-ran the analyses including those who responded to the 2021 Census by proxy (increasing the study population to N=41,198,320); there were higher proportions of proxy responses in some sociodemographic groups, for example those in bad health and those with lower-level qualifications, which may have increased bias in these variables when excluding proxy responses from the main analysis.

Second, we restricted the analyses to people living in private households only at the time of Census 2021 (N=28,339,665); individuals living in communal establishments included those living in hospitals, care homes, prisons and defence bases^23^, whose mental health may systematically differ from those in private households.

Finally, we included inpatient admissions from HES and PEDW for self-harm with undetermined intent within our outcome measure for self-harm (ICD-10 codes Y10-Y34; N=136,370 individuals had at least one recording for self-harm); intent may not always have been captured in the administrative hospital records. Emergency attendances in ECDS and EDDS without a record of intent were not included in the sensitivity analysis because of the high level of missing data on intent in these datasets (17.9% of ECDS records and 36.4% of EDDS records).

### Role of the funding source

There was no external funding for this study.

## Results

### Descriptive statistics

The study population included 28,659,465 individuals (59.0% of all people aged 16 years and over in Census 2021), including 1,105,325 (3.9%) who identified as LGB+ (Table 1). The mean age of the cohort was 48.1 years, 53.7% were female, and 84.2% were White. Figure 1 shows how the final study population was defined. During the study period, 135,035 individuals (0.5%) had at least one recording for intentional self-harm and 7,150 individuals (0.02%) died by suicide.

**Figure 1:**
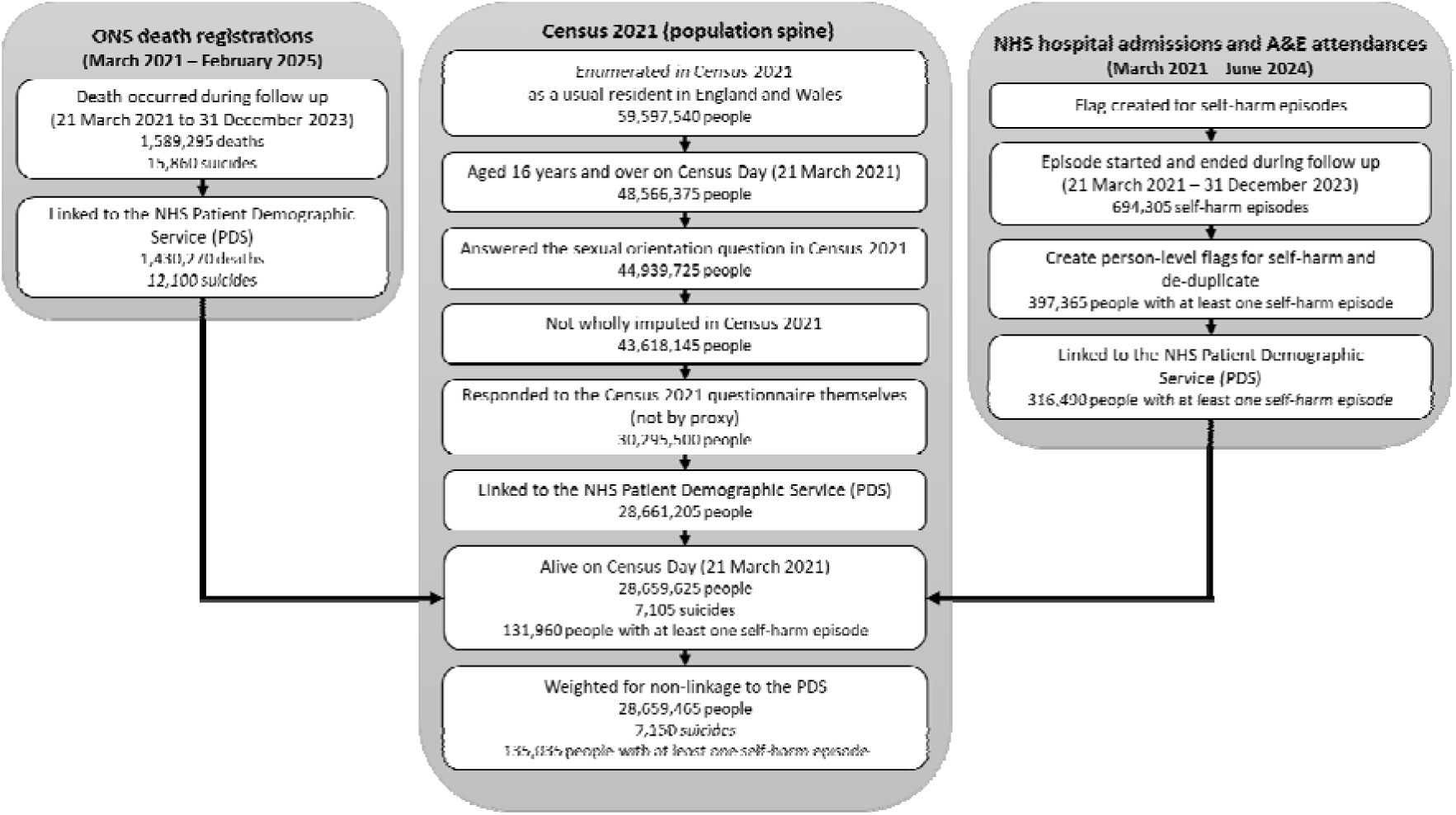
Study population flow

**Table 1:**
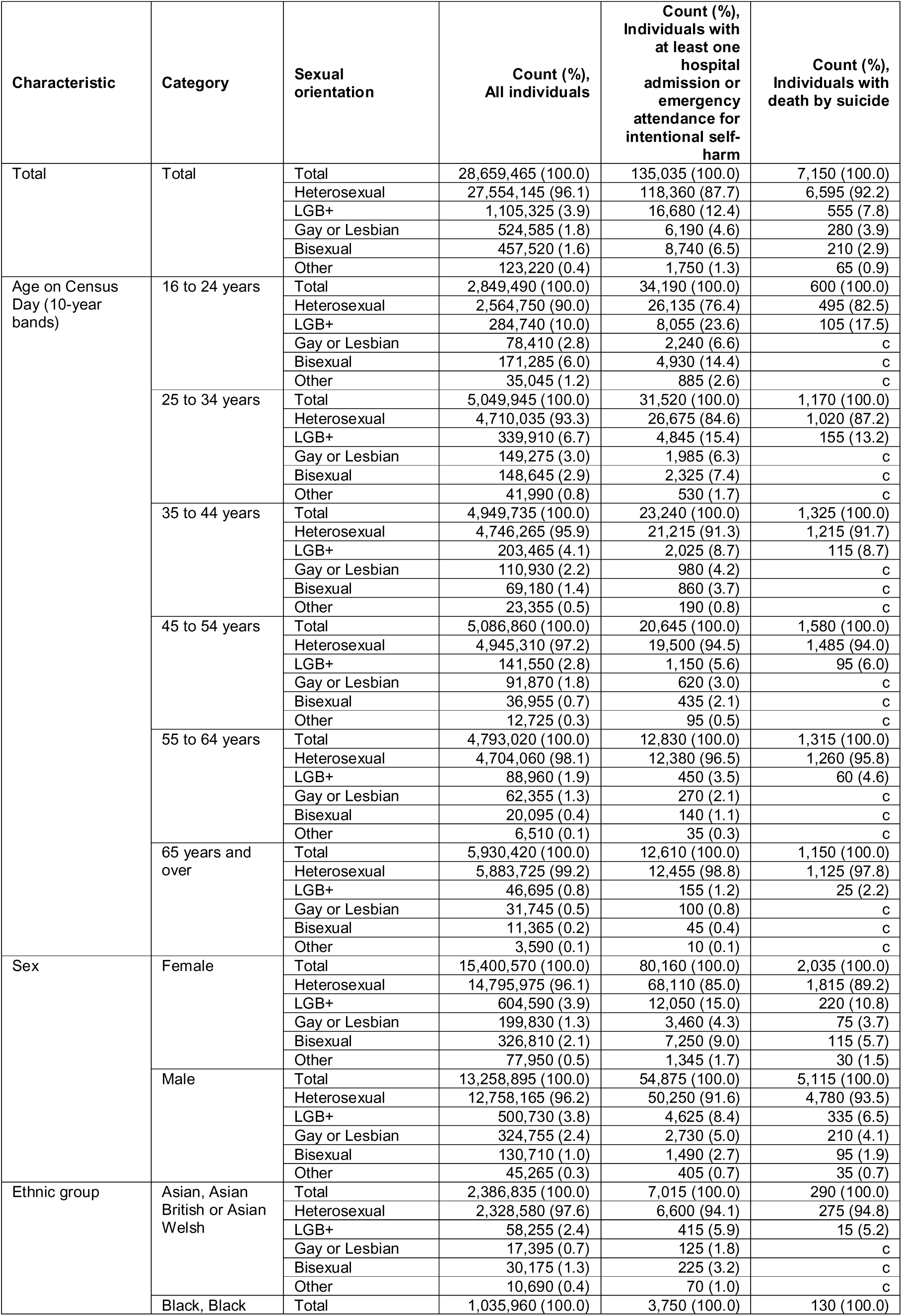
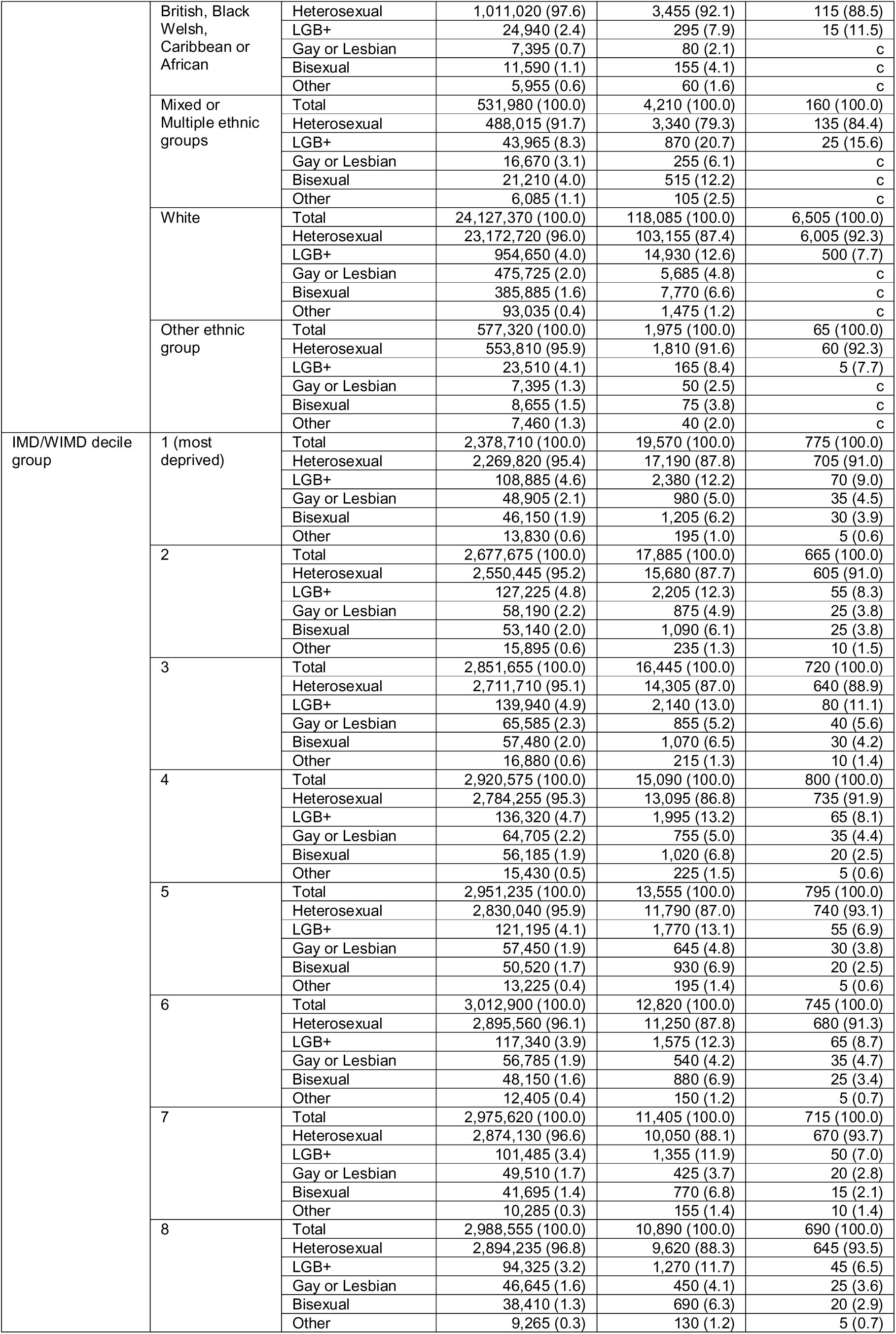
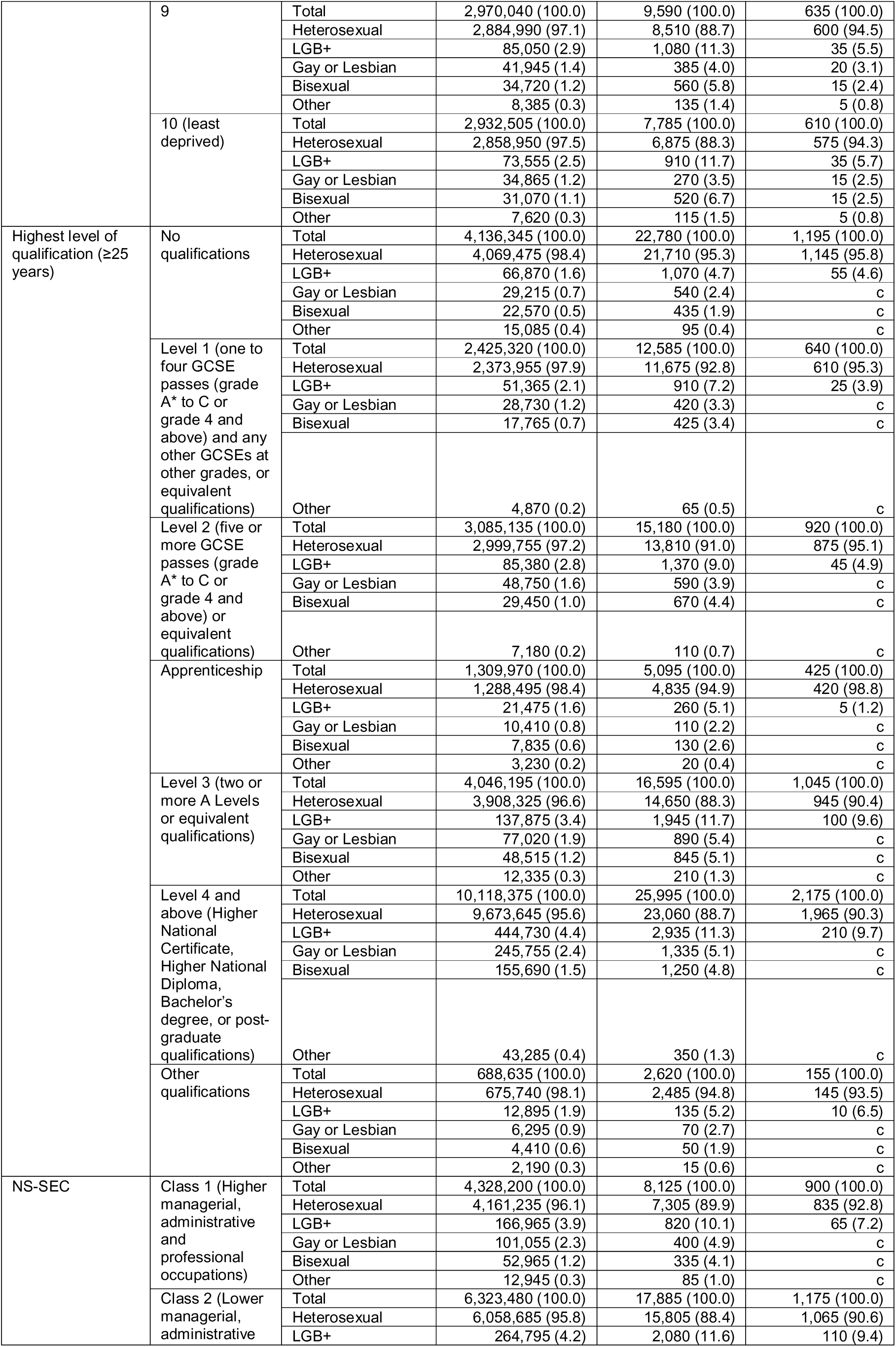
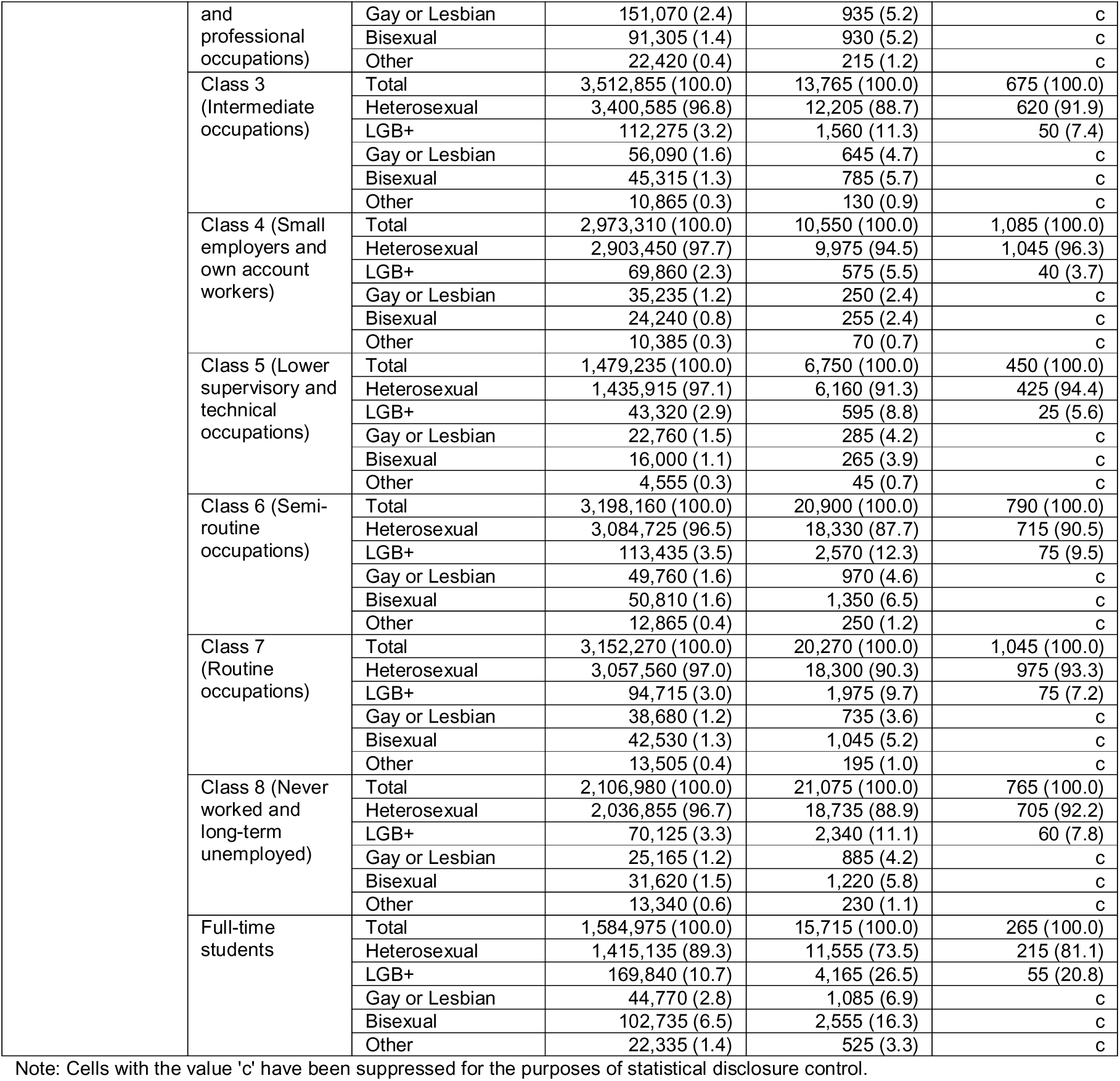
Characteristics of the study population.

Greater proportions of younger individuals identified as LGB+ (e.g., 10.0% of those aged 16-24 years compared with 0.8% of those ≥65 years). The Mixed ethnicity group had the highest proportion of LGB+ individuals (8.3%) while the Asian and Black ethnic groups had the lowest (2.4%). Higher proportions of LGB+ individuals lived in more deprived areas, with 4.6% reporting to be LGB+ in IMD/WIMD decile 1 (most deprived) compared with 2.5% in decile 10 (least deprived). Groups with higher education levels were more likely to identify as LGB+ (e.g., 4.4% of those holding a qualification at Level 4 and above (Higher National Certificate, Higher National Diploma, Bachelor’s degree and post-graduate qualifications), compared with 1.6% of those with no qualifications).

### SO differences in intentional self-harm

Among LGB+ individuals, 16,680 (1.5%) had at least one record for intentional self-harm during the study period, compared with 118,360 (0.4%) in the heterosexual group.

The rate of self-harm was more than twice as high among LGB+ individuals (RR 2.52, 95% CI 2.48-2.56) compared with heterosexual peers, with age-standardised rates of 1,508.9 per 100,000 people (hereafter /100,000) for the LGB+ population and 598.4/100,000 for the heterosexual population (Figure 2, Table 2).

**Figure 2:**
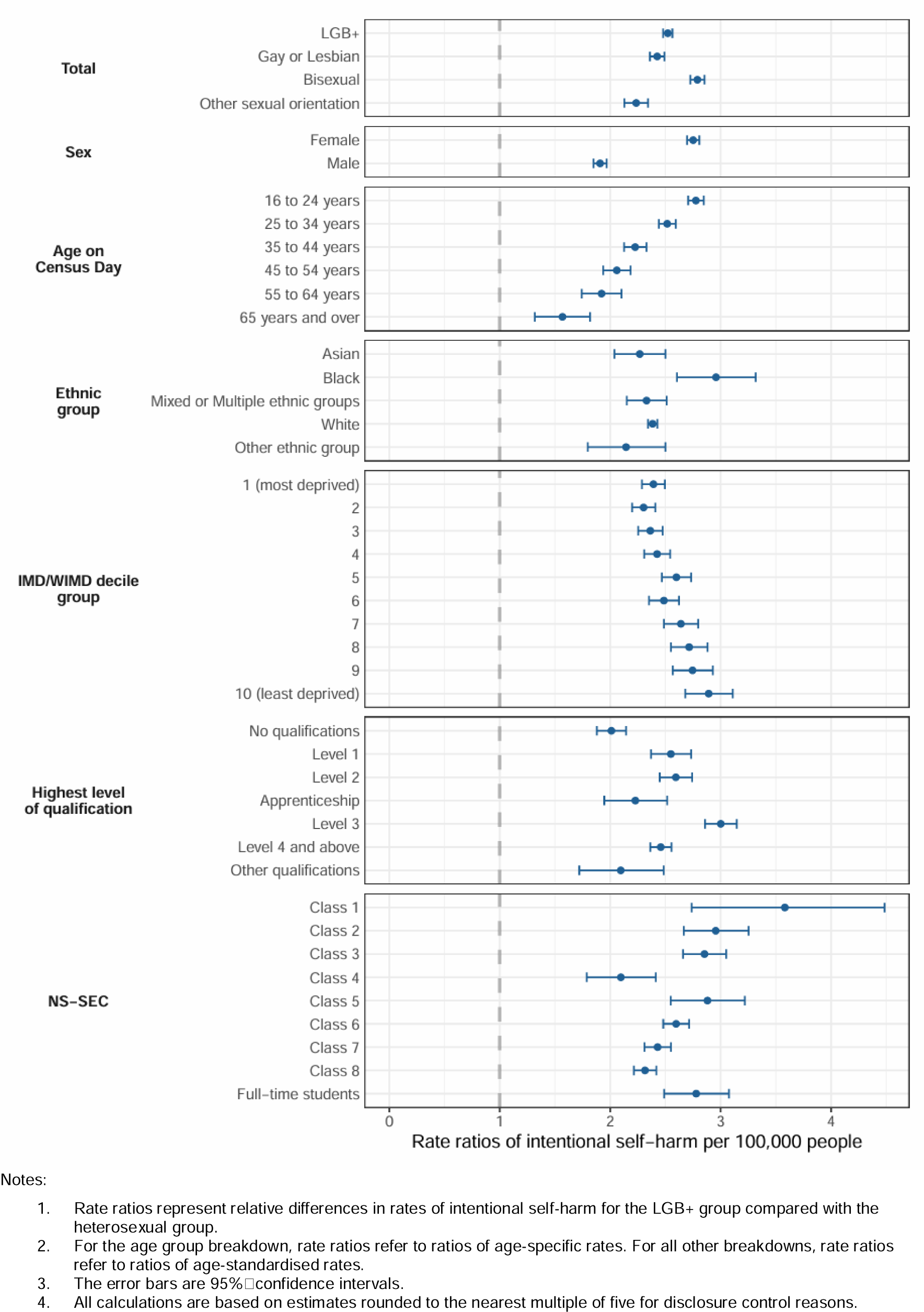
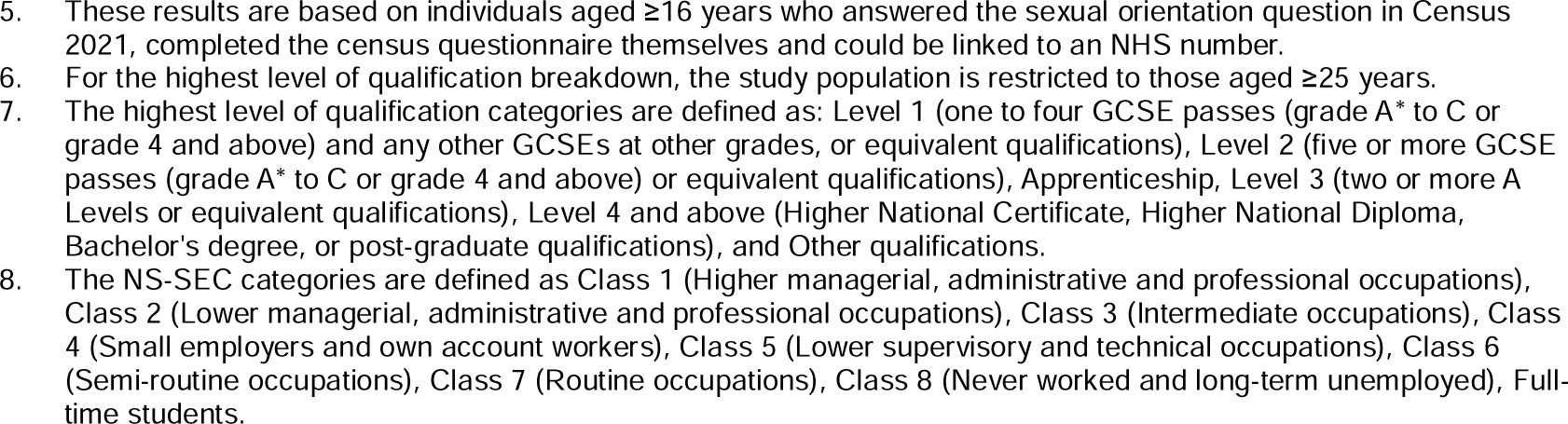
Rate ratios of intentional self-harm per 100,000 people for the LGB+ group relative to the heterosexual group

**Table 2:**
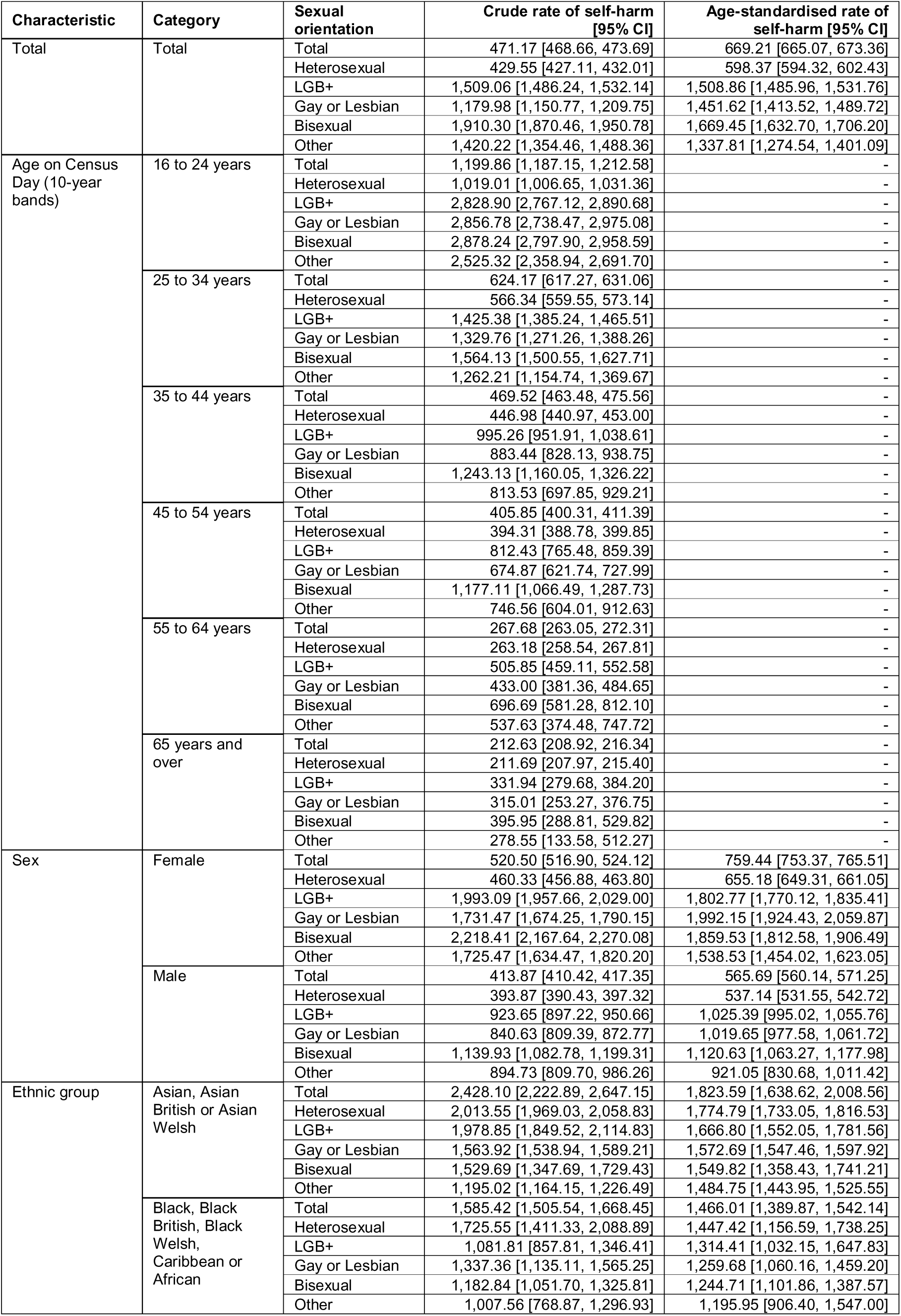
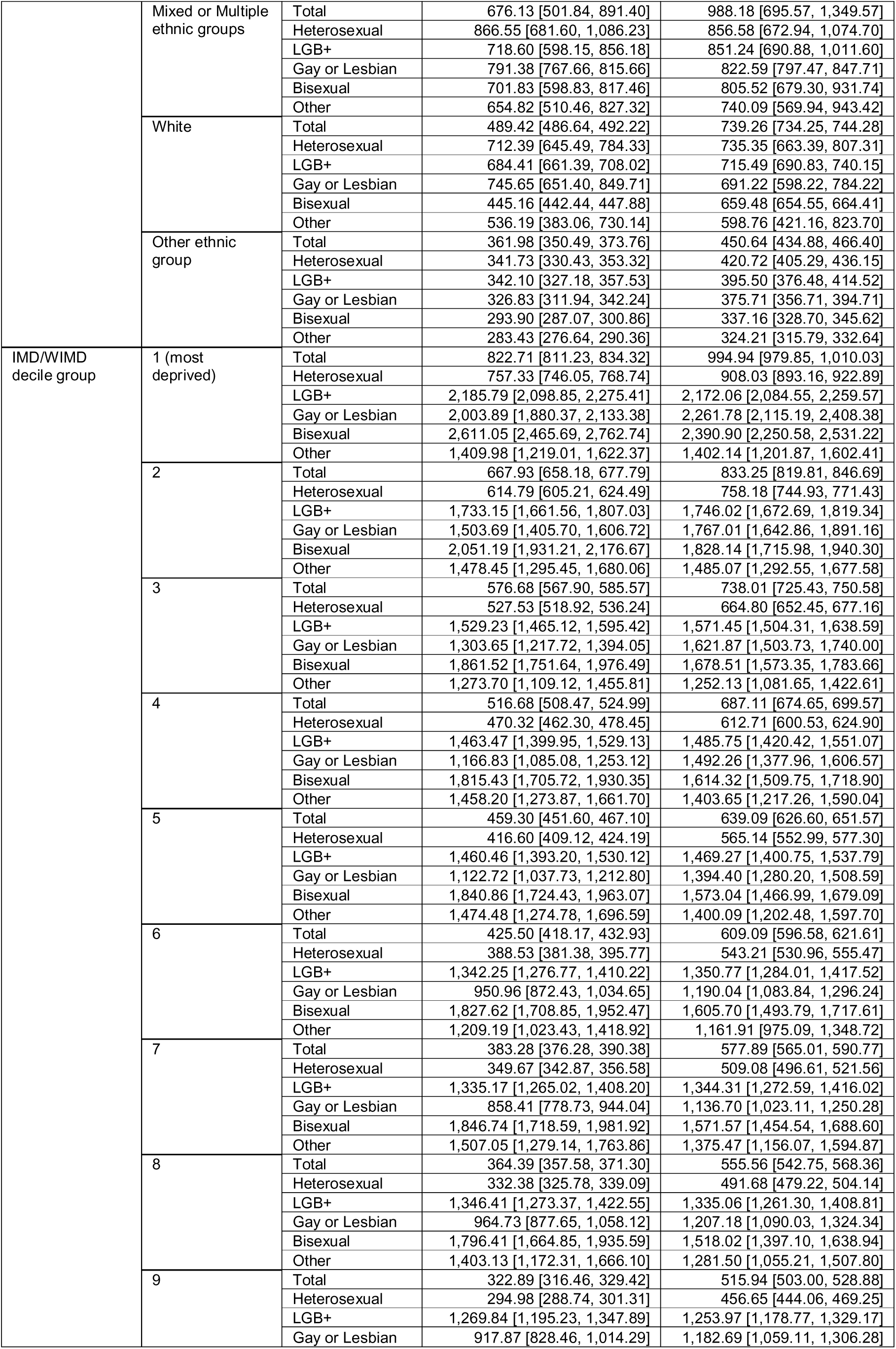
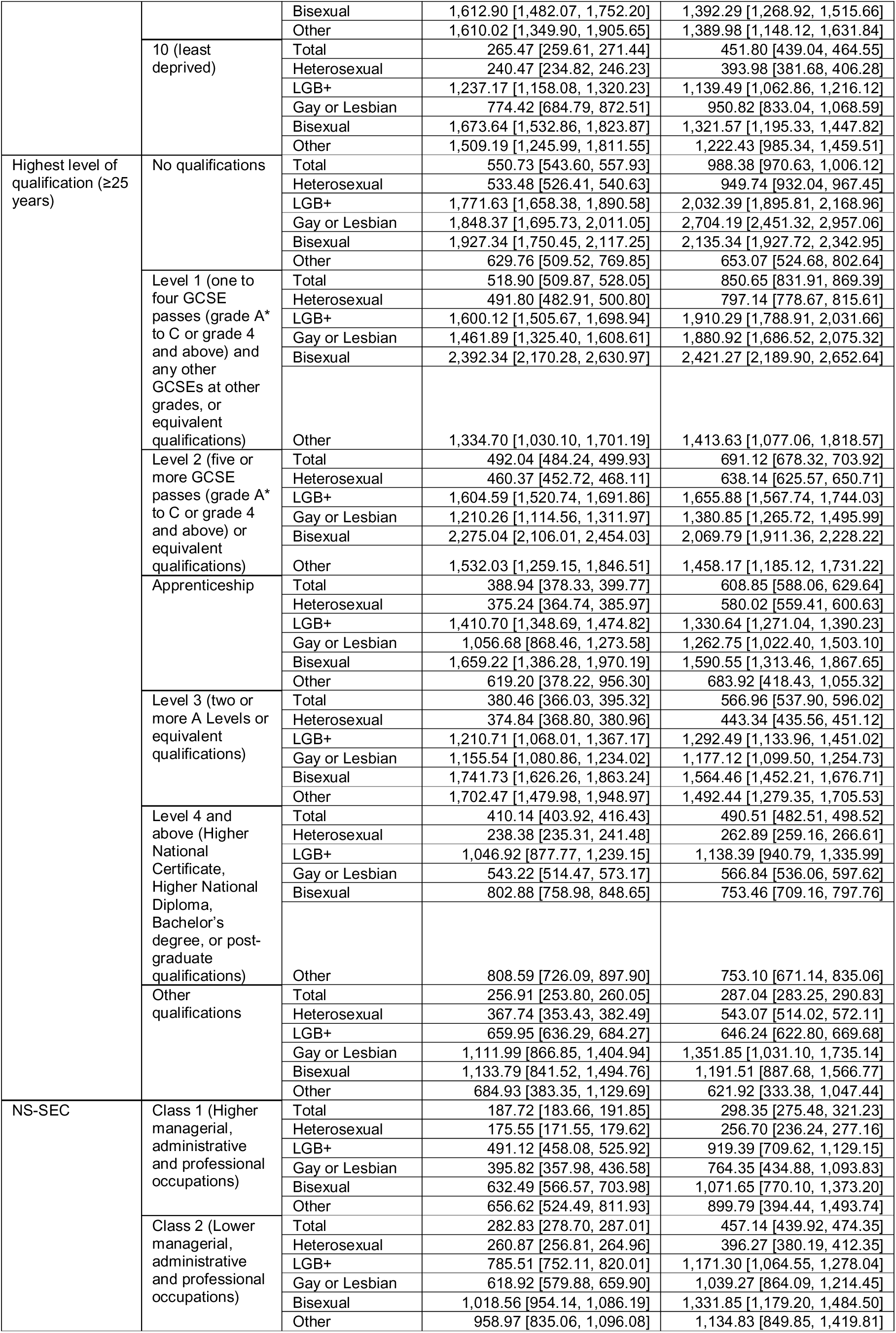
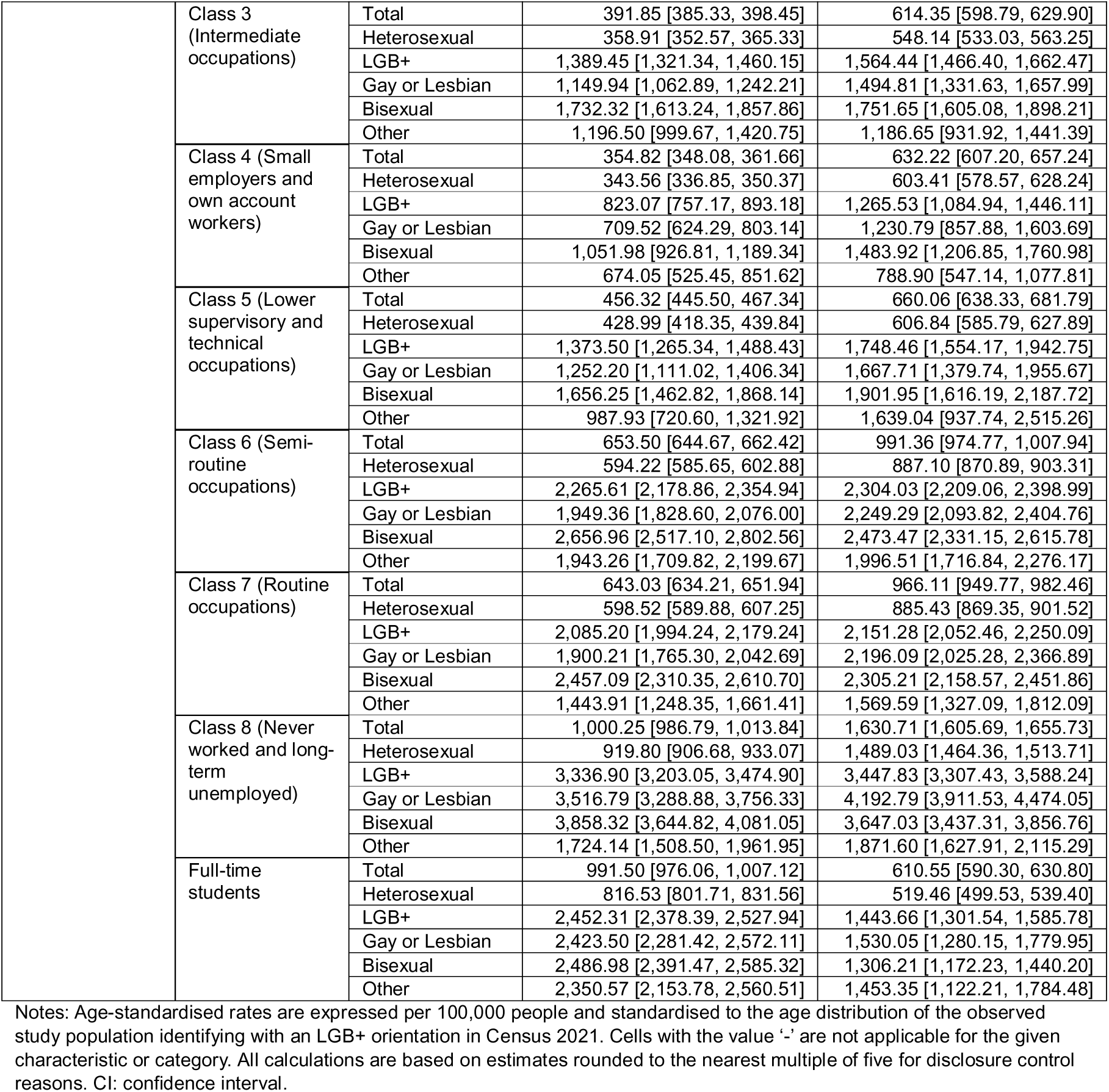
Crude and age-standardised rates of self-harm per 100,000 people.

Within the LGB+ population, the age-standardised rate of self-harm was highest for bisexual individuals (1,669.5/100,000; 2.79 times higher than heterosexual), followed by gay or lesbian individuals (1,451.6/100,000; 2.43 times higher than heterosexual) and those who identified with other sexual orientations (1,337.8/100,000; 2.24 times higher than heterosexual).

Age-standardised rates of self-harm were higher for females (759.4/100,000) than for males (565.7/100,000). Both females and males identifying as LGB+ had significantly higher rates of self-harm compared with those identifying as heterosexual. The rate of self-harm was significantly higher among LGB+ females (RR 2.75, 2.70-2.81) than LGB+ males (RR 1.91, 1.85-1.97) relative to heterosexual peers.

Age-specific rates of self-harm were higher for younger age groups than for older age groups (e.g., 1,199.9/100,000 aged 16-24 years, 212.6/100,000 aged ≥65 years) across all sexual orientations. Within each age group, LGB+ individuals were significantly more likely to have self-harmed compared with heterosexual peers, though RRs decreased with increasing age (e.g., RR 2.78, 2.71-2.85 among 16-24 years, RR 1.57, 1.32-1.82 among ≥65 years).

Age-standardised rates of self-harm were highest in the Mixed (822.6/100,000) and White (739.3/100,000) ethnic groups. In all five ethnic groups, LGB+ individuals had significantly higher rates of self-harm compared with heterosexual peers, with RRs highest among Black LGB+ individuals (RR 2.96, 2.61-3.32). LGB+ individuals born in the UK (RR 2.49, 2.45-2.54) and those born outside the UK (RR 2.36, 2.23-2.49) had similar relative increases in rates compared with heterosexual peers (Supplementary Table 6).

For the socioeconomic indicators IMD/WIMD, highest education level and NS-SEC, we found clear trends in rates of self-harm increasing with decreasing socioeconomic status for both heterosexual and LGB+ groups. For example, the age-standardised rate of self-harm in IMD/WIMD decile 1 (most deprived) was 994.9/100,000, decreasing to 451.8/100,000 in decile 10 (least deprived). Across all IMD/WIMD decile groups and education levels, LGB+ individuals were 2-3 times more likely to self-harm compared with heterosexual peers. Similarly, within each of the eight occupational classes (NS-SEC), LGB+ individuals were more than twice as likely to self-harm compared with heterosexual peers, with the highest RR observed for LGB+ individuals in Class 1 (higher managerial, administrative and professional occupations)^24^ (RR 3.58, 2.74-4.48).

### SO differences in suicide death

Among LGB+ individuals, 555 (0.05%) had at least one record for suicide death during the study period, compared with 6,595 (0.02%) in the heterosexual group.

The rate of suicide was 2.17 (1.98-2.37) times higher for people LGB+ individuals compared with heterosexual, with age-standardised rates of 50.3/100,000 for the LGB+ population and 23.1/100,000 for the heterosexual population (Figure 3, Table 3). Within the LGB+ population, differences in rates and RRs of suicide compared with heterosexual peers were not statistically significant.

**Figure 3:**
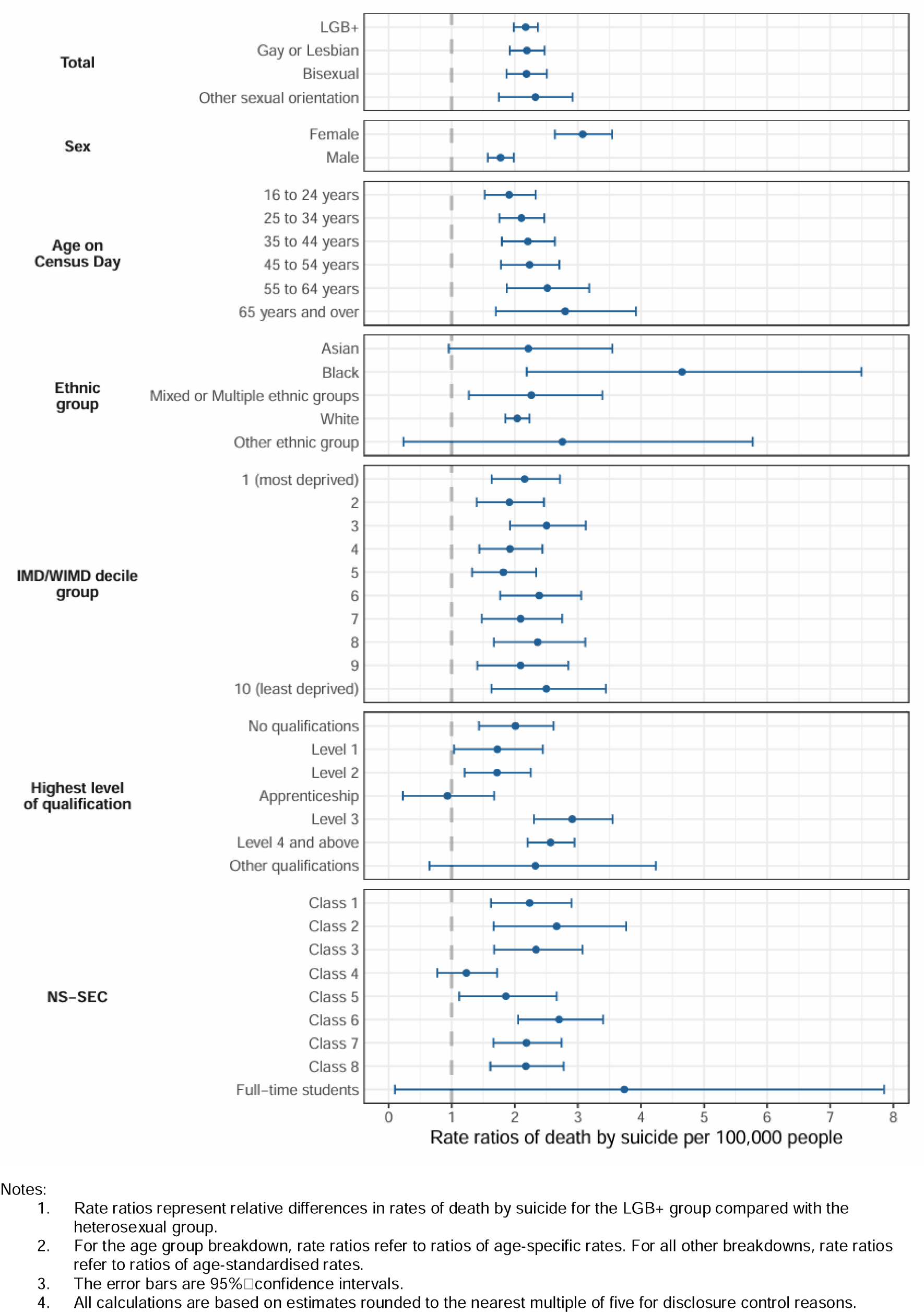
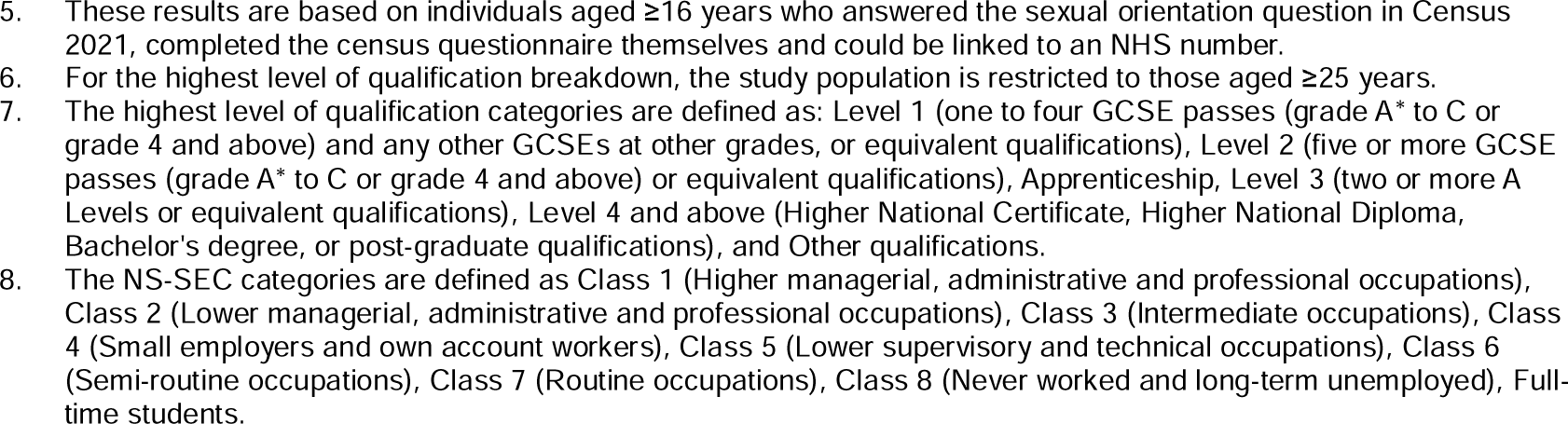
Rate ratios of death by suicide per 100,000 people for the LGB+ group relative to the heterosexual group

**Table 3:**
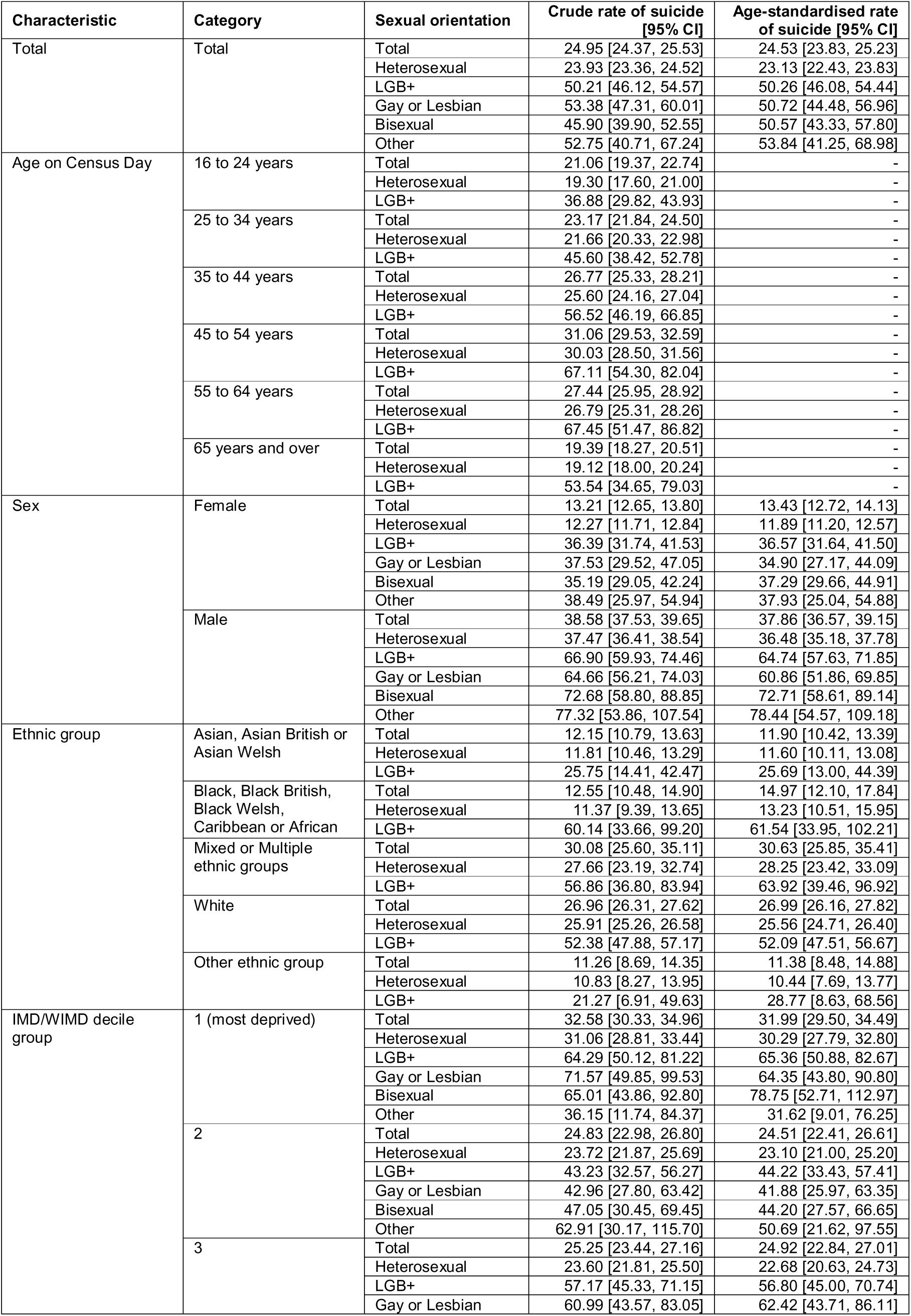
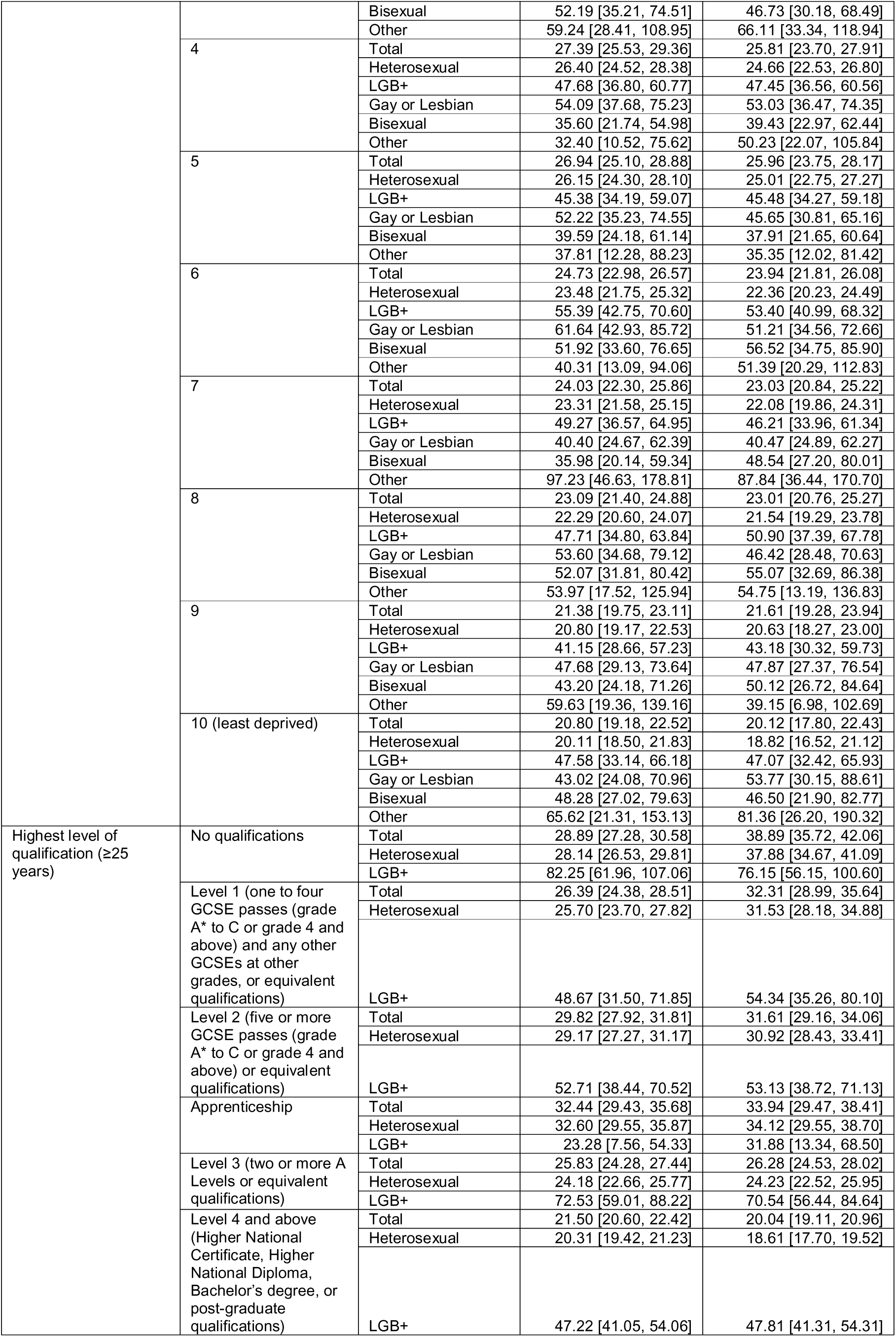
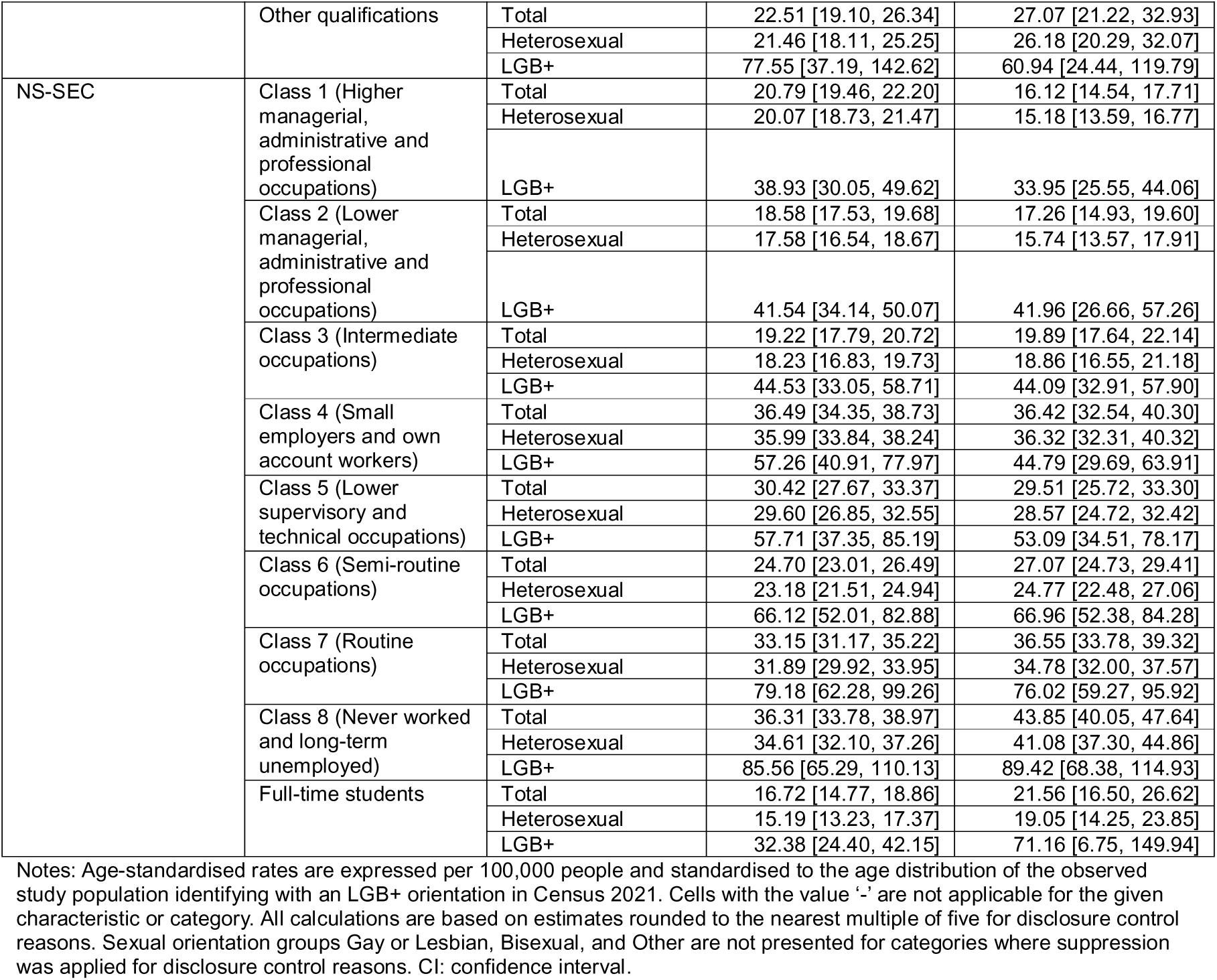
Crude and age-standardised rates of suicide per 100,000 people.

Age-standardised rates of suicide were higher for males (37.9/100,000) than for females (13.4/100,000). Both females and males identifying as LGB+ had significantly higher rates of suicide compared with heterosexual peers. The rate of suicide was higher among LGB+ females (RR 3.08, 2.64-3.54) than LGB+ males (RR 1.77, 1.57-1.98) relative to heterosexual peers.

Age-specific rates of suicide were highest for people aged 45-54 years (31.1/100,000). Within each age group, LGB+ individuals had significantly higher rates of suicide compared with heterosexual peers (e.g., RR 1.91, 1.52-2.33 for 16-24 years, RR 2.80, 1.70-3.92 for ≥65 years). Relative increases in rates of suicide for the LGB+ population compared with the heterosexual population were highest for older age groups (e.g., RR 2.80, 1.70-3.92 for ≥65 years).

Age-standardised rates of suicide were highest in the Mixed (30.6/100,000) and White (26.7/100,000) ethnic groups. Black LGB+ individuals had the highest RRs for suicide relative to heterosexual peers (RR 4.65, 2.19-7.50), and more than twice as high as observed for LGB+ peers from other ethnic groups (e.g., RR 2.04, 1.85-2.23 for White LGB+ individuals).

For the socioeconomic indicators IMD/WIMD, highest education level and NS-SEC, age-standardised rates of suicide increased with decreasing socioeconomic status (e.g., 32.0/100,000 in IMD/WIMD decile 1 (most deprived), falling to 20.1/100,000 in decile 10 (least deprived)). Within each IMD/WIMD decile group, LGB+ individuals were significantly more likely to have died by suicide compared with heterosexuals (e.g., RR 2.16, 1.63-2.72 in decile 1; RR 2.50, 1.63-3.45 in decile 10). LGB+ individuals with the highest qualification levels had the greatest relative increase in rates of suicide compared with heterosexual peers (RR 2.91, 2.30-3.55 for LGB+ individuals holding a qualification at Level 3 (two or more A Levels or equivalent qualifications); RR 2.57, 2.20-2.95 for LGB+ individuals holding a qualification at Level 4 and above (Higher National Certificate, Higher National Diploma, Bachelor’s degree and post-graduate qualifications)). LGB+ individuals were at higher risk of suicide than heterosexuals within all NS-SEC groups, though differences were not found to be statistically significant for those in Class 4 (small employers and own account workers) and full-time students. Further, differences in RRs between NS-SEC classes were not statistically significant.

### Sensitivity analyses

The findings were robust to (i) inclusion of proxy responses to Census 2021, (ii) restriction to people living in private households at the time of Census 2021, and (iii) inclusion of inpatient admissions from HES and PEDW for self-harm with undetermined intent within our outcome measure for self-harm (ICD-10 codes Y10-Y34) (Supplementary Tables 11-18).

## Discussion

### Principal findings

This study has produced the first national population-based estimates of SO inequalities in hospitalisation for intentional self-harm and death by suicide in England and Wales, and wider Europe. Both self-harm and suicide were 2-3 times higher in sexual minority groups compared with heterosexual peers, with the highest risk observed for bisexual and female LGB+ groups. This pattern was observed consistently across all sociodemographic and other factors. This study also reports for the first time SO inequalities in self-harm and suicide risk by faith group, and multiple indicators of socioeconomic position and household situation.

Our study corroborates previous findings on SO inequalities in self-harm and suicide including a recent meta-analysis which found 3.8-4.4 times higher odds for attempted suicide, suicide ideation and self-harm in >87,000 SMI individuals compared with heterosexual peers^3^. The majority of studies examining SO disparities in self-harm and suicide originate from the USA (58-71% of studies included in systematic reviews and meta-analyses originate from the USA^3^), with only one UK national survey reporting higher odds for self-harm and suicide in adult LGB+ groups but not accounting for the range of factors in our study^15^.

Mental health inequalities in sexual minority groups are explained by the “Minority stress theory”, which predicates that sexual minority individuals experience unique and long-term stress processes, which increases the risk for a wide range of mental health problems including self-harm and suicide^25^. Recent decades have witnessed greater social acceptance of and better equality rights for individuals with SMI (such as The Equality Act 2010, marriage rights, and adoption rights in the UK). Despite this, our findings indicate that sexual minority groups remain at increased risk of self-harm and suicide compared with heterosexuals.

When broken down by different demographic and socio-economic characteristics, risk of intentional self-harm was significantly higher for LGB+ individuals compared with heterosexual peers in every subgroup. A similar pattern was observed for suicide, though not always statistically significant for all subgroups. In the UK, ethnic minority adolescents and young adults have better mental health than White peers, but this is limited to those identifying as heterosexual^9^. Our analysis found that the risk of self-harm and suicide for Black LGB+ individuals was particularly high compared with Black heterosexual individuals. Limited research shows that ethnic minority and SMI individuals face a different set of challenges including ethnic- and cultural-specific obligations which are difficult to navigate over the life course, leading to worse mental health compared with heterosexual peers^9,26^.

### Strengths and limitations

The analysis uses a nationally representative population-level linked dataset, making it the largest study to date to examine rates of intentional self-harm and suicide by sexual orientation. This study covers all usual residents of England and Wales aged ≥16 years who self-reported their sexual orientation in Census 2021 and linked to an NHS number. The 2021 Census covers 97% of the population^27^ and provides high quality data on demographic and socio-economic factors for the population of England and Wales. This enabled us to compare risk of self-harm and suicide for sexual minority subgroups, and crucially by sociodemographic, geographical, socioeconomic and health-related factors, including analysis of multiple minoritised individuals. For the analysis of self-harm in particular, we had the statistical power to detect many small differences across groups.

We acknowledge limitations. Some people will be omitted from the study population due to not completing the 2021 Census, such as non-respondents (which may be biased towards some groups e.g., those with no fixed address^27^), and those who migrated into England and Wales after the 2021 Census date.

Additionally, Census respondents who did not link to an NHS number could not be included in the study population. To mitigate against this, we adjusted for non-linkage by applying post-stratification weights by numerous factors potentially related to both linkage propensity and adverse outcomes^20^.

Our estimates of self-harm are based on people who were admitted to an NHS hospital or had an emergency attendance for intentional self-harm in England or Wales. However, reported statistics are likely a significant underestimate^28,29^. We acknowledge that NHS hospital data will not cover the whole self-harming population, such as those who did not attend hospital or those accessing private healthcare. Additionally, the recording of intent in administrative health records can be inconsistent, with 17.9% of ECDS records missing both the chief complaint and injury intent, and 36.4% of EDDS records missing a reason for admission or recording an “unknown” reason for admission.

## Conclusions

This study provides the first national population-based estimates of intentional self-harm and suicide by sexual orientation in England and Wales. Our findings show that LGB+ individuals are at a substantially increased risk of self-harm and suicide compared with heterosexual peers. This analysis also identified key groups of individuals at an increased risk of self-harm and suicide. These findings are important for informing government prevention programs and further research supporting the mental health of sexual minority groups. Future work should explore further comparisons by sexual orientation such as deaths from other causes, diagnoses of severe mental illnesses and access to mental healthcare services.

## Contributors

ES, VN and AK conceived the study. HB, ES and AK designed the study. HB led the analysis with support from ES, VN, DA and AK. All authors quality assured the outcomes and the interpretations of them. All authors contributed to the critical revision of the manuscript and approved the final version.

## Declaration of interests

All authors declare no competing interests.

## Funding source

There was no external funding for this study.

## Data sharing

The source data are not publicly available and are subject to controlled access due to their sensitive nature. Non-linked death registrations data are available to Accredited Researchers through the Secure Research Service (SRS). Details of the application requirements and process, and the use of data, are available at https://www.ons.gov.uk/aboutus/whatwedo/statistics/requestingstatistics/secureresearchservice. The non-linked HES and ECDS data are held by NHS England and can be accessed through the NHS Secure Data Environment: https://digital.nhs.uk/services/secure-data-environment-service. The non-linked PEDW and EDDS data are held by DHCW.

## Code sharing

The code for this study is publicly available in GitHub: https://github.com/ONS-Health-modelling-hub/self-harm_suicide_sexual_orientation.

## Data Availability

All data relating to this work has been published as an Office for National Statistics (ONS) dataset.

https://www.ons.gov.uk/peoplepopulationandcommunity/healthandsocialcare/mentalhealth/datasets/selfharmandsuicidebysexualorientationenglandandwales

## Notes

### Competing Interest Statement

The authors have declared no competing interest.

### Funding Statement

This study did not receive any external funding.

### Author Declarations

The datasets used in the study were individual-level and had been de-identified prior to use in the study.

