## Supplementary material for "Sexual orientation inequalities in self-harm and suicidality in England and Wales – A national population-based study"

**Supplementary Figure 1: Datasets used in this study**

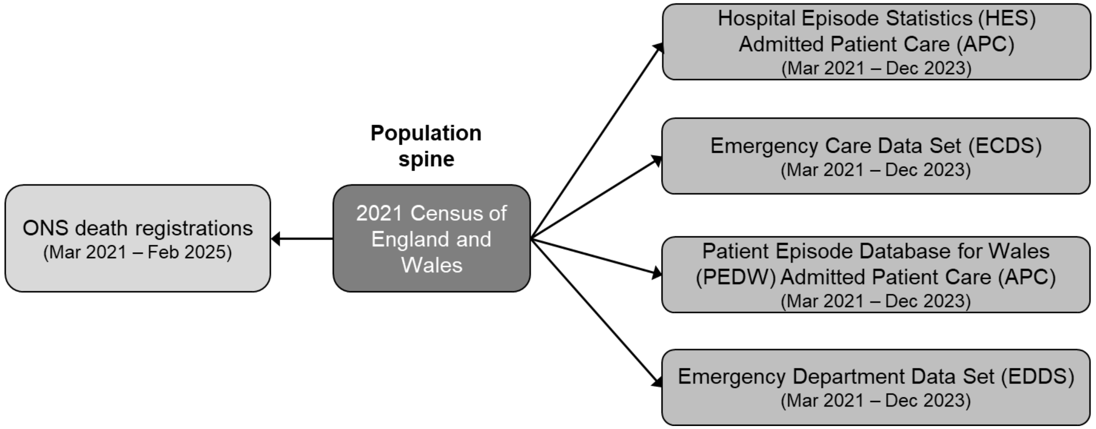


**Supplementary Table 1: Sample flow**

| Sample | Count, Individuals | Count, Individuals with at least one hospital admission or emergency attendance for intentional self-harm | Count, Individuals with death by suicide |
|---|---|---|---|
| All people with a hospital episode recorded in the Hospital Episode Statistics (HES) Admitted Patient Care (APC), Emergency Care Data Set (ECDS), Patient Episode Database for Wales (PEDW) Admitted Patient Care (APC) or Emergency Department Data Set (EDDS) data which started and ended between 21 March 2021 to 31 December 2023 | 24,290,465 | 397,365 | - |
| All people with a hospital episode recorded in the HES APC, ECDS, PEDW APC or EDDS data which started and ended between 21 March 2021 to 31 December 2023, who linked to the NHS Personal Demographics Service (PDS) 2019 | 20,126,070 | 316,490 | - |
| Deaths occurring between 21 March 2021 to 31 December 2023 and registered by 10 February 2025 | 1,589,295 | - | 15,860 |
| Deaths occurring between 21 March 2021 to 31 December 2023, registered by 10 February 2025 and linked to the NHS PDS 2019 | 1,430,270 | - | 12,100 |
| All people enumerated in Census 2021 who were usual residents in England and Wales | 59,597,540 | - | - |
| Aged 16 years and over | 48,566,375 | - | - |
| Answered the sexual orientation question in Census 2021 | 44,939,725 | - | - |
| Not wholly imputed | 43,618,145 | - | - |
| Answered the Census 2021 questionnaire themselves (not by proxy) | 30,295,500 | - | - |
| Linked to the NHS PDS (unweighted) | 28,661,205 | - | - |
| Linked to the NHS PDS (weighted for non-linkage to the PDS) | 28,660,990 | - | - |
| Alive on 21 March 2021 (unweighted) | 28,659,625 | 131,960 | 7,105 |
| Study population: Alive on 21 March 2021 (weighted for non-linkage to the PDS) | 28,659,465 | 135,035 | 7,150 |

Note: Cells with the value '-' are not applicable for the given sample.

**Supplementary Table 2: List of codes used to define intentional self-harm**

| Data source | Code type | Code and description |
|---|---|---|
| Hospital Episode Statistics (HES) Admitted Patient Care (APC) | International Classification of Diseases, Tenth Revision (ICD-10) | X60 Intentional self-poisoning by and exposure to nonopioid analgesics, antipyretics and antirheumatics<br>X61 Intentional self-poisoning by and exposure to antiepileptic, sedative-hypnotic, antiparkinsonism and psychotropic drugs, not elsewhere classified<br>X62 Intentional self-poisoning by and exposure to narcotics and psychodysleptics [hallucinogens], not elsewhere classified<br>X63 Intentional self-poisoning by and exposure to other drugs acting on the autonomic nervous system<br>X64 Intentional self-poisoning by and exposure to other and unspecified drugs, medicaments and biological substances<br>X65 Intentional self-poisoning by and exposure to alcohol<br>X66 Intentional self-poisoning by and exposure to organic solvents and halogenated hydrocarbons and their vapours<br>X67 Intentional self-poisoning by and exposure to carbon monoxide and other gases and vapours<br>X68 Intentional self-poisoning by and exposure to pesticides<br>X69 Intentional self-poisoning by and exposure to other and unspecified chemicals and noxious substances<br>X70 Intentional self-harm by hanging, strangulation and suffocation<br>X71 Intentional self-harm by drowning and submersion<br>X72 Intentional self-harm by handgun discharge<br>X73 Intentional self-harm by rifle, shotgun and larger firearm discharge<br>X74 Intentional self-harm by other and unspecified firearm discharge<br>X75 Intentional self-harm by explosive material<br>X76 Intentional self-harm by smoke, fire and flames<br>X77 Intentional self-harm by steam, hot vapours and hot objects<br>X78 Intentional self-harm by sharp object<br>X79 Intentional self-harm by blunt object<br>X80 Intentional self-harm by jumping from a high place<br>X81 Intentional self-harm by jumping or lying before moving object<br>X82 Intentional self-harm by crashing of motor vehicle<br>X83 Intentional self-harm by other specified means<br>X84 Intentional self-harm by unspecified means |
| Patient Episode Database for Wales (PEDW) Admitted Patient Care (APC) | International Classification of Diseases, Tenth Revision (ICD-10) | X60 Intentional self-poisoning by and exposure to nonopioid analgesics, antipyretics and antirheumatics<br>X61 Intentional self-poisoning by and exposure to antiepileptic, sedative-hypnotic, antiparkinsonism and psychotropic drugs, not elsewhere classified<br>X62 Intentional self-poisoning by and exposure to narcotics and psychodysleptics [hallucinogens], not elsewhere classified<br>X63 Intentional self-poisoning by and exposure to other drugs acting on the autonomic nervous system<br>X64 Intentional self-poisoning by and exposure to other and unspecified drugs, medicaments and biological substances<br>X65 Intentional self-poisoning by and exposure to alcohol<br>X66 Intentional self-poisoning by and exposure to organic solvents and halogenated hydrocarbons and their vapours<br>X67 Intentional self-poisoning by and exposure to carbon monoxide and other gases and vapours<br>X68 Intentional self-poisoning by and exposure to pesticides<br>X69 Intentional self-poisoning by and exposure to other and unspecified chemicals and noxious substances<br>X70 Intentional self-harm by hanging, strangulation and suffocation<br>X71 Intentional self-harm by drowning and submersion<br>X72 Intentional self-harm by handgun discharge<br>X73 Intentional self-harm by rifle, shotgun and larger firearm discharge<br>X74 Intentional self-harm by other and unspecified firearm discharge<br>X75 Intentional self-harm by explosive material<br>X76 Intentional self-harm by smoke, fire and flames<br>X77 Intentional self-harm by steam, hot vapours and hot objects<br>X78 Intentional self-harm by sharp object<br>X79 Intentional self-harm by blunt object<br>X80 Intentional self-harm by jumping from a high place<br>X81 Intentional self-harm by jumping or lying before moving object<br>X82 Intentional self-harm by crashing of motor vehicle<br>X83 Intentional self-harm by other specified means<br>X84 Intentional self-harm by unspecified means |
| Emergency Care Data Set (ECDS) | Chief complaint | 248062006 Self-injurious behaviour (finding) |
| | Injury intent | 276853009 Self-inflicted injury (disorder) |
| Emergency Department Data Set (EDDS) | Attendance group | 13 Deliberate self-harm |

**Supplementary Table 3: List of International Classification of Diseases, Tenth Revision (ICD-10) codes used to define death by suicide**

| ICD-10 code | Description |
|---|---|
| X60 | Intentional self-poisoning by and exposure to nonopioid analgesics, antipyretics and antirheumatics |
| X61 | Intentional self-poisoning by and exposure to antiepileptic, sedative-hypnotic, antiparkinsonism and psychotropic drugs, not elsewhere classified |
| X62 | Intentional self-poisoning by and exposure to narcotics and psychodysleptics [hallucinogens], not elsewhere classified |
| X63 | Intentional self-poisoning by and exposure to other drugs acting on the autonomic nervous system |
| X64 | Intentional self-poisoning by and exposure to other and unspecified drugs, medicaments and biological substances |
| X65 | Intentional self-poisoning by and exposure to alcohol |
| X66 | Intentional self-poisoning by and exposure to organic solvents and halogenated hydrocarbons and their vapours |
| X67 | Intentional self-poisoning by and exposure to carbon monoxide and other gases and vapours |
| X68 | Intentional self-poisoning by and exposure to pesticides |
| X69 | Intentional self-poisoning by and exposure to other and unspecified chemicals and noxious substances |
| X70 | Intentional self-harm by hanging, strangulation and suffocation |
| X71 | Intentional self-harm by drowning and submersion |
| X72 | Intentional self-harm by handgun discharge |
| X73 | Intentional self-harm by rifle, shotgun and larger firearm discharge |
| X74 | Intentional self-harm by other and unspecified firearm discharge |
| X75 | Intentional self-harm by explosive material |
| X76 | Intentional self-harm by smoke, fire and flames |
| X77 | Intentional self-harm by steam, hot vapours and hot objects |
| X78 | Intentional self-harm by sharp object |
| X79 | Intentional self-harm by blunt object |
| X80 | Intentional self-harm by jumping from a high place |
| X81 | Intentional self-harm by jumping or lying before moving object |
| X82 | Intentional self-harm by crashing of motor vehicle |
| X83 | Intentional self-harm by other specified means |
| X84 | Intentional self-harm by unspecified means |
| Y10 | Poisoning by and exposure to nonopioid analgesics, antipyretics and antirheumatics, undetermined intent |
| Y11 | Poisoning by and exposure to antiepileptic, sedative-hypnotic, antiparkinsonism and psychotropic drugs, not elsewhere classified, undetermined intent |
| Y12 | Poisoning by and exposure to narcotics and psychodysleptics [hallucinogens], not elsewhere classified, undetermined intent |
| Y13 | Poisoning by and exposure to other drugs acting on the autonomic nervous system, undetermined intent |
| Y14 | Poisoning by and exposure to other and unspecified drugs, medicaments and biological substances, undetermined intent |
| Y15 | Poisoning by and exposure to alcohol, undetermined intent |
| Y16 | Poisoning by and exposure to organic solvents and halogenated hydrocarbons and their vapours, undetermined intent |
| Y17 | Poisoning by and exposure to carbon monoxide and other gases and vapours, undetermined intent |
| Y18 | Poisoning by and exposure to pesticides, undetermined intent |
| Y19 | Poisoning by and exposure to other and unspecified chemicals and noxious substances, undetermined intent |
| Y20 | Hanging, strangulation and suffocation, undetermined intent |
| Y21 | Drowning and submersion, undetermined intent |
| Y22 | Handgun discharge, undetermined intent |
| Y23 | Rifle, shotgun and larger firearm discharge, undetermined intent |
| Y24 | Other and unspecified firearm discharge, undetermined intent |
| Y25 | Contact with explosive material, undetermined intent |
| Y26 | Exposure to smoke, fire and flames, undetermined intent |
| Y27 | Contact with steam, hot vapours and hot objects, undetermined intent |
| Y28 | Contact with sharp object, undetermined intent |
| Y29 | Contact with blunt object, undetermined intent |
| Y30 | Falling, jumping or pushed from a high place, undetermined intent |
| Y31 | Falling, lying or running before or into moving object, undetermined intent |
| Y32 | Crashing of motor vehicle, undetermined intent |
| Y33 | Other specified events, undetermined intent |
| Y34 | Unspecified event, undetermined intent |

Note: Deaths by suicide were sourced from the Office for National Statistics (ONS) death registrations.

**Supplementary Table 4: Classifications of population characteristics**

| Characteristic | Data source | Classification |
|---|---|---|
| Sexual orientation | Census 2021 | Heterosexual<br>Gay or Lesbian<br>Bisexual<br>Other<br>LGB+ (Gay or Lesbian, Bisexual, or Other) |

| | | |
|---|---|---|
| Age on Census Day (10-year bands) | Census 2021 | 16 to 24 years<br>25 to 34 years<br>35 to 44 years<br>45 to 54 years<br>55 to 64 years<br>65 years and over |
| Sex | Census 2021 | Female<br>Male |
| Region | Census 2021 | England: North East<br>England: North West<br>England: Yorkshire and the Humber<br>England: East Midlands<br>England: West Midlands<br>England: East of England<br>England: London<br>England: South East<br>England: South West<br>Wales |
| Rural/urban classification | Census 2021 | Urban<br>Rural |
| Ethnic group | Census 2021 | Asian, Asian British or Asian Welsh<br>Black, Black British, Black Welsh, Caribbean or African<br>Mixed or Multiple ethnic groups<br>White<br>Other ethnic group |
| Country of birth | Census 2021 | Born in the UK<br>Born outside the UK |
| English language proficiency | Census 2021 | Main language is English<br>Main language is not English |
| Religion | Census 2021 | No religion<br>Christian<br>Buddhist<br>Hindu<br>Jewish<br>Muslim<br>Sikh<br>Other religion<br>Not answered |
| Index of Multiple Deprivation (IMD)/Welsh Index of Multiple Deprivation (WIMD) decile group | Census 2021 | 1 (most deprived)<br>2<br>3<br>4<br>5<br>6<br>7<br>8<br>9<br>10 (least deprived) |
| Highest level of qualification (≥25 years) | Census 2021 | No academic or professional qualifications<br>Level 1: one to four GCSE passes (grade A* to C or grade 4 and above) and any other GCSEs at other grades, or equivalent qualifications<br>Level 2: five or more GCSE passes (grade A* to C or grade 4 and above) or equivalent qualifications<br>Apprenticeship<br>Level 3: two or more A Levels or equivalent qualifications<br>Level 4 and above: Higher National Certificate, Higher National Diploma, Bachelor’s degree, or post-graduate qualifications<br>Other qualifications |
| National Statistics Socio-Economic Classification (NS-SEC) | Census 2021 | Class 1: Higher managerial, administrative and professional occupations<br>Class 2: Lower managerial, administrative and professional occupations<br>Class 3: Intermediate occupations<br>Class 4: Small employers and own account workers<br>Class 5: Lower supervisory and technical occupations<br>Class 6: Semi-routine occupations<br>Class 7: Routine occupations<br>Class 8: Never worked and long-term unemployed<br>Full-time students |
| Household National Statistics Socio-Economic Classification (NS-SEC) | Census 2021 | Class 1: Higher managerial, administrative and professional occupations<br>Class 2: Lower managerial, administrative and professional occupations |

| | | |
|---|---|---|
| | | Class 3: Intermediate occupations<br>Class 4: Small employers and own account workers<br>Class 5: Lower supervisory and technical occupations<br>Class 6: Semi-routine occupations<br>Class 7: Routine occupations<br>Class 8: Never worked and long-term unemployed<br>Full-time students<br>Living in a communal establishment |
| Lone person household indicator | Census 2021 | Lives alone<br>Does not live alone<br>Living in a communal establishment |
| Household size | Census 2021 | One person in household<br>Two people in household<br>Three to five people in household<br>Six or more people in household<br>Living in a communal establishment |
| Household tenure | Census 2021 | Owned or shared ownership<br>Social rented<br>Private rented or living rent free<br>Living in a communal establishment |
| Economic activity | Census 2021 | Economically active<br>Economically inactive |
| General health (detailed) | Census 2021 | Very good health<br>Good health<br>Fair health<br>Bad health<br>Very bad health |
| General health (aggregated) | Census 2021 | Good health<br>Not good health |
| Disability status | Census 2021 | Day-to-day activities not limited<br>Day-to-day activities limited a little<br>Day-to-day activities limited a lot |
| Residence type | Census 2021 | Living in a private household<br>Living in a communal establishment |

Notes: The age group 65 years and over has been aggregated for disclosure control reasons.
Further information on highest level of qualification is available from:
https://www.ons.gov.uk/census/census2021dictionary/variablesbytopic/educationvariablescensus2021/highestlevelofqualification.
Further information on NS-SEC is available from:
https://www.ons.gov.uk/census/census2021dictionary/variablesbytopic/labourmarketvariablescensus2021/nationalstatisticssocio-economicclassificationns-sec.
Further information on IMD is available from: https://www.gov.uk/government/statistics/english-indices-of-deprivation-2019.
Further information on WIMD is available from: https://www.gov.wales/welsh-index-multiple-deprivation.

**Supplementary Table 5: Characteristics of the study population by region, rural/urban classification, country of birth, English language proficiency, religion, household NS-SEC, lone person household indicator, household size, household tenure, economic activity, general health, disability, and residence type**

| **Characteristic** | **Category** | **Sexual orientation** | **Count (%), Individuals** | **Count (%), Individuals with at least one hospital admission or emergency attendance for intentional self-harm** | **Count (%), Individuals with death by suicide** |
|---|---|---|---|---|---|
| Region | England: North East | Total | 1,330,595 (100.0) | 8,215 (100.0) | 420 (100.0) |
| | | Heterosexual | 1,283,770 (96.5) | 7,280 (88.6) | 390 (92.9) |
| | | LGB+ | 46,825 (3.5) | 935 (11.4) | 35 (8.3) |
| | | Gay or Lesbian | 23,665 (1.8) | 425 (5.2) | c |
| | | Bisexual | 18,945 (1.4) | 420 (5.1) | c |
| | | Other | 4,215 (0.3) | 90 (1.1) | c |
| | England: North West | Total | 3,659,340 (100.0) | 16,475 (100.0) | 1,065 (100.0) |
| | | Heterosexual | 3,521,115 (96.2) | 14,290 (86.7) | 965 (90.6) |
| | | LGB+ | 138,225 (3.8) | 2,185 (13.3) | 100 (9.4) |
| | | Gay or Lesbian | 71,195 (1.9) | 950 (5.8) | c |
| | | Bisexual | 53,820 (1.5) | 1,065 (6.5) | c |
| | | Other | 13,210 (0.4) | 175 (1.1) | c |
| | England: Yorkshire and The Humber | Total | 2,627,670 (100.0) | 18,185 (100.0) | 745 (100.0) |
| | | Heterosexual | 2,529,495 (96.3) | 16,345 (89.9) | 685 (91.9) |
| | | LGB+ | 98,175 (3.7) | 1,840 (10.1) | 55 (7.4) |

| | | | | | |
|---|---|---|---|---|---|
| | | Gay or Lesbian | 44,200 (1.7) | 700 (3.8) | c |
| | | Bisexual | 42,885 (1.6) | 945 (5.2) | c |
| | | Other | 11,085 (0.4) | 190 (1.0) | c |
| | England: East Midlands | Total | 2,290,650 (100.0) | 12,170 (100.0) | 605 (100.0) |
| | | Heterosexual | 2,211,910 (96.6) | 10,660 (87.6) | 555 (91.7) |
| | | LGB+ | 78,740 (3.4) | 1,510 (12.4) | 50 (8.3) |
| | | Gay or Lesbian | 34,430 (1.5) | 525 (4.3) | c |
| | | Bisexual | 35,525 (1.6) | 835 (6.9) | c |
| | | Other | 8,785 (0.4) | 150 (1.2) | c |
| | England: West Midlands | Total | 2,684,810 (100.0) | 10,380 (100.0) | 675 (100.0) |
| | | Heterosexual | 2,600,505 (96.9) | 9,140 (88.1) | 645 (95.6) |
| | | LGB+ | 84,310 (3.1) | 1,240 (11.9) | 30 (4.4) |
| | | Gay or Lesbian | 38,695 (1.4) | 470 (4.5) | c |
| | | Bisexual | 35,760 (1.3) | 655 (6.3) | c |
| | | Other | 9,855 (0.4) | 115 (1.1) | c |
| | England: East of England | Total | 3,121,770 (100.0) | 14,970 (100.0) | 680 (100.0) |
| | | Heterosexual | 3,022,385 (96.8) | 13,275 (88.7) | 630 (92.6) |
| | | LGB+ | 99,385 (3.2) | 1,695 (11.3) | 45 (6.6) |
| | | Gay or Lesbian | 44,105 (1.4) | 575 (3.8) | c |
| | | Bisexual | 43,470 (1.4) | 930 (6.2) | c |
| | | Other | 11,810 (0.4) | 190 (1.3) | c |
| | England: London | Total | 4,329,065 (100.0) | 16,565 (100.0) | 720 (100.0) |
| | | Heterosexual | 4,096,810 (94.6) | 14,600 (88.1) | 640 (88.9) |
| | | LGB+ | 232,255 (5.4) | 1,965 (11.9) | 80 (11.1) |
| | | Gay or Lesbian | 119,415 (2.8) | 740 (4.5) | c |
| | | Bisexual | 83,985 (1.9) | 955 (5.8) | c |
| | | Other | 28,855 (0.7) | 275 (1.7) | c |
| | England: South East | Total | 4,642,855 (100.0) | 20,470 (100.0) | 1,175 (100.0) |
| | | Heterosexual | 4,470,840 (96.3) | 17,740 (86.7) | 1,095 (93.2) |
| | | LGB+ | 172,015 (3.7) | 2,730 (13.3) | 80 (6.8) |
| | | Gay or Lesbian | 80,120 (1.7) | 900 (4.4) | c |
| | | Bisexual | 73,040 (1.6) | 1,530 (7.5) | c |
| | | Other | 18,850 (0.4) | 300 (1.5) | c |
| | England: South West | Total | 2,760,745 (100.0) | 12,870 (100.0) | 745 (100.0) |
| | | Heterosexual | 2,653,295 (96.1) | 10,900 (84.7) | 700 (94.0) |
| | | LGB+ | 107,450 (3.9) | 1,970 (15.3) | 50 (6.7) |
| | | Gay or Lesbian | 46,135 (1.7) | 665 (5.2) | c |
| | | Bisexual | 49,505 (1.8) | 1,085 (8.4) | c |
| | | Other | 11,805 (0.4) | 220 (1.7) | c |
| | Wales | Total | 1,211,965 (100.0) | 4,735 (100.0) | 315 (100.0) |
| | | Heterosexual | 1,164,015 (96.0) | 4,130 (87.2) | 285 (90.5) |
| | | LGB+ | 47,950 (4.0) | 605 (12.8) | 30 (9.5) |
| | | Gay or Lesbian | 22,625 (1.9) | 245 (5.2) | c |
| | | Bisexual | 20,585 (1.7) | 320 (6.8) | c |
| | | Other | 4,745 (0.4) | 40 (0.8) | c |
| Rural/urban classification | Urban | Total | 23,366,335 (100.0) | 115,495 (100.0) | 5,760 (100.0) |
| | | Heterosexual | 22,395,100 (95.8) | 100,890 (87.4) | 5,280 (91.7) |
| | | LGB+ | 971,235 (4.2) | 14,605 (12.6) | 480 (8.3) |
| | | Gay or Lesbian | 457,000 (2.0) | 5,425 (4.7) | 240 (4.2) |
| | | Bisexual | 403,390 (1.7) | 7,625 (6.6) | 185 (3.2) |
| | | Other | 110,845 (0.5) | 1,550 (1.3) | 60 (1.0) |
| | Rural | Total | 5,293,135 (100.0) | 19,545 (100.0) | 1,390 (100.0) |
| | | Heterosexual | 5,159,045 (97.5) | 17,470 (89.4) | 1,315 (94.6) |
| | | LGB+ | 134,090 (2.5) | 2,075 (10.6) | 75 (5.4) |
| | | Gay or Lesbian | 67,585 (1.3) | 765 (3.9) | 40 (2.9) |
| | | Bisexual | 54,130 (1.0) | 1,115 (5.7) | 25 (1.8) |
| | | Other | 12,370 (0.2) | 195 (1.0) | 5 (0.4) |
| Country of birth | Born in the UK | Total | 23,138,360 (100.0) | 119,820 (100.0) | 6,270 (100.0) |
| | | Heterosexual | 22,260,930 (96.2) | 104,645 (87.3) | 5,775 (92.1) |
| | | LGB+ | 877,430 (3.8) | 15,175 (12.7) | 495 (7.9) |
| | | Gay or Lesbian | 433,385 (1.9) | 5,710 (4.8) | 255 (4.1) |
| | | Bisexual | 360,775 (1.6) | 7,945 (6.6) | 180 (2.9) |
| | | Other | 83,265 (0.4) | 1,515 (1.3) | 55 (0.9) |
| | Born outside the UK | Total | 5,521,105 (100.0) | 15,220 (100.0) | 880 (100.0) |
| | | Heterosexual | 5,293,215 (95.9) | 13,715 (90.1) | 820 (93.2) |
| | | LGB+ | 227,895 (4.1) | 1,505 (9.9) | 65 (7.4) |
| | | Gay or Lesbian | 91,200 (1.7) | 480 (3.2) | 25 (2.8) |
| | | Bisexual | 96,745 (1.8) | 795 (5.2) | 30 (3.4) |
| | | Other | 39,950 (0.7) | 230 (1.5) | 10 (1.1) |
| English language proficiency | Main language is English | Total | 25,978,125 (100.0) | 128,585 (100.0) | 6,800 (100.0) |
| | | Heterosexual | 24,976,765 (96.1) | 112,410 (87.4) | 6,260 (92.1) |

| | | | | | |
|---|---|---|---|---|---|
| | | LGB+ | 1,001,360 (3.9) | 16,175 (12.6) | 540 (7.9) |
| | | Gay or Lesbian | 491,205 (1.9) | 6,055 (4.7) | 275 (4.0) |
| | | Bisexual | 411,395 (1.6) | 8,490 (6.6) | 205 (3.0) |
| | | Other | 98,760 (0.4) | 1,635 (1.3) | 60 (0.9) |
| | Main language is not English | Total | 2,681,340 (100.0) | 6,450 (100.0) | 350 (100.0) |
| | | Heterosexual | 2,577,380 (96.1) | 5,950 (92.2) | 335 (95.7) |
| | | LGB+ | 103,965 (3.9) | 500 (7.8) | 15 (4.3) |
| | | Gay or Lesbian | 33,380 (1.2) | 135 (2.1) | 5 (1.4) |
| | | Bisexual | 46,125 (1.7) | 250 (3.9) | 5 (1.4) |
| | | Other | 24,460 (0.9) | 115 (1.8) | 5 (1.4) |
| Religion | No religion | Total | 10,732,605 (100.0) | 68,720 (100.0) | 3,520 (100.0) |
| | | Heterosexual | 10,031,320 (93.5) | 57,380 (83.5) | 3,150 (89.5) |
| | | LGB+ | 701,285 (6.5) | 11,340 (16.5) | 370 (10.5) |
| | | Gay or Lesbian | 328,620 (3.1) | 4,120 (6.0) | c |
| | | Bisexual | 305,285 (2.8) | 6,115 (8.9) | c |
| | | Other | 67,380 (0.6) | 1,105 (1.6) | c |
| | Christian | Total | 14,299,140 (100.0) | 51,940 (100.0) | 2,955 (100.0) |
| | | Heterosexual | 14,022,860 (98.1) | 48,410 (93.2) | 2,830 (95.8) |
| | | LGB+ | 276,275 (1.9) | 3,525 (6.8) | 130 (4.4) |
| | | Gay or Lesbian | 157,940 (1.1) | 1,570 (3.0) | c |
| | | Bisexual | 91,305 (0.6) | 1,665 (3.2) | c |
| | | Other | 27,035 (0.2) | 290 (0.6) | c |
| | Buddhist | Total | 149,690 (100.0) | 690 (100.0) | 50 (100.0) |
| | | Heterosexual | 136,525 (91.2) | 495 (71.7) | 45 (90.0) |
| | | LGB+ | 13,165 (8.8) | 195 (28.3) | 10 (20.0) |
| | | Gay or Lesbian | 5,640 (3.8) | 60 (8.7) | c |
| | | Bisexual | 5,725 (3.8) | 100 (14.5) | c |
| | | Other | 1,800 (1.2) | 35 (5.1) | c |
| | Hindu | Total | 509,375 (100.0) | 1,230 (100.0) | 65 (100.0) |
| | | Heterosexual | 498,540 (97.9) | 1,165 (94.7) | 65 (100.0) |
| | | LGB+ | 10,835 (2.1) | 60 (4.9) | 5 (7.7) |
| | | Gay or Lesbian | 1,945 (0.4) | 15 (1.2) | c |
| | | Bisexual | 7,135 (1.4) | 40 (3.3) | c |
| | | Other | 1,750 (0.3) | 5 (0.4) | c |
| | Jewish | Total | 126,115 (100.0) | 560 (100.0) | 30 (100.0) |
| | | Heterosexual | 119,135 (94.5) | 470 (83.9) | 25 (83.3) |
| | | LGB+ | 6,975 (5.5) | 90 (16.1) | 5 (16.7) |
| | | Gay or Lesbian | 3,385 (2.7) | 30 (5.4) | c |
| | | Bisexual | 2,735 (2.2) | 50 (8.9) | c |
| | | Other | 860 (0.7) | 10 (1.8) | c |
| | Muslim | Total | 1,509,160 (100.0) | 5,325 (100.0) | 85 (100.0) |
| | | Heterosexual | 1,482,735 (98.2) | 5,130 (96.3) | 80 (94.1) |
| | | LGB+ | 26,425 (1.8) | 190 (3.6) | 5 (5.9) |
| | | Gay or Lesbian | 3,975 (0.3) | 45 (0.8) | c |
| | | Bisexual | 12,780 (0.8) | 100 (1.9) | c |
| | | Other | 9,670 (0.6) | 45 (0.8) | c |
| | Sikh | Total | 249,655 (100.0) | 820 (100.0) | 70 (100.0) |
| | | Heterosexual | 245,185 (98.2) | 785 (95.7) | 65 (92.9) |
| | | LGB+ | 4,475 (1.8) | 35 (4.3) | 5 (7.1) |
| | | Gay or Lesbian | 835 (0.3) | 10 (1.2) | c |
| | | Bisexual | 2,355 (0.9) | 10 (1.2) | c |
| | | Other | 1,285 (0.5) | 10 (1.2) | c |
| | Other religion | Total | 201,225 (100.0) | 1,735 (100.0) | 95 (100.0) |
| | | Heterosexual | 165,730 (82.4) | 970 (55.9) | 75 (78.9) |
| | | LGB+ | 35,495 (17.6) | 765 (44.1) | 20 (21.1) |
| | | Gay or Lesbian | 8,455 (4.2) | 170 (9.8) | c |
| | | Bisexual | 17,955 (8.9) | 410 (23.6) | c |
| | | Other | 9,090 (4.5) | 190 (11.0) | c |
| | Not answered | Total | 882,505 (100.0) | 4,025 (100.0) | 275 (100.0) |
| | | Heterosexual | 852,110 (96.6) | 3,550 (88.2) | 260 (94.5) |
| | | LGB+ | 30,395 (3.4) | 475 (11.8) | 15 (5.5) |
| | | Gay or Lesbian | 13,790 (1.6) | 175 (4.3) | c |
| | | Bisexual | 12,255 (1.4) | 245 (6.1) | c |
| | | Other | 4,350 (0.5) | 55 (1.4) | c |
| Household NS-SEC | Class 1 (Higher managerial, administrative and professional occupations) | Total | 7,224,420 (100.0) | 17,125 (100.0) | 1,375 (100.0) |
| | | Heterosexual | 6,936,510 (96.0) | 14,925 (87.2) | 1,265 (92.0) |
| | | LGB+ | 287,910 (4.0) | 2,205 (12.9) | 110 (8.0) |
| | | Gay or Lesbian | 157,010 (2.2) | 840 (4.9) | c |
| | | Bisexual | 104,825 (1.5) | 1,100 (6.4) | c |
| | | Other | 26,075 (0.4) | 265 (1.5) | c |
| | | Total | 7,994,445 (100.0) | 28,115 (100.0) | 1,635 (100.0) |

| | | | | | |
|---|---|---|---|---|---|
| | Class 2 (Lower managerial, administrative and professional occupations) | Heterosexual | 7,669,120 (95.9) | 24,560 (87.4) | 1,495 (91.4) |
| | | LGB+ | 325,325 (4.1) | 3,550 (12.6) | 140 (8.6) |
| | | Gay or Lesbian | 173,375 (2.2) | 1,445 (5.1) | c |
| | | Bisexual | 121,205 (1.5) | 1,720 (6.1) | c |
| | | Other | 30,745 (0.4) | 390 (1.4) | c |
| | Class 3 (Intermediate occupations) | Total | 3,551,725 (100.0) | 16,550 (100.0) | 810 (100.0) |
| | | Heterosexual | 3,443,335 (96.9) | 14,690 (88.8) | 755 (93.2) |
| | | LGB+ | 108,390 (3.1) | 1,860 (11.2) | 55 (6.8) |
| | | Gay or Lesbian | 51,290 (1.4) | 775 (4.7) | c |
| | | Bisexual | 45,360 (1.3) | 900 (5.4) | c |
| | | Other | 11,740 (0.3) | 180 (1.1) | c |
| | Class 4 (Small employers and own account workers) | Total | 2,794,170 (100.0) | 12,815 (100.0) | 920 (100.0) |
| | | Heterosexual | 2,722,625 (97.4) | 11,935 (93.1) | 880 (95.7) |
| | | LGB+ | 71,545 (2.6) | 880 (6.9) | 35 (3.8) |
| | | Gay or Lesbian | 30,595 (1.1) | 335 (2.6) | c |
| | | Bisexual | 28,725 (1.0) | 430 (3.4) | c |
| | | Other | 12,225 (0.4) | 110 (0.9) | c |
| | Class 5 (Lower supervisory and technical occupations) | Total | 1,326,605 (100.0) | 7,555 (100.0) | 400 (100.0) |
| | | Heterosexual | 1,284,420 (96.8) | 6,790 (89.9) | 375 (93.8) |
| | | LGB+ | 42,185 (3.2) | 765 (10.1) | 25 (6.2) |
| | | Gay or Lesbian | 18,910 (1.4) | 315 (4.2) | c |
| | | Bisexual | 18,045 (1.4) | 400 (5.3) | c |
| | | Other | 5,230 (0.4) | 50 (0.7) | c |
| | Class 6 (Semi-routine occupations) | Total | 2,258,230 (100.0) | 18,110 (100.0) | 675 (100.0) |
| | | Heterosexual | 2,175,160 (96.3) | 15,880 (87.7) | 615 (91.1) |
| | | LGB+ | 83,070 (3.7) | 2,230 (12.3) | 65 (9.6) |
| | | Gay or Lesbian | 34,365 (1.5) | 855 (4.7) | c |
| | | Bisexual | 38,565 (1.7) | 1,155 (6.4) | c |
| | | Other | 10,140 (0.4) | 220 (1.2) | c |
| | Class 7 (Routine occupations) | Total | 1,663,240 (100.0) | 13,945 (100.0) | 670 (100.0) |
| | | Heterosexual | 1,605,255 (96.5) | 12,450 (89.3) | 615 (91.8) |
| | | LGB+ | 57,990 (3.5) | 1,495 (10.7) | 55 (8.2) |
| | | Gay or Lesbian | 21,570 (1.3) | 520 (3.7) | c |
| | | Bisexual | 27,295 (1.6) | 825 (5.9) | c |
| | | Other | 9,120 (0.5) | 150 (1.1) | c |
| | Class 8 (Never worked and long-term unemployed) | Total | 1,178,230 (100.0) | 14,230 (100.0) | 485 (100.0) |
| | | Heterosexual | 1,138,610 (96.6) | 12,665 (89.0) | 445 (91.8) |
| | | LGB+ | 39,625 (3.4) | 1,565 (11.0) | 40 (8.2) |
| | | Gay or Lesbian | 15,185 (1.3) | 610 (4.3) | c |
| | | Bisexual | 17,290 (1.5) | 815 (5.7) | c |
| | | Other | 7,150 (0.6) | 140 (1.0) | c |
| | Full-time students | Total | 348,600 (100.0) | 3,155 (100.0) | 80 (100.0) |
| | | Heterosexual | 295,750 (84.8) | 2,050 (65.0) | 60 (75.0) |
| | | LGB+ | 52,845 (15.2) | 1,105 (35.0) | 20 (25.0) |
| | | Gay or Lesbian | 13,075 (3.8) | 275 (8.7) | c |
| | | Bisexual | 33,185 (9.5) | 695 (22.0) | c |
| | | Other | 6,585 (1.9) | 135 (4.3) | c |
| | Living in a communal establishment | Total | 319,805 (100.0) | 3,435 (100.0) | 100 (100.0) |
| | | Heterosexual | 283,355 (88.6) | 2,410 (70.2) | 85 (85.0) |
| | | LGB+ | 36,445 (11.4) | 1,025 (29.8) | 15 (15.0) |
| | | Gay or Lesbian | 9,215 (2.9) | 225 (6.6) | c |
| | | Bisexual | 23,020 (7.2) | 695 (20.2) | c |
| | | Other | 4,210 (1.3) | 105 (3.1) | c |
| Lone person household indicator | Lives alone | Total | 4,600,305 (100.0) | 35,250 (100.0) | 2,555 (100.0) |
| | | Heterosexual | 4,363,790 (94.9) | 30,480 (86.5) | 2,315 (90.6) |
| | | LGB+ | 236,515 (5.1) | 4,770 (13.5) | 245 (9.6) |
| | | Gay or Lesbian | 134,830 (2.9) | 2,115 (6.0) | c |
| | | Bisexual | 78,515 (1.7) | 2,175 (6.2) | c |
| | | Other | 23,170 (0.5) | 475 (1.3) | c |
| | Does not live alone | Total | 23,739,360 (100.0) | 96,350 (100.0) | 4,495 (100.0) |
| | | Heterosexual | 22,906,995 (96.5) | 85,470 (88.7) | 4,200 (93.4) |
| | | LGB+ | 832,365 (3.5) | 10,885 (11.3) | 300 (6.7) |
| | | Gay or Lesbian | 380,540 (1.6) | 3,850 (4.0) | c |
| | | Bisexual | 355,985 (1.5) | 5,870 (6.1) | c |
| | | Other | 95,840 (0.4) | 1,165 (1.2) | c |
| | Living in a communal establishment | Total | 319,805 (100.0) | 3,435 (100.0) | 100 (100.0) |
| | | Heterosexual | 283,355 (88.6) | 2,410 (70.2) | 85 (85.0) |
| | | LGB+ | 36,445 (11.4) | 1,025 (29.8) | 15 (15.0) |
| | | Gay or Lesbian | 9,215 (2.9) | 225 (6.6) | c |
| | | Bisexual | 23,020 (7.2) | 695 (20.2) | c |

| | | | | | |
|---|---|---|---|---|---|
| | | Other | 4,210 (1.3) | 105 (3.1) | c |
| Household size | One person in household | Total | 4,600,305 (100.0) | 35,250 (100.0) | 2,555 (100.0) |
| | | Heterosexual | 4,363,790 (94.9) | 30,480 (86.5) | 2,315 (90.6) |
| | | LGB+ | 236,515 (5.1) | 4,770 (13.5) | 245 (9.6) |
| | | Gay or Lesbian | 134,830 (2.9) | 2,115 (6.0) | c |
| | | Bisexual | 78,515 (1.7) | 2,175 (6.2) | c |
| | | Other | 23,170 (0.5) | 475 (1.3) | c |
| | Two people in household | Total | 10,142,025 (100.0) | 36,480 (100.0) | 2,105 (100.0) |
| | | Heterosexual | 9,726,660 (95.9) | 32,095 (88.0) | 1,950 (92.6) |
| | | LGB+ | 415,365 (4.1) | 4,385 (12.0) | 160 (7.6) |
| | | Gay or Lesbian | 239,535 (2.4) | 1,830 (5.0) | c |
| | | Bisexual | 141,265 (1.4) | 2,140 (5.9) | c |
| | | Other | 34,560 (0.3) | 420 (1.2) | c |
| | Three to five people in household | Total | 12,340,900 (100.0) | 53,305 (100.0) | 2,215 (100.0) |
| | | Heterosexual | 11,964,450 (96.9) | 47,495 (89.1) | 2,085 (94.1) |
| | | LGB+ | 376,450 (3.1) | 5,810 (10.9) | 135 (6.1) |
| | | Gay or Lesbian | 131,220 (1.1) | 1,840 (3.5) | c |
| | | Bisexual | 192,115 (1.6) | 3,310 (6.2) | c |
| | | Other | 53,115 (0.4) | 660 (1.2) | c |
| | Six or more people in household | Total | 1,256,435 (100.0) | 6,565 (100.0) | 175 (100.0) |
| | | Heterosexual | 1,215,885 (96.8) | 5,880 (89.6) | 165 (94.3) |
| | | LGB+ | 40,550 (3.2) | 685 (10.4) | 5 (2.9) |
| | | Gay or Lesbian | 9,785 (0.8) | 180 (2.7) | c |
| | | Bisexual | 22,600 (1.8) | 415 (6.3) | c |
| | | Other | 8,160 (0.6) | 85 (1.3) | c |
| | Living in a communal establishment | Total | 319,805 (100.0) | 3,435 (100.0) | 100 (100.0) |
| | | Heterosexual | 283,355 (88.6) | 2,410 (70.2) | 85 (85.0) |
| | | LGB+ | 36,445 (11.4) | 1,025 (29.8) | 15 (15.0) |
| | | Gay or Lesbian | 9,215 (2.9) | 225 (6.6) | c |
| | | Bisexual | 23,020 (7.2) | 695 (20.2) | c |
| | | Other | 4,210 (1.3) | 105 (3.1) | c |
| Household tenure | Owned or shared ownership | Total | 18,590,085 (100.0) | 50,600 (100.0) | 3,820 (100.0) |
| | | Heterosexual | 18,092,870 (97.3) | 46,395 (91.7) | 3,625 (94.9) |
| | | LGB+ | 497,215 (2.7) | 4,205 (8.3) | 195 (5.1) |
| | | Gay or Lesbian | 284,235 (1.5) | 1,725 (3.4) | c |
| | | Bisexual | 167,880 (0.9) | 1,990 (3.9) | c |
| | | Other | 45,100 (0.2) | 490 (1.0) | c |
| | Social rented | Total | 4,048,710 (100.0) | 44,390 (100.0) | 1,500 (100.0) |
| | | Heterosexual | 3,888,600 (96.0) | 39,245 (88.4) | 1,370 (91.3) |
| | | LGB+ | 160,110 (4.0) | 5,145 (11.6) | 130 (8.7) |
| | | Gay or Lesbian | 69,005 (1.7) | 2,020 (4.6) | c |
| | | Bisexual | 71,845 (1.8) | 2,680 (6.0) | c |
| | | Other | 19,260 (0.5) | 445 (1.0) | c |
| | Private rented or living rent free | Total | 5,700,870 (100.0) | 36,615 (100.0) | 1,735 (100.0) |
| | | Heterosexual | 5,289,315 (92.8) | 30,310 (82.8) | 1,515 (87.3) |
| | | LGB+ | 411,550 (7.2) | 6,305 (17.2) | 215 (12.4) |
| | | Gay or Lesbian | 162,125 (2.8) | 2,225 (6.1) | c |
| | | Bisexual | 194,775 (3.4) | 3,375 (9.2) | c |
| | | Other | 54,650 (1.0) | 705 (1.9) | c |
| | Living in a communal establishment | Total | 319,805 (100.0) | 3,435 (100.0) | 100 (100.0) |
| | | Heterosexual | 283,355 (88.6) | 2,410 (70.2) | 85 (85.0) |
| | | LGB+ | 36,445 (11.4) | 1,025 (29.8) | 15 (15.0) |
| | | Gay or Lesbian | 9,215 (2.9) | 225 (6.6) | c |
| | | Bisexual | 23,020 (7.2) | 695 (20.2) | c |
| | | Other | 4,210 (1.3) | 105 (3.1) | c |
| Economic activity | Economically active | Total | 18,575,200 (100.0) | 69,280 (100.0) | 4,085 (100.0) |
| | | Heterosexual | 17,769,215 (95.7) | 60,675 (87.6) | 3,735 (91.4) |
| | | LGB+ | 805,985 (4.3) | 8,605 (12.4) | 345 (8.4) |
| | | Gay or Lesbian | 406,400 (2.2) | 3,400 (4.9) | 180 (4.4) |
| | | Bisexual | 317,585 (1.7) | 4,345 (6.3) | 125 (3.1) |
| | | Other | 81,995 (0.4) | 855 (1.2) | 40 (1.0) |
| | Economically inactive | Total | 10,084,270 (100.0) | 65,760 (100.0) | 3,065 (100.0) |
| | | Heterosexual | 9,784,925 (97.0) | 57,685 (87.7) | 2,860 (93.3) |
| | | LGB+ | 299,340 (3.0) | 8,075 (12.3) | 210 (6.9) |
| | | Gay or Lesbian | 118,185 (1.2) | 2,790 (4.2) | 105 (3.4) |
| | | Bisexual | 139,935 (1.4) | 4,395 (6.7) | 85 (2.8) |
| | | Other | 41,225 (0.4) | 890 (1.4) | 20 (0.7) |
| General health (aggregated) | Good health | Total | 23,169,210 (100.0) | 80,175 (100.0) | 4,495 (100.0) |
| | | Heterosexual | 22,302,530 (96.3) | 71,160 (88.8) | 4,180 (93.0) |
| | | LGB+ | 866,680 (3.7) | 9,020 (11.3) | 315 (7.0) |
| | | Gay or Lesbian | 427,600 (1.8) | 3,525 (4.4) | 185 (4.1) |

| | | | | | |
|---|---|---|---|---|---|
| | | Bisexual | 349,125 (1.5) | 4,705 (5.9) | 105 (2.3) |
| | | Other | 89,960 (0.4) | 785 (1.0) | 30 (0.7) |
| | Not good health | Total | 5,490,255 (100.0) | 54,860 (100.0) | 2,655 (100.0) |
| | | Heterosexual | 5,251,610 (95.7) | 47,200 (86.0) | 2,415 (91.0) |
| | | LGB+ | 238,645 (4.3) | 7,660 (14.0) | 240 (9.0) |
| | | Gay or Lesbian | 96,985 (1.8) | 2,665 (4.9) | 95 (3.6) |
| | | Bisexual | 108,395 (2.0) | 4,035 (7.4) | 110 (4.1) |
| | | Other | 33,260 (0.6) | 960 (1.7) | 35 (1.3) |
| General health (detailed) | Very good health | Total | 11,919,820 (100.0) | 33,480 (100.0) | 1,890 (100.0) |
| | | Heterosexual | 11,498,565 (96.5) | 30,370 (90.7) | 1,760 (93.1) |
| | | LGB+ | 421,255 (3.5) | 3,110 (9.3) | 130 (6.9) |
| | | Gay or Lesbian | 224,690 (1.9) | 1,380 (4.1) | c |
| | | Bisexual | 156,015 (1.3) | 1,500 (4.5) | c |
| | | Other | 40,545 (0.3) | 230 (0.7) | c |
| | Good health | Total | 11,249,390 (100.0) | 46,695 (100.0) | 2,605 (100.0) |
| | | Heterosexual | 10,803,965 (96.0) | 40,785 (87.3) | 2,420 (92.9) |
| | | LGB+ | 445,425 (4.0) | 5,905 (12.6) | 185 (7.1) |
| | | Gay or Lesbian | 202,910 (1.8) | 2,150 (4.6) | c |
| | | Bisexual | 193,105 (1.7) | 3,205 (6.9) | c |
| | | Other | 49,415 (0.4) | 555 (1.2) | c |
| | Fair health | Total | 4,008,345 (100.0) | 32,685 (100.0) | 1,565 (100.0) |
| | | Heterosexual | 3,840,595 (95.8) | 27,980 (85.6) | 1,425 (91.1) |
| | | LGB+ | 167,750 (4.2) | 4,705 (14.4) | 145 (9.3) |
| | | Gay or Lesbian | 67,995 (1.7) | 1,605 (4.9) | c |
| | | Bisexual | 77,840 (1.9) | 2,565 (7.8) | c |
| | | Other | 21,915 (0.5) | 535 (1.6) | c |
| | Bad health | Total | 1,175,695 (100.0) | 16,770 (100.0) | 790 (100.0) |
| | | Heterosexual | 1,119,055 (95.2) | 14,505 (86.5) | 720 (91.1) |
| | | LGB+ | 56,640 (4.8) | 2,265 (13.5) | 70 (8.9) |
| | | Gay or Lesbian | 22,750 (1.9) | 790 (4.7) | c |
| | | Bisexual | 25,130 (2.1) | 1,150 (6.9) | c |
| | | Other | 8,760 (0.7) | 320 (1.9) | c |
| | Very bad health | Total | 306,215 (100.0) | 5,405 (100.0) | 300 (100.0) |
| | | Heterosexual | 291,960 (95.3) | 4,715 (87.2) | 270 (90.0) |
| | | LGB+ | 14,255 (4.7) | 690 (12.8) | 25 (8.3) |
| | | Gay or Lesbian | 6,240 (2.0) | 270 (5.0) | c |
| | | Bisexual | 5,425 (1.8) | 315 (5.8) | c |
| | | Other | 2,585 (0.8) | 105 (1.9) | c |
| Disability | Day-to-day activities limited a lot | Total | 1,976,155 (100.0) | 31,200 (100.0) | 1,350 (100.0) |
| | | Heterosexual | 1,876,155 (94.9) | 26,310 (84.3) | 1,205 (89.3) |
| | | LGB+ | 100,005 (5.1) | 4,895 (15.7) | 145 (10.7) |
| | | Gay or Lesbian | 40,150 (2.0) | 1,685 (5.4) | 60 (4.4) |
| | | Bisexual | 44,795 (2.3) | 2,550 (8.2) | 65 (4.8) |
| | | Other | 15,060 (0.8) | 660 (2.1) | 20 (1.5) |
| | Day-to-day activities limited a little | Total | 3,226,275 (100.0) | 32,840 (100.0) | 1,400 (100.0) |
| | | Heterosexual | 3,002,375 (93.1) | 26,930 (82.0) | 1,250 (89.3) |
| | | LGB+ | 223,905 (6.9) | 5,910 (18.0) | 155 (11.1) |
| | | Gay or Lesbian | 78,960 (2.4) | 1,920 (5.8) | 70 (5.0) |
| | | Bisexual | 115,035 (3.6) | 3,360 (10.2) | 65 (4.6) |
| | | Other | 29,905 (0.9) | 630 (1.9) | 20 (1.4) |
| | Day-to-day activities not limited | Total | 23,457,035 (100.0) | 70,995 (100.0) | 4,400 (100.0) |
| | | Heterosexual | 22,675,615 (96.7) | 65,120 (91.7) | 4,145 (94.2) |
| | | LGB+ | 781,420 (3.3) | 5,875 (8.3) | 260 (5.9) |
| | | Gay or Lesbian | 405,475 (1.7) | 2,585 (3.6) | 150 (3.4) |
| | | Bisexual | 297,690 (1.3) | 2,830 (4.0) | 85 (1.9) |
| | | Other | 78,255 (0.3) | 460 (0.6) | 25 (0.6) |
| Residence type | Living in a private household | Total | 28,339,665 (100.0) | 131,605 (100.0) | 7,050 (100.0) |
| | | Heterosexual | 27,270,785 (96.2) | 115,950 (88.1) | 6,510 (92.3) |
| | | LGB+ | 1,068,880 (3.8) | 15,655 (11.9) | 540 (7.7) |
| | | Gay or Lesbian | 515,370 (1.8) | 5,965 (4.5) | c |
| | | Bisexual | 434,500 (1.5) | 8,045 (6.1) | c |
| | | Other | 119,005 (0.4) | 1,640 (1.2) | c |
| | Living in a communal establishment | Total | 319,805 (100.0) | 3,435 (100.0) | 100 (100.0) |
| | | Heterosexual | 283,355 (88.6) | 2,410 (70.2) | 85 (85.0) |
| | | LGB+ | 36,445 (11.4) | 1,025 (29.8) | 15 (15.0) |
| | | Gay or Lesbian | 9,215 (2.9) | 225 (6.6) | c |
| | | Bisexual | 23,020 (7.2) | 695 (20.2) | c |
| | | Other | 4,210 (1.3) | 105 (3.1) | c |

Note: Cells with the value 'c' have been suppressed for the purposes of statistical disclosure control.

**Supplementary Table 6: Crude and age-standardised rates of self-harm per 100,000 people by region, rural/urban classification, country of birth, English language proficiency, religion, household NS-SEC, lone person household indicator, household size, household tenure, economic activity, general health, disability, and residence type**

| Characteristic | Category | Sexual orientation | Crude rate of self-harm [95% CI] | Age-standardised rate of self-harm [95% CI] |
|---|---|---|---|---|
| Region | England: North East | Total | 617.39 [604.11, 630.89] | 944.93 [921.52, 968.34] |
| | | Heterosexual | 567.08 [554.13, 580.26] | 866.82 [843.52, 890.12] |
| | | LGB+ | 1,996.80 [1,870.84, 2,129.00] | 1,938.67 [1,814.01, 2,063.32] |
| | | Gay or Lesbian | 1,795.90 [1,629.20, 1,975.03] | 2,019.85 [1,822.33, 2,217.36] |
| | | Bisexual | 2,216.94 [2,009.97, 2,439.45] | 1,983.61 [1,774.05, 2,193.18] |
| | | Other | 2,135.23 [1,716.98, 2,624.56] | 2,031.35 [1,626.28, 2,505.26] |
| | England: North West | Total | 450.22 [443.37, 457.15] | 648.35 [636.83, 659.87] |
| | | Heterosexual | 405.84 [399.21, 412.55] | 570.94 [559.78, 582.11] |
| | | LGB+ | 1,580.76 [1,515.16, 1,648.46] | 1,570.34 [1,504.51, 1,636.17] |
| | | Gay or Lesbian | 1,334.36 [1,250.85, 1,421.99] | 1,549.41 [1,446.31, 1,652.51] |
| | | Bisexual | 1,978.82 [1,861.74, 2,101.33] | 1,751.55 [1,639.62, 1,863.48] |
| | | Other | 1,324.75 [1,135.74, 1,536.23] | 1,224.84 [1,041.15, 1,408.52] |
| | England: Yorkshire and The Humber | Total | 692.06 [682.04, 702.19] | 902.37 [886.96, 917.77] |
| | | Heterosexual | 646.18 [636.31, 656.16] | 829.30 [814.00, 844.60] |
| | | LGB+ | 1,874.20 [1,789.54, 1,961.84] | 1,800.85 [1,718.06, 1,883.64] |
| | | Gay or Lesbian | 1,583.71 [1,468.55, 1,705.51] | 1,767.74 [1,633.45, 1,902.02] |
| | | Bisexual | 2,203.57 [2,065.29, 2,348.66] | 1,945.78 [1,810.87, 2,080.69] |
| | | Other | 1,714.03 [1,478.96, 1,975.84] | 1,586.41 [1,356.44, 1,816.37] |
| | England: East Midlands | Total | 531.29 [521.89, 540.81] | 777.47 [761.81, 793.13] |
| | | Heterosexual | 481.94 [472.83, 491.17] | 696.04 [680.74, 711.33] |
| | | LGB+ | 1,917.70 [1,822.19, 2,016.93] | 1,832.74 [1,739.77, 1,925.71] |
| | | Gay or Lesbian | 1,524.83 [1,397.17, 1,661.02] | 1,780.99 [1,624.97, 1,937.01] |
| | | Bisexual | 2,350.46 [2,193.71, 2,515.44] | 2,034.70 [1,888.03, 2,181.38] |
| | | Other | 1,707.46 [1,445.15, 2,003.61] | 1,510.46 [1,264.07, 1,756.85] |
| | England: West Midlands | Total | 386.62 [379.22, 394.13] | 554.62 [542.50, 566.75] |
| | | Heterosexual | 351.47 [344.30, 358.75] | 496.04 [484.27, 507.82] |
| | | LGB+ | 1,470.76 [1,390.03, 1,554.96] | 1,425.53 [1,346.02, 1,505.03] |
| | | Gay or Lesbian | 1,214.63 [1,107.28, 1,329.57] | 1,422.96 [1,290.39, 1,555.52] |
| | | Bisexual | 1,831.66 [1,694.05, 1,977.46] | 1,565.95 [1,440.87, 1,691.03] |
| | | Other | 1,166.92 [963.41, 1,400.71] | 1,085.39 [884.12, 1,286.66] |
| | England: East of England | Total | 479.54 [471.88, 487.28] | 707.28 [693.54, 721.02] |
| | | Heterosexual | 439.22 [431.78, 446.76] | 630.43 [617.05, 643.80] |
| | | LGB+ | 1,705.49 [1,625.25, 1,788.66] | 1,697.17 [1,616.26, 1,778.09] |
| | | Gay or Lesbian | 1,303.71 [1,199.31, 1,414.76] | 1,575.17 [1,441.22, 1,709.11] |
| | | Bisexual | 2,139.41 [2,004.10, 2,281.45] | 1,898.28 [1,771.53, 2,025.02] |
| | | Other | 1,608.81 [1,388.17, 1,854.54] | 1,546.23 [1,324.43, 1,768.04] |
| | England: London | Total | 382.65 [376.84, 388.52] | 467.33 [459.16, 475.50] |
| | | Heterosexual | 356.37 [350.62, 362.20] | 425.43 [417.40, 433.46] |
| | | LGB+ | 846.05 [809.05, 884.31] | 979.64 [934.43, 1,024.85] |
| | | Gay or Lesbian | 619.69 [575.84, 665.99] | 895.43 [815.35, 975.51] |
| | | Bisexual | 1,137.11 [1,066.12, 1,211.58] | 1,092.34 [1,020.86, 1,163.83] |
| | | Other | 953.04 [843.72, 1,072.60] | 1,007.99 [885.14, 1,130.85] |
| | England: South East | Total | 440.89 [434.87, 446.97] | 685.90 [674.74, 697.07] |
| | | Heterosexual | 396.79 [390.98, 402.68] | 601.20 [590.36, 612.04] |
| | | LGB+ | 1,587.07 [1,528.09, 1,647.75] | 1,588.26 [1,528.52, 1,648.01] |
| | | Gay or Lesbian | 1,123.32 [1,051.12, 1,199.17] | 1,444.07 [1,344.95, 1,543.20] |
| | | Bisexual | 2,094.74 [1,991.08, 2,202.40] | 1,774.69 [1,681.71, 1,867.67] |
| | | Other | 1,591.51 [1,416.49, 1,782.18] | 1,461.96 [1,294.54, 1,629.38] |
| | England: South West | Total | 466.18 [458.16, 474.30] | 752.22 [737.37, 767.07] |
| | | Heterosexual | 410.81 [403.13, 418.60] | 654.77 [640.32, 669.23] |
| | | LGB+ | 1,833.41 [1,753.34, 1,916.20] | 1,776.40 [1,697.44, 1,855.36] |
| | | Gay or Lesbian | 1,441.42 [1,333.94, 1,555.26] | 1,793.57 [1,653.66, 1,933.49] |
| | | Bisexual | 2,191.70 [2,063.21, 2,326.09] | 1,886.39 [1,766.69, 2,006.09] |
| | | Other | 1,863.62 [1,625.48, 2,126.83] | 1,675.30 [1,448.98, 1,901.62] |
| | Wales | Total | 390.69 [379.64, 401.98] | 581.46 [563.17, 599.74] |
| | | Heterosexual | 354.81 [344.07, 365.80] | 529.56 [511.40, 547.71] |
| | | LGB+ | 1,261.73 [1,163.18, 1,366.40] | 1,192.58 [1,097.03, 1,288.14] |
| | | Gay or Lesbian | 1,082.87 [951.51, 1,227.30] | 1,252.49 [1,090.58, 1,414.40] |
| | | Bisexual | 1,554.53 [1,388.85, 1,734.53] | 1,321.33 [1,162.73, 1,479.92] |
| | | Other | 842.99 [602.25, 1,147.92] | 731.29 [521.15, 997.46] |
| Rural/urban classification | Urban | Total | 494.28 [491.43, 497.14] | 674.38 [669.92, 678.83] |
| | | Heterosexual | 450.50 [447.72, 453.29] | 603.43 [599.07, 607.79] |

| | | | | |
|---|---|---|---|---|
| | | LGB+ | 1,503.76 [1,479.47, 1,528.34] | 1,494.03 [1,469.79, 1,518.27] |
| | | Gay or Lesbian | 1,187.09 [1,155.71, 1,219.11] | 1,441.53 [1,401.11, 1,481.95] |
| | | Bisexual | 1,890.23 [1,848.04, 1,933.14] | 1,648.41 [1,609.35, 1,687.48] |
| | | Other | 1,398.35 [1,329.59, 1,469.74] | 1,324.72 [1,258.14, 1,391.30] |
| | Rural | Total | 369.25 [364.09, 374.47] | 646.66 [635.19, 658.14] |
| | | Heterosexual | 338.63 [333.63, 343.69] | 577.07 [565.92, 588.22] |
| | | LGB+ | 1,547.47 [1,481.59, 1,615.52] | 1,642.43 [1,570.25, 1,714.61] |
| | | Gay or Lesbian | 1,131.91 [1,053.11, 1,215.04] | 1,546.46 [1,428.47, 1,664.45] |
| | | Bisexual | 2,059.86 [1,940.71, 2,184.40] | 1,843.78 [1,731.29, 1,956.26] |
| | | Other | 1,576.39 [1,362.89, 1,813.86] | 1,488.82 [1,275.04, 1,702.61] |
| Country of birth | Born in the UK | Total | 517.84 [514.91, 520.78] | 765.88 [760.87, 770.89] |
| | | Heterosexual | 470.08 [467.24, 472.94] | 684.43 [679.52, 689.34] |
| | | LGB+ | 1,729.48 [1,702.07, 1,757.22] | 1,705.39 [1,678.20, 1,732.57] |
| | | Gay or Lesbian | 1,317.54 [1,283.58, 1,352.16] | 1,602.13 [1,558.83, 1,645.43] |
| | | Bisexual | 2,202.20 [2,154.04, 2,251.17] | 1,915.00 [1,869.95, 1,960.05] |
| | | Other | 1,819.49 [1,729.01, 1,913.48] | 1,642.46 [1,557.68, 1,727.25] |
| | Born outside the UK | Total | 275.67 [271.31, 280.08] | 339.68 [333.29, 346.08] |
| | | Heterosexual | 259.11 [254.79, 263.48] | 311.27 [305.00, 317.54] |
| | | LGB+ | 660.39 [627.45, 694.62] | 735.34 [697.07, 773.62] |
| | | Gay or Lesbian | 526.32 [480.28, 575.58] | 701.68 [626.48, 776.89] |
| | | Bisexual | 821.75 [765.61, 880.91] | 795.88 [739.26, 852.51] |
| | | Other | 575.72 [503.71, 655.13] | 650.75 [564.20, 737.29] |
| English language proficiency | Main language is English | Total | 494.97 [492.27, 497.69] | 722.65 [718.07, 727.23] |
| | | Heterosexual | 450.06 [447.43, 452.70] | 645.60 [641.11, 650.09] |
| | | LGB+ | 1,615.30 [1,590.50, 1,640.39] | 1,609.96 [1,585.14, 1,634.78] |
| | | Gay or Lesbian | 1,232.68 [1,201.83, 1,264.13] | 1,522.76 [1,482.46, 1,563.07] |
| | | Bisexual | 2,063.71 [2,020.04, 2,108.08] | 1,796.60 [1,756.11, 1,837.09] |
| | | Other | 1,655.53 [1,576.25, 1,737.77] | 1,512.84 [1,438.32, 1,587.35] |
| | Main language is not English | Total | 240.55 [234.72, 246.49] | 278.62 [270.74, 286.50] |
| | | Heterosexual | 230.85 [225.03, 236.80] | 263.00 [255.18, 270.83] |
| | | LGB+ | 480.93 [439.69, 524.99] | 536.24 [486.58, 585.89] |
| | | Gay or Lesbian | 404.43 [339.09, 478.70] | 580.64 [461.81, 699.48] |
| | | Bisexual | 542.01 [476.90, 613.52] | 545.76 [476.20, 615.33] |
| | | Other | 470.16 [388.16, 564.35] | 504.71 [406.28, 603.14] |
| Religion | No religion | Total | 640.29 [635.51, 645.10] | 797.98 [791.58, 804.38] |
| | | Heterosexual | 572.01 [567.34, 576.71] | 711.15 [704.78, 717.53] |
| | | LGB+ | 1,617.03 [1,587.41, 1,647.07] | 1,486.95 [1,459.28, 1,514.62] |
| | | Gay or Lesbian | 1,253.73 [1,215.73, 1,292.61] | 1,409.56 [1,365.39, 1,453.73] |
| | | Bisexual | 2,003.05 [1,953.15, 2,053.89] | 1,664.66 [1,617.59, 1,711.74] |
| | | Other | 1,639.95 [1,544.67, 1,739.57] | 1,407.47 [1,319.90, 1,495.04] |
| | Christian | Total | 363.24 [360.12, 366.38] | 577.91 [571.10, 584.71] |
| | | Heterosexual | 345.22 [342.15, 348.31] | 534.57 [527.92, 541.22] |
| | | LGB+ | 1,275.90 [1,234.13, 1,318.73] | 1,579.60 [1,524.66, 1,634.54] |
| | | Gay or Lesbian | 994.05 [945.48, 1,044.46] | 1,521.83 [1,431.59, 1,612.07] |
| | | Bisexual | 1,823.56 [1,737.01, 1,913.30] | 1,847.57 [1,758.34, 1,936.80] |
| | | Other | 1,072.68 [952.77, 1,203.52] | 1,169.39 [1,033.07, 1,305.72] |
| | Buddhist | Total | 460.95 [427.20, 496.67] | 615.39 [560.08, 670.71] |
| | | Heterosexual | 362.57 [331.33, 395.96] | 442.11 [393.18, 491.03] |
| | | LGB+ | 1,481.20 [1,280.59, 1,704.32] | 1,789.44 [1,522.92, 2,055.97] |
| | | Gay or Lesbian | 1,063.83 [811.81, 1,369.36] | 1,416.08 [1,004.54, 1,915.01] |
| | | Bisexual | 1,746.72 [1,421.21, 2,124.49] | 1,883.83 [1,510.16, 2,257.50] |
| | | Other | 1,944.44 [1,354.38, 2,704.25] | 1,998.98 [1,371.42, 2,807.08] |
| | Hindu | Total | 241.47 [228.16, 255.35] | 298.76 [279.15, 318.38] |
| | | Heterosexual | 233.68 [220.45, 247.50] | 287.37 [267.91, 306.83] |
| | | LGB+ | 553.76 [422.58, 712.80] | 688.92 [519.88, 893.87] |
| | | Gay or Lesbian | 771.21 [431.64, 1,271.99] | 910.61 [426.43, 1,624.65] |
| | | Bisexual | 560.62 [400.51, 763.40] | 696.73 [493.31, 954.37] |
| | | Other | 285.71 [92.77, 666.76] | 524.28 [198.57, 1,167.53] |
| | Jewish | Total | 444.04 [408.02, 482.39] | 595.15 [527.38, 662.92] |
| | | Heterosexual | 394.51 [359.65, 431.84] | 467.96 [404.86, 531.06] |
| | | LGB+ | 1,290.32 [1,037.57, 1,586.03] | 1,433.35 [1,149.41, 1,765.55] |
| | | Gay or Lesbian | 886.26 [597.96, 1,265.20] | 1,189.85 [768.24, 1,743.98] |
| | | Bisexual | 1,828.15 [1,356.89, 2,410.19] | 1,711.37 [1,260.44, 2,268.30] |
| | | Other | 1,162.79 [557.60, 2,138.41] | 1,182.54 [538.04, 2,221.55] |
| | Muslim | Total | 352.85 [343.43, 362.45] | 386.83 [375.86, 397.79] |
| | | Heterosexual | 345.98 [336.58, 355.58] | 379.60 [368.65, 390.56] |
| | | LGB+ | 719.02 [620.41, 828.84] | 864.22 [734.66, 993.79] |
| | | Gay or Lesbian | 1,132.08 [825.74, 1,514.81] | 1,309.97 [921.50, 1,795.34] |
| | | Bisexual | 782.47 [636.65, 951.70] | 903.39 [719.30, 1,087.49] |
| | | Other | 465.36 [339.43, 622.68] | 619.25 [438.31, 845.32] |

| | | | | |
|---|---|---|---|---|
| | Sikh | Total | 328.45 [306.35, 351.73] | 371.14 [342.07, 400.20] |
| | | Heterosexual | 320.17 [298.16, 343.37] | 360.41 [331.48, 389.35] |
| | | LGB+ | 782.12 [544.78, 1,087.74] | 793.17 [530.15, 1,131.85] |
| | | Gay or Lesbian | 1,197.60 [574.30, 2,202.44] | 1,508.04 [713.63, 2,788.71] |
| | | Bisexual | 424.63 [203.63, 780.91] | 487.49 [244.28, 879.56] |
| | | Other | 778.21 [373.18, 1,431.16] | 959.24 [390.16, 1,876.67] |
| | Other religion | Total | 862.22 [822.12, 903.77] | 1,217.79 [1,153.07, 1,282.50] |
| | | Heterosexual | 585.29 [549.03, 623.31] | 749.43 [688.95, 809.92] |
| | | LGB+ | 2,155.23 [2,005.19, 2,313.53] | 2,100.50 [1,950.79, 2,250.21] |
| | | Gay or Lesbian | 2,010.64 [1,719.75, 2,336.65] | 2,223.87 [1,879.25, 2,568.49] |
| | | Bisexual | 2,283.49 [2,067.78, 2,515.58] | 2,112.44 [1,903.08, 2,321.80] |
| | | Other | 2,090.21 [1,803.55, 2,409.48] | 1,981.65 [1,695.83, 2,267.47] |
| | Not answered | Total | 456.09 [442.11, 470.40] | 636.07 [613.19, 658.95] |
| | | Heterosexual | 416.61 [403.02, 430.55] | 569.90 [547.62, 592.17] |
| | | LGB+ | 1,562.76 [1,425.36, 1,709.82] | 1,573.57 [1,431.95, 1,715.19] |
| | | Gay or Lesbian | 1,269.04 [1,087.98, 1,471.61] | 1,520.05 [1,278.74, 1,761.36] |
| | | Bisexual | 1,999.18 [1,756.67, 2,265.82] | 1,789.59 [1,554.76, 2,024.41] |
| | | Other | 1,264.37 [952.49, 1,645.75] | 1,281.58 [967.44, 1,665.73] |
| Household NS-SEC | Class 1 (Higher managerial, administrative and professional occupations) | Total | 237.04 [233.51, 240.62] | 363.69 [356.70, 370.68] |
| | | Heterosexual | 215.17 [211.73, 218.65] | 316.51 [309.78, 323.24] |
| | | LGB+ | 765.86 [734.23, 798.51] | 884.13 [846.00, 922.27] |
| | | Gay or Lesbian | 535.00 [499.43, 572.43] | 807.51 [742.55, 872.47] |
| | | Bisexual | 1,049.37 [988.26, 1,113.26] | 977.39 [917.90, 1,036.89] |
| | | Other | 1,016.30 [897.61, 1,146.32] | 962.84 [844.33, 1,081.36] |
| | Class 2 (Lower managerial, administrative and professional occupations) | Total | 351.68 [347.58, 355.82] | 513.04 [505.77, 520.31] |
| | | Heterosexual | 320.25 [316.25, 324.28] | 455.47 [448.39, 462.55] |
| | | LGB+ | 1,091.22 [1,055.61, 1,127.72] | 1,203.02 [1,162.61, 1,243.43] |
| | | Gay or Lesbian | 833.45 [791.03, 877.56] | 1,123.92 [1,057.65, 1,190.19] |
| | | Bisexual | 1,419.08 [1,352.80, 1,487.77] | 1,317.02 [1,253.22, 1,380.82] |
| | | Other | 1,268.50 [1,145.71, 1,400.86] | 1,238.79 [1,113.03, 1,364.55] |
| | Class 3 (Intermediate occupations) | Total | 465.97 [458.90, 473.12] | 684.73 [672.63, 696.82] |
| | | Heterosexual | 426.62 [419.75, 433.58] | 616.93 [605.12, 628.75] |
| | | LGB+ | 1,716.03 [1,638.92, 1,795.82] | 1,708.21 [1,630.29, 1,786.12] |
| | | Gay or Lesbian | 1,511.02 [1,406.49, 1,621.25] | 1,728.88 [1,603.48, 1,854.28] |
| | | Bisexual | 1,984.13 [1,856.60, 2,118.11] | 1,772.12 [1,652.07, 1,892.17] |
| | | Other | 1,533.22 [1,317.41, 1,774.30] | 1,459.42 [1,245.17, 1,673.68] |
| | Class 4 (Small employers and own account workers) | Total | 458.63 [450.73, 466.64] | 659.12 [644.95, 673.28] |
| | | Heterosexual | 438.36 [430.53, 446.30] | 622.95 [608.88, 637.01] |
| | | LGB+ | 1,230.00 [1,150.06, 1,314.02] | 1,396.90 [1,300.97, 1,492.82] |
| | | Gay or Lesbian | 1,094.95 [980.82, 1,218.71] | 1,509.46 [1,331.69, 1,687.22] |
| | | Bisexual | 1,496.95 [1,358.79, 1,645.36] | 1,494.75 [1,350.56, 1,638.93] |
| | | Other | 899.80 [739.52, 1,084.50] | 1,020.88 [826.03, 1,215.74] |
| | Class 5 (Lower supervisory and technical occupations) | Total | 569.50 [556.73, 582.49] | 785.77 [765.17, 806.38] |
| | | Heterosexual | 528.64 [516.14, 541.37] | 718.60 [698.39, 738.81] |
| | | LGB+ | 1,813.44 [1,687.20, 1,946.63] | 1,860.73 [1,728.38, 1,993.09] |
| | | Gay or Lesbian | 1,665.79 [1,486.89, 1,860.28] | 1,933.94 [1,709.06, 2,158.82] |
| | | Bisexual | 2,216.68 [2,004.75, 2,444.92] | 2,057.67 [1,851.92, 2,263.42] |
| | | Other | 956.02 [709.58, 1,260.40] | 1,024.54 [761.22, 1,349.75] |
| | Class 6 (Semi-routine occupations) | Total | 801.96 [790.32, 813.72] | 1,153.48 [1,134.64, 1,172.31] |
| | | Heterosexual | 730.06 [718.75, 741.51] | 1,038.09 [1,019.65, 1,056.54] |
| | | LGB+ | 2,684.48 [2,574.21, 2,798.27] | 2,649.44 [2,539.27, 2,759.61] |
| | | Gay or Lesbian | 2,488.00 [2,324.00, 2,660.51] | 2,814.88 [2,623.71, 3,006.06] |
| | | Bisexual | 2,994.94 [2,824.69, 3,172.78] | 2,723.01 [2,561.22, 2,884.79] |
| | | Other | 2,169.63 [1,892.38, 2,476.06] | 2,137.56 [1,855.02, 2,420.09] |
| | Class 7 (Routine occupations) | Total | 838.42 [824.57, 852.46] | 1,173.04 [1,150.65, 1,195.42] |
| | | Heterosexual | 775.58 [762.01, 789.32] | 1,072.05 [1,050.00, 1,094.10] |
| | | LGB+ | 2,578.03 [2,448.99, 2,712.11] | 2,618.87 [2,485.68, 2,752.06] |
| | | Gay or Lesbian | 2,410.76 [2,207.98, 2,627.15] | 2,761.89 [2,517.71, 3,006.06] |
| | | Bisexual | 3,022.53 [2,819.77, 3,236.02] | 2,822.50 [2,626.48, 3,018.52] |
| | | Other | 1,644.74 [1,392.06, 1,930.01] | 1,696.24 [1,422.93, 1,969.55] |
| | Class 8 (Never worked and long-term unemployed) | Total | 1,207.74 [1,187.98, 1,227.75] | 2,191.39 [2,149.76, 2,233.03] |
| | | Heterosexual | 1,112.32 [1,093.03, 1,131.86] | 2,005.96 [1,964.83, 2,047.08] |
| | | LGB+ | 3,949.53 [3,756.25, 4,150.17] | 4,422.73 [4,202.14, 4,643.32] |
| | | Gay or Lesbian | 4,017.12 [3,704.62, 4,348.94] | 5,511.13 [5,050.80, 5,971.47] |
| | | Bisexual | 4,713.71 [4,395.60, 5,048.75] | 4,617.88 [4,298.77, 4,936.98] |
| | | Other | 1,958.04 [1,647.14, 2,310.57] | 2,245.73 [1,866.78, 2,624.67] |
| | Full-time students | Total | 905.05 [873.74, 937.19] | 789.27 [747.45, 831.08] |
| | | Heterosexual | 693.15 [663.47, 723.82] | 659.44 [618.43, 700.45] |
| | | LGB+ | 2,091.02 [1,969.53, 2,218.04] | 1,751.54 [1,487.21, 2,015.87] |
| | | Gay or Lesbian | 2,103.25 [1,861.99, 2,367.10] | 1,920.29 [1,447.94, 2,392.63] |

| | | | | |
|---|---|---|---|---|
| | | Bisexual | 2,094.32 [1,941.49, 2,255.98] | 1,555.40 [1,308.38, 1,802.41] |
| | | Other | 2,050.11 [1,718.89, 2,426.56] | 1,768.11 [1,007.43, 2,528.80] |
| | Living in a communal establishment | Total | 1,074.09 [1,038.47, 1,110.62] | 1,547.24 [1,464.96, 1,629.52] |
| | | Heterosexual | 850.52 [816.90, 885.17] | 1,376.18 [1,294.11, 1,458.25] |
| | | LGB+ | 2,812.46 [2,642.89, 2,990.04] | 3,133.73 [2,661.41, 3,606.06] |
| | | Gay or Lesbian | 2,441.67 [2,133.03, 2,782.42] | 2,481.32 [1,928.66, 3,033.98] |
| | | Bisexual | 3,019.11 [2,798.80, 3,252.16] | 4,727.74 [3,637.25, 5,818.23] |
| | | Other | 2,494.06 [2,039.90, 3,019.21] | 1,361.57 [868.38, 1,854.75] |
| Lone person household indicator | Lives alone | Total | 766.25 [758.28, 774.30] | 1,856.18 [1,817.62, 1,894.74] |
| | | Heterosexual | 698.48 [690.66, 706.36] | 1,613.46 [1,575.20, 1,651.72] |
| | | LGB+ | 2,016.79 [1,959.95, 2,074.85] | 3,329.42 [3,195.00, 3,463.85] |
| | | Gay or Lesbian | 1,568.64 [1,502.49, 1,636.95] | 3,402.67 [3,146.59, 3,658.76] |
| | | Bisexual | 2,770.17 [2,654.96, 2,889.09] | 3,590.41 [3,406.69, 3,774.12] |
| | | Other | 2,050.06 [1,869.83, 2,242.99] | 2,783.74 [2,434.19, 3,133.30] |
| | Does not live alone | Total | 405.87 [403.31, 408.44] | 572.75 [568.64, 576.87] |
| | | Heterosexual | 373.12 [370.62, 375.63] | 519.47 [515.43, 523.51] |
| | | LGB+ | 1,307.72 [1,283.27, 1,332.52] | 1,253.00 [1,229.40, 1,276.61] |
| | | Gay or Lesbian | 1,011.72 [980.01, 1,044.19] | 1,198.24 [1,159.07, 1,237.41] |
| | | Bisexual | 1,648.95 [1,607.03, 1,691.68] | 1,385.73 [1,347.88, 1,423.57] |
| | | Other | 1,215.57 [1,146.76, 1,287.43] | 1,095.83 [1,032.08, 1,159.57] |
| | Living in a communal establishment | Total | 1,074.09 [1,038.47, 1,110.62] | 1,547.24 [1,464.96, 1,629.52] |
| | | Heterosexual | 850.52 [816.90, 885.17] | 1,376.18 [1,294.11, 1,458.25] |
| | | LGB+ | 2,812.46 [2,642.89, 2,990.04] | 3,133.73 [2,661.41, 3,606.06] |
| | | Gay or Lesbian | 2,441.67 [2,133.03, 2,782.42] | 2,481.32 [1,928.66, 3,033.98] |
| | | Bisexual | 3,019.11 [2,798.80, 3,252.16] | 4,727.74 [3,637.25, 5,818.23] |
| | | Other | 2,494.06 [2,039.90, 3,019.21] | 1,361.57 [868.38, 1,854.75] |
| Household size | One person in household | Total | 766.25 [758.28, 774.30] | 1,856.18 [1,817.62, 1,894.74] |
| | | Heterosexual | 698.48 [690.66, 706.36] | 1,613.46 [1,575.20, 1,651.72] |
| | | LGB+ | 2,016.79 [1,959.95, 2,074.85] | 3,329.42 [3,195.00, 3,463.85] |
| | | Gay or Lesbian | 1,568.64 [1,502.49, 1,636.95] | 3,402.67 [3,146.59, 3,658.76] |
| | | Bisexual | 2,770.17 [2,654.96, 2,889.09] | 3,590.41 [3,406.69, 3,774.12] |
| | | Other | 2,050.06 [1,869.83, 2,242.99] | 2,783.74 [2,434.19, 3,133.30] |
| | Two people in household | Total | 359.69 [356.01, 363.40] | 713.97 [702.92, 725.02] |
| | | Heterosexual | 329.97 [326.37, 333.60] | 639.61 [628.59, 650.62] |
| | | LGB+ | 1,055.70 [1,024.68, 1,087.42] | 1,333.13 [1,287.15, 1,379.11] |
| | | Gay or Lesbian | 763.98 [729.37, 799.80] | 1,272.44 [1,189.09, 1,355.78] |
| | | Bisexual | 1,514.88 [1,451.37, 1,580.46] | 1,508.81 [1,438.46, 1,579.15] |
| | | Other | 1,215.28 [1,101.82, 1,337.25] | 1,339.91 [1,197.54, 1,482.28] |
| | Three to five people in household | Total | 431.94 [428.28, 435.62] | 547.58 [542.47, 552.69] |
| | | Heterosexual | 396.97 [393.41, 400.55] | 501.21 [496.18, 506.24] |
| | | LGB+ | 1,543.37 [1,503.93, 1,583.57] | 1,255.15 [1,219.84, 1,290.46] |
| | | Gay or Lesbian | 1,402.23 [1,338.88, 1,467.79] | 1,230.36 [1,171.65, 1,289.08] |
| | | Bisexual | 1,722.93 [1,664.73, 1,782.64] | 1,357.56 [1,302.50, 1,412.62] |
| | | Other | 1,242.59 [1,149.58, 1,341.11] | 1,001.23 [922.56, 1,079.91] |
| | Six or more people in household | Total | 522.51 [509.95, 535.31] | 530.17 [516.71, 543.63] |
| | | Heterosexual | 483.60 [471.32, 496.12] | 493.85 [480.56, 507.14] |
| | | LGB+ | 1,689.27 [1,565.12, 1,820.65] | 1,240.30 [1,132.69, 1,347.91] |
| | | Gay or Lesbian | 1,839.55 [1,580.62, 2,128.80] | 1,335.64 [1,115.15, 1,556.14] |
| | | Bisexual | 1,836.28 [1,663.84, 2,021.74] | 1,351.16 [1,195.20, 1,507.11] |
| | | Other | 1,041.67 [832.05, 1,288.04] | 885.45 [690.18, 1,114.95] |
| | Living in a communal establishment | Total | 1,074.09 [1,038.47, 1,110.62] | 1,547.24 [1,464.96, 1,629.52] |
| | | Heterosexual | 850.52 [816.90, 885.17] | 1,376.18 [1,294.11, 1,458.25] |
| | | LGB+ | 2,812.46 [2,642.89, 2,990.04] | 3,133.73 [2,661.41, 3,606.06] |
| | | Gay or Lesbian | 2,441.67 [2,133.03, 2,782.42] | 2,481.32 [1,928.66, 3,033.98] |
| | | Bisexual | 3,019.11 [2,798.80, 3,252.16] | 4,727.74 [3,637.25, 5,818.23] |
| | | Other | 2,494.06 [2,039.90, 3,019.21] | 1,361.57 [868.38, 1,854.75] |
| Household tenure | Owned or shared ownership | Total | 272.19 [269.82, 274.57] | 427.67 [422.66, 432.69] |
| | | Heterosexual | 256.43 [254.10, 258.77] | 390.56 [385.65, 395.48] |
| | | LGB+ | 845.71 [820.34, 871.67] | 976.14 [945.43, 1,006.85] |
| | | Gay or Lesbian | 606.89 [578.59, 636.22] | 914.03 [864.46, 963.61] |
| | | Bisexual | 1,185.37 [1,133.86, 1,238.62] | 1,057.39 [1,009.45, 1,105.33] |
| | | Other | 1,086.47 [992.39, 1,187.07] | 1,041.34 [947.25, 1,135.44] |
| | Social rented | Total | 1,096.40 [1,086.22, 1,106.65] | 1,371.89 [1,357.76, 1,386.02] |
| | | Heterosexual | 1,009.23 [999.27, 1,019.27] | 1,247.46 [1,233.64, 1,261.29] |
| | | LGB+ | 3,213.42 [3,126.20, 3,302.44] | 3,274.35 [3,184.50, 3,364.20] |
| | | Gay or Lesbian | 2,927.32 [2,801.05, 3,057.83] | 3,340.82 [3,190.99, 3,490.64] |
| | | Bisexual | 3,730.25 [3,590.35, 3,874.21] | 3,502.83 [3,368.37, 3,637.29] |
| | | Other | 2,310.49 [2,100.78, 2,535.47] | 2,393.90 [2,169.46, 2,618.35] |
| | Private rented or living rent free | Total | 642.27 [635.71, 648.88] | 716.75 [708.91, 724.59] |
| | | Heterosexual | 573.04 [566.61, 579.53] | 633.08 [625.41, 640.75] |

| | | | | |
|---|---|---|---|---|
| | | LGB+ | 1,532.01 [1,494.43, 1,570.30] | 1,486.32 [1,446.03, 1,526.61] |
| | | Gay or Lesbian | 1,372.40 [1,315.96, 1,430.63] | 1,525.61 [1,453.54, 1,597.67] |
| | | Bisexual | 1,732.77 [1,674.80, 1,792.23] | 1,548.89 [1,487.70, 1,610.07] |
| | | Other | 1,290.03 [1,196.55, 1,388.87] | 1,220.02 [1,121.06, 1,318.98] |
| | Living in a communal establishment | Total | 1,074.09 [1,038.47, 1,110.62] | 1,547.24 [1,464.96, 1,629.52] |
| | | Heterosexual | 850.52 [816.90, 885.17] | 1,376.18 [1,294.11, 1,458.25] |
| | | LGB+ | 2,812.46 [2,642.89, 2,990.04] | 3,133.73 [2,661.41, 3,606.06] |
| | | Gay or Lesbian | 2,441.67 [2,133.03, 2,782.42] | 2,481.32 [1,928.66, 3,033.98] |
| | | Bisexual | 3,019.11 [2,798.80, 3,252.16] | 4,727.74 [3,637.25, 5,818.23] |
| | | Other | 2,494.06 [2,039.90, 3,019.21] | 1,361.57 [868.38, 1,854.75] |
| Economic activity | Economically active | Total | 372.97 [370.20, 375.76] | 544.12 [539.15, 549.08] |
| | | Heterosexual | 341.46 [338.75, 344.19] | 488.13 [483.27, 492.99] |
| | | LGB+ | 1,067.64 [1,045.20, 1,090.44] | 1,183.83 [1,156.83, 1,210.83] |
| | | Gay or Lesbian | 836.61 [808.73, 865.22] | 1,131.79 [1,087.21, 1,176.37] |
| | | Bisexual | 1,368.14 [1,327.76, 1,409.43] | 1,268.65 [1,225.65, 1,311.66] |
| | | Other | 1,042.75 [974.01, 1,115.05] | 1,114.36 [1,034.36, 1,194.36] |
| | Economically inactive | Total | 652.10 [647.13, 657.11] | 1,317.90 [1,305.42, 1,330.38] |
| | | Heterosexual | 589.53 [584.73, 594.36] | 1,182.23 [1,169.97, 1,194.49] |
| | | LGB+ | 2,697.60 [2,639.08, 2,757.09] | 2,969.78 [2,898.02, 3,041.55] |
| | | Gay or Lesbian | 2,360.71 [2,273.91, 2,449.96] | 3,363.79 [3,226.58, 3,501.01] |
| | | Bisexual | 3,140.74 [3,048.57, 3,235.00] | 3,108.25 [3,001.00, 3,215.51] |
| | | Other | 2,158.88 [2,019.36, 2,305.51] | 2,070.12 [1,923.57, 2,216.66] |
| General health (aggregated) | Good health | Total | 1,017.89 [1,006.91, 1,028.96] | 1,872.68 [1,849.20, 1,896.16] |
| | | Heterosexual | 896.96 [886.28, 907.73] | 1,736.90 [1,711.30, 1,762.49] |
| | | LGB+ | 728.53 [720.02, 737.12] | 1,691.66 [1,663.02, 1,720.29] |
| | | Gay or Lesbian | 824.37 [797.38, 852.04] | 1,020.26 [985.13, 1,055.40] |
| | | Bisexual | 1,347.65 [1,309.42, 1,386.72] | 1,077.21 [1,044.36, 1,110.05] |
| | | Other | 872.61 [812.63, 935.85] | 790.44 [734.39, 846.48] |
| | Not good health | Total | 1,325.70 [1,292.10, 1,359.95] | 1,297.25 [1,264.14, 1,330.35] |
| | | Heterosexual | 1,040.75 [1,019.38, 1,062.46] | 1,008.32 [987.49, 1,029.16] |
| | | LGB+ | 751.84 [732.73, 771.31] | 767.35 [747.70, 787.00] |
| | | Gay or Lesbian | 2,747.85 [2,644.50, 2,854.20] | 3,957.97 [3,792.90, 4,123.04] |
| | | Bisexual | 3,722.50 [3,608.51, 3,839.16] | 3,612.82 [3,500.61, 3,725.03] |
| | | Other | 2,886.35 [2,706.63, 3,074.87] | 2,904.68 [2,720.80, 3,088.55] |
| General health (detailed) | Very good health | Total | 4,840.41 [4,485.93, 5,215.45] | 6,672.96 [6,091.39, 7,254.54] |
| | | Heterosexual | 4,894.76 [4,758.58, 5,033.83] | 5,331.55 [5,180.73, 5,482.36] |
| | | LGB+ | 3,998.94 [3,835.93, 4,167.10] | 4,786.28 [4,578.53, 4,994.03] |
| | | Gay or Lesbian | 614.18 [582.20, 647.46] | 745.78 [704.81, 786.75] |
| | | Bisexual | 961.45 [913.40, 1,011.36] | 749.66 [708.23, 791.09] |
| | | Other | 567.27 [496.32, 645.52] | 529.40 [459.32, 599.49] |
| | Good health | Total | 1,765.10 [1,718.35, 1,812.80] | 4,317.84 [4,121.46, 4,514.23] |
| | | Heterosexual | 1,614.95 [1,569.18, 1,661.71] | 3,819.14 [3,617.70, 4,020.58] |
| | | LGB+ | 3,209.79 [3,138.31, 3,282.49] | 3,556.67 [3,476.13, 3,637.22] |
| | | Gay or Lesbian | 1,059.58 [1,015.26, 1,105.34] | 1,354.69 [1,294.71, 1,414.68] |
| | | Bisexual | 1,659.72 [1,602.75, 1,718.19] | 1,348.37 [1,299.11, 1,397.62] |
| | | Other | 1,123.14 [1,031.63, 1,220.59] | 1,005.34 [920.80, 1,089.88] |
| | Fair health | Total | 1,578.82 [1,561.35, 1,596.44] | 3,508.28 [3,456.53, 3,560.02] |
| | | Heterosexual | 1,426.39 [1,404.88, 1,448.14] | 3,372.97 [3,293.04, 3,452.90] |
| | | LGB+ | 1,402.34 [1,385.44, 1,419.39] | 3,104.64 [3,050.75, 3,158.54] |
| | | Gay or Lesbian | 2,360.47 [2,246.39, 2,478.84] | 3,373.56 [3,197.49, 3,549.63] |
| | | Bisexual | 3,295.22 [3,168.92, 3,425.27] | 3,039.83 [2,919.92, 3,159.74] |
| | | Other | 2,441.25 [2,238.74, 2,657.16] | 2,318.02 [2,120.18, 2,515.85] |
| | Bad health | Total | 1,296.18 [1,275.17, 1,317.45] | 3,030.76 [2,945.32, 3,116.20] |
| | | Heterosexual | 2,804.77 [2,725.19, 2,886.08] | 2,990.21 [2,904.39, 3,076.03] |
| | | LGB+ | 2,639.51 [2,572.64, 2,707.68] | 2,445.07 [2,381.46, 2,508.69] |
| | | Gay or Lesbian | 3,472.53 [3,234.57, 3,723.36] | 5,248.26 [4,819.32, 5,677.20] |
| | | Bisexual | 4,576.20 [4,315.51, 4,848.53] | 4,873.96 [4,586.68, 5,161.23] |
| | | Other | 3,652.97 [3,263.65, 4,075.95] | 3,983.68 [3,537.16, 4,430.19] |
| | Very bad health | Total | 999.22 [990.88, 1,007.62] | 2,264.23 [2,237.65, 2,290.80] |
| | | Heterosexual | 898.77 [890.68, 906.92] | 2,011.16 [1,983.56, 2,038.75] |
| | | LGB+ | 815.42 [806.61, 824.31] | 1,902.98 [1,875.35, 1,930.62] |
| | | Gay or Lesbian | 4,326.92 [3,826.15, 4,875.02] | 7,446.11 [6,253.36, 8,638.86] |
| | | Bisexual | 5,806.45 [5,182.87, 6,484.39] | 6,978.05 [6,133.88, 7,822.22] |
| | | Other | 4,061.90 [3,322.23, 4,917.17] | 5,137.62 [4,056.50, 6,218.74] |
| Disability | Day-to-day activities limited a lot | Total | 415.09 [411.33, 418.87] | 717.48 [709.73, 725.24] |
| | | Heterosexual | 738.27 [712.55, 764.68] | 709.60 [684.56, 734.64] |
| | | LGB+ | 377.50 [373.85, 381.18] | 648.99 [641.22, 656.77] |
| | | Gay or Lesbian | 4,196.76 [3,998.75, 4,402.04] | 5,744.51 [5,449.02, 6,039.99] |
| | | Bisexual | 5,692.60 [5,473.77, 5,917.93] | 5,485.75 [5,271.45, 5,700.06] |
| | | Other | 4,382.47 [4,054.46, 4,729.95] | 4,350.61 [4,017.70, 4,683.53] |

| | | | | |
|---|---|---|---|---|
| | Day-to-day activities limited a little | Total | 346.04 [343.65, 348.44] | 492.56 [488.77, 496.36] |
| | | Heterosexual | 319.07 [316.73, 321.42] | 451.67 [447.92, 455.42] |
| | | LGB+ | 302.66 [300.44, 304.89] | 423.06 [419.50, 426.61] |
| | | Gay or Lesbian | 2,431.61 [2,324.05, 2,542.87] | 2,838.33 [2,709.58, 2,967.08] |
| | | Bisexual | 2,920.85 [2,822.91, 3,021.32] | 2,492.36 [2,399.71, 2,585.01] |
| | | Other | 2,106.67 [1,945.36, 2,277.79] | 1,799.41 [1,652.05, 1,946.77] |
| | Day-to-day activities not limited | Total | 287.18 [284.98, 289.40] | 400.03 [396.47, 403.59] |
| | | Heterosexual | 280.88 [277.88, 283.90] | 357.25 [353.19, 361.32] |
| | | LGB+ | 264.12 [261.16, 267.11] | 336.45 [332.41, 340.50] |
| | | Gay or Lesbian | 637.52 [613.18, 662.58] | 805.89 [772.83, 838.94] |
| | | Bisexual | 950.65 [915.95, 986.34] | 797.16 [766.48, 827.84] |
| | | Other | 587.82 [535.32, 644.08] | 566.69 [514.42, 618.97] |
| Residence type | Living in a private household | Total | 464.38 [461.88, 466.90] | 666.77 [662.54, 671.00] |
| | | Heterosexual | 425.18 [422.74, 427.63] | 598.25 [594.11, 602.39] |
| | | LGB+ | 1,464.62 [1,441.76, 1,487.74] | 1,515.25 [1,491.43, 1,539.07] |
| | | Gay or Lesbian | 1,157.42 [1,128.23, 1,187.17] | 1,468.58 [1,428.58, 1,508.58] |
| | | Bisexual | 1,851.55 [1,811.31, 1,892.46] | 1,668.17 [1,630.46, 1,705.87] |
| | | Other | 1,378.09 [1,312.20, 1,446.44] | 1,339.67 [1,274.37, 1,404.97] |
| | Living in a communal establishment | Total | 1,074.09 [1,038.47, 1,110.62] | 1,547.24 [1,464.96, 1,629.52] |
| | | Heterosexual | 850.52 [816.90, 885.17] | 1,376.18 [1,294.11, 1,458.25] |
| | | LGB+ | 2,812.46 [2,642.89, 2,990.04] | 3,133.73 [2,661.41, 3,606.06] |
| | | Gay or Lesbian | 2,441.67 [2,133.03, 2,782.42] | 2,481.32 [1,928.66, 3,033.98] |
| | | Bisexual | 3,019.11 [2,798.80, 3,252.16] | 4,727.74 [3,637.25, 5,818.23] |
| | | Other | 2,494.06 [2,039.90, 3,019.21] | 1,361.57 [868.38, 1,854.75] |

Notes: Age-standardised rates are expressed per 100,000 people and standardised to the age distribution of the observed study population identifying with an LGB+ orientation in Census 2021. All calculations are based on estimates rounded to the nearest multiple of five for disclosure control reasons. CI: confidence interval.

**Supplementary Table 7: Crude and age-standardised rates of suicide per 100,000 people by region, rural/urban classification, country of birth, English language proficiency, religion, household NS-SEC, lone person household indicator, household size, household tenure, economic activity, general health (detailed), disability, and residence type**

| Characteristic | Category | Sexual orientation | Crude rate of suicide [95% CI] | Age-standardised rate of suicide [95% CI] |
|---|---|---|---|---|
| Region | England: North East | Total | 31.56 [28.62, 34.73] | 35.90 [31.68, 40.12] |
| | | Heterosexual | 30.38 [27.44, 33.55] | 34.42 [30.11, 38.74] |
| | | LGB+ | 74.75 [52.06, 103.95] | 71.93 [49.35, 101.00] |
| | England: North West | Total | 29.10 [27.38, 30.91] | 30.05 [27.86, 32.25] |
| | | Heterosexual | 27.41 [25.70, 29.19] | 27.44 [25.29, 29.59] |
| | | LGB+ | 72.35 [58.86, 87.99] | 72.99 [58.74, 87.24] |
| | England: Yorkshire and The Humber | Total | 28.35 [26.35, 30.46] | 28.39 [25.93, 30.86] |
| | | Heterosexual | 27.08 [25.09, 29.19] | 26.82 [24.34, 29.29] |
| | | LGB+ | 56.02 [42.20, 72.92] | 60.13 [45.55, 77.95] |
| | England: East Midlands | Total | 26.41 [24.35, 28.60] | 25.75 [23.25, 28.26] |
| | | Heterosexual | 25.09 [23.05, 27.27] | 23.61 [21.17, 26.05] |
| | | LGB+ | 63.50 [47.13, 83.72] | 61.09 [44.96, 81.01] |
| | England: West Midlands | Total | 25.14 [23.28, 27.11] | 24.10 [21.91, 26.29] |
| | | Heterosexual | 24.80 [22.93, 26.79] | 23.88 [21.64, 26.13] |
| | | LGB+ | 35.58 [24.01, 50.80] | 39.31 [26.85, 55.67] |
| | England: East of England | Total | 21.78 [20.18, 23.48] | 21.61 [19.48, 23.73] |
| | | Heterosexual | 20.84 [19.25, 22.54] | 20.19 [18.09, 22.29] |
| | | LGB+ | 45.28 [33.03, 60.59] | 47.24 [34.73, 62.87] |
| | England: London | Total | 16.63 [15.44, 17.89] | 16.09 [14.73, 17.45] |
| | | Heterosexual | 15.62 [14.43, 16.88] | 14.90 [13.55, 16.26] |
| | | LGB+ | 34.44 [27.31, 42.87] | 34.48 [27.22, 43.06] |
| | England: South East | Total | 25.31 [23.88, 26.80] | 24.67 [22.83, 26.51] |
| | | Heterosexual | 24.49 [23.06, 25.99] | 23.51 [21.64, 25.38] |
| | | LGB+ | 46.51 [36.88, 57.88] | 46.44 [36.79, 57.84] |
| | England: South West | Total | 26.99 [25.08, 28.99] | 26.85 [24.39, 29.30] |
| | | Heterosexual | 26.38 [24.46, 28.41] | 26.02 [23.48, 28.55] |
| | | LGB+ | 46.53 [34.54, 61.35] | 46.60 [34.32, 61.76] |
| | Wales | Total | 25.99 [23.20, 29.03] | 25.02 [21.68, 28.37] |
| | | Heterosexual | 24.48 [21.72, 27.50] | 22.99 [19.72, 26.26] |
| | | LGB+ | 62.57 [42.21, 89.32] | 61.69 [41.08, 88.78] |
| Rural/urban classification | Urban | Total | 24.65 [24.02, 25.30] | 24.33 [23.58, 25.08] |
| | | Heterosexual | 23.58 [22.94, 24.22] | 22.91 [22.16, 23.66] |
| | | LGB+ | 49.42 [45.10, 54.05] | 49.79 [45.34, 54.24] |
| | | Gay or Lesbian | 52.52 [46.08, 59.60] | 49.11 [42.59, 55.62] |
| | | Bisexual | 45.86 [39.49, 52.97] | 50.25 [42.49, 58.01] |
| | | Other | 54.13 [41.31, 69.68] | 54.87 [41.48, 71.10] |
| | Rural | Total | 26.26 [24.90, 27.68] | 25.53 [23.60, 27.46] |
| | | Heterosexual | 25.49 [24.13, 26.91] | 24.20 [22.27, 26.12] |
| | | LGB+ | 55.93 [43.99, 70.11] | 54.41 [42.41, 68.67] |
| | | Gay or Lesbian | 59.18 [42.28, 80.59] | 63.47 [44.32, 87.73] |
| | | Bisexual | 46.19 [29.89, 68.18] | 52.26 [33.98, 76.93] |
| | | Other | 40.42 [13.12, 94.33] | 44.34 [14.66, 102.96] |
| Country of birth | Born in the UK | Total | 27.10 [26.43, 27.78] | 27.35 [26.51, 28.19] |
| | | Heterosexual | 25.94 [25.28, 26.62] | 25.73 [24.88, 26.57] |
| | | LGB+ | 56.41 [51.55, 61.61] | 56.73 [51.72, 61.75] |
| | | Gay or Lesbian | 58.84 [51.84, 66.52] | 55.98 [48.82, 63.14] |
| | | Bisexual | 49.89 [42.87, 57.74] | 56.34 [47.56, 65.12] |
| | | Other | 66.05 [49.76, 85.98] | 71.57 [53.59, 93.57] |
| | Born outside the UK | Total | 15.94 [14.90, 17.03] | 15.66 [14.43, 16.89] |
| | | Heterosexual | 15.49 [14.45, 16.59] | 14.97 [13.73, 16.20] |
| | | LGB+ | 28.52 [22.01, 36.35] | 29.07 [22.12, 37.44] |
| | | Gay or Lesbian | 27.41 [17.74, 40.47] | 30.70 [17.91, 47.96] |
| | | Bisexual | 31.01 [20.92, 44.27] | 33.12 [21.81, 47.98] |
| | | Other | 25.03 [12.00, 46.03] | 23.49 [9.69, 45.72] |
| English language proficiency | Main language is English | Total | 26.18 [25.56, 26.81] | 26.23 [25.45, 27.00] |
| | | Heterosexual | 25.06 [24.45, 25.69] | 24.67 [23.89, 25.45] |
| | | LGB+ | 53.93 [49.47, 58.67] | 53.89 [49.34, 58.44] |
| | | Gay or Lesbian | 55.98 [49.56, 63.01] | 53.23 [46.59, 59.87] |
| | | Bisexual | 49.83 [43.24, 57.14] | 55.43 [47.38, 63.49] |
| | | Other | 60.75 [46.36, 78.20] | 62.89 [47.52, 81.52] |
| | Main language is not English | Total | 13.05 [11.72, 14.49] | 12.29 [10.85, 13.74] |
| | | Heterosexual | 13.00 [11.64, 14.47] | 12.12 [10.65, 13.60] |
| | | LGB+ | 14.43 [8.08, 23.80] | 17.45 [9.57, 29.06] |
| | | Gay or Lesbian | 14.98 [4.86, 34.96] | 17.24 [5.43, 40.57] |

| | | | | |
|---|---|---|---|---|
| | | Bisexual | 10.84 [3.52, 25.30] | 12.10 [3.25, 29.59] |
| | | Other | 20.44 [6.64, 47.70] | 25.57 [8.43, 59.41] |
| Religion | No religion | Total | 32.80 [31.72, 33.90] | 31.13 [30.00, 32.25] |
| | | Heterosexual | 31.40 [30.31, 32.52] | 29.41 [28.27, 30.56] |
| | | LGB+ | 52.76 [47.52, 58.42] | 55.96 [50.15, 61.78] |
| | Christian | Total | 20.67 [19.93, 21.42] | 20.34 [19.25, 21.44] |
| | | Heterosexual | 20.18 [19.44, 20.94] | 19.56 [18.47, 20.65] |
| | | LGB+ | 47.05 [39.31, 55.87] | 43.62 [35.42, 51.82] |
| | Buddhist | Total | 33.40 [24.79, 44.04] | 27.85 [19.23, 38.50] |
| | | Heterosexual | 32.96 [24.04, 44.10] | 23.54 [15.44, 33.67] |
| | | LGB+ | 75.96 [36.43, 139.69] | 62.49 [27.01, 119.68] |
| | Hindu | Total | 12.76 [9.85, 16.26] | 13.16 [9.68, 17.35] |
| | | Heterosexual | 13.04 [10.06, 16.62] | 12.55 [9.15, 16.65] |
| | | LGB+ | 46.15 [14.98, 107.69] | 31.10 [2.83, 86.93] |
| | Jewish | Total | 23.79 [16.05, 33.96] | 18.39 [11.30, 27.72] |
| | | Heterosexual | 20.98 [13.58, 30.98] | 16.22 [9.64, 25.09] |
| | | LGB+ | 71.68 [23.28, 167.29] | 41.61 [5.08, 113.75] |
| | Muslim | Total | 5.63 [4.50, 6.96] | 5.64 [4.44, 7.06] |
| | | Heterosexual | 5.40 [4.28, 6.72] | 5.46 [4.27, 6.86] |
| | | LGB+ | 18.92 [6.14, 44.16] | 15.25 [2.11, 41.20] |
| | Sikh | Total | 28.04 [21.86, 35.43] | 25.52 [18.94, 33.40] |
| | | Heterosexual | 26.51 [20.46, 33.79] | 24.29 [17.89, 31.99] |
| | | LGB+ | 111.73 [36.28, 260.74] | 88.81 [12.25, 240.01] |
| | Other religion | Total | 47.21 [38.20, 57.71] | 48.52 [37.62, 61.21] |
| | | Heterosexual | 45.25 [35.60, 56.73] | 46.33 [33.19, 61.94] |
| | | LGB+ | 56.35 [34.42, 87.02] | 52.52 [31.65, 81.72] |
| | Not answered | Total | 31.16 [27.59, 35.07] | 31.44 [26.86, 36.02] |
| | | Heterosexual | 30.51 [26.92, 34.46] | 30.67 [25.99, 35.36] |
| | | LGB+ | 49.35 [27.62, 81.40] | 49.10 [27.39, 81.11] |
| Household NS-SEC | Class 1 (Higher managerial, administrative and professional occupations) | Total | 19.03 [18.04, 20.07] | 17.68 [16.42, 18.94] |
| | | Heterosexual | 18.24 [17.25, 19.27] | 16.51 [15.25, 17.76] |
| | | LGB+ | 38.21 [31.40, 46.05] | 37.70 [30.42, 44.98] |
| | Class 2 (Lower managerial, administrative and professional occupations) | Total | 20.45 [19.47, 21.47] | 19.48 [18.27, 20.69] |
| | | Heterosexual | 19.49 [18.52, 20.51] | 18.36 [17.13, 19.58] |
| | | LGB+ | 43.03 [36.20, 50.78] | 41.07 [34.21, 47.93] |
| | Class 3 (Intermediate occupations) | Total | 22.81 [21.26, 24.43] | 23.60 [21.60, 25.59] |
| | | Heterosexual | 21.93 [20.39, 23.55] | 22.60 [20.58, 24.61] |
| | | LGB+ | 50.74 [38.23, 66.05] | 50.35 [37.80, 65.69] |
| | Class 4 (Small employers and own account workers) | Total | 32.93 [30.83, 35.12] | 31.11 [28.44, 33.78] |
| | | Heterosexual | 32.32 [30.22, 34.53] | 30.44 [27.75, 33.13] |
| | | LGB+ | 48.92 [34.07, 68.04] | 47.95 [32.74, 67.54] |
| | Class 5 (Lower supervisory and technical occupations) | Total | 30.15 [27.27, 33.26] | 30.47 [26.83, 34.11] |
| | | Heterosexual | 29.20 [26.32, 32.31] | 29.03 [25.40, 32.66] |
| | | LGB+ | 59.26 [38.35, 87.48] | 61.68 [40.36, 90.46] |
| | Class 6 (Semi-routine occupations) | Total | 29.89 [27.68, 32.23] | 32.37 [29.47, 35.26] |
| | | Heterosexual | 28.27 [26.08, 30.60] | 30.06 [27.18, 32.94] |
| | | LGB+ | 78.25 [60.39, 99.73] | 75.96 [58.30, 97.20] |
| | Class 7 (Routine occupations) | Total | 40.28 [37.29, 43.45] | 42.40 [38.58, 46.21] |
| | | Heterosexual | 38.31 [35.34, 41.46] | 39.81 [36.00, 43.61] |
| | | LGB+ | 94.84 [71.45, 123.45] | 94.39 [70.97, 123.03] |
| | Class 8 (Never worked and long-term unemployed) | Total | 41.16 [37.58, 45.00] | 53.34 [47.31, 59.38] |
| | | Heterosexual | 39.08 [35.54, 42.89] | 49.45 [43.48, 55.42] |
| | | LGB+ | 100.95 [72.12, 137.46] | 105.87 [75.27, 144.62] |
| | Full-time students | Total | 22.95 [18.20, 28.56] | 28.19 [19.08, 38.96] |
| | | Heterosexual | 20.29 [15.48, 26.11] | 24.89 [16.81, 34.69] |
| | | LGB+ | 37.85 [23.12, 58.45] | 95.93 [.00, 263.93] |
| | Living in a communal establishment | Total | 31.27 [25.44, 38.03] | 61.99 [44.73, 79.24] |
| | | Heterosexual | 30.00 [23.96, 37.09] | 61.45 [44.48, 81.40] |
| | | LGB+ | 41.16 [23.04, 67.88] | 43.92 [8.90, 95.55] |
| Lone person household indicator | Lives alone | Total | 55.54 [53.41, 57.74] | 72.21 [66.52, 77.90] |
| | | Heterosexual | 53.05 [50.91, 55.26] | 66.12 [60.58, 71.67] |
| | | LGB+ | 103.59 [91.02, 117.40] | 115.88 [94.81, 136.95] |
| | Does not live alone | Total | 18.93 [18.39, 19.50] | 19.00 [18.33, 19.67] |
| | | Heterosexual | 18.34 [17.78, 18.90] | 18.16 [17.49, 18.83] |
| | | LGB+ | 36.04 [32.08, 40.36] | 36.29 [32.14, 40.44] |
| | Living in a communal establishment | Total | 31.27 [25.44, 38.03] | 61.99 [44.73, 79.24] |
| | | Heterosexual | 30.00 [23.96, 37.09] | 61.45 [44.48, 81.40] |
| | | LGB+ | 41.16 [23.04, 67.88] | 43.92 [8.90, 95.55] |

| | | | | |
|---|---|---|---|---|
| Household size | One person in household | Total | 55.54 [53.41, 57.74] | 72.21 [66.52, 77.90] |
| | | Heterosexual | 53.05 [50.91, 55.26] | 66.12 [60.58, 71.67] |
| | | LGB+ | 103.59 [91.02, 117.40] | 115.88 [94.81, 136.95] |
| | Two people in household | Total | 20.76 [19.88, 21.66] | 22.20 [20.78, 23.62] |
| | | Heterosexual | 20.05 [19.17, 20.96] | 21.14 [19.67, 22.61] |
| | | LGB+ | 38.52 [32.78, 44.97] | 35.64 [29.87, 41.42] |
| | Three to five people in household | Total | 17.95 [17.21, 18.71] | 18.34 [17.47, 19.20] |
| | | Heterosexual | 17.43 [16.69, 18.19] | 17.61 [16.74, 18.48] |
| | | LGB+ | 35.86 [30.07, 42.45] | 39.58 [31.27, 47.90] |
| | Six or more people in household | Total | 13.93 [11.94, 16.15] | 13.96 [11.77, 16.15] |
| | | Heterosexual | 13.57 [11.58, 15.81] | 13.80 [11.58, 16.02] |
| | | LGB+ | 12.33 [4.00, 28.78] | 18.73 [5.97, 43.94] |
| | Living in a communal establishment | Total | 31.27 [25.44, 38.03] | 61.99 [44.73, 79.24] |
| | | Heterosexual | 30.00 [23.96, 37.09] | 61.45 [44.48, 81.40] |
| | | LGB+ | 41.16 [23.04, 67.88] | 43.92 [8.90, 95.55] |
| Household tenure | Owned or shared ownership | Total | 20.55 [19.90, 21.21] | 19.37 [18.46, 20.28] |
| | | Heterosexual | 20.04 [19.39, 20.70] | 18.66 [17.74, 19.58] |
| | | LGB+ | 39.22 [33.91, 45.13] | 37.07 [31.60, 42.53] |
| | Social rented | Total | 37.05 [35.20, 38.97] | 36.98 [34.84, 39.11] |
| | | Heterosexual | 35.23 [33.39, 37.15] | 34.76 [32.64, 36.89] |
| | | LGB+ | 81.19 [67.84, 96.41] | 80.39 [66.48, 94.30] |
| | Private rented or living rent free | Total | 30.43 [29.02, 31.90] | 29.46 [28.02, 30.90] |
| | | Heterosexual | 28.64 [27.22, 30.12] | 27.30 [25.86, 28.73] |
| | | LGB+ | 52.24 [45.49, 59.71] | 60.26 [51.33, 69.19] |
| | Living in a communal establishment | Total | 31.27 [25.44, 38.03] | 61.99 [44.73, 79.24] |
| | | Heterosexual | 30.00 [23.96, 37.09] | 61.45 [44.48, 81.40] |
| | | LGB+ | 41.16 [23.04, 67.88] | 43.92 [8.90, 95.55] |
| Economic activity | Economically active | Total | 21.99 [21.32, 22.68] | 21.21 [20.41, 22.01] |
| | | Heterosexual | 21.02 [20.35, 21.70] | 20.09 [19.29, 20.90] |
| | | LGB+ | 42.80 [38.41, 47.57] | 43.35 [38.45, 48.25] |
| | | Gay or Lesbian | 44.29 [38.06, 51.26] | 43.97 [36.47, 51.47] |
| | | Bisexual | 39.36 [32.76, 46.90] | 45.02 [36.22, 53.82] |
| | | Other | 48.78 [34.85, 66.43] | 51.63 [35.81, 71.65] |
| | Economically inactive | Total | 30.39 [29.33, 31.49] | 41.70 [39.61, 43.79] |
| | | Heterosexual | 29.23 [28.17, 30.32] | 39.14 [37.05, 41.23] |
| | | LGB+ | 70.15 [60.99, 80.31] | 83.12 [70.89, 95.35] |
| | | Gay or Lesbian | 88.84 [72.67, 107.55] | 108.82 [84.40, 133.23] |
| | | Bisexual | 60.74 [48.52, 75.11] | 73.38 [56.79, 92.87] |
| | | Other | 48.51 [29.63, 74.93] | 56.08 [34.23, 86.65] |
| General health (aggregated) | Good health | Total | 19.40 [18.84, 19.98] | 19.28 [18.61, 19.94] |
| | | Heterosexual | 18.74 [18.18, 19.32] | 18.52 [17.85, 19.19] |
| | | LGB+ | 36.35 [32.44, 40.59] | 36.82 [32.74, 40.90] |
| | | Gay or Lesbian | 43.26 [37.25, 49.97] | 41.64 [35.40, 47.87] |
| | | Bisexual | 30.08 [24.60, 36.41] | 34.37 [27.06, 41.68] |
| | | Other | 33.35 [22.50, 47.61] | 33.23 [21.47, 48.68] |
| | Not good health | Total | 48.36 [46.54, 50.23] | 64.25 [60.39, 68.10] |
| | | Heterosexual | 45.99 [44.17, 47.86] | 59.29 [55.28, 63.31] |
| | | LGB+ | 100.57 [88.25, 114.13] | 101.36 [88.30, 114.41] |
| | | Gay or Lesbian | 97.95 [79.25, 119.74] | 96.62 [75.58, 121.13] |
| | | Bisexual | 101.48 [83.40, 122.31] | 99.83 [80.87, 118.80] |
| | | Other | 105.23 [73.30, 146.35] | 111.46 [78.46, 153.95] |
| General health (detailed) | Very good health | Total | 15.86 [15.15, 16.59] | 15.48 [14.71, 16.24] |
| | | Heterosexual | 15.31 [14.60, 16.04] | 14.87 [14.10, 15.63] |
| | | LGB+ | 30.86 [25.78, 36.64] | 30.69 [25.32, 36.05] |
| | Good health | Total | 23.16 [22.28, 24.06] | 24.72 [23.49, 25.95] |
| | | Heterosexual | 22.40 [21.52, 23.31] | 23.94 [22.66, 25.21] |
| | | LGB+ | 41.53 [35.76, 47.97] | 42.40 [36.32, 48.48] |
| | Fair health | Total | 39.04 [37.13, 41.03] | 52.48 [48.50, 56.47] |
| | | Heterosexual | 37.10 [35.20, 39.08] | 48.58 [44.43, 52.74] |
| | | LGB+ | 86.44 [72.94, 101.71] | 86.05 [71.79, 100.30] |
| | Bad health | Total | 67.19 [62.59, 72.05] | 93.64 [82.23, 105.04] |
| | | Heterosexual | 64.34 [59.73, 69.22] | 86.46 [74.39, 98.53] |
| | | LGB+ | 123.59 [96.34, 156.15] | 128.96 [99.08, 164.68] |
| | Very bad health | Total | 97.97 [87.20, 109.71] | 141.14 [112.80, 169.47] |
| | | Heterosexual | 92.48 [81.78, 104.19] | 132.28 [102.64, 161.91] |
| | | LGB+ | 175.38 [113.49, 258.89] | 189.48 [115.93, 288.74] |
| Disability | Day-to-day activities limited a lot | Total | 68.31 [64.72, 72.06] | 96.42 [88.75, 104.08] |
| | | Heterosexual | 64.23 [60.65, 67.96] | 88.53 [80.49, 96.56] |
| | | LGB+ | 144.99 [122.35, 170.61] | 145.04 [120.98, 169.10] |
| | | Gay or Lesbian | 149.44 [114.04, 192.36] | 155.44 [114.95, 204.52] |
| | | Bisexual | 145.11 [111.99, 184.95] | 139.84 [107.43, 178.83] |

| | | | | |
|---|---|---|---|---|
| | | Other | 132.80 [81.12, 205.10] | 125.42 [74.95, 196.03] |
| | Day-to-day activities limited a little | Total | 43.39 [41.15, 45.73] | 53.01 [49.49, 56.53] |
| | | Heterosexual | 41.63 [39.36, 44.01] | 51.35 [47.48, 55.22] |
| | | LGB+ | 69.23 [58.76, 81.02] | 71.92 [60.34, 83.50] |
| | | Gay or Lesbian | 88.65 [69.11, 112.01] | 93.00 [72.00, 118.11] |
| | | Bisexual | 56.50 [43.61, 72.02] | 64.28 [48.01, 83.86] |
| | | Other | 66.88 [40.85, 103.29] | 76.38 [43.92, 121.78] |
| | Day-to-day activities not limited | Total | 18.76 [18.21, 19.32] | 18.05 [17.41, 18.69] |
| | | Heterosexual | 18.28 [17.73, 18.84] | 17.51 [16.86, 18.17] |
| | | LGB+ | 33.27 [29.35, 37.57] | 32.80 [28.80, 36.81] |
| | | Gay or Lesbian | 36.99 [31.31, 43.41] | 33.68 [28.01, 39.35] |
| | | Bisexual | 28.55 [22.81, 35.31] | 31.67 [24.83, 39.70] |
| | | Other | 31.95 [20.67, 47.16] | 33.39 [21.54, 49.39] |
| Residence type | Living in a private household | Total | 24.88 [24.30, 25.46] | 24.35 [23.65, 25.06] |
| | | Heterosexual | 23.87 [23.30, 24.46] | 22.97 [22.26, 23.68] |
| | | LGB+ | 50.52 [46.35, 54.97] | 50.26 [46.02, 54.50] |
| | Living in a communal establishment | Total | 31.27 [25.44, 38.03] | 61.99 [44.73, 79.24] |
| | | Heterosexual | 30.00 [23.96, 37.09] | 61.45 [44.48, 81.40] |
| | | LGB+ | 41.16 [23.04, 67.88] | 43.92 [8.90, 95.55] |

Notes: Age-standardised rates are expressed per 100,000 people and standardised to the age distribution of the observed study population identifying with an LGB+ orientation in Census 2021. Sexual orientation groups Gay or Lesbian, Bisexual, and Other are not presented for categories where suppression was applied for the purposes of statistical disclosure control. All calculations are based on estimates rounded to the nearest multiple of five for disclosure control reasons. CI: confidence interval.

### Supplementary Table 8: Ratios of age-specific and age-standardised rates of self-harm and suicide per 100,000 people

| Characteristic | Category | Sexual orientation | Rate ratio denominator | Rate ratio for self-harm [95% CI] | Rate ratio for suicide [95% CI] |
|---|---|---|---|---|---|
| Total | Total | LGB+ | Heterosexual | 2.52 [2.48, 2.56] | 2.17 [1.98, 2.37] |
| | | Gay or Lesbian | Heterosexual | 2.43 [2.36, 2.49] | 2.19 [1.92, 2.47] |
| | | Bisexual | Heterosexual | 2.79 [2.73, 2.85] | 2.19 [1.87, 2.51] |
| | | Other | Heterosexual | 2.24 [2.13, 2.34] | 2.33 [1.75, 2.92] |
| Age on Census Day (10-year bands) | 16 to 24 years | LGB+ | Heterosexual | 2.78 [2.71, 2.85] | 1.91 [1.52, 2.33] |
| | | Gay or Lesbian | Heterosexual | 2.80 [2.68, 2.93] | c |
| | | Bisexual | Heterosexual | 2.82 [2.74, 2.91] | c |
| | | Other | Heterosexual | 2.48 [2.31, 2.65] | c |
| | 25 to 34 years | LGB+ | Heterosexual | 2.52 [2.44, 2.59] | 2.11 [1.76, 2.47] |
| | | Gay or Lesbian | Heterosexual | 2.35 [2.24, 2.46] | c |
| | | Bisexual | Heterosexual | 2.76 [2.64, 2.88] | c |
| | | Other | Heterosexual | 2.23 [2.04, 2.42] | c |
| | 35 to 44 years | LGB+ | Heterosexual | 2.23 [2.13, 2.33] | 2.21 [1.79, 2.64] |
| | | Gay or Lesbian | Heterosexual | 1.98 [1.85, 2.10] | c |
| | | Bisexual | Heterosexual | 2.78 [2.59, 2.97] | c |
| | | Other | Heterosexual | 1.82 [1.56, 2.08] | c |
| | 45 to 54 years | LGB+ | Heterosexual | 2.06 [1.94, 2.19] | 2.24 [1.78, 2.71] |
| | | Gay or Lesbian | Heterosexual | 1.71 [1.58, 1.85] | c |
| | | Bisexual | Heterosexual | 2.99 [2.70, 3.27] | c |
| | | Other | Heterosexual | 1.89 [1.51, 2.27] | c |
| | 55 to 64 years | LGB+ | Heterosexual | 1.92 [1.74, 2.10] | 2.52 [1.87, 3.18] |
| | | Gay or Lesbian | Heterosexual | 1.65 [1.45, 1.85] | c |
| | | Bisexual | Heterosexual | 2.65 [2.21, 3.09] | c |
| | | Other | Heterosexual | 2.04 [1.36, 2.73] | c |
| | 65 years and over | LGB+ | Heterosexual | 1.57 [1.32, 1.82] | 2.80 [1.70, 3.92] |
| | | Gay or Lesbian | Heterosexual | 1.49 [1.20, 1.78] | c |
| | | Bisexual | Heterosexual | 1.87 [1.32, 2.42] | c |
| | | Other | Heterosexual | 1.32 [0.50, 2.13] | c |
| Sex | Female | LGB+ | Heterosexual | 2.75 [2.70, 2.81] | 3.08 [2.64, 3.54] |
| | | Gay or Lesbian | Heterosexual | 3.04 [2.93, 3.15] | 2.94 [2.24, 3.66] |
| | | Bisexual | Heterosexual | 2.84 [2.76, 2.91] | 3.14 [2.47, 3.81] |
| | | Other | Heterosexual | 2.35 [2.22, 2.48] | 3.19 [1.99, 4.41] |
| | Male | LGB+ | Heterosexual | 1.91 [1.85, 1.97] | 1.77 [1.57, 1.98] |
| | | Gay or Lesbian | Heterosexual | 1.90 [1.82, 1.98] | 1.67 [1.42, 1.92] |
| | | Bisexual | Heterosexual | 2.09 [1.98, 2.20] | 1.99 [1.58, 2.41] |
| | | Other | Heterosexual | 1.71 [1.54, 1.88] | 2.15 [1.43, 2.87] |
| Region | England: North East | LGB+ | Heterosexual | 2.24 [2.08, 2.39] | 2.09 [1.36, 2.89] |
| | | Gay or Lesbian | Heterosexual | 2.33 [2.10, 2.57] | c |
| | | Bisexual | Heterosexual | 2.29 [2.04, 2.54] | c |
| | | Other | Heterosexual | 2.34 [1.84, 2.84] | c |

| | | | | | |
|---|---|---|---|---|---|
| | England: North West | LGB+ | Heterosexual | 2.75 [2.62, 2.88] | 2.66 [2.11, 3.23] |
| | | Gay or Lesbian | Heterosexual | 2.71 [2.53, 2.90] | c |
| | | Bisexual | Heterosexual | 3.07 [2.86, 3.27] | c |
| | | Other | Heterosexual | 2.15 [1.82, 2.47] | c |
| | England: Yorkshire and The Humber | LGB+ | Heterosexual | 2.17 [2.07, 2.28] | 2.24 [1.64, 2.88] |
| | | Gay or Lesbian | Heterosexual | 2.13 [1.97, 2.30] | c |
| | | Bisexual | Heterosexual | 2.35 [2.18, 2.52] | c |
| | | Other | Heterosexual | 1.91 [1.64, 2.19] | c |
| | England: East Midlands | LGB+ | Heterosexual | 2.63 [2.49, 2.78] | 2.59 [1.83, 3.40] |
| | | Gay or Lesbian | Heterosexual | 2.56 [2.33, 2.79] | c |
| | | Bisexual | Heterosexual | 2.92 [2.70, 3.14] | c |
| | | Other | Heterosexual | 2.17 [1.81, 2.53] | c |
| | England: West Midlands | LGB+ | Heterosexual | 2.87 [2.70, 3.05] | 1.65 [1.06, 2.26] |
| | | Gay or Lesbian | Heterosexual | 2.87 [2.59, 3.15] | c |
| | | Bisexual | Heterosexual | 3.16 [2.89, 3.42] | c |
| | | Other | Heterosexual | 2.19 [1.78, 2.60] | c |
| | England: East of England | LGB+ | Heterosexual | 2.69 [2.55, 2.83] | 2.34 [1.65, 3.08] |
| | | Gay or Lesbian | Heterosexual | 2.50 [2.28, 2.72] | c |
| | | Bisexual | Heterosexual | 3.01 [2.80, 3.22] | c |
| | | Other | Heterosexual | 2.45 [2.10, 2.81] | c |
| | England: London | LGB+ | Heterosexual | 2.30 [2.19, 2.42] | 2.31 [1.77, 2.89] |
| | | Gay or Lesbian | Heterosexual | 2.10 [1.91, 2.30] | c |
| | | Bisexual | Heterosexual | 2.57 [2.39, 2.74] | c |
| | | Other | Heterosexual | 2.37 [2.08, 2.66] | c |
| | England: South East | LGB+ | Heterosexual | 2.64 [2.53, 2.75] | 1.98 [1.53, 2.45] |
| | | Gay or Lesbian | Heterosexual | 2.40 [2.23, 2.57] | c |
| | | Bisexual | Heterosexual | 2.95 [2.79, 3.11] | c |
| | | Other | Heterosexual | 2.43 [2.15, 2.71] | c |
| | England: South West | LGB+ | Heterosexual | 2.71 [2.58, 2.85] | 1.79 [1.27, 2.35] |
| | | Gay or Lesbian | Heterosexual | 2.74 [2.52, 2.96] | c |
| | | Bisexual | Heterosexual | 2.88 [2.69, 3.08] | c |
| | | Other | Heterosexual | 2.56 [2.21, 2.91] | c |
| | Wales | LGB+ | Heterosexual | 2.25 [2.06, 2.45] | 2.68 [1.68, 3.81] |
| | | Gay or Lesbian | Heterosexual | 2.37 [2.05, 2.68] | c |
| | | Bisexual | Heterosexual | 2.50 [2.19, 2.81] | c |
| | | Other | Heterosexual | 1.38 [0.95, 1.82] | c |
| Ethnic group | Asian, Asian British or Asian Welsh | LGB+ | Heterosexual | 2.27 [2.04, 2.50] | 2.21 [0.95, 3.54] |
| | | Gay or Lesbian | Heterosexual | 2.63 [2.13, 3.13] | c |
| | | Bisexual | Heterosexual | 2.13 [1.84, 2.42] | c |
| | | Other | Heterosexual | 2.28 [1.72, 2.84] | c |
| | Black, Black British, Black Welsh, Caribbean or African | LGB+ | Heterosexual | 2.96 [2.61, 3.32] | 4.65 [2.19, 7.50] |
| | | Gay or Lesbian | Heterosexual | 3.12 [2.41, 3.85] | c |
| | | Bisexual | Heterosexual | 2.99 [2.51, 3.48] | c |
| | | Other | Heterosexual | 2.84 [2.11, 3.59] | c |
| | Mixed or Multiple ethnic groups | LGB+ | Heterosexual | 2.33 [2.15, 2.51] | 2.26 [1.27, 3.39] |
| | | Gay or Lesbian | Heterosexual | 2.17 [1.89, 2.44] | c |
| | | Bisexual | Heterosexual | 2.55 [2.28, 2.83] | c |
| | | Other | Heterosexual | 2.02 [1.61, 2.44] | c |
| | White | LGB+ | Heterosexual | 2.38 [2.34, 2.43] | 2.04 [1.85, 2.23] |
| | | Gay or Lesbian | Heterosexual | 2.25 [2.19, 2.32] | c |
| | | Bisexual | Heterosexual | 2.69 [2.62, 2.76] | c |
| | | Other | Heterosexual | 2.22 [2.11, 2.34] | c |
| | Other ethnic group | LGB+ | Heterosexual | 2.14 [1.80, 2.50] | 2.76 [0.24, 5.77] |
| | | Gay or Lesbian | Heterosexual | 2.63 [1.79, 3.48] | c |
| | | Bisexual | Heterosexual | 2.28 [1.76, 2.82] | c |
| | | Other | Heterosexual | 1.59 [1.08, 2.12] | c |
| Rural/urban classification | Urban | LGB+ | Heterosexual | 2.48 [2.43, 2.52] | 2.17 [1.97, 2.38] |
| | | Gay or Lesbian | Heterosexual | 2.39 [2.32, 2.46] | 2.14 [1.85, 2.44] |
| | | Bisexual | Heterosexual | 2.73 [2.66, 2.80] | 2.19 [1.85, 2.54] |
| | | Other | Heterosexual | 2.20 [2.08, 2.31] | 2.40 [1.77, 3.03] |
| | Rural | LGB+ | Heterosexual | 2.85 [2.71, 2.98] | 2.25 [1.71, 2.82] |
| | | Gay or Lesbian | Heterosexual | 2.68 [2.47, 2.89] | 2.62 [1.75, 3.53] |
| | | Bisexual | Heterosexual | 3.20 [2.99, 3.40] | 2.16 [1.31, 3.03] |
| | | Other | Heterosexual | 2.58 [2.20, 2.95] | 1.83 [0.24, 3.45] |
| Country of birth | Born in the UK | LGB+ | Heterosexual | 2.49 [2.45, 2.54] | 2.21 [2.00, 2.41] |
| | | Gay or Lesbian | Heterosexual | 2.34 [2.28, 2.41] | 2.18 [1.89, 2.47] |
| | | Bisexual | Heterosexual | 2.80 [2.73, 2.87] | 2.19 [1.84, 2.54] |
| | | Other | Heterosexual | 2.40 [2.28, 2.52] | 2.78 [2.03, 3.54] |
| | Born outside the UK | LGB+ | Heterosexual | 2.36 [2.23, 2.49] | 1.94 [1.44, 2.47] |
| | | Gay or Lesbian | Heterosexual | 2.25 [2.01, 2.50] | 2.05 [1.10, 3.04] |
| | | Bisexual | Heterosexual | 2.56 [2.37, 2.75] | 2.21 [1.37, 3.08] |

| | | | | | |
|---|---|---|---|---|---|
| | | Other | Heterosexual | 2.09 [1.81, 2.37] | 1.57 [0.47, 2.69] |
| English language proficiency | Main language is English | LGB+ | Heterosexual | 2.49 [2.45, 2.54] | 2.18 [1.99, 2.38] |
| | | Gay or Lesbian | Heterosexual | 2.36 [2.29, 2.42] | 2.16 [1.88, 2.44] |
| | | Bisexual | Heterosexual | 2.78 [2.72, 2.85] | 2.25 [1.91, 2.58] |
| | | Other | Heterosexual | 2.34 [2.23, 2.46] | 2.55 [1.88, 3.22] |
| | Main language is not English | LGB+ | Heterosexual | 2.04 [1.84, 2.24] | 1.44 [0.69, 2.23] |
| | | Gay or Lesbian | Heterosexual | 2.21 [1.75, 2.67] | 1.42 [0.15, 2.72] |
| | | Bisexual | Heterosexual | 2.08 [1.80, 2.35] | 1.00 [0.05, 1.97] |
| | | Other | Heterosexual | 1.92 [1.54, 2.30] | 2.11 [0.28, 4.01] |
| Religion | No religion | LGB+ | Heterosexual | 2.09 [2.05, 2.13] | 1.90 [1.70, 2.12] |
| | | Gay or Lesbian | Heterosexual | 1.98 [1.92, 2.05] | c |
| | | Bisexual | Heterosexual | 2.34 [2.27, 2.41] | c |
| | | Other | Heterosexual | 1.98 [1.86, 2.10] | c |
| | Christian | LGB+ | Heterosexual | 2.95 [2.85, 3.06] | 2.23 [1.80, 2.67] |
| | | Gay or Lesbian | Heterosexual | 2.85 [2.67, 3.02] | c |
| | | Bisexual | Heterosexual | 3.46 [3.28, 3.63] | c |
| | | Other | Heterosexual | 2.19 [1.93, 2.44] | c |
| | Buddhist | LGB+ | Heterosexual | 4.05 [3.34, 4.86] | 2.65 [0.83, 5.35] |
| | | Gay or Lesbian | Heterosexual | 3.20 [2.18, 4.30] | c |
| | | Bisexual | Heterosexual | 4.26 [3.34, 5.28] | c |
| | | Other | Heterosexual | 4.52 [2.93, 6.20] | c |
| | Hindu | LGB+ | Heterosexual | 2.40 [1.75, 3.06] | 2.48 [0.00, 5.84] |
| | | Gay or Lesbian | Heterosexual | 3.17 [1.23, 5.13] | c |
| | | Bisexual | Heterosexual | 2.42 [1.65, 3.23] | c |
| | | Other | Heterosexual | 1.82 [0.35, 3.31] | c |
| | Jewish | LGB+ | Heterosexual | 3.06 [2.34, 3.89] | 2.57 [0.00, 6.80] |
| | | Gay or Lesbian | Heterosexual | 2.54 [1.53, 3.65] | c |
| | | Bisexual | Heterosexual | 3.66 [2.56, 4.89] | c |
| | | Other | Heterosexual | 2.53 [0.88, 4.27] | c |
| | Muslim | LGB+ | Heterosexual | 2.28 [1.93, 2.63] | 2.79 [0.00, 6.22] |
| | | Gay or Lesbian | Heterosexual | 3.45 [2.34, 4.57] | c |
| | | Bisexual | Heterosexual | 2.38 [1.90, 2.87] | c |
| | | Other | Heterosexual | 1.63 [1.12, 2.15] | c |
| | Sikh | LGB+ | Heterosexual | 2.20 [1.40, 3.03] | 3.66 [0.00, 8.36] |
| | | Gay or Lesbian | Heterosexual | 4.18 [1.57, 6.86] | c |
| | | Bisexual | Heterosexual | 1.35 [0.55, 2.17] | c |
| | | Other | Heterosexual | 2.66 [0.78, 4.59] | c |
| | Other religion | LGB+ | Heterosexual | 2.80 [2.52, 3.13] | 1.13 [0.59, 1.90] |
| | | Gay or Lesbian | Heterosexual | 2.97 [2.47, 3.51] | c |
| | | Bisexual | Heterosexual | 2.82 [2.47, 3.20] | c |
| | | Other | Heterosexual | 2.64 [2.22, 3.10] | c |
| | Not answered | LGB+ | Heterosexual | 2.76 [2.49, 3.04] | 1.60 [0.78, 2.50] |
| | | Gay or Lesbian | Heterosexual | 2.67 [2.23, 3.11] | c |
| | | Bisexual | Heterosexual | 3.14 [2.71, 3.58] | c |
| | | Other | Heterosexual | 2.25 [1.66, 2.85] | c |
| IMD/WIMD decile group | 1 (most deprived) | LGB+ | Heterosexual | 2.39 [2.29, 2.50] | 2.16 [1.63, 2.72] |
| | | Gay or Lesbian | Heterosexual | 2.49 [2.32, 2.66] | 2.12 [1.38, 2.90] |
| | | Bisexual | Heterosexual | 2.63 [2.47, 2.79] | 2.60 [1.64, 3.58] |
| | | Other | Heterosexual | 1.54 [1.32, 1.77] | 1.04 [0.08, 2.02] |
| | 2 | LGB+ | Heterosexual | 2.30 [2.20, 2.41] | 1.91 [1.40, 2.46] |
| | | Gay or Lesbian | Heterosexual | 2.33 [2.16, 2.50] | 1.81 [1.05, 2.61] |
| | | Bisexual | Heterosexual | 2.41 [2.26, 2.57] | 1.91 [1.11, 2.75] |
| | | Other | Heterosexual | 1.96 [1.71, 2.22] | 2.19 [0.69, 3.73] |
| | 3 | LGB+ | Heterosexual | 2.36 [2.25, 2.47] | 2.50 [1.92, 3.12] |
| | | Gay or Lesbian | Heterosexual | 2.44 [2.26, 2.62] | 2.75 [1.85, 3.71] |
| | | Bisexual | Heterosexual | 2.52 [2.36, 2.69] | 2.06 [1.25, 2.91] |
| | | Other | Heterosexual | 1.88 [1.63, 2.14] | 2.92 [1.19, 4.69] |
| | 4 | LGB+ | Heterosexual | 2.42 [2.31, 2.54] | 1.92 [1.44, 2.44] |
| | | Gay or Lesbian | Heterosexual | 2.44 [2.24, 2.63] | 2.15 [1.41, 2.93] |
| | | Bisexual | Heterosexual | 2.63 [2.46, 2.81] | 1.60 [0.84, 2.37] |
| | | Other | Heterosexual | 2.29 [1.98, 2.60] | 2.04 [0.56, 3.55] |
| | 5 | LGB+ | Heterosexual | 2.60 [2.47, 2.73] | 1.82 [1.32, 2.34] |
| | | Gay or Lesbian | Heterosexual | 2.47 [2.26, 2.68] | 1.83 [1.17, 2.52] |
| | | Bisexual | Heterosexual | 2.78 [2.59, 2.98] | 1.52 [0.77, 2.27] |
| | | Other | Heterosexual | 2.48 [2.13, 2.83] | 1.41 [0.21, 2.64] |
| | 6 | LGB+ | Heterosexual | 2.49 [2.35, 2.62] | 2.39 [1.77, 3.05] |
| | | Gay or Lesbian | Heterosexual | 2.19 [1.99, 2.39] | 2.29 [1.46, 3.15] |
| | | Bisexual | Heterosexual | 2.96 [2.74, 3.17] | 2.53 [1.44, 3.66] |
| | | Other | Heterosexual | 2.14 [1.79, 2.49] | 2.30 [0.50, 4.15] |
| | 7 | LGB+ | Heterosexual | 2.64 [2.49, 2.80] | 2.09 [1.48, 2.75] |
| | | Gay or Lesbian | Heterosexual | 2.23 [2.00, 2.46] | 1.83 [1.03, 2.67] |

| | | | | | |
|---|---|---|---|---|---|
| | | Bisexual | Heterosexual | 3.09 [2.85, 3.33] | 2.20 [1.09, 3.36] |
| | | Other | Heterosexual | 2.70 [2.27, 3.14] | 3.98 [1.19, 6.86] |
| | 8 | LGB+ | Heterosexual | 2.72 [2.55, 2.88] | 2.36 [1.67, 3.12] |
| | | Gay or Lesbian | Heterosexual | 2.46 [2.21, 2.70] | 2.16 [1.23, 3.13] |
| | | Bisexual | Heterosexual | 3.09 [2.83, 3.35] | 2.56 [1.37, 3.80] |
| | | Other | Heterosexual | 2.61 [2.14, 3.07] | 2.54 [0.02, 5.12] |
| | 9 | LGB+ | Heterosexual | 2.75 [2.57, 2.93] | 2.09 [1.40, 2.85] |
| | | Gay or Lesbian | Heterosexual | 2.59 [2.31, 2.87] | 2.32 [1.19, 3.52] |
| | | Bisexual | Heterosexual | 3.05 [2.77, 3.33] | 2.43 [1.12, 3.80] |
| | | Other | Heterosexual | 3.04 [2.51, 3.59] | 1.90 [0.00, 3.98] |
| | 10 (least deprived) | LGB+ | Heterosexual | 2.89 [2.68, 3.11] | 2.50 [1.63, 3.45] |
| | | Gay or Lesbian | Heterosexual | 2.41 [2.11, 2.72] | 2.86 [1.41, 4.39] |
| | | Bisexual | Heterosexual | 3.35 [3.02, 3.70] | 2.47 [0.94, 4.05] |
| | | Other | Heterosexual | 3.10 [2.50, 3.72] | 4.32 [0.53, 8.24] |
| Highest level of qualification (≥25 years) | No qualifications | LGB+ | Heterosexual | 2.01 [1.88, 2.14] | 2.01 [1.43, 2.62] |
| | | Gay or Lesbian | Heterosexual | 2.85 [2.58, 3.12] | c |
| | | Bisexual | Heterosexual | 2.25 [2.03, 2.47] | c |
| | | Other | Heterosexual | 0.69 [0.55, 0.83] | c |
| | Level 1 (one to four GCSE passes (grade A* to C or grade 4 and above) and any other GCSEs at other grades, or equivalent qualifications) | LGB+ | Heterosexual | 2.55 [2.37, 2.73] | 1.72 [1.04, 2.44] |
| | | Gay or Lesbian | Heterosexual | 2.36 [2.11, 2.61] | c |
| | | Bisexual | Heterosexual | 3.04 [2.74, 3.34] | c |
| | | Other | Heterosexual | 1.77 [1.32, 2.23] | c |
| | Level 2 (five or more GCSE passes (grade A* to C or grade 4 and above) or equivalent qualifications) | LGB+ | Heterosexual | 2.59 [2.45, 2.74] | 1.72 [1.21, 2.25] |
| | | Gay or Lesbian | Heterosexual | 2.16 [1.98, 2.35] | c |
| | | Bisexual | Heterosexual | 3.24 [2.99, 3.50] | c |
| | | Other | Heterosexual | 2.29 [1.85, 2.72] | c |
| | Apprenticeship | LGB+ | Heterosexual | 2.23 [1.95, 2.52] | 0.93 [0.23, 1.67] |
| | | Gay or Lesbian | Heterosexual | 2.18 [1.76, 2.60] | c |
| | | Bisexual | Heterosexual | 2.74 [2.26, 3.23] | c |
| | | Other | Heterosexual | 1.18 [0.67, 1.70] | c |
| | Level 3 (two or more A Levels or equivalent qualifications) | LGB+ | Heterosexual | 3.00 [2.86, 3.15] | 2.91 [2.30, 3.55] |
| | | Gay or Lesbian | Heterosexual | 2.66 [2.48, 2.84] | c |
| | | Bisexual | Heterosexual | 3.53 [3.27, 3.79] | c |
| | | Other | Heterosexual | 3.37 [2.88, 3.85] | c |
| | Level 4 and above (Higher National Certificate, Higher National Diploma, Bachelor's degree, or post-graduate qualifications) | LGB+ | Heterosexual | 2.46 [2.36, 2.55] | 2.57 [2.20, 2.95] |
| | | Gay or Lesbian | Heterosexual | 2.16 [2.04, 2.28] | c |
| | | Bisexual | Heterosexual | 2.87 [2.69, 3.04] | c |
| | | Other | Heterosexual | 2.86 [2.55, 3.18] | c |
| | Other qualifications | LGB+ | Heterosexual | 2.10 [1.72, 2.48] | 2.33 [0.65, 4.24] |
| | | Gay or Lesbian | Heterosexual | 2.49 [1.85, 3.14] | c |
| | | Bisexual | Heterosexual | 2.19 [1.59, 2.81] | c |
| | | Other | Heterosexual | 1.15 [0.53, 1.76] | c |
| NS-SEC | Class 1 (Higher managerial, administrative and professional occupations) | LGB+ | Heterosexual | 3.58 [2.74, 4.48] | 2.24 [1.62, 2.90] |
| | | Gay or Lesbian | Heterosexual | 2.98 [1.70, 4.30] | c |
| | | Bisexual | Heterosexual | 4.17 [2.97, 5.42] | c |
| | | Other | Heterosexual | 3.51 [1.44, 5.64] | c |
| | Class 2 (Lower managerial, administrative and professional occupations) | LGB+ | Heterosexual | 2.96 [2.67, 3.25] | 2.67 [1.66, 3.76] |
| | | Gay or Lesbian | Heterosexual | 2.62 [2.17, 3.08] | c |
| | | Bisexual | Heterosexual | 3.36 [2.95, 3.77] | c |
| | | Other | Heterosexual | 2.86 [2.14, 3.60] | c |
| | Class 3 (Intermediate occupations) | LGB+ | Heterosexual | 2.85 [2.66, 3.05] | 2.34 [1.67, 3.07] |
| | | Gay or Lesbian | Heterosexual | 2.73 [2.42, 3.04] | c |
| | | Bisexual | Heterosexual | 3.20 [2.92, 3.48] | c |
| | | Other | Heterosexual | 2.16 [1.70, 2.64] | c |
| | | LGB+ | Heterosexual | 2.10 [1.79, 2.41] | 1.23 [0.77, 1.72] |

| | | | | | |
|---|---|---|---|---|---|
| | Class 4 (Small employers and own account workers) | Gay or Lesbian | Heterosexual | 2.04 [1.42, 2.66] | c |
| | | Bisexual | Heterosexual | 2.46 [1.99, 2.93] | c |
| | | Other | Heterosexual | 1.31 [0.88, 1.74] | c |
| | Class 5 (Lower supervisory and technical occupations) | LGB+ | Heterosexual | 2.88 [2.55, 3.22] | 1.86 [1.12, 2.66] |
| | | Gay or Lesbian | Heterosexual | 2.75 [2.27, 3.24] | c |
| | | Bisexual | Heterosexual | 3.13 [2.65, 3.62] | c |
| | | Other | Heterosexual | 2.70 [1.44, 3.95] | c |
| | Class 6 (Semi-routine occupations) | LGB+ | Heterosexual | 2.60 [2.48, 2.72] | 2.70 [2.05, 3.40] |
| | | Gay or Lesbian | Heterosexual | 2.54 [2.36, 2.72] | c |
| | | Bisexual | Heterosexual | 2.79 [2.62, 2.96] | c |
| | | Other | Heterosexual | 2.25 [1.93, 2.57] | c |
| | Class 7 (Routine occupations) | LGB+ | Heterosexual | 2.43 [2.31, 2.55] | 2.19 [1.66, 2.74] |
| | | Gay or Lesbian | Heterosexual | 2.48 [2.28, 2.68] | c |
| | | Bisexual | Heterosexual | 2.60 [2.43, 2.78] | c |
| | | Other | Heterosexual | 1.77 [1.50, 2.05] | c |
| | Class 8 (Never worked and long-term unemployed) | LGB+ | Heterosexual | 2.32 [2.21, 2.42] | 2.18 [1.61, 2.78] |
| | | Gay or Lesbian | Heterosexual | 2.82 [2.62, 3.01] | c |
| | | Bisexual | Heterosexual | 2.45 [2.30, 2.60] | c |
| | | Other | Heterosexual | 1.26 [1.09, 1.42] | c |
| | Full-time students | LGB+ | Heterosexual | 2.78 [2.49, 3.07] | 3.74 [0.10, 7.85] |
| | | Gay or Lesbian | Heterosexual | 2.95 [2.45, 3.44] | c |
| | | Bisexual | Heterosexual | 2.51 [2.24, 2.79] | c |
| | | Other | Heterosexual | 2.80 [2.15, 3.45] | c |
| Household NS-SEC | Class 1 (Higher managerial, administrative and professional occupations) | LGB+ | Heterosexual | 2.79 [2.66, 2.93] | 2.28 [1.82, 2.77] |
| | | Gay or Lesbian | Heterosexual | 2.55 [2.34, 2.77] | c |
| | | Bisexual | Heterosexual | 3.09 [2.89, 3.29] | c |
| | | Other | Heterosexual | 3.04 [2.66, 3.42] | c |
| | Class 2 (Lower managerial, administrative and professional occupations) | LGB+ | Heterosexual | 2.64 [2.54, 2.74] | 2.24 [1.84, 2.65] |
| | | Gay or Lesbian | Heterosexual | 2.47 [2.32, 2.62] | c |
| | | Bisexual | Heterosexual | 2.89 [2.74, 3.04] | c |
| | | Other | Heterosexual | 2.72 [2.44, 3.00] | c |
| | Class 3 (Intermediate occupations) | LGB+ | Heterosexual | 2.77 [2.63, 2.91] | 2.23 [1.62, 2.88] |
| | | Gay or Lesbian | Heterosexual | 2.80 [2.59, 3.01] | c |
| | | Bisexual | Heterosexual | 2.87 [2.67, 3.07] | c |
| | | Other | Heterosexual | 2.37 [2.02, 2.72] | c |
| | Class 4 (Small employers and own account workers) | LGB+ | Heterosexual | 2.24 [2.08, 2.41] | 1.58 [1.02, 2.15] |
| | | Gay or Lesbian | Heterosexual | 2.42 [2.14, 2.71] | c |
| | | Bisexual | Heterosexual | 2.40 [2.16, 2.64] | c |
| | | Other | Heterosexual | 1.64 [1.32, 1.96] | c |
| | Class 5 (Lower supervisory and technical occupations) | LGB+ | Heterosexual | 2.59 [2.39, 2.79] | 2.12 [1.29, 3.03] |
| | | Gay or Lesbian | Heterosexual | 2.69 [2.37, 3.02] | c |
| | | Bisexual | Heterosexual | 2.86 [2.57, 3.16] | c |
| | | Other | Heterosexual | 1.43 [1.03, 1.82] | c |
| | Class 6 (Semi-routine occupations) | LGB+ | Heterosexual | 2.55 [2.44, 2.67] | 2.53 [1.87, 3.23] |
| | | Gay or Lesbian | Heterosexual | 2.71 [2.52, 2.90] | c |
| | | Bisexual | Heterosexual | 2.62 [2.46, 2.79] | c |
| | | Other | Heterosexual | 2.06 [1.78, 2.33] | c |
| | Class 7 (Routine occupations) | LGB+ | Heterosexual | 2.44 [2.31, 2.58] | 2.37 [1.72, 3.07] |
| | | Gay or Lesbian | Heterosexual | 2.58 [2.34, 2.81] | c |
| | | Bisexual | Heterosexual | 2.63 [2.44, 2.82] | c |
| | | Other | Heterosexual | 1.58 [1.33, 1.84] | c |
| | Class 8 (Never worked and long-term unemployed) | LGB+ | Heterosexual | 2.20 [2.09, 2.32] | 2.14 [1.44, 2.91] |
| | | Gay or Lesbian | Heterosexual | 2.75 [2.51, 2.99] | c |
| | | Bisexual | Heterosexual | 2.30 [2.14, 2.47] | c |
| | | Other | Heterosexual | 1.12 [0.93, 1.31] | c |
| | Full-time students | LGB+ | Heterosexual | 2.66 [2.24, 3.10] | 3.85 [0.00, 10.32] |
| | | Gay or Lesbian | Heterosexual | 2.91 [2.18, 3.66] | c |
| | | Bisexual | Heterosexual | 2.36 [1.97, 2.77] | c |
| | | Other | Heterosexual | 2.68 [1.53, 3.86] | c |
| | Living in a communal establishment | LGB+ | Heterosexual | 2.28 [1.92, 2.65] | 0.71 [0.06, 1.50] |
| | | Gay or Lesbian | Heterosexual | 1.80 [1.39, 2.22] | c |
| | | Bisexual | Heterosexual | 3.44 [2.63, 4.27] | c |
| | | Other | Heterosexual | 0.99 [0.63, 1.36] | c |
| Lone person household indicator | Lives alone | LGB+ | Heterosexual | 2.06 [1.97, 2.16] | 1.75 [1.42, 2.12] |
| | | Gay or Lesbian | Heterosexual | 2.11 [1.94, 2.28] | c |
| | | Bisexual | Heterosexual | 2.23 [2.10, 2.35] | c |
| | | Other | Heterosexual | 1.73 [1.51, 1.95] | c |
| | Does not live alone | LGB+ | Heterosexual | 2.41 [2.36, 2.46] | 2.00 [1.76, 2.24] |
| | | Gay or Lesbian | Heterosexual | 2.31 [2.23, 2.38] | c |

| | | | | | |
|---|---|---|---|---|---|
| | | Bisexual | Heterosexual | 2.67 [2.59, 2.74] | c |
| | | Other | Heterosexual | 2.11 [1.99, 2.23] | c |
| | Living in a communal establishment | LGB+ | Heterosexual | 2.28 [1.92, 2.66] | 0.71 [0.06, 1.51] |
| | | Gay or Lesbian | Heterosexual | 1.80 [1.39, 2.22] | c |
| | | Bisexual | Heterosexual | 3.44 [2.63, 4.28] | c |
| | | Other | Heterosexual | 0.99 [0.63, 1.36] | c |
| Household size | One person in household | LGB+ | Heterosexual | 2.06 [1.97, 2.16] | 1.75 [1.41, 2.12] |
| | | Gay or Lesbian | Heterosexual | 2.11 [1.94, 2.28] | c |
| | | Bisexual | Heterosexual | 2.23 [2.10, 2.35] | c |
| | | Other | Heterosexual | 1.73 [1.51, 1.95] | c |
| | Two people in household | LGB+ | Heterosexual | 2.08 [2.00, 2.17] | 1.69 [1.40, 1.99] |
| | | Gay or Lesbian | Heterosexual | 1.99 [1.85, 2.12] | c |
| | | Bisexual | Heterosexual | 2.36 [2.24, 2.48] | c |
| | | Other | Heterosexual | 2.09 [1.87, 2.32] | c |
| | Three to five people in household | LGB+ | Heterosexual | 2.50 [2.43, 2.58] | 2.25 [1.77, 2.74] |
| | | Gay or Lesbian | Heterosexual | 2.45 [2.33, 2.57] | c |
| | | Bisexual | Heterosexual | 2.71 [2.60, 2.82] | c |
| | | Other | Heterosexual | 2.00 [1.84, 2.16] | c |
| | Six or more people in household | LGB+ | Heterosexual | 2.51 [2.28, 2.74] | 1.36 [0.15, 2.63] |
| | | Gay or Lesbian | Heterosexual | 2.70 [2.25, 3.16] | c |
| | | Bisexual | Heterosexual | 2.74 [2.41, 3.06] | c |
| | | Other | Heterosexual | 1.79 [1.37, 2.22] | c |
| | Living in a communal establishment | LGB+ | Heterosexual | 2.28 [1.92, 2.65] | 0.71 [0.05, 1.50] |
| | | Gay or Lesbian | Heterosexual | 1.80 [1.39, 2.23] | c |
| | | Bisexual | Heterosexual | 3.44 [2.63, 4.27] | c |
| | | Other | Heterosexual | 0.99 [0.63, 1.36] | c |
| Household tenure | Owned or shared ownership | LGB+ | Heterosexual | 2.50 [2.41, 2.58] | 1.99 [1.68, 2.30] |
| | | Gay or Lesbian | Heterosexual | 2.34 [2.21, 2.47] | c |
| | | Bisexual | Heterosexual | 2.71 [2.58, 2.83] | c |
| | | Other | Heterosexual | 2.67 [2.42, 2.91] | c |
| | Social rented | LGB+ | Heterosexual | 2.62 [2.55, 2.70] | 2.31 [1.90, 2.75] |
| | | Gay or Lesbian | Heterosexual | 2.68 [2.55, 2.80] | c |
| | | Bisexual | Heterosexual | 2.81 [2.70, 2.92] | c |
| | | Other | Heterosexual | 1.92 [1.74, 2.10] | c |
| | Private rented or living rent free | LGB+ | Heterosexual | 2.35 [2.28, 2.42] | 2.21 [1.87, 2.56] |
| | | Gay or Lesbian | Heterosexual | 2.41 [2.29, 2.53] | c |
| | | Bisexual | Heterosexual | 2.45 [2.35, 2.55] | c |
| | | Other | Heterosexual | 1.93 [1.77, 2.09] | c |
| | Living in a communal establishment | LGB+ | Heterosexual | 2.28 [1.92, 2.65] | 0.71 [0.06, 1.50] |
| | | Gay or Lesbian | Heterosexual | 1.80 [1.40, 2.23] | c |
| | | Bisexual | Heterosexual | 3.44 [2.63, 4.27] | c |
| | | Other | Heterosexual | 0.99 [0.63, 1.36] | c |
| Economic activity | Economically active | LGB+ | Heterosexual | 2.43 [2.37, 2.49] | 2.16 [1.90, 2.42] |
| | | Gay or Lesbian | Heterosexual | 2.32 [2.22, 2.41] | 2.19 [1.81, 2.58] |
| | | Bisexual | Heterosexual | 2.60 [2.51, 2.69] | 2.24 [1.80, 2.69] |
| | | Other | Heterosexual | 2.28 [2.12, 2.45] | 2.57 [1.71, 3.43] |
| | Economically inactive | LGB+ | Heterosexual | 2.51 [2.45, 2.58] | 2.12 [1.80, 2.46] |
| | | Gay or Lesbian | Heterosexual | 2.85 [2.73, 2.97] | 2.78 [2.15, 3.43] |
| | | Bisexual | Heterosexual | 2.63 [2.54, 2.72] | 1.87 [1.42, 2.34] |
| | | Other | Heterosexual | 1.75 [1.63, 1.88] | 1.43 [0.80, 2.07] |
| General health (detailed) | Very good health | LGB+ | Heterosexual | 2.11 [2.03, 2.19] | 2.06 [1.69, 2.45] |
| | | Gay or Lesbian | Heterosexual | 2.22 [2.09, 2.34] | c |
| | | Bisexual | Heterosexual | 2.23 [2.10, 2.35] | c |
| | | Other | Heterosexual | 1.57 [1.36, 1.78] | c |
| | Good health | LGB+ | Heterosexual | 2.00 [1.94, 2.06] | 1.77 [1.51, 2.05] |
| | | Gay or Lesbian | Heterosexual | 2.09 [1.99, 2.18] | c |
| | | Bisexual | Heterosexual | 2.08 [2.00, 2.16] | c |
| | | Other | Heterosexual | 1.55 [1.42, 1.68] | c |
| | Fair health | LGB+ | Heterosexual | 1.77 [1.71, 1.83] | 1.77 [1.45, 2.12] |
| | | Gay or Lesbian | Heterosexual | 1.99 [1.89, 2.10] | c |
| | | Bisexual | Heterosexual | 1.80 [1.72, 1.87] | c |
| | | Other | Heterosexual | 1.37 [1.25, 1.49] | c |
| | Bad health | LGB+ | Heterosexual | 1.58 [1.50, 1.66] | 1.49 [1.09, 1.95] |
| | | Gay or Lesbian | Heterosexual | 1.73 [1.58, 1.88] | c |
| | | Bisexual | Heterosexual | 1.61 [1.50, 1.72] | c |
| | | Other | Heterosexual | 1.31 [1.16, 1.47] | c |
| | Very bad health | LGB+ | Heterosexual | 1.75 [1.57, 1.93] | 1.43 [0.79, 2.22] |
| | | Gay or Lesbian | Heterosexual | 1.95 [1.62, 2.28] | c |
| | | Bisexual | Heterosexual | 1.83 [1.59, 2.07] | c |
| | | Other | Heterosexual | 1.35 [1.06, 1.64] | c |

| | | | | | |
|---|---|---|---|---|---|
| General health (aggregated) | Good health | LGB+ | Heterosexual | 2.23 [2.18, 2.28] | 1.99 [1.76, 2.22] |
| | | Gay or Lesbian | Heterosexual | 2.26 [2.18, 2.34] | 2.25 [1.90, 2.60] |
| | | Bisexual | Heterosexual | 2.38 [2.31, 2.46] | 1.86 [1.46, 2.26] |
| | | Other | Heterosexual | 1.75 [1.63, 1.88] | 1.79 [1.10, 2.50] |
| | Not good health | LGB+ | Heterosexual | 1.77 [1.72, 1.82] | 1.71 [1.47, 1.97] |
| | | Gay or Lesbian | Heterosexual | 1.97 [1.88, 2.05] | 1.63 [1.25, 2.03] |
| | | Bisexual | Heterosexual | 1.80 [1.74, 1.86] | 1.68 [1.35, 2.03] |
| | | Other | Heterosexual | 1.44 [1.35, 1.54] | 1.88 [1.27, 2.52] |
| Disability | Day-to-day activities limited a lot | LGB+ | Heterosexual | 1.72 [1.66, 1.77] | 1.64 [1.34, 1.96] |
| | | Gay or Lesbian | Heterosexual | 1.85 [1.75, 1.95] | 1.76 [1.25, 2.29] |
| | | Bisexual | Heterosexual | 1.77 [1.69, 1.84] | 1.58 [1.18, 2.01] |
| | | Other | Heterosexual | 1.40 [1.29, 1.51] | 1.42 [0.77, 2.09] |
| | Day-to-day activities limited a little | LGB+ | Heterosexual | 1.41 [1.37, 1.45] | 1.40 [1.16, 1.66] |
| | | Gay or Lesbian | Heterosexual | 1.63 [1.56, 1.71] | 1.81 [1.37, 2.28] |
| | | Bisexual | Heterosexual | 1.43 [1.38, 1.49] | 1.25 [0.91, 1.61] |
| | | Other | Heterosexual | 1.04 [0.95, 1.12] | 1.49 [0.78, 2.22] |
| | Day-to-day activities not limited | LGB+ | Heterosexual | 1.92 [1.87, 1.97] | 1.87 [1.64, 2.12] |
| | | Gay or Lesbian | Heterosexual | 2.01 [1.93, 2.10] | 1.92 [1.59, 2.26] |
| | | Bisexual | Heterosexual | 1.99 [1.91, 2.07] | 1.81 [1.39, 2.22] |
| | | Other | Heterosexual | 1.42 [1.29, 1.55] | 1.91 [1.15, 2.67] |
| Residence type | Living in a private household | LGB+ | Heterosexual | 2.53 [2.49, 2.58] | 2.19 [1.99, 2.39] |
| | | Gay or Lesbian | Heterosexual | 2.45 [2.39, 2.52] | c |
| | | Bisexual | Heterosexual | 2.79 [2.72, 2.85] | c |
| | | Other | Heterosexual | 2.24 [2.13, 2.35] | c |
| | Living in a communal establishment | LGB+ | Heterosexual | 2.28 [1.92, 2.65] | 0.71 [0.06, 1.52] |
| | | Gay or Lesbian | Heterosexual | 1.80 [1.39, 2.23] | c |
| | | Bisexual | Heterosexual | 3.44 [2.63, 4.27] | c |
| | | Other | Heterosexual | 0.99 [0.63, 1.36] | c |

Notes: Rate ratios were calculated using age-specific rates for the age on Census Day variable, and age-standardised rates for all other variables. Cells with the value 'c' have been suppressed for the purposes of statistical disclosure control. All calculations are based on estimates rounded to the nearest multiple of five for disclosure control reasons. CI: confidence interval.

### Supplementary Table 9: Monthly counts of hospital admissions and emergency attendances for intentional self-harm

| **Month** | **Count of hospital admissions and emergency attendances for intentional self-harm** |
|---|---|
| April 2021 | 17,920 |
| May 2021 | 21,955 |
| June 2021 | 21,305 |
| July 2021 | 23,430 |
| August 2021 | 22,640 |
| September 2021 | 22,725 |
| October 2021 | 20,975 |
| November 2021 | 20,825 |
| December 2021 | 21,980 |
| January 2022 | 20,765 |
| February 2022 | 19,005 |
| March 2022 | 21,360 |
| April 2022 | 21,760 |
| May 2022 | 18,570 |
| June 2022 | 13,390 |
| July 2022 | 17,925 |
| August 2022 | 19,755 |
| September 2022 | 21,115 |
| October 2022 | 20,885 |
| November 2022 | 19,150 |
| December 2022 | 19,700 |
| January 2023 | 19,200 |
| February 2023 | 16,475 |
| March 2023 | 16,480 |
| April 2023 | 17,025 |
| May 2023 | 21,180 |
| June 2023 | 21,305 |
| July 2023 | 23,645 |
| August 2023 | 22,735 |
| September 2023 | 21,005 |
| October 2023 | 18,730 |
| November 2023 | 19,480 |

| | |
|---|---|
| December 2023 | 19,025 |

### Supplementary Table 10: Monthly counts of deaths by suicide

| Month | Count of deaths by suicide |
|---|---|
| April 2021 | 495 |
| May 2021 | 500 |
| June 2021 | 560 |
| July 2021 | 510 |
| August 2021 | 470 |
| September 2021 | 485 |
| October 2021 | 480 |
| November 2021 | 505 |
| December 2021 | 450 |
| January 2022 | 500 |
| February 2022 | 460 |
| March 2022 | 540 |
| April 2022 | 475 |
| May 2022 | 525 |
| June 2022 | 485 |
| July 2022 | 530 |
| August 2022 | 495 |
| September 2022 | 460 |
| October 2022 | 485 |
| November 2022 | 435 |
| December 2022 | 475 |
| January 2023 | 455 |
| February 2023 | 425 |
| March 2023 | 470 |
| April 2023 | 490 |
| May 2023 | 500 |
| June 2023 | 460 |
| July 2023 | 470 |
| August 2023 | 430 |
| September 2023 | 440 |
| October 2023 | 430 |
| November 2023 | 420 |
| December 2023 | 370 |

**Supplementary Table 11: Sensitivity analysis including proxy responses to the sexual orientation question in Census 2021, crude and age-standardised rates of self-harm per 100,000 people**

| Characteristic | Category | Sexual orientation | Count (%), Individuals | Count (%), Individuals with at least one hospital admission or emergency attendance for intentional self-harm | Crude rate of self-harm [95% CI] | Age-standardised rate of self-harm [95% CI] |
|---|---|---|---|---|---|---|
| Total | Total | Total | 41,198,325 (100.0) | 216,180 (100.0) | 524.73 [522.52, 526.95] | 735.62 [732.10, 739.15] |
| | | Heterosexual | 39,817,525 (96.6) | 192,500 (89.0) | 483.46 [481.30, 485.62] | 669.73 [666.27, 673.18] |
| | | LGB+ | 1,380,800 (3.4) | 23,680 (11.0) | 1,714.95 [1,693.17, 1,736.93] | 1,715.10 [1,693.26, 1,736.95] |
| | | Gay or Lesbian | 674,555 (1.6) | 9,220 (4.3) | 1,366.83 [1,339.07, 1,395.02] | 1,658.91 [1,623.69, 1,694.12] |
| | | Bisexual | 559,255 (1.4) | 12,165 (5.6) | 2,175.22 [2,136.73, 2,214.22] | 1,891.43 [1,856.24, 1,926.61] |
| | | Other | 146,990 (0.4) | 2,295 (1.1) | 1,561.33 [1,498.10, 1,626.54] | 1,480.31 [1,419.30, 1,541.31] |
| Age on Census Day (10-year bands) | 16 to 24 years | Total | 5,105,765 (100.0) | 68,060 (100.0) | 1,333.00 [1,322.99, 1,343.02] | - |
| | | Heterosexual | 4,719,460 (92.4) | 55,505 (81.6) | 1,176.09 [1,166.30, 1,185.87] | - |
| | | LGB+ | 386,305 (7.6) | 12,550 (18.4) | 3,248.73 [3,191.89, 3,305.57] | - |
| | | Gay or Lesbian | 118,690 (2.3) | 3,885 (5.7) | 3,273.23 [3,170.30, 3,376.16] | - |
| | | Bisexual | 222,655 (4.4) | 7,405 (10.9) | 3,325.77 [3,250.02, 3,401.52] | - |
| | | Other | 44,960 (0.9) | 1,260 (1.9) | 2,802.49 [2,647.75, 2,957.24] | - |
| | 25 to 34 years | Total | 6,807,450 (100.0) | 45,520 (100.0) | 668.68 [662.54, 674.82] | - |
| | | Heterosexual | 6,398,210 (94.0) | 39,315 (86.4) | 614.47 [608.39, 620.54] | - |
| | | LGB+ | 409,240 (6.0) | 6,205 (13.6) | 1,516.23 [1,478.50, 1,553.95] | - |
| | | Gay or Lesbian | 188,300 (2.8) | 2,690 (5.9) | 1,428.57 [1,374.59, 1,482.56] | - |
| | | Bisexual | 172,810 (2.5) | 2,880 (6.3) | 1,666.57 [1,605.70, 1,727.44] | - |
| | | Other | 48,130 (0.7) | 635 (1.4) | 1,319.34 [1,216.72, 1,421.96] | - |
| | 35 to 44 years | Total | 6,608,235 (100.0) | 32,445 (100.0) | 490.98 [485.64, 496.32] | - |
| | | Heterosexual | 6,365,385 (96.3) | 29,895 (92.1) | 469.65 [464.33, 474.97] | - |
| | | LGB+ | 242,850 (3.7) | 2,550 (7.9) | 1,050.03 [1,009.28, 1,090.79] | - |
| | | Gay or Lesbian | 136,095 (2.1) | 1,280 (3.9) | 940.52 [888.99, 992.04] | - |
| | | Bisexual | 80,355 (1.2) | 1,055 (3.3) | 1,312.92 [1,233.70, 1,392.15] | - |
| | | Other | 26,405 (0.4) | 215 (0.7) | 814.24 [705.40, 923.08] | - |
| | 45 to 54 years | Total | 6,805,755 (100.0) | 28,990 (100.0) | 425.96 [421.06, 430.87] | - |
| | | Heterosexual | 6,635,310 (97.5) | 27,470 (94.8) | 414.00 [409.10, 418.89] | - |
| | | LGB+ | 170,445 (2.5) | 1,520 (5.2) | 891.78 [846.95, 936.62] | - |
| | | Gay or Lesbian | 111,990 (1.6) | 845 (2.9) | 754.53 [703.66, 805.41] | - |
| | | Bisexual | 43,725 (0.6) | 560 (1.9) | 1,280.73 [1,174.66, 1,386.81] | - |
| | | Other | 14,730 (0.2) | 120 (0.4) | 814.66 [668.90, 960.43] | - |
| | 55 to 64 years | Total | 6,480,905 (100.0) | 18,795 (100.0) | 290.01 [285.86, 294.15] | - |
| | | Heterosexual | 6,372,060 (98.3) | 18,190 (96.8) | 285.46 [281.32, 289.61] | - |
| | | LGB+ | 108,845 (1.7) | 605 (3.2) | 555.84 [511.54, 600.13] | - |
| | | Gay or Lesbian | 76,585 (1.2) | 365 (1.9) | 476.59 [427.70, 525.49] | - |
| | | Bisexual | 24,360 (0.4) | 195 (1.0) | 800.49 [688.14, 912.85] | - |
| | | Other | 7,895 (0.1) | 45 (0.2) | 569.98 [415.75, 762.68] | - |
| | 65 years and over | Total | 9,390,215 (100.0) | 22,370 (100.0) | 238.23 [235.10, 241.35] | - |

| | | | | | | |
|---|---|---|---|---|---|---|
| | | Heterosexual | 9,327,105 (99.3) | 22,120 (98.9) | 237.16 [234.03, 240.28] | - |
| | | LGB+ | 63,110 (0.7) | 250 (1.1) | 396.13 [347.03, 445.24] | - |
| | | Gay or Lesbian | 42,895 (0.5) | 160 (0.7) | 373.00 [315.21, 430.80] | - |
| | | Bisexual | 15,350 (0.2) | 70 (0.3) | 456.03 [355.50, 576.16] | - |
| | | Other | 4,865 (0.1) | 15 (0.1) | 308.32 [172.57, 508.54] | - |
| Sex | Female | Total | 21,319,600 (100.0) | 120,875 (100.0) | 566.97 [563.77, 570.17] | 831.69 [826.37, 837.01] |
| | | Heterosexual | 20,563,625 (96.5) | 103,845 (85.9) | 504.99 [501.93, 508.07] | 728.94 [723.78, 734.11] |
| | | LGB+ | 755,980 (3.5) | 17,030 (14.1) | 2,252.71 [2,219.00, 2,286.80] | 2,027.76 [1,996.90, 2,058.62] |
| | | Gay or Lesbian | 266,445 (1.2) | 5,330 (4.4) | 2,000.41 [1,947.07, 2,054.85] | 2,222.90 [2,162.41, 2,283.40] |
| | | Bisexual | 397,245 (1.9) | 9,950 (8.2) | 2,504.75 [2,455.77, 2,554.46] | 2,077.11 [2,032.59, 2,121.62] |
| | | Other | 92,290 (0.4) | 1,750 (1.4) | 1,896.20 [1,808.39, 1,987.17] | 1,699.10 [1,617.54, 1,780.65] |
| | Male | Total | 19,878,720 (100.0) | 95,305 (100.0) | 479.43 [476.39, 482.49] | 642.41 [637.77, 647.06] |
| | | Heterosexual | 19,253,900 (96.9) | 88,650 (93.0) | 460.43 [457.40, 463.47] | 616.85 [612.18, 621.51] |
| | | LGB+ | 624,820 (3.1) | 6,655 (7.0) | 1,065.11 [1,039.67, 1,091.01] | 1,189.62 [1,160.27, 1,218.96] |
| | | Gay or Lesbian | 408,110 (2.1) | 3,890 (4.1) | 953.17 [923.45, 983.61] | 1,163.99 [1,124.18, 1,203.80] |
| | | Bisexual | 162,010 (0.8) | 2,215 (2.3) | 1,367.20 [1,310.85, 1,425.35] | 1,342.08 [1,285.74, 1,398.42] |
| | | Other | 54,695 (0.3) | 545 (0.6) | 996.43 [914.52, 1,083.71] | 1,033.27 [945.90, 1,120.63] |
| Region | England: North East | Total | 1,882,045 (100.0) | 12,935 (100.0) | 687.28 [675.49, 699.23] | 1,023.74 [1,004.03, 1,043.46] |
| | | Heterosexual | 1,823,065 (96.9) | 11,645 (90.0) | 638.76 [627.21, 650.47] | 955.06 [935.39, 974.73] |
| | | LGB+ | 58,980 (3.1) | 1,290 (10.0) | 2,187.18 [2,069.44, 2,309.88] | 2,104.45 [1,989.11, 2,219.80] |
| | | Gay or Lesbian | 30,715 (1.6) | 585 (4.5) | 1,904.61 [1,753.37, 2,065.39] | 2,069.98 [1,900.11, 2,239.85] |
| | | Bisexual | 23,150 (1.2) | 590 (4.6) | 2,548.60 [2,347.07, 2,762.80] | 2,281.92 [2,079.75, 2,484.09] |
| | | Other | 5,110 (0.3) | 110 (0.9) | 2,152.64 [1,769.21, 2,594.52] | 2,036.80 [1,652.31, 2,421.28] |
| | England: North West | Total | 5,141,245 (100.0) | 25,885 (100.0) | 503.48 [497.36, 509.65] | 705.06 [695.36, 714.76] |
| | | Heterosexual | 4,969,855 (96.7) | 22,865 (88.3) | 460.07 [454.13, 466.08] | 635.28 [625.82, 644.74] |
| | | LGB+ | 171,390 (3.3) | 3,020 (11.7) | 1,762.06 [1,699.77, 1,826.05] | 1,737.05 [1,675.08, 1,799.01] |
| | | Gay or Lesbian | 90,745 (1.8) | 1,375 (5.3) | 1,515.23 [1,436.19, 1,597.49] | 1,744.64 [1,649.80, 1,839.48] |
| | | Bisexual | 64,905 (1.3) | 1,420 (5.5) | 2,187.81 [2,075.49, 2,304.64] | 1,923.77 [1,817.70, 2,029.84] |
| | | Other | 15,735 (0.3) | 225 (0.9) | 1,429.93 [1,249.18, 1,629.49] | 1,315.69 [1,141.41, 1,489.97] |
| | England: Yorkshire and The Humber | Total | 3,796,000 (100.0) | 28,405 (100.0) | 748.29 [739.61, 757.04] | 960.67 [947.72, 973.63] |
| | | Heterosexual | 3,672,585 (96.7) | 25,835 (91.0) | 703.46 [694.90, 712.09] | 892.07 [879.22, 904.92] |
| | | LGB+ | 123,415 (3.3) | 2,570 (9.0) | 2,082.40 [2,002.67, 2,164.50] | 2,004.05 [1,926.23, 2,081.86] |
| | | Gay or Lesbian | 57,680 (1.5) | 1,025 (3.6) | 1,777.05 [1,669.91, 1,889.25] | 1,955.86 [1,833.77, 2,077.95] |
| | | Bisexual | 52,530 (1.4) | 1,295 (4.6) | 2,465.26 [2,332.80, 2,603.28] | 2,162.78 [2,035.26, 2,290.30] |
| | | Other | 13,205 (0.3) | 255 (0.9) | 1,931.09 [1,701.32, 2,183.23] | 1,802.94 [1,577.50, 2,028.39] |
| | England: East Midlands | Total | 3,407,150 (100.0) | 20,085 (100.0) | 589.50 [581.37, 597.71] | 855.67 [842.38, 868.95] |
| | | Heterosexual | 3,304,925 (97.0) | 17,880 (89.0) | 541.01 [533.11, 549.00] | 778.57 [765.55, 791.59] |
| | | LGB+ | 102,225 (3.0) | 2,205 (11.0) | 2,157.01 [2,067.91, 2,248.96] | 2,066.41 [1,979.72, 2,153.10] |
| | | Gay or Lesbian | 46,555 (1.4) | 835 (4.2) | 1,793.58 [1,673.97, 1,919.47] | 2,054.92 [1,912.94, 2,196.89] |
| | | Bisexual | 44,920 (1.3) | 1,175 (5.9) | 2,615.76 [2,468.32, 2,769.71] | 2,261.43 [2,124.57, 2,398.28] |
| | | Other | 10,745 (0.3) | 190 (0.9) | 1,768.26 [1,525.76, 2,038.36] | 1,616.21 [1,385.14, 1,847.28] |
| | England: West Midlands | Total | 4,059,830 (100.0) | 17,750 (100.0) | 437.21 [430.80, 443.69] | 621.54 [611.29, 631.79] |
| | | Heterosexual | 3,949,510 (97.3) | 15,870 (89.4) | 401.82 [395.59, 408.12] | 565.59 [555.58, 575.59] |
| | | LGB+ | 110,320 (2.7) | 1,880 (10.6) | 1,704.13 [1,627.96, 1,782.95] | 1,654.17 [1,579.30, 1,729.04] |
| | | Gay or Lesbian | 52,630 (1.3) | 745 (4.2) | 1,415.54 [1,315.71, 1,520.94] | 1,614.77 [1,496.33, 1,733.21] |

| | | | | | | |
|---|---|---|---|---|---|---|
| | | Bisexual | 45,650 (1.1) | 980 (5.5) | 2,146.77 [2,014.45, 2,285.50] | 1,832.98 [1,713.13, 1,952.83] |
| | | Other | 12,045 (0.3) | 155 (0.9) | 1,286.84 [1,092.23, 1,506.13] | 1,213.85 [1,021.79, 1,405.91] |
| | England: East of England | Total | 4,428,750 (100.0) | 23,830 (100.0) | 538.08 [531.26, 544.95] | 768.63 [757.27, 779.98] |
| | | Heterosexual | 4,304,030 (97.2) | 21,390 (89.8) | 496.98 [490.34, 503.68] | 699.11 [688.00, 710.22] |
| | | LGB+ | 124,720 (2.8) | 2,440 (10.2) | 1,956.38 [1,879.52, 2,035.58] | 1,917.30 [1,841.04, 1,993.56] |
| | | Gay or Lesbian | 57,060 (1.3) | 870 (3.7) | 1,524.71 [1,425.07, 1,629.49] | 1,786.80 [1,665.02, 1,908.58] |
| | | Bisexual | 53,515 (1.2) | 1,320 (5.5) | 2,466.60 [2,335.31, 2,603.34] | 2,148.08 [2,027.01, 2,269.15] |
| | | Other | 14,145 (0.3) | 250 (1.0) | 1,767.41 [1,555.09, 2,000.62] | 1,662.78 [1,453.65, 1,871.92] |
| | England: London | Total | 5,783,795 (100.0) | 24,535 (100.0) | 424.20 [418.91, 429.54] | 522.06 [514.67, 529.45] |
| | | Heterosexual | 5,512,495 (95.3) | 21,980 (89.6) | 398.73 [393.48, 404.04] | 482.30 [475.01, 489.59] |
| | | LGB+ | 271,300 (4.7) | 2,555 (10.4) | 941.76 [905.60, 979.00] | 1,138.07 [1,091.47, 1,184.66] |
| | | Gay or Lesbian | 141,750 (2.5) | 995 (4.1) | 701.94 [659.00, 746.95] | 1,088.11 [1,004.50, 1,171.71] |
| | | Bisexual | 96,680 (1.7) | 1,225 (5.0) | 1,267.07 [1,197.10, 1,340.06] | 1,243.83 [1,172.20, 1,315.46] |
| | | Other | 32,870 (0.6) | 330 (1.3) | 1,003.95 [898.55, 1,118.33] | 1,117.80 [993.06, 1,242.55] |
| | England: South East | Total | 6,487,835 (100.0) | 32,685 (100.0) | 503.79 [498.34, 509.28] | 751.68 [742.33, 761.03] |
| | | Heterosexual | 6,274,920 (96.7) | 28,815 (88.2) | 459.21 [453.92, 464.54] | 676.01 [666.87, 685.15] |
| | | LGB+ | 212,915 (3.3) | 3,870 (11.8) | 1,817.63 [1,760.81, 1,875.81] | 1,801.37 [1,744.46, 1,858.28] |
| | | Gay or Lesbian | 101,865 (1.6) | 1,375 (4.2) | 1,349.83 [1,279.41, 1,423.11] | 1,689.00 [1,596.57, 1,781.43] |
| | | Bisexual | 88,480 (1.4) | 2,090 (6.4) | 2,362.12 [2,261.92, 2,465.61] | 1,978.94 [1,890.22, 2,067.66] |
| | | Other | 22,575 (0.3) | 405 (1.2) | 1,794.02 [1,623.53, 1,977.54] | 1,631.10 [1,470.08, 1,792.12] |
| | England: South West | Total | 4,051,285 (100.0) | 21,240 (100.0) | 524.28 [517.25, 531.38] | 829.34 [816.79, 841.89] |
| | | Heterosexual | 3,914,520 (96.6) | 18,400 (86.6) | 470.04 [463.28, 476.89] | 738.43 [726.18, 750.69] |
| | | LGB+ | 136,765 (3.4) | 2,840 (13.4) | 2,076.55 [2,000.88, 2,154.36] | 2,005.87 [1,931.68, 2,080.06] |
| | | Gay or Lesbian | 61,230 (1.5) | 980 (4.6) | 1,600.52 [1,501.87, 1,703.95] | 1,971.21 [1,845.36, 2,097.05] |
| | | Bisexual | 61,170 (1.5) | 1,560 (7.3) | 2,550.27 [2,425.27, 2,680.04] | 2,173.78 [2,059.45, 2,288.11] |
| | | Other | 14,365 (0.4) | 295 (1.4) | 2,053.60 [1,825.92, 2,301.83] | 1,843.78 [1,629.44, 2,058.13] |
| | Wales | Total | 2,160,385 (100.0) | 8,825 (100.0) | 408.49 [400.01, 417.11] | 628.33 [613.76, 642.89] |
| | | Heterosexual | 2,091,620 (96.8) | 7,815 (88.6) | 373.63 [365.40, 382.01] | 575.00 [560.61, 589.38] |
| | | LGB+ | 68,765 (3.2) | 1,010 (11.4) | 1,468.77 [1,379.57, 1,562.22] | 1,414.12 [1,326.64, 1,501.60] |
| | | Gay or Lesbian | 34,320 (1.6) | 430 (4.9) | 1,252.91 [1,137.27, 1,377.12] | 1,460.95 [1,319.28, 1,602.62] |
| | | Bisexual | 28,250 (1.3) | 505 (5.7) | 1,787.61 [1,635.08, 1,950.54] | 1,484.37 [1,344.95, 1,623.80] |
| | | Other | 6,190 (0.3) | 75 (0.8) | 1,211.63 [953.03, 1,518.79] | 1,067.23 [834.66, 1,343.47] |
| Ethnic group | Asian, Asian British or Asian Welsh | Total | 3,371,415 (100.0) | 10,655 (100.0) | 316.04 [310.07, 322.10] | 370.13 [362.54, 377.73] |
| | | Heterosexual | 3,302,465 (98.0) | 10,140 (95.2) | 307.04 [301.10, 313.08] | 358.37 [350.81, 365.93] |
| | | LGB+ | 68,950 (2.0) | 515 (4.8) | 746.92 [683.79, 814.30] | 829.76 [756.68, 902.83] |
| | | Gay or Lesbian | 21,095 (0.6) | 160 (1.5) | 758.47 [645.50, 885.53] | 1,005.90 [832.64, 1,179.16] |
| | | Bisexual | 35,490 (1.1) | 275 (2.6) | 774.87 [685.98, 872.07] | 778.53 [685.27, 871.79] |
| | | Other | 12,365 (0.4) | 80 (0.8) | 646.99 [513.02, 805.23] | 790.07 [622.46, 988.07] |
| | Black, Black British, Black Welsh, Caribbean or African | Total | 1,432,710 (100.0) | 5,800 (100.0) | 404.83 [394.48, 415.38] | 494.04 [480.29, 507.80] |
| | | Heterosexual | 1,402,805 (97.9) | 5,405 (93.2) | 385.30 [375.10, 395.71] | 466.77 [453.25, 480.30] |
| | | LGB+ | 29,910 (2.1) | 390 (6.7) | 1,303.91 [1,177.70, 1,439.97] | 1,422.28 [1,280.24, 1,564.33] |
| | | Gay or Lesbian | 9,205 (0.6) | 115 (2.0) | 1,249.32 [1,031.44, 1,499.62] | 1,559.44 [1,264.40, 1,854.48] |
| | | Bisexual | 13,815 (1.0) | 205 (3.5) | 1,483.89 [1,287.70, 1,701.52] | 1,433.83 [1,234.08, 1,633.57] |
| | | Other | 6,885 (0.5) | 70 (1.2) | 1,016.70 [792.57, 1,284.54] | 1,274.84 [993.85, 1,610.63] |
| | | Total | 782,190 (100.0) | 7,225 (100.0) | 923.69 [902.51, 945.24] | 900.12 [878.99, 921.25] |

| | | | | | | |
|---|---|---|---|---|---|---|
| | Mixed or Multiple ethnic groups | Heterosexual | 727,450 (93.0) | 6,025 (83.4) | 828.24 [807.45, 849.42] | 810.92 [790.02, 831.83] |
| | | LGB+ | 54,740 (7.0) | 1,200 (16.6) | 2,192.18 [2,069.89, 2,319.81] | 1,840.84 [1,733.04, 1,948.64] |
| | | Gay or Lesbian | 21,685 (2.8) | 365 (5.1) | 1,683.19 [1,514.92, 1,865.04] | 1,676.59 [1,503.96, 1,849.22] |
| | | Bisexual | 25,760 (3.3) | 700 (9.7) | 2,717.39 [2,519.79, 2,926.37] | 2,050.34 [1,874.55, 2,226.13] |
| | | Other | 7,295 (0.9) | 135 (1.9) | 1,850.58 [1,551.59, 2,190.39] | 1,575.20 [1,298.85, 1,851.54] |
| | White | Total | 34,847,830 (100.0) | 189,755 (100.0) | 544.52 [542.08, 546.98] | 802.71 [798.55, 806.88] |
| | | Heterosexual | 33,648,035 (96.6) | 168,405 (88.7) | 500.49 [498.10, 502.89] | 729.37 [725.28, 733.47] |
| | | LGB+ | 1,199,795 (3.4) | 21,350 (11.3) | 1,779.47 [1,755.68, 1,803.50] | 1,782.23 [1,758.32, 1,806.14] |
| | | Gay or Lesbian | 613,485 (1.8) | 8,505 (4.5) | 1,386.34 [1,357.03, 1,416.12] | 1,691.52 [1,654.08, 1,728.95] |
| | | Bisexual | 474,185 (1.4) | 10,895 (5.7) | 2,297.63 [2,254.68, 2,341.18] | 2,004.86 [1,965.09, 2,044.63] |
| | | Other | 112,125 (0.3) | 1,950 (1.0) | 1,739.13 [1,662.79, 1,818.07] | 1,611.38 [1,538.76, 1,683.99] |
| | Other ethnic group | Total | 764,175 (100.0) | 2,750 (100.0) | 359.87 [346.54, 373.57] | 423.73 [406.46, 441.00] |
| | | Heterosexual | 736,770 (96.4) | 2,525 (91.8) | 342.71 [329.47, 356.35] | 400.89 [383.76, 418.03] |
| | | LGB+ | 27,405 (3.6) | 225 (8.2) | 821.02 [717.24, 935.60] | 964.85 [832.84, 1,096.85] |
| | | Gay or Lesbian | 9,080 (1.2) | 75 (2.7) | 825.99 [649.69, 1,035.39] | 1,281.98 [966.72, 1,656.42] |
| | | Bisexual | 10,005 (1.3) | 90 (3.3) | 899.55 [723.34, 1,105.70] | 951.31 [766.04, 1,168.07] |
| | | Other | 8,320 (1.1) | 55 (2.0) | 661.06 [498.00, 860.46] | 724.10 [532.32, 958.64] |
| Rural/urban classification | Urban | Total | 33,391,510 (100.0) | 182,530 (100.0) | 546.64 [544.13, 549.15] | 740.29 [736.47, 744.11] |
| | | Heterosexual | 32,189,260 (96.4) | 162,130 (88.8) | 503.68 [501.23, 506.14] | 674.08 [670.33, 677.84] |
| | | LGB+ | 1,202,250 (3.6) | 20,400 (11.2) | 1,696.82 [1,673.61, 1,720.27] | 1,694.35 [1,671.09, 1,717.61] |
| | | Gay or Lesbian | 582,585 (1.7) | 7,955 (4.4) | 1,365.47 [1,335.62, 1,395.81] | 1,643.93 [1,606.30, 1,681.55] |
| | | Bisexual | 488,625 (1.5) | 10,435 (5.7) | 2,135.58 [2,094.80, 2,176.96] | 1,861.38 [1,823.87, 1,898.90] |
| | | Other | 131,035 (0.4) | 2,010 (1.1) | 1,533.94 [1,467.61, 1,602.50] | 1,465.96 [1,401.33, 1,530.59] |
| | Rural | Total | 7,806,815 (100.0) | 33,650 (100.0) | 431.03 [426.44, 435.66] | 717.32 [708.14, 726.50] |
| | | Heterosexual | 7,628,265 (97.7) | 30,370 (90.3) | 398.12 [393.66, 402.63] | 653.47 [644.50, 662.43] |
| | | LGB+ | 178,550 (2.3) | 3,285 (9.8) | 1,839.82 [1,777.44, 1,903.83] | 1,882.02 [1,816.27, 1,947.78] |
| | | Gay or Lesbian | 91,970 (1.2) | 1,265 (3.8) | 1,375.45 [1,300.69, 1,453.39] | 1,775.36 [1,672.73, 1,878.00] |
| | | Bisexual | 70,630 (0.9) | 1,730 (5.1) | 2,449.38 [2,335.31, 2,567.59] | 2,123.23 [2,018.08, 2,228.38] |
| | | Other | 15,950 (0.2) | 285 (0.8) | 1,786.83 [1,585.39, 2,006.79] | 1,629.27 [1,434.07, 1,824.46] |
| Country of birth | Born in the UK | Total | 33,751,745 (100.0) | 194,515 (100.0) | 576.31 [573.75, 578.88] | 823.03 [818.90, 827.15] |
| | | Heterosexual | 32,638,615 (96.7) | 172,660 (88.8) | 529.01 [526.51, 531.51] | 748.77 [744.72, 752.82] |
| | | LGB+ | 1,113,130 (3.3) | 21,855 (11.2) | 1,963.38 [1,937.44, 1,989.59] | 1,916.80 [1,891.32, 1,942.28] |
| | | Gay or Lesbian | 563,890 (1.7) | 8,605 (4.4) | 1,526.01 [1,493.93, 1,558.60] | 1,804.94 [1,765.73, 1,844.15] |
| | | Bisexual | 447,290 (1.3) | 11,225 (5.8) | 2,509.56 [2,463.34, 2,556.42] | 2,154.51 [2,111.82, 2,197.19] |
| | | Other | 101,950 (0.3) | 2,025 (1.0) | 1,986.27 [1,900.69, 2,074.71] | 1,783.44 [1,703.79, 1,863.09] |
| | Born outside the UK | Total | 7,446,575 (100.0) | 21,665 (100.0) | 290.94 [287.08, 294.84] | 378.35 [372.18, 384.52] |
| | | Heterosexual | 7,178,910 (96.4) | 19,835 (91.6) | 276.30 [272.46, 280.17] | 351.90 [345.81, 357.99] |
| | | LGB+ | 267,665 (3.6) | 1,825 (8.4) | 681.82 [650.90, 713.84] | 808.51 [769.23, 847.79] |
| | | Gay or Lesbian | 110,665 (1.5) | 615 (2.8) | 555.73 [512.67, 601.44] | 797.90 [717.95, 877.85] |
| | | Bisexual | 111,965 (1.5) | 940 (4.3) | 839.55 [786.73, 894.98] | 863.85 [807.26, 920.43] |
| | | Other | 45,040 (0.6) | 270 (1.2) | 599.47 [530.09, 675.40] | 698.72 [610.88, 786.55] |
| English language proficiency | Main language is English | Total | 37,542,140 (100.0) | 207,015 (100.0) | 551.42 [549.05, 553.80] | 784.21 [780.39, 788.04] |
| | | Heterosexual | 36,283,540 (96.6) | 183,945 (88.9) | 506.97 [504.65, 509.29] | 713.55 [709.79, 717.31] |
| | | LGB+ | 1,258,600 (3.4) | 23,070 (11.1) | 1,832.99 [1,809.41, 1,856.80] | 1,816.86 [1,793.41, 1,840.32] |
| | | Gay or Lesbian | 633,445 (1.7) | 9,040 (4.4) | 1,427.12 [1,397.85, 1,456.84] | 1,727.21 [1,690.32, 1,764.11] |

| | | | | | | |
|---|---|---|---|---|---|---|
| | | Bisexual | 505,645 (1.3) | 11,870 (5.7) | 2,347.50 [2,305.45, 2,390.11] | 2,025.85 [1,987.28, 2,064.41] |
| | | Other | 119,505 (0.3) | 2,160 (1.0) | 1,807.46 [1,732.03, 1,885.32] | 1,650.68 [1,580.04, 1,721.32] |
| | Main language is not English | Total | 3,656,185 (100.0) | 9,165 (100.0) | 250.67 [245.57, 255.86] | 304.47 [296.83, 312.11] |
| | | Heterosexual | 3,533,985 (96.7) | 8,555 (93.3) | 242.08 [236.98, 247.26] | 289.62 [282.02, 297.22] |
| | | LGB+ | 122,200 (3.3) | 610 (6.7) | 499.18 [460.35, 540.42] | 588.23 [537.44, 639.02] |
| | | Gay or Lesbian | 41,110 (1.1) | 180 (2.0) | 437.85 [376.22, 506.70] | 641.90 [521.44, 762.35] |
| | | Bisexual | 53,610 (1.5) | 295 (3.2) | 550.27 [489.26, 616.78] | 593.49 [522.92, 664.05] |
| | | Other | 27,485 (0.8) | 135 (1.5) | 491.18 [411.82, 581.37] | 538.85 [439.75, 637.95] |
| Religion | No religion | Total | 15,126,775 (100.0) | 107,795 (100.0) | 712.61 [708.36, 716.88] | 866.83 [861.38, 872.28] |
| | | Heterosexual | 14,258,160 (94.3) | 91,880 (85.2) | 644.40 [640.24, 648.58] | 784.77 [779.36, 790.19] |
| | | LGB+ | 868,615 (5.7) | 15,915 (14.8) | 1,832.23 [1,803.87, 1,860.92] | 1,681.91 [1,655.51, 1,708.31] |
| | | Gay or Lesbian | 417,475 (2.8) | 6,075 (5.6) | 1,455.18 [1,418.81, 1,492.24] | 1,619.35 [1,577.80, 1,660.90] |
| | | Bisexual | 370,120 (2.4) | 8,380 (7.8) | 2,264.13 [2,215.91, 2,313.14] | 1,863.85 [1,819.05, 1,908.65] |
| | | Other | 81,025 (0.5) | 1,460 (1.4) | 1,801.91 [1,710.66, 1,896.77] | 1,537.88 [1,455.20, 1,620.55] |
| | Christian | Total | 20,881,940 (100.0) | 85,930 (100.0) | 411.50 [408.76, 414.26] | 651.08 [645.44, 656.72] |
| | | Heterosexual | 20,524,925 (98.3) | 80,570 (93.8) | 392.55 [389.84, 395.27] | 609.67 [604.14, 615.21] |
| | | LGB+ | 357,015 (1.7) | 5,360 (6.2) | 1,501.34 [1,461.41, 1,542.08] | 1,818.26 [1,767.63, 1,868.89] |
| | | Gay or Lesbian | 208,870 (1.0) | 2,450 (2.9) | 1,172.98 [1,126.99, 1,220.37] | 1,711.90 [1,635.29, 1,788.51] |
| | | Bisexual | 115,935 (0.6) | 2,510 (2.9) | 2,165.01 [2,081.13, 2,251.40] | 2,143.62 [2,058.93, 2,228.31] |
| | | Other | 32,210 (0.2) | 395 (0.5) | 1,226.33 [1,108.36, 1,353.43] | 1,330.45 [1,197.30, 1,463.60] |
| | Buddhist | Total | 202,525 (100.0) | 970 (100.0) | 478.95 [449.28, 510.07] | 683.00 [630.02, 735.98] |
| | | Heterosexual | 186,835 (92.3) | 715 (73.7) | 382.69 [355.15, 411.80] | 504.00 [456.64, 551.36] |
| | | LGB+ | 15,690 (7.7) | 255 (26.3) | 1,625.24 [1,431.86, 1,837.45] | 2,087.01 [1,805.51, 2,368.51] |
| | | Gay or Lesbian | 6,925 (3.4) | 85 (8.8) | 1,227.44 [980.43, 1,517.74] | 1,821.19 [1,351.61, 2,373.10] |
| | | Bisexual | 6,670 (3.3) | 125 (12.9) | 1,874.06 [1,559.95, 2,232.86] | 2,076.86 [1,693.58, 2,460.14] |
| | | Other | 2,095 (1.0) | 45 (4.6) | 2,147.97 [1,566.75, 2,874.15] | 2,540.72 [1,833.42, 3,424.44] |
| | Hindu | Total | 693,140 (100.0) | 1,745 (100.0) | 251.75 [240.08, 263.85] | 322.45 [304.38, 340.51] |
| | | Heterosexual | 680,310 (98.1) | 1,675 (96.0) | 246.21 [234.56, 258.29] | 312.20 [294.23, 330.17] |
| | | LGB+ | 12,830 (1.9) | 75 (4.3) | 584.57 [459.80, 732.76] | 736.94 [570.06, 935.14] |
| | | Gay or Lesbian | 2,335 (0.3) | 20 (1.1) | 856.53 [523.19, 1,322.84] | 1,083.38 [558.03, 1,818.32] |
| | | Bisexual | 8,460 (1.2) | 50 (2.9) | 591.02 [438.66, 779.18] | 732.34 [532.98, 978.58] |
| | | Other | 2,030 (0.3) | 10 (0.6) | 492.61 [236.23, 905.93] | 510.35 [194.53, 1,019.47] |
| | Jewish | Total | 178,245 (100.0) | 835 (100.0) | 468.46 [437.22, 501.34] | 581.88 [530.04, 633.71] |
| | | Heterosexual | 169,850 (95.3) | 720 (86.2) | 423.90 [393.50, 456.03] | 483.65 [434.77, 532.54] |
| | | LGB+ | 8,395 (4.7) | 115 (13.8) | 1,369.86 [1,130.96, 1,644.31] | 1,523.04 [1,241.49, 1,804.60] |
| | | Gay or Lesbian | 4,135 (2.3) | 35 (4.2) | 846.43 [589.57, 1,177.18] | 1,114.15 [753.43, 1,578.65] |
| | | Bisexual | 3,275 (1.8) | 65 (7.8) | 1,984.73 [1,531.77, 2,529.70] | 1,955.76 [1,504.87, 2,498.25] |
| | | Other | 985 (0.6) | 10 (1.2) | 1,015.23 [486.84, 1,867.04] | 1,309.90 [661.95, 2,354.47] |
| | Muslim | Total | 2,162,660 (100.0) | 8,275 (100.0) | 382.63 [374.43, 390.97] | 419.60 [410.10, 429.11] |
| | | Heterosexual | 2,131,220 (98.5) | 8,020 (96.9) | 376.31 [368.12, 384.64] | 412.38 [402.89, 421.87] |
| | | LGB+ | 31,440 (1.5) | 255 (3.1) | 811.07 [714.57, 916.97] | 988.78 [860.44, 1,117.13] |
| | | Gay or Lesbian | 4,990 (0.2) | 60 (0.7) | 1,202.40 [917.56, 1,547.73] | 1,550.04 [1,162.79, 2,019.53] |
| | | Bisexual | 15,405 (0.7) | 130 (1.6) | 843.88 [705.06, 1,002.04] | 994.87 [817.22, 1,172.52] |
| | | Other | 11,040 (0.5) | 60 (0.7) | 543.48 [414.73, 699.56] | 715.70 [532.22, 938.15] |
| | Sikh | Total | 354,395 (100.0) | 1,230 (100.0) | 347.07 [327.94, 367.02] | 402.34 [376.49, 428.20] |

| | | | | | | |
|---|---|---|---|---|---|---|
| | | Heterosexual | 348,995 (98.5) | 1,185 (96.3) | 339.55 [320.49, 359.44] | 392.26 [366.52, 418.01] |
| | | LGB+ | 5,400 (1.5) | 45 (3.7) | 833.33 [607.84, 1,115.07] | 921.77 [653.27, 1,257.23] |
| | | Gay or Lesbian | 1,020 (0.3) | 20 (1.6) | 1,960.78 [1,197.70, 3,028.27] | 2,318.38 [1,320.64, 3,714.15] |
| | | Bisexual | 2,885 (0.8) | 15 (1.2) | 519.93 [291.00, 857.55] | 475.04 [236.95, 826.18] |
| | | Other | 1,490 (0.4) | 15 (1.2) | 1,006.71 [563.45, 1,660.42] | 934.21 [404.35, 1,715.62] |
| | Other religion | Total | 256,045 (100.0) | 2,275 (100.0) | 888.52 [852.38, 925.79] | 1,294.57 [1,233.89, 1,355.25] |
| | | Heterosexual | 214,080 (83.6) | 1,330 (58.5) | 621.26 [588.32, 655.57] | 836.53 [779.30, 893.75] |
| | | LGB+ | 41,965 (16.4) | 940 (41.3) | 2,239.96 [2,099.04, 2,387.86] | 2,259.94 [2,114.19, 2,405.70] |
| | | Gay or Lesbian | 10,150 (4.0) | 205 (9.0) | 2,019.70 [1,752.67, 2,315.92] | 2,380.58 [2,045.55, 2,715.61] |
| | | Bisexual | 21,115 (8.2) | 510 (22.4) | 2,415.34 [2,210.24, 2,634.36] | 2,298.51 [2,094.74, 2,502.28] |
| | | Other | 10,700 (4.2) | 225 (9.9) | 2,102.80 [1,837.00, 2,396.26] | 2,094.19 [1,815.16, 2,373.21] |
| | Not answered | Total | 1,342,600 (100.0) | 7,125 (100.0) | 530.69 [518.44, 543.15] | 731.12 [711.94, 750.29] |
| | | Heterosexual | 1,303,150 (97.1) | 6,400 (89.8) | 491.12 [479.16, 503.30] | 669.51 [650.73, 688.30] |
| | | LGB+ | 39,450 (2.9) | 725 (10.2) | 1,837.77 [1,706.41, 1,976.55] | 1,864.61 [1,728.99, 2,000.22] |
| | | Gay or Lesbian | 18,650 (1.4) | 265 (3.7) | 1,420.91 [1,254.97, 1,602.69] | 1,677.52 [1,467.10, 1,887.94] |
| | | Bisexual | 15,390 (1.1) | 385 (5.4) | 2,501.62 [2,257.95, 2,764.43] | 2,261.62 [2,025.82, 2,497.43] |
| | | Other | 5,405 (0.4) | 75 (1.1) | 1,387.60 [1,091.44, 1,739.37] | 1,375.71 [1,079.37, 1,727.68] |
| IMD/WIMD decile group | 1 (most deprived) | Total | 3,697,945 (100.0) | 31,680 (100.0) | 856.69 [847.28, 866.18] | 1,055.26 [1,042.71, 1,067.82] |
| | | Heterosexual | 3,552,730 (96.1) | 28,180 (89.0) | 793.19 [783.96, 802.51] | 970.43 [958.06, 982.80] |
| | | LGB+ | 145,215 (3.9) | 3,500 (11.0) | 2,410.22 [2,331.02, 2,491.42] | 2,375.41 [2,296.60, 2,454.23] |
| | | Gay or Lesbian | 67,800 (1.8) | 1,510 (4.8) | 2,227.14 [2,116.21, 2,342.37] | 2,454.04 [2,327.84, 2,580.25] |
| | | Bisexual | 60,455 (1.6) | 1,710 (5.4) | 2,828.55 [2,696.06, 2,965.87] | 2,580.41 [2,453.47, 2,707.36] |
| | | Other | 16,960 (0.5) | 275 (0.9) | 1,621.46 [1,435.47, 1,824.87] | 1,623.15 [1,429.82, 1,816.48] |
| | 2 | Total | 3,911,785 (100.0) | 28,020 (100.0) | 716.30 [707.93, 724.73] | 901.97 [890.44, 913.51] |
| | | Heterosexual | 3,752,330 (95.9) | 24,915 (88.9) | 663.99 [655.77, 672.28] | 829.39 [818.00, 840.77] |
| | | LGB+ | 159,455 (4.1) | 3,110 (11.1) | 1,950.39 [1,882.44, 2,020.17] | 1,971.84 [1,902.25, 2,041.43] |
| | | Gay or Lesbian | 75,145 (1.9) | 1,275 (4.6) | 1,696.72 [1,604.85, 1,792.47] | 1,960.30 [1,848.18, 2,072.42] |
| | | Bisexual | 65,475 (1.7) | 1,525 (5.4) | 2,329.13 [2,213.69, 2,449.04] | 2,096.47 [1,987.78, 2,205.17] |
| | | Other | 18,835 (0.5) | 310 (1.1) | 1,645.87 [1,467.74, 1,839.67] | 1,662.19 [1,474.64, 1,849.75] |
| | 3 | Total | 4,064,925 (100.0) | 25,745 (100.0) | 633.35 [625.63, 641.13] | 818.62 [807.59, 829.66] |
| | | Heterosexual | 3,893,450 (95.8) | 22,775 (88.5) | 584.96 [577.38, 592.60] | 748.46 [737.59, 759.33] |
| | | LGB+ | 171,475 (4.2) | 2,975 (11.6) | 1,734.95 [1,673.16, 1,798.43] | 1,799.04 [1,733.89, 1,864.18] |
| | | Gay or Lesbian | 82,610 (2.0) | 1,230 (4.8) | 1,488.92 [1,406.87, 1,574.52] | 1,830.84 [1,721.75, 1,939.92] |
| | | Bisexual | 69,120 (1.7) | 1,470 (5.7) | 2,126.74 [2,019.39, 2,238.30] | 1,927.90 [1,825.46, 2,030.34] |
| | | Other | 19,745 (0.5) | 270 (1.0) | 1,367.43 [1,209.18, 1,540.65] | 1,383.43 [1,216.68, 1,550.18] |
| | 4 | Total | 4,152,080 (100.0) | 23,655 (100.0) | 569.71 [562.48, 577.02] | 765.38 [754.46, 776.30] |
| | | Heterosexual | 3,985,065 (96.0) | 20,885 (88.3) | 524.08 [517.00, 531.24] | 695.38 [684.64, 706.11] |
| | | LGB+ | 167,015 (4.0) | 2,775 (11.7) | 1,661.53 [1,600.28, 1,724.52] | 1,700.98 [1,637.48, 1,764.49] |
| | | Gay or Lesbian | 81,310 (2.0) | 1,095 (4.6) | 1,346.70 [1,268.10, 1,428.89] | 1,708.87 [1,601.58, 1,816.16] |
| | | Bisexual | 67,530 (1.6) | 1,400 (5.9) | 2,073.15 [1,965.97, 2,184.66] | 1,859.85 [1,757.85, 1,961.86] |
| | | Other | 18,175 (0.4) | 275 (1.2) | 1,513.07 [1,339.50, 1,702.88] | 1,484.52 [1,308.10, 1,660.94] |
| | 5 | Total | 4,195,725 (100.0) | 21,445 (100.0) | 511.12 [504.30, 518.00] | 712.45 [701.62, 723.28] |
| | | Heterosexual | 4,046,600 (96.4) | 19,030 (88.7) | 470.27 [463.61, 477.00] | 645.86 [635.24, 656.48] |
| | | LGB+ | 149,120 (3.6) | 2,415 (11.3) | 1,619.50 [1,555.55, 1,685.41] | 1,642.93 [1,577.39, 1,708.48] |
| | | Gay or Lesbian | 72,645 (1.7) | 935 (4.4) | 1,287.08 [1,205.89, 1,372.30] | 1,601.61 [1,493.87, 1,709.35] |

| | | | | | | |
|---|---|---|---|---|---|---|
| | | Bisexual | 60,800 (1.4) | 1,230 (5.7) | 2,023.03 [1,911.54, 2,139.32] | 1,725.40 [1,624.83, 1,825.98] |
| | | Other | 15,675 (0.4) | 255 (1.2) | 1,626.79 [1,433.23, 1,839.21] | 1,557.27 [1,365.06, 1,749.48] |
| | 6 | Total | 4,290,295 (100.0) | 20,615 (100.0) | 480.50 [473.97, 487.11] | 686.56 [675.75, 697.36] |
| | | Heterosexual | 4,145,555 (96.6) | 18,340 (89.0) | 442.40 [436.02, 448.85] | 622.90 [612.30, 633.51] |
| | | LGB+ | 144,745 (3.4) | 2,270 (11.0) | 1,568.28 [1,504.42, 1,634.15] | 1,587.88 [1,522.54, 1,653.22] |
| | | Gay or Lesbian | 72,075 (1.7) | 825 (4.0) | 1,144.64 [1,067.86, 1,225.49] | 1,441.15 [1,338.04, 1,544.26] |
| | | Bisexual | 57,895 (1.3) | 1,240 (6.0) | 2,141.81 [2,024.24, 2,264.42] | 1,860.03 [1,751.24, 1,968.83] |
| | | Other | 14,770 (0.3) | 210 (1.0) | 1,421.80 [1,235.99, 1,627.66] | 1,352.59 [1,167.95, 1,537.22] |
| | 7 | Total | 4,237,365 (100.0) | 18,425 (100.0) | 434.82 [428.57, 441.15] | 643.91 [633.01, 654.80] |
| | | Heterosexual | 4,111,035 (97.0) | 16,505 (89.6) | 401.48 [395.38, 407.65] | 583.53 [572.87, 594.19] |
| | | LGB+ | 126,325 (3.0) | 1,920 (10.4) | 1,519.89 [1,452.66, 1,589.43] | 1,529.11 [1,460.67, 1,597.56] |
| | | Gay or Lesbian | 63,370 (1.5) | 665 (3.6) | 1,049.39 [971.14, 1,132.27] | 1,337.23 [1,231.24, 1,443.22] |
| | | Bisexual | 50,655 (1.2) | 1,060 (5.8) | 2,092.59 [1,968.49, 2,222.45] | 1,771.37 [1,659.32, 1,883.42] |
| | | Other | 12,305 (0.3) | 195 (1.1) | 1,584.72 [1,370.09, 1,823.44] | 1,474.06 [1,266.49, 1,681.64] |
| | 8 | Total | 4,258,970 (100.0) | 17,725 (100.0) | 416.18 [410.08, 422.35] | 619.92 [609.16, 630.68] |
| | | Heterosexual | 4,141,200 (97.2) | 15,900 (89.7) | 383.95 [378.00, 389.96] | 562.05 [551.52, 572.57] |
| | | LGB+ | 117,765 (2.8) | 1,830 (10.3) | 1,553.94 [1,483.55, 1,626.81] | 1,526.07 [1,455.84, 1,596.30] |
| | | Gay or Lesbian | 59,885 (1.4) | 670 (3.8) | 1,118.81 [1,035.69, 1,206.83] | 1,391.83 [1,282.30, 1,501.35] |
| | | Bisexual | 46,730 (1.1) | 985 (5.6) | 2,107.85 [1,978.26, 2,243.71] | 1,723.85 [1,609.11, 1,838.60] |
| | | Other | 11,150 (0.3) | 175 (1.0) | 1,569.51 [1,345.58, 1,820.05] | 1,415.79 [1,201.69, 1,629.89] |
| | 9 | Total | 4,222,750 (100.0) | 15,780 (100.0) | 373.69 [367.88, 379.57] | 569.90 [559.27, 580.52] |
| | | Heterosexual | 4,115,680 (97.5) | 14,195 (90.0) | 344.90 [339.25, 350.62] | 517.48 [507.09, 527.88] |
| | | LGB+ | 107,070 (2.5) | 1,590 (10.1) | 1,485.01 [1,412.91, 1,559.84] | 1,447.25 [1,375.68, 1,518.82] |
| | | Gay or Lesbian | 54,355 (1.3) | 585 (3.7) | 1,076.26 [990.80, 1,167.12] | 1,340.73 [1,228.52, 1,452.95] |
| | | Bisexual | 42,540 (1.0) | 825 (5.2) | 1,939.35 [1,809.26, 2,076.33] | 1,642.98 [1,522.65, 1,763.30] |
| | | Other | 10,175 (0.2) | 175 (1.1) | 1,719.90 [1,474.51, 1,994.45] | 1,499.56 [1,270.85, 1,728.27] |
| | 10 (least deprived) | Total | 4,166,490 (100.0) | 13,080 (100.0) | 313.93 [308.58, 319.36] | 498.21 [487.87, 508.56] |
| | | Heterosexual | 4,073,880 (97.8) | 11,775 (90.0) | 289.04 [283.84, 294.31] | 450.39 [440.31, 460.47] |
| | | LGB+ | 92,610 (2.2) | 1,305 (10.0) | 1,409.14 [1,333.71, 1,487.71] | 1,284.74 [1,213.00, 1,356.47] |
| | | Gay or Lesbian | 45,365 (1.1) | 435 (3.3) | 958.89 [870.88, 1,053.38] | 1,138.98 [1,028.76, 1,249.19] |
| | | Bisexual | 38,050 (0.9) | 720 (5.5) | 1,892.25 [1,756.54, 2,035.66] | 1,447.61 [1,330.38, 1,564.84] |
| | | Other | 9,195 (0.2) | 150 (1.1) | 1,631.32 [1,380.71, 1,914.27] | 1,286.16 [1,067.15, 1,505.17] |
| Highest level of qualification (≥25 years) | No qualifications | Total | 6,640,315 (100.0) | 36,870 (100.0) | 555.24 [549.59, 560.94] | 1,062.43 [1,047.11, 1,077.76] |
| | | Heterosexual | 6,552,600 (98.7) | 35,315 (95.8) | 538.95 [533.34, 544.60] | 1,022.11 [1,006.85, 1,037.37] |
| | | LGB+ | 87,720 (1.3) | 1,555 (4.2) | 1,772.69 [1,685.66, 1,863.04] | 2,164.22 [2,049.92, 2,278.53] |
| | | Gay or Lesbian | 40,610 (0.6) | 815 (2.2) | 2,006.89 [1,871.46, 2,149.54] | 2,953.36 [2,731.16, 3,175.56] |
| | | Bisexual | 29,125 (0.4) | 625 (1.7) | 2,145.92 [1,980.96, 2,320.96] | 2,397.36 [2,201.65, 2,593.07] |
| | | Other | 17,980 (0.3) | 115 (0.3) | 639.60 [528.06, 767.74] | 677.51 [550.38, 804.64] |
| | Level 1 (one to four GCSE passes (grade A* to C or grade 4 and above) and any other GCSEs at other grades, or equivalent qualifications) | Total | 3,547,145 (100.0) | 19,265 (100.0) | 543.11 [535.47, 550.84] | 865.77 [850.63, 880.91] |
| | | Heterosexual | 3,480,285 (98.1) | 18,015 (93.5) | 517.63 [510.10, 525.24] | 818.77 [803.79, 833.75] |
| | | LGB+ | 66,860 (1.9) | 1,250 (6.5) | 1,869.58 [1,767.36, 1,976.17] | 2,083.05 [1,964.49, 2,201.60] |
| | | Gay or Lesbian | 38,635 (1.1) | 615 (3.2) | 1,591.82 [1,468.48, 1,722.75] | 1,925.97 [1,764.01, 2,087.94] |
| | | Bisexual | 22,335 (0.6) | 555 (2.9) | 2,484.89 [2,282.43, 2,700.49] | 2,477.34 [2,270.17, 2,684.52] |
| | | Other | 5,895 (0.2) | 80 (0.4) | 1,357.08 [1,076.08, 1,689.01] | 1,455.03 [1,150.66, 1,814.56] |

| | | | | | | |
|---|---|---|---|---|---|---|
| | Level 2 (five or more GCSE passes (grade A* to C or grade 4 and above) or equivalent qualifications) | Total | 4,308,070 (100.0) | 21,885 (100.0) | 508.00 [501.29, 514.78] | 698.42 [687.77, 709.07] |
| | | Heterosexual | 4,199,885 (97.5) | 20,095 (91.8) | 478.47 [471.87, 485.13] | 650.56 [640.08, 661.04] |
| | | LGB+ | 108,190 (2.5) | 1,790 (8.2) | 1,654.50 [1,578.73, 1,732.96] | 1,690.34 [1,611.74, 1,768.93] |
| | | Gay or Lesbian | 63,675 (1.5) | 825 (3.8) | 1,295.64 [1,208.73, 1,387.15] | 1,442.55 [1,341.69, 1,543.42] |
| | | Bisexual | 35,980 (0.8) | 825 (3.8) | 2,292.94 [2,139.13, 2,454.89] | 2,076.70 [1,933.27, 2,220.12] |
| | | Other | 8,535 (0.2) | 140 (0.6) | 1,640.30 [1,379.85, 1,935.62] | 1,560.90 [1,302.23, 1,819.57] |
| | Apprenticeship | Total | 2,013,560 (100.0) | 8,110 (100.0) | 402.77 [394.05, 411.63] | 630.45 [613.04, 647.86] |
| | | Heterosexual | 1,986,265 (98.6) | 7,745 (95.5) | 389.93 [381.29, 398.71] | 602.99 [585.74, 620.25] |
| | | LGB+ | 27,295 (1.4) | 360 (4.4) | 1,318.92 [1,186.18, 1,462.46] | 1,425.23 [1,276.43, 1,574.02] |
| | | Gay or Lesbian | 13,950 (0.7) | 165 (2.0) | 1,182.80 [1,009.20, 1,377.68] | 1,374.66 [1,157.80, 1,591.51] |
| | | Bisexual | 9,590 (0.5) | 170 (2.1) | 1,772.68 [1,516.22, 2,060.10] | 1,750.90 [1,485.56, 2,016.24] |
| | | Other | 3,760 (0.2) | 25 (0.3) | 664.89 [430.28, 981.51] | 785.54 [518.90, 1,145.38] |
| | Level 3 (two or more A Levels or equivalent qualifications) | Total | 5,447,045 (100.0) | 22,920 (100.0) | 420.78 [415.35, 426.26] | 496.72 [489.84, 503.59] |
| | | Heterosexual | 5,279,280 (96.9) | 20,495 (89.4) | 388.22 [382.92, 393.57] | 454.04 [447.33, 460.74] |
| | | LGB+ | 167,765 (3.1) | 2,420 (10.6) | 1,442.49 [1,385.59, 1,501.14] | 1,355.41 [1,300.88, 1,409.95] |
| | | Gay or Lesbian | 96,380 (1.8) | 1,150 (5.0) | 1,193.19 [1,125.22, 1,264.20] | 1,201.03 [1,131.43, 1,270.62] |
| | | Bisexual | 56,845 (1.0) | 1,025 (4.5) | 1,803.15 [1,694.44, 1,917.01] | 1,613.00 [1,507.59, 1,718.42] |
| | | Other | 14,540 (0.3) | 245 (1.1) | 1,685.01 [1,480.60, 1,909.74] | 1,476.60 [1,280.82, 1,672.37] |
| | Level 4 and above (Higher National Certificate, Higher National Diploma, Bachelor’s degree, or post-graduate qualifications) | Total | 13,103,645 (100.0) | 34,990 (100.0) | 267.02 [264.23, 269.84] | 296.77 [293.39, 300.16] |
| | | Heterosexual | 12,583,195 (96.0) | 31,420 (89.8) | 249.70 [246.94, 252.47] | 274.20 [270.86, 277.54] |
| | | LGB+ | 520,445 (4.0) | 3,570 (10.2) | 685.95 [663.63, 708.83] | 671.19 [649.11, 693.27] |
| | | Gay or Lesbian | 294,395 (2.2) | 1,680 (4.8) | 570.66 [543.70, 598.62] | 591.81 [563.19, 620.43] |
| | | Bisexual | 177,250 (1.4) | 1,490 (4.3) | 840.62 [798.47, 884.41] | 787.02 [744.58, 829.46] |
| | | Other | 48,805 (0.4) | 405 (1.2) | 829.83 [750.97, 914.72] | 769.61 [691.41, 847.81] |
| | Other qualifications | Total | 1,032,785 (100.0) | 4,085 (100.0) | 395.53 [383.50, 407.85] | 606.38 [580.90, 631.86] |
| | | Heterosexual | 1,016,560 (98.4) | 3,900 (95.5) | 383.65 [371.70, 395.88] | 582.25 [556.83, 607.68] |
| | | LGB+ | 16,220 (1.6) | 180 (4.4) | 1,109.74 [953.54, 1,284.23] | 1,248.22 [1,062.37, 1,434.06] |
| | | Gay or Lesbian | 8,225 (0.8) | 90 (2.2) | 1,094.22 [879.89, 1,344.99] | 1,383.90 [1,095.29, 1,721.57] |
| | | Bisexual | 5,485 (0.5) | 75 (1.8) | 1,367.37 [1,075.52, 1,714.00] | 1,354.79 [1,061.37, 1,703.29] |
| | | Other | 2,515 (0.2) | 20 (0.5) | 795.23 [485.75, 1,228.17] | 753.92 [448.53, 1,181.14] |
| NS-SEC | Class 1 (Higher managerial, administrative and professional occupations) | Total | 5,565,200 (100.0) | 10,945 (100.0) | 196.67 [193.00, 200.39] | 318.13 [298.52, 337.74] |
| | | Heterosexual | 5,373,260 (96.6) | 10,005 (91.4) | 186.20 [182.57, 189.88] | 287.78 [269.09, 306.47] |
| | | LGB+ | 191,940 (3.4) | 940 (8.6) | 489.74 [458.93, 522.07] | 932.16 [728.75, 1,135.56] |
| | | Gay or Lesbian | 118,370 (2.1) | 475 (4.3) | 401.28 [366.00, 439.05] | 818.25 [502.58, 1,133.93] |
| | | Bisexual | 59,135 (1.1) | 375 (3.4) | 634.14 [571.58, 701.69] | 1,088.26 [785.59, 1,390.92] |
| | | Other | 14,435 (0.3) | 90 (0.8) | 623.48 [501.36, 766.37] | 833.59 [367.31, 1,379.12] |
| | Class 2 (Lower managerial, administrative and professional occupations) | Total | 8,415,665 (100.0) | 24,790 (100.0) | 294.57 [290.91, 298.26] | 483.06 [468.20, 497.93] |
| | | Heterosexual | 8,100,895 (96.3) | 22,210 (89.6) | 274.17 [270.57, 277.80] | 431.23 [416.99, 445.47] |
| | | LGB+ | 314,770 (3.7) | 2,580 (10.4) | 819.65 [788.32, 851.90] | 1,277.53 [1,166.18, 1,388.88] |
| | | Gay or Lesbian | 184,505 (2.2) | 1,200 (4.8) | 650.39 [614.11, 688.26] | 1,146.20 [971.08, 1,321.31] |
| | | Bisexual | 104,755 (1.2) | 1,120 (4.5) | 1,069.16 [1,007.45, 1,133.66] | 1,434.75 [1,274.72, 1,594.78] |
| | | Other | 25,510 (0.3) | 260 (1.0) | 1,019.21 [899.08, 1,150.92] | 1,310.73 [961.55, 1,659.92] |
| | Class 3 (Intermediate occupations) | Total | 4,821,360 (100.0) | 19,600 (100.0) | 406.52 [400.85, 412.26] | 641.45 [628.20, 654.69] |
| | | Heterosexual | 4,682,910 (97.1) | 17,630 (89.9) | 376.48 [370.94, 382.07] | 582.23 [569.32, 595.14] |

| | | | | | | |
|---|---|---|---|---|---|---|
| | | LGB+ | 138,450 (2.9) | 1,975 (10.1) | 1,426.51 [1,364.28, 1,490.84] | 1,679.74 [1,582.41, 1,777.07] |
| | | Gay or Lesbian | 72,050 (1.5) | 850 (4.3) | 1,179.74 [1,101.75, 1,261.79] | 1,520.76 [1,377.02, 1,664.50] |
| | | Bisexual | 53,715 (1.1) | 965 (4.9) | 1,796.52 [1,684.94, 1,913.54] | 1,908.87 [1,759.23, 2,058.51] |
| | | Other | 12,685 (0.3) | 160 (0.8) | 1,261.33 [1,073.46, 1,472.62] | 1,463.31 [1,135.16, 1,791.47] |
| | Class 4 (Small employers and own account workers) | Total | 4,352,905 (100.0) | 16,650 (100.0) | 382.50 [376.71, 388.36] | 704.90 [683.54, 726.25] |
| | | Heterosexual | 4,266,820 (98.0) | 15,900 (95.5) | 372.64 [366.87, 378.48] | 681.94 [660.68, 703.20] |
| | | LGB+ | 86,085 (2.0) | 750 (4.5) | 871.23 [809.99, 935.88] | 1,395.82 [1,209.98, 1,581.66] |
| | | Gay or Lesbian | 45,170 (1.0) | 345 (2.1) | 763.78 [685.30, 848.78] | 1,356.33 [1,006.04, 1,706.62] |
| | | Bisexual | 28,905 (0.7) | 320 (1.9) | 1,107.07 [989.09, 1,235.26] | 1,570.72 [1,299.71, 1,841.73] |
| | | Other | 12,010 (0.3) | 90 (0.5) | 749.38 [602.59, 921.11] | 1,023.74 [678.46, 1,427.70] |
| | Class 5 (Lower supervisory and technical occupations) | Total | 2,251,985 (100.0) | 10,770 (100.0) | 478.24 [469.25, 487.36] | 690.02 [672.51, 707.52] |
| | | Heterosexual | 2,197,490 (97.6) | 9,960 (92.5) | 453.24 [444.39, 462.23] | 646.53 [629.36, 663.71] |
| | | LGB+ | 54,500 (2.4) | 810 (7.5) | 1,486.24 [1,385.64, 1,592.21] | 1,882.53 [1,697.98, 2,067.09] |
| | | Gay or Lesbian | 29,855 (1.3) | 415 (3.9) | 1,390.05 [1,259.51, 1,530.44] | 1,859.77 [1,582.37, 2,137.17] |
| | | Bisexual | 19,370 (0.9) | 345 (3.2) | 1,781.10 [1,598.10, 1,979.32] | 2,028.61 [1,756.43, 2,300.80] |
| | | Other | 5,270 (0.2) | 55 (0.5) | 1,043.64 [786.21, 1,358.45] | 1,637.65 [939.61, 2,491.28] |
| | Class 6 (Semi-routine occupations) | Total | 4,745,535 (100.0) | 31,560 (100.0) | 665.05 [657.73, 672.42] | 1,018.98 [1,005.24, 1,032.71] |
| | | Heterosexual | 4,597,870 (96.9) | 28,070 (88.9) | 610.50 [603.38, 617.68] | 924.44 [910.98, 937.90] |
| | | LGB+ | 147,665 (3.1) | 3,490 (11.1) | 2,363.46 [2,285.69, 2,443.20] | 2,443.29 [2,355.50, 2,531.09] |
| | | Gay or Lesbian | 68,880 (1.5) | 1,420 (4.5) | 2,061.56 [1,955.71, 2,171.64] | 2,357.72 [2,222.40, 2,493.04] |
| | | Bisexual | 63,445 (1.3) | 1,755 (5.6) | 2,766.18 [2,638.26, 2,898.69] | 2,612.64 [2,479.31, 2,745.98] |
| | | Other | 15,340 (0.3) | 320 (1.0) | 2,086.05 [1,863.73, 2,327.59] | 2,144.27 [1,869.67, 2,418.88] |
| | Class 7 (Routine occupations) | Total | 4,963,690 (100.0) | 33,130 (100.0) | 667.45 [660.28, 674.67] | 1,033.29 [1,019.74, 1,046.85] |
| | | Heterosexual | 4,839,745 (97.5) | 30,345 (91.6) | 627.00 [619.96, 634.09] | 960.20 [946.84, 973.57] |
| | | LGB+ | 123,945 (2.5) | 2,785 (8.4) | 2,246.96 [2,164.28, 2,332.00] | 2,374.52 [2,281.44, 2,467.60] |
| | | Gay or Lesbian | 54,185 (1.1) | 1,125 (3.4) | 2,076.22 [1,956.65, 2,201.18] | 2,409.37 [2,258.00, 2,560.75] |
| | | Bisexual | 53,790 (1.1) | 1,415 (4.3) | 2,630.60 [2,495.31, 2,771.32] | 2,536.40 [2,396.85, 2,675.94] |
| | | Other | 15,970 (0.3) | 245 (0.7) | 1,534.13 [1,348.02, 1,738.74] | 1,696.25 [1,454.72, 1,937.78] |
| | Class 8 (Never worked and long-term unemployed) | Total | 3,273,875 (100.0) | 34,935 (100.0) | 1,067.08 [1,055.92, 1,078.33] | 1,724.24 [1,704.08, 1,744.40] |
| | | Heterosexual | 3,177,115 (97.0) | 31,260 (89.5) | 983.91 [973.03, 994.88] | 1,583.11 [1,563.26, 1,602.97] |
| | | LGB+ | 96,765 (3.0) | 3,680 (10.5) | 3,803.03 [3,681.14, 3,927.93] | 3,874.93 [3,748.66, 4,001.21] |
| | | Gay or Lesbian | 36,165 (1.1) | 1,415 (4.1) | 3,912.62 [3,711.39, 4,121.93] | 4,421.23 [4,187.47, 4,655.00] |
| | | Bisexual | 43,345 (1.3) | 1,920 (5.5) | 4,429.58 [4,233.64, 4,632.25] | 4,181.28 [3,988.85, 4,373.70] |
| | | Other | 17,255 (0.5) | 345 (1.0) | 1,999.42 [1,793.98, 2,221.94] | 2,164.54 [1,931.98, 2,397.11] |
| | Full-time students | Total | 2,808,105 (100.0) | 33,800 (100.0) | 1,203.66 [1,190.86, 1,216.56] | 657.76 [640.14, 675.39] |
| | | Heterosexual | 2,581,420 (91.9) | 27,125 (80.3) | 1,050.78 [1,038.31, 1,063.36] | 575.37 [557.94, 592.80] |
| | | LGB+ | 226,685 (8.1) | 6,670 (19.7) | 2,942.41 [2,872.21, 3,013.89] | 1,550.07 [1,427.11, 1,673.03] |
| | | Gay or Lesbian | 65,380 (2.3) | 1,980 (5.9) | 3,028.45 [2,896.51, 3,164.85] | 1,661.50 [1,442.87, 1,880.14] |
| | | Bisexual | 132,795 (4.7) | 3,955 (11.7) | 2,978.27 [2,886.17, 3,072.57] | 1,435.80 [1,317.13, 1,554.46] |
| | | Other | 28,510 (1.0) | 740 (2.2) | 2,595.58 [2,411.91, 2,789.52] | 1,450.94 [1,172.02, 1,729.86] |
| Household NS-SEC | Class 1 (Higher managerial, administrative and professional occupations) | Total | 10,270,425 (100.0) | 29,510 (100.0) | 287.33 [284.06, 290.63] | 422.07 [416.38, 427.75] |
| | | Heterosexual | 9,913,780 (96.5) | 26,265 (89.0) | 264.93 [261.74, 268.16] | 381.73 [376.18, 387.28] |
| | | LGB+ | 356,645 (3.5) | 3,250 (11.0) | 911.27 [880.21, 943.15] | 1,051.43 [1,014.38, 1,088.47] |
| | | Gay or Lesbian | 198,130 (1.9) | 1,280 (4.3) | 646.04 [611.13, 682.43] | 983.18 [921.99, 1,044.36] |
| | | Bisexual | 127,180 (1.2) | 1,615 (5.5) | 1,269.85 [1,208.67, 1,333.33] | 1,154.92 [1,097.00, 1,212.85] |

| | | | | | | |
|---|---|---|---|---|---|---|
| | | Other | 31,340 (0.3) | 350 (1.2) | 1,116.78 [1,002.84, 1,240.13] | 1,064.93 [950.84, 1,179.03] |
| | Class 2 (Lower managerial, administrative and professional occupations) | Total | 11,489,885 (100.0) | 47,165 (100.0) | 410.49 [406.80, 414.21] | 580.18 [574.20, 586.16] |
| | | Heterosexual | 11,078,640 (96.4) | 41,915 (88.9) | 378.34 [374.73, 381.98] | 528.27 [522.40, 534.14] |
| | | LGB+ | 411,245 (3.6) | 5,250 (11.1) | 1,276.61 [1,242.31, 1,311.62] | 1,382.29 [1,344.43, 1,420.14] |
| | | Gay or Lesbian | 223,345 (1.9) | 2,210 (4.7) | 989.50 [948.67, 1,031.63] | 1,300.85 [1,241.46, 1,360.23] |
| | | Bisexual | 150,600 (1.3) | 2,510 (5.3) | 1,666.67 [1,602.10, 1,733.17] | 1,499.28 [1,439.06, 1,559.49] |
| | | Other | 37,300 (0.3) | 530 (1.1) | 1,420.91 [1,302.50, 1,547.20] | 1,383.09 [1,263.22, 1,502.96] |
| | Class 3 (Intermediate occupations) | Total | 5,198,695 (100.0) | 27,125 (100.0) | 521.77 [515.57, 528.01] | 743.50 [733.55, 753.44] |
| | | Heterosexual | 5,057,880 (97.3) | 24,380 (89.9) | 482.02 [475.99, 488.11] | 680.59 [670.84, 690.33] |
| | | LGB+ | 140,810 (2.7) | 2,745 (10.1) | 1,949.44 [1,877.18, 2,023.76] | 1,890.62 [1,819.72, 1,961.52] |
| | | Gay or Lesbian | 68,410 (1.3) | 1,180 (4.4) | 1,724.89 [1,627.87, 1,826.19] | 1,904.18 [1,794.09, 2,014.26] |
| | | Bisexual | 57,795 (1.1) | 1,320 (4.9) | 2,283.93 [2,162.37, 2,410.55] | 1,964.72 [1,853.54, 2,075.91] |
| | | Other | 14,605 (0.3) | 245 (0.9) | 1,677.51 [1,474.01, 1,901.24] | 1,581.99 [1,383.08, 1,780.90] |
| | Class 4 (Small employers and own account workers) | Total | 3,976,970 (100.0) | 20,645 (100.0) | 519.11 [512.06, 526.24] | 747.65 [735.46, 759.83] |
| | | Heterosexual | 3,885,775 (97.7) | 19,255 (93.3) | 495.53 [488.55, 502.57] | 709.04 [696.94, 721.14] |
| | | LGB+ | 91,200 (2.3) | 1,390 (6.7) | 1,524.12 [1,445.04, 1,606.41] | 1,644.13 [1,554.86, 1,733.40] |
| | | Gay or Lesbian | 40,305 (1.0) | 530 (2.6) | 1,314.97 [1,205.39, 1,431.84] | 1,665.04 [1,513.53, 1,816.55] |
| | | Bisexual | 36,400 (0.9) | 700 (3.4) | 1,923.08 [1,783.24, 2,070.97] | 1,790.53 [1,653.56, 1,927.50] |
| | | Other | 14,495 (0.4) | 160 (0.8) | 1,103.83 [939.42, 1,288.74] | 1,225.77 [1,032.01, 1,419.53] |
| | Class 5 (Lower supervisory and technical occupations) | Total | 1,954,560 (100.0) | 12,115 (100.0) | 619.83 [608.84, 630.97] | 875.23 [857.33, 893.14] |
| | | Heterosexual | 1,900,420 (97.2) | 11,005 (90.8) | 579.08 [568.31, 590.00] | 810.02 [792.38, 827.66] |
| | | LGB+ | 54,140 (2.8) | 1,110 (9.2) | 2,050.24 [1,931.39, 2,174.49] | 2,072.17 [1,950.12, 2,194.21] |
| | | Gay or Lesbian | 25,075 (1.3) | 470 (3.9) | 1,874.38 [1,708.73, 2,051.75] | 2,138.75 [1,939.99, 2,337.51] |
| | | Bisexual | 22,850 (1.2) | 565 (4.7) | 2,472.65 [2,272.94, 2,685.20] | 2,260.14 [2,069.51, 2,450.76] |
| | | Other | 6,215 (0.3) | 75 (0.6) | 1,206.76 [949.19, 1,512.68] | 1,227.72 [961.38, 1,544.05] |
| | Class 6 (Semi-routine occupations) | Total | 3,340,505 (100.0) | 27,930 (100.0) | 836.10 [826.32, 845.97] | 1,213.51 [1,197.63, 1,229.39] |
| | | Heterosexual | 3,232,530 (96.8) | 24,760 (88.7) | 765.96 [756.45, 775.56] | 1,103.69 [1,088.10, 1,119.28] |
| | | LGB+ | 107,975 (3.2) | 3,170 (11.3) | 2,935.86 [2,834.54, 3,039.88] | 2,827.57 [2,728.57, 2,926.57] |
| | | Gay or Lesbian | 46,520 (1.4) | 1,280 (4.6) | 2,751.50 [2,602.82, 2,906.47] | 2,987.74 [2,823.04, 3,152.45] |
| | | Bisexual | 49,115 (1.5) | 1,590 (5.7) | 3,237.30 [3,080.11, 3,400.43] | 2,862.25 [2,716.36, 3,008.14] |
| | | Other | 12,340 (0.4) | 295 (1.1) | 2,390.60 [2,125.55, 2,679.56] | 2,313.65 [2,049.35, 2,577.94] |
| | Class 7 (Routine occupations) | Total | 2,439,520 (100.0) | 20,800 (100.0) | 852.63 [841.08, 864.29] | 1,251.04 [1,231.27, 1,270.81] |
| | | Heterosexual | 2,366,810 (97.0) | 18,780 (90.3) | 793.47 [782.16, 804.90] | 1,152.56 [1,133.06, 1,172.07] |
| | | LGB+ | 72,705 (3.0) | 2,020 (9.7) | 2,778.35 [2,658.50, 2,902.21] | 2,803.54 [2,680.57, 2,926.51] |
| | | Gay or Lesbian | 28,295 (1.2) | 745 (3.6) | 2,632.97 [2,447.28, 2,829.03] | 2,981.00 [2,761.99, 3,200.00] |
| | | Bisexual | 33,815 (1.4) | 1,095 (5.3) | 3,238.21 [3,049.23, 3,435.84] | 2,993.75 [2,812.63, 3,174.88] |
| | | Other | 10,595 (0.4) | 180 (0.9) | 1,698.91 [1,459.78, 1,966.05] | 1,777.05 [1,516.98, 2,037.13] |
| | Class 8 (Never worked and long-term unemployed) | Total | 1,655,055 (100.0) | 20,330 (100.0) | 1,228.36 [1,211.53, 1,245.36] | 2,273.94 [2,237.49, 2,310.40] |
| | | Heterosexual | 1,605,800 (97.0) | 18,230 (89.7) | 1,135.26 [1,118.84, 1,151.86] | 2,091.21 [2,055.20, 2,127.21] |
| | | LGB+ | 49,260 (3.0) | 2,100 (10.3) | 4,263.09 [4,082.69, 4,449.41] | 4,750.79 [4,545.57, 4,956.02] |
| | | Gay or Lesbian | 19,460 (1.2) | 825 (4.1) | 4,239.47 [3,955.07, 4,538.90] | 5,613.44 [5,210.95, 6,015.92] |
| | | Bisexual | 21,365 (1.3) | 1,085 (5.3) | 5,078.40 [4,780.68, 5,389.80] | 5,049.03 [4,747.41, 5,350.66] |
| | | Other | 8,435 (0.5) | 185 (0.9) | 2,193.24 [1,888.57, 2,533.07] | 2,508.01 [2,137.13, 2,878.89] |
| | Full-time students | Total | 386,235 (100.0) | 3,700 (100.0) | 957.97 [927.34, 989.34] | 845.28 [806.32, 884.24] |
| | | Heterosexual | 329,545 (85.3) | 2,485 (67.2) | 754.07 [724.71, 784.31] | 714.87 [676.48, 753.26] |

| | | | | | | |
|---|---|---|---|---|---|---|
| | | LGB+ | 56,690 (14.7) | 1,215 (32.8) | 2,143.24 [2,024.40, 2,267.22] | 1,812.23 [1,575.94, 2,048.52] |
| | | Gay or Lesbian | 14,205 (3.7) | 315 (8.5) | 2,217.53 [1,979.38, 2,476.44] | 2,051.21 [1,611.47, 2,490.96] |
| | | Bisexual | 35,465 (9.2) | 760 (20.5) | 2,142.96 [1,993.29, 2,300.88] | 1,626.48 [1,398.29, 1,854.67] |
| | | Other | 7,020 (1.8) | 140 (3.8) | 1,994.30 [1,677.64, 2,353.35] | 1,698.33 [1,070.69, 2,325.96] |
| | Living in a communal establishment | Total | 486,475 (100.0) | 6,860 (100.0) | 1,410.14 [1,376.97, 1,443.92] | 2,627.65 [2,551.66, 2,703.64] |
| | | Heterosexual | 446,340 (91.7) | 5,425 (79.1) | 1,215.44 [1,183.31, 1,248.22] | 2,414.70 [2,338.27, 2,491.12] |
| | | LGB+ | 40,135 (8.3) | 1,435 (20.9) | 3,575.43 [3,392.81, 3,765.33] | 4,733.46 [4,339.05, 5,127.88] |
| | | Gay or Lesbian | 10,810 (2.2) | 380 (5.5) | 3,515.26 [3,170.67, 3,887.10] | 4,297.19 [3,732.49, 4,861.88] |
| | | Bisexual | 24,675 (5.1) | 920 (13.4) | 3,728.47 [3,491.40, 3,977.40] | 5,770.71 [5,089.57, 6,451.86] |
| | | Other | 4,650 (1.0) | 135 (2.0) | 2,903.23 [2,434.17, 3,436.32] | 2,540.42 [1,807.95, 3,272.90] |
| Lone person household indicator | Lives alone | Total | 5,954,715 (100.0) | 45,170 (100.0) | 758.56 [751.58, 765.59] | 2,147.80 [2,100.37, 2,195.22] |
| | | Heterosexual | 5,697,830 (95.7) | 39,795 (88.1) | 698.42 [691.58, 705.32] | 1,873.22 [1,826.05, 1,920.40] |
| | | LGB+ | 256,880 (4.3) | 5,370 (11.9) | 2,090.47 [2,034.93, 2,147.14] | 3,830.03 [3,663.78, 3,996.28] |
| | | Gay or Lesbian | 147,065 (2.5) | 2,405 (5.3) | 1,635.33 [1,570.62, 1,702.02] | 3,934.17 [3,614.02, 4,254.31] |
| | | Bisexual | 85,070 (1.4) | 2,445 (5.4) | 2,874.10 [2,761.30, 2,990.33] | 4,106.43 [3,884.59, 4,328.27] |
| | | Other | 24,750 (0.4) | 520 (1.2) | 2,101.01 [1,924.28, 2,289.60] | 3,162.76 [2,726.80, 3,598.72] |
| | Does not live alone | Total | 34,757,135 (100.0) | 164,150 (100.0) | 472.28 [470.00, 474.57] | 649.45 [645.98, 652.92] |
| | | Heterosexual | 33,673,350 (96.9) | 147,280 (89.7) | 437.38 [435.15, 439.62] | 597.93 [594.51, 601.35] |
| | | LGB+ | 1,083,785 (3.1) | 16,875 (10.3) | 1,557.04 [1,533.64, 1,580.72] | 1,471.98 [1,449.70, 1,494.26] |
| | | Gay or Lesbian | 516,680 (1.5) | 6,435 (3.9) | 1,245.45 [1,215.21, 1,276.26] | 1,434.37 [1,398.61, 1,470.13] |
| | | Bisexual | 449,515 (1.3) | 8,800 (5.4) | 1,957.67 [1,916.97, 1,999.00] | 1,601.74 [1,565.96, 1,637.51] |
| | | Other | 117,590 (0.3) | 1,640 (1.0) | 1,394.68 [1,327.99, 1,463.85] | 1,250.40 [1,188.99, 1,311.80] |
| | Living in a communal establishment | Total | 486,475 (100.0) | 6,860 (100.0) | 1,410.14 [1,376.97, 1,443.92] | 2,627.65 [2,551.66, 2,703.64] |
| | | Heterosexual | 446,340 (91.7) | 5,425 (79.1) | 1,215.44 [1,183.31, 1,248.22] | 2,414.70 [2,338.27, 2,491.12] |
| | | LGB+ | 40,135 (8.3) | 1,435 (20.9) | 3,575.43 [3,392.81, 3,765.33] | 4,733.46 [4,339.05, 5,127.88] |
| | | Gay or Lesbian | 10,810 (2.2) | 380 (5.5) | 3,515.26 [3,170.67, 3,887.10] | 4,297.19 [3,732.49, 4,861.88] |
| | | Bisexual | 24,675 (5.1) | 920 (13.4) | 3,728.47 [3,491.40, 3,977.40] | 5,770.71 [5,089.57, 6,451.86] |
| | | Other | 4,650 (1.0) | 135 (2.0) | 2,903.23 [2,434.17, 3,436.32] | 2,540.42 [1,807.95, 3,272.90] |
| Household size | One person in household | Total | 5,954,715 (100.0) | 45,170 (100.0) | 758.56 [751.58, 765.59] | 2,147.80 [2,100.37, 2,195.22] |
| | | Heterosexual | 5,697,830 (95.7) | 39,795 (88.1) | 698.42 [691.58, 705.32] | 1,873.22 [1,826.05, 1,920.40] |
| | | LGB+ | 256,880 (4.3) | 5,370 (11.9) | 2,090.47 [2,034.93, 2,147.14] | 3,830.03 [3,663.78, 3,996.28] |
| | | Gay or Lesbian | 147,065 (2.5) | 2,405 (5.3) | 1,635.33 [1,570.62, 1,702.02] | 3,934.17 [3,614.02, 4,254.31] |
| | | Bisexual | 85,070 (1.4) | 2,445 (5.4) | 2,874.10 [2,761.30, 2,990.33] | 4,106.43 [3,884.59, 4,328.27] |
| | | Other | 24,750 (0.4) | 520 (1.2) | 2,101.01 [1,924.28, 2,289.60] | 3,162.76 [2,726.80, 3,598.72] |
| | Two people in household | Total | 14,026,325 (100.0) | 54,485 (100.0) | 388.45 [385.19, 391.72] | 808.55 [798.78, 818.31] |
| | | Heterosexual | 13,512,625 (96.3) | 48,495 (89.0) | 358.89 [355.70, 362.10] | 730.88 [721.21, 740.56] |
| | | LGB+ | 513,705 (3.7) | 5,990 (11.0) | 1,166.04 [1,136.69, 1,195.95] | 1,572.14 [1,525.38, 1,618.89] |
| | | Gay or Lesbian | 305,270 (2.2) | 2,620 (4.8) | 858.26 [825.70, 891.76] | 1,525.41 [1,443.93, 1,606.90] |
| | | Bisexual | 167,505 (1.2) | 2,815 (5.2) | 1,680.55 [1,619.03, 1,743.80] | 1,740.71 [1,670.10, 1,811.31] |
| | | Other | 40,925 (0.3) | 555 (1.0) | 1,356.14 [1,245.65, 1,473.81] | 1,564.94 [1,420.42, 1,709.47] |
| | Three to five people in household | Total | 18,767,340 (100.0) | 97,180 (100.0) | 517.81 [514.56, 521.08] | 623.65 [619.46, 627.85] |
| | | Heterosexual | 18,251,775 (97.3) | 87,460 (90.0) | 479.19 [476.02, 482.37] | 579.07 [574.92, 583.21] |
| | | LGB+ | 515,565 (2.7) | 9,725 (10.0) | 1,886.28 [1,848.97, 1,924.15] | 1,483.08 [1,450.27, 1,515.90] |
| | | Gay or Lesbian | 196,230 (1.0) | 3,450 (3.6) | 1,758.14 [1,699.96, 1,817.81] | 1,491.33 [1,438.00, 1,544.65] |
| | | Bisexual | 252,715 (1.3) | 5,320 (5.5) | 2,105.14 [2,048.95, 2,162.48] | 1,573.10 [1,522.43, 1,623.78] |

| | | | | | | |
|---|---|---|---|---|---|---|
| | | Other | 66,625 (0.4) | 955 (1.0) | 1,433.40 [1,343.92, 1,527.27] | 1,144.82 [1,068.16, 1,221.47] |
| | Six or more people in household | Total | 1,963,470 (100.0) | 12,485 (100.0) | 635.86 [624.76, 647.12] | 627.25 [615.62, 638.87] |
| | | Heterosexual | 1,908,955 (97.2) | 11,325 (90.7) | 593.26 [582.38, 604.29] | 590.74 [579.23, 602.26] |
| | | LGB+ | 54,515 (2.8) | 1,160 (9.3) | 2,127.85 [2,007.15, 2,253.92] | 1,486.91 [1,387.57, 1,586.25] |
| | | Gay or Lesbian | 15,185 (0.8) | 365 (2.9) | 2,403.69 [2,163.39, 2,663.38] | 1,696.03 [1,481.69, 1,910.36] |
| | | Bisexual | 29,295 (1.5) | 665 (5.3) | 2,270.01 [2,100.74, 2,449.30] | 1,559.03 [1,417.52, 1,700.54] |
| | | Other | 10,040 (0.5) | 130 (1.0) | 1,294.82 [1,081.82, 1,537.49] | 1,055.31 [864.38, 1,246.24] |
| | Living in a communal establishment | Total | 486,475 (100.0) | 6,860 (100.0) | 1,410.14 [1,376.97, 1,443.92] | 2,627.65 [2,551.66, 2,703.64] |
| | | Heterosexual | 446,340 (91.7) | 5,425 (79.1) | 1,215.44 [1,183.31, 1,248.22] | 2,414.70 [2,338.27, 2,491.12] |
| | | LGB+ | 40,135 (8.3) | 1,435 (20.9) | 3,575.43 [3,392.81, 3,765.33] | 4,733.46 [4,339.05, 5,127.88] |
| | | Gay or Lesbian | 10,810 (2.2) | 380 (5.5) | 3,515.26 [3,170.67, 3,887.10] | 4,297.19 [3,732.49, 4,861.88] |
| | | Bisexual | 24,675 (5.1) | 920 (13.4) | 3,728.47 [3,491.40, 3,977.40] | 5,770.71 [5,089.57, 6,451.86] |
| | | Other | 4,650 (1.0) | 135 (2.0) | 2,903.23 [2,434.17, 3,436.32] | 2,540.42 [1,807.95, 3,272.90] |
| Household tenure | Owned or shared ownership | Total | 27,298,305 (100.0) | 89,320 (100.0) | 327.20 [325.06, 329.35] | 498.08 [494.01, 502.15] |
| | | Heterosexual | 26,653,165 (97.6) | 82,485 (92.3) | 309.48 [307.37, 311.59] | 463.21 [459.21, 467.22] |
| | | LGB+ | 645,140 (2.4) | 6,835 (7.7) | 1,059.46 [1,034.49, 1,084.88] | 1,179.33 [1,150.63, 1,208.03] |
| | | Gay or Lesbian | 373,305 (1.4) | 2,920 (3.3) | 782.20 [754.09, 811.10] | 1,134.24 [1,089.33, 1,179.14] |
| | | Bisexual | 215,560 (0.8) | 3,215 (3.6) | 1,491.46 [1,440.35, 1,543.93] | 1,262.59 [1,217.26, 1,307.93] |
| | | Other | 56,270 (0.2) | 695 (0.8) | 1,235.12 [1,144.99, 1,330.46] | 1,161.61 [1,073.69, 1,249.53] |
| | Social rented | Total | 6,069,620 (100.0) | 69,220 (100.0) | 1,140.43 [1,131.95, 1,148.96] | 1,418.33 [1,406.79, 1,429.88] |
| | | Heterosexual | 5,853,620 (96.4) | 61,825 (89.3) | 1,056.18 [1,047.87, 1,064.54] | 1,305.27 [1,293.93, 1,316.62] |
| | | LGB+ | 216,000 (3.6) | 7,395 (10.7) | 3,423.61 [3,346.02, 3,502.55] | 3,369.72 [3,292.08, 3,447.36] |
| | | Gay or Lesbian | 96,265 (1.6) | 2,990 (4.3) | 3,106.01 [2,995.67, 3,219.38] | 3,348.89 [3,226.68, 3,471.10] |
| | | Bisexual | 95,410 (1.6) | 3,780 (5.5) | 3,961.85 [3,836.55, 4,090.20] | 3,633.52 [3,514.12, 3,752.92] |
| | | Other | 24,330 (0.4) | 625 (0.9) | 2,568.85 [2,371.37, 2,778.37] | 2,554.33 [2,351.95, 2,756.71] |
| | Private rented or living rent free | Total | 7,343,925 (100.0) | 50,780 (100.0) | 691.46 [685.45, 697.50] | 793.32 [786.01, 800.63] |
| | | Heterosexual | 6,864,400 (93.5) | 42,765 (84.2) | 623.00 [617.11, 628.93] | 709.47 [702.29, 716.65] |
| | | LGB+ | 479,525 (6.5) | 8,020 (15.8) | 1,672.49 [1,636.08, 1,709.50] | 1,681.12 [1,641.02, 1,721.23] |
| | | Gay or Lesbian | 194,170 (2.6) | 2,925 (5.8) | 1,506.41 [1,452.31, 1,562.01] | 1,722.96 [1,653.40, 1,792.52] |
| | | Bisexual | 223,610 (3.0) | 4,250 (8.4) | 1,900.63 [1,843.91, 1,958.65] | 1,745.55 [1,685.58, 1,805.52] |
| | | Other | 61,740 (0.8) | 840 (1.7) | 1,360.54 [1,270.08, 1,455.75] | 1,340.99 [1,241.30, 1,440.68] |
| | Living in a communal establishment | Total | 486,475 (100.0) | 6,860 (100.0) | 1,410.14 [1,376.97, 1,443.92] | 2,627.65 [2,551.66, 2,703.64] |
| | | Heterosexual | 446,340 (91.7) | 5,425 (79.1) | 1,215.44 [1,183.31, 1,248.22] | 2,414.70 [2,338.27, 2,491.12] |
| | | LGB+ | 40,135 (8.3) | 1,435 (20.9) | 3,575.43 [3,392.81, 3,765.33] | 4,733.46 [4,339.05, 5,127.88] |
| | | Gay or Lesbian | 10,810 (2.2) | 380 (5.5) | 3,515.26 [3,170.67, 3,887.10] | 4,297.19 [3,732.49, 4,861.88] |
| | | Bisexual | 24,675 (5.1) | 920 (13.4) | 3,728.47 [3,491.40, 3,977.40] | 5,770.71 [5,089.57, 6,451.86] |
| | | Other | 4,650 (1.0) | 135 (2.0) | 2,903.23 [2,434.17, 3,436.32] | 2,540.42 [1,807.95, 3,272.90] |
| Economic activity | Economically active | Total | 25,443,735 (100.0) | 104,760 (100.0) | 411.73 [409.24, 414.23] | 594.12 [589.91, 598.33] |
| | | Heterosexual | 24,463,115 (96.1) | 93,355 (89.1) | 381.62 [379.17, 384.07] | 545.16 [541.02, 549.31] |
| | | LGB+ | 980,620 (3.9) | 11,405 (10.9) | 1,163.04 [1,141.79, 1,184.58] | 1,323.61 [1,296.80, 1,350.42] |
| | | Gay or Lesbian | 510,450 (2.0) | 4,760 (4.5) | 932.51 [906.21, 959.39] | 1,268.43 [1,225.82, 1,311.04] |
| | | Bisexual | 375,785 (1.5) | 5,590 (5.3) | 1,487.55 [1,448.81, 1,527.07] | 1,404.33 [1,363.63, 1,445.03] |
| | | Other | 94,385 (0.4) | 1,050 (1.0) | 1,112.46 [1,046.19, 1,181.84] | 1,241.23 [1,158.91, 1,323.55] |
| | Economically inactive | Total | 15,754,590 (100.0) | 111,425 (100.0) | 707.25 [703.11, 711.42] | 1,411.96 [1,401.34, 1,422.58] |
| | | Heterosexual | 15,354,410 (97.5) | 99,145 (89.0) | 645.71 [641.70, 649.74] | 1,282.78 [1,272.33, 1,293.24] |

| | | | | | | |
|---|---|---|---|---|---|---|
| | | LGB+ | 400,180 (2.5) | 12,280 (11.0) | 3,068.62 [3,014.58, 3,123.38] | 3,247.43 [3,182.72, 3,312.15] |
| | | Gay or Lesbian | 164,110 (1.0) | 4,460 (4.0) | 2,717.69 [2,638.51, 2,798.64] | 3,627.27 [3,507.92, 3,746.63] |
| | | Bisexual | 183,470 (1.2) | 6,575 (5.9) | 3,583.69 [3,497.59, 3,671.38] | 3,402.60 [3,305.32, 3,499.87] |
| | | Other | 52,605 (0.3) | 1,245 (1.1) | 2,366.70 [2,237.04, 2,501.90] | 2,210.98 [2,077.26, 2,344.69] |
| General health (detailed) | Very good health | Total | 16,934,985 (100.0) | 57,960 (100.0) | 342.25 [339.47, 345.05] | 404.17 [400.75, 407.60] |
| | | Heterosexual | 16,399,805 (96.8) | 53,140 (91.7) | 324.03 [321.28, 326.80] | 384.33 [380.91, 387.74] |
| | | LGB+ | 535,180 (3.2) | 4,820 (8.3) | 900.63 [875.38, 926.42] | 833.10 [809.38, 856.83] |
| | | Gay or Lesbian | 292,080 (1.7) | 2,210 (3.8) | 756.64 [725.42, 788.86] | 865.94 [829.00, 902.87] |
| | | Bisexual | 194,705 (1.1) | 2,285 (3.9) | 1,173.57 [1,125.94, 1,222.70] | 872.50 [832.92, 912.08] |
| | | Other | 48,395 (0.3) | 330 (0.6) | 681.89 [610.29, 759.57] | 611.85 [544.04, 679.67] |
| | Good health | Total | 15,579,320 (100.0) | 71,955 (100.0) | 461.86 [458.49, 465.25] | 823.83 [816.79, 830.86] |
| | | Heterosexual | 15,036,325 (96.5) | 63,660 (88.5) | 423.37 [420.09, 426.68] | 755.62 [748.58, 762.66] |
| | | LGB+ | 542,995 (3.5) | 8,295 (11.5) | 1,527.64 [1,494.94, 1,560.87] | 1,522.11 [1,489.33, 1,554.89] |
| | | Gay or Lesbian | 254,750 (1.6) | 3,180 (4.4) | 1,248.28 [1,205.27, 1,292.44] | 1,621.88 [1,562.98, 1,680.78] |
| | | Bisexual | 230,325 (1.5) | 4,390 (6.1) | 1,906.00 [1,850.03, 1,963.23] | 1,565.05 [1,516.70, 1,613.40] |
| | | Other | 57,920 (0.4) | 725 (1.0) | 1,251.73 [1,162.26, 1,346.25] | 1,143.65 [1,059.71, 1,227.58] |
| | Fair health | Total | 6,156,315 (100.0) | 51,020 (100.0) | 828.74 [821.57, 835.97] | 2,082.18 [2,058.22, 2,106.13] |
| | | Heterosexual | 5,946,370 (96.6) | 44,550 (87.3) | 749.20 [742.26, 756.19] | 1,881.63 [1,857.06, 1,906.19] |
| | | LGB+ | 209,945 (3.4) | 6,470 (12.7) | 3,081.76 [3,007.12, 3,157.78] | 3,360.71 [3,278.31, 3,443.11] |
| | | Gay or Lesbian | 88,170 (1.4) | 2,310 (4.5) | 2,619.94 [2,514.18, 2,729.01] | 3,764.69 [3,601.61, 3,927.78] |
| | | Bisexual | 95,245 (1.5) | 3,465 (6.8) | 3,637.99 [3,517.85, 3,761.18] | 3,408.88 [3,293.61, 3,524.14] |
| | | Other | 26,535 (0.4) | 700 (1.4) | 2,638.03 [2,446.19, 2,840.90] | 2,561.00 [2,370.18, 2,751.83] |
| | Bad health | Total | 1,963,555 (100.0) | 26,500 (100.0) | 1,349.59 [1,333.39, 1,365.94] | 3,479.32 [3,415.79, 3,542.84] |
| | | Heterosexual | 1,890,340 (96.3) | 23,395 (88.3) | 1,237.61 [1,221.80, 1,253.57] | 3,149.94 [3,084.05, 3,215.83] |
| | | LGB+ | 73,215 (3.7) | 3,105 (11.7) | 4,240.93 [4,093.06, 4,392.78] | 5,212.25 [5,019.18, 5,405.31] |
| | | Gay or Lesbian | 30,720 (1.6) | 1,135 (4.3) | 3,694.66 [3,482.82, 3,916.02] | 5,681.32 [5,301.32, 6,061.32] |
| | | Bisexual | 31,695 (1.6) | 1,575 (5.9) | 4,969.24 [4,726.83, 5,220.86] | 5,404.12 [5,131.13, 5,677.10] |
| | | Other | 10,805 (0.6) | 400 (1.5) | 3,701.99 [3,348.05, 4,083.17] | 4,042.06 [3,634.78, 4,449.34] |
| | Very bad health | Total | 564,145 (100.0) | 8,745 (100.0) | 1,550.13 [1,517.81, 1,582.97] | 4,109.05 [3,969.02, 4,249.08] |
| | | Heterosexual | 544,685 (96.6) | 7,755 (88.7) | 1,423.76 [1,392.25, 1,455.81] | 3,657.56 [3,517.09, 3,798.03] |
| | | LGB+ | 19,460 (3.4) | 990 (11.3) | 5,087.36 [4,775.35, 5,414.40] | 7,147.12 [6,632.54, 7,661.70] |
| | | Gay or Lesbian | 8,840 (1.6) | 395 (4.5) | 4,468.33 [4,038.49, 4,931.46] | 7,620.56 [6,648.19, 8,592.92] |
| | | Bisexual | 7,290 (1.3) | 455 (5.2) | 6,241.43 [5,681.04, 6,842.14] | 7,640.76 [6,872.73, 8,408.79] |
| | | Other | 3,330 (0.6) | 145 (1.7) | 4,354.35 [3,674.47, 5,123.57] | 5,469.66 [4,487.21, 6,452.10] |
| General health (aggregated) | Good health | Total | 32,514,310 (100.0) | 129,920 (100.0) | 399.58 [397.41, 401.76] | 548.97 [545.74, 552.21] |
| | | Heterosexual | 31,436,130 (96.7) | 116,800 (89.9) | 371.55 [369.42, 373.68] | 510.46 [507.26, 513.67] |
| | | LGB+ | 1,078,175 (3.3) | 13,115 (10.1) | 1,216.41 [1,195.68, 1,237.41] | 1,169.55 [1,149.50, 1,189.60] |
| | | Gay or Lesbian | 546,830 (1.7) | 5,385 (4.1) | 984.77 [958.64, 1,011.43] | 1,189.30 [1,156.59, 1,222.00] |
| | | Bisexual | 425,030 (1.3) | 6,675 (5.1) | 1,570.48 [1,533.03, 1,608.61] | 1,239.47 [1,207.80, 1,271.14] |
| | | Other | 106,315 (0.3) | 1,055 (0.8) | 992.33 [933.35, 1,054.07] | 896.06 [841.30, 950.82] |
| | Not good health | Total | 8,684,015 (100.0) | 86,265 (100.0) | 993.38 [986.76, 1,000.03] | 2,442.63 [2,420.03, 2,465.24] |
| | | Heterosexual | 8,381,395 (96.5) | 75,695 (87.7) | 903.13 [896.71, 909.59] | 2,202.59 [2,179.42, 2,225.75] |
| | | LGB+ | 302,620 (3.5) | 10,565 (12.2) | 3,491.18 [3,424.92, 3,558.39] | 3,960.92 [3,884.25, 4,037.58] |
| | | Gay or Lesbian | 127,725 (1.5) | 3,835 (4.4) | 3,002.54 [2,908.26, 3,099.11] | 4,374.40 [4,223.35, 4,525.45] |
| | | Bisexual | 134,225 (1.5) | 5,490 (6.4) | 4,090.15 [3,982.66, 4,199.80] | 4,043.22 [3,935.64, 4,150.81] |

| | | | | | | |
|---|---|---|---|---|---|---|
| | | Other | 40,670 (0.5) | 1,240 (1.4) | 3,048.93 [2,881.57, 3,223.48] | 3,122.38 [2,947.87, 3,296.90] |
| Disability | Day-to-day activities limited a lot | Total | 3,441,160 (100.0) | 50,100 (100.0) | 1,455.90 [1,443.18, 1,468.71] | 3,271.86 [3,235.29, 3,308.43] |
| | | Heterosexual | 3,307,285 (96.1) | 43,235 (86.3) | 1,307.27 [1,294.97, 1,319.65] | 2,922.07 [2,885.10, 2,959.04] |
| | | LGB+ | 133,875 (3.9) | 6,865 (13.7) | 5,127.92 [5,007.32, 5,250.68] | 5,554.02 [5,421.44, 5,686.61] |
| | | Gay or Lesbian | 55,805 (1.6) | 2,500 (5.0) | 4,479.89 [4,305.98, 4,659.01] | 5,964.87 [5,716.93, 6,212.81] |
| | | Bisexual | 58,395 (1.7) | 3,520 (7.0) | 6,027.91 [5,830.41, 6,230.40] | 5,788.79 [5,596.05, 5,981.53] |
| | | Other | 19,675 (0.6) | 845 (1.7) | 4,294.79 [4,010.06, 4,594.40] | 4,249.21 [3,962.36, 4,536.06] |
| | Day-to-day activities limited a little | Total | 4,765,825 (100.0) | 50,805 (100.0) | 1,066.03 [1,056.78, 1,075.34] | 2,028.54 [2,008.29, 2,048.78] |
| | | Heterosexual | 4,491,165 (94.2) | 42,685 (84.0) | 950.42 [941.43, 959.48] | 1,889.17 [1,867.66, 1,910.68] |
| | | LGB+ | 274,660 (5.8) | 8,120 (16.0) | 2,956.38 [2,892.43, 3,021.40] | 2,773.96 [2,712.63, 2,835.28] |
| | | Gay or Lesbian | 100,800 (2.1) | 2,745 (5.4) | 2,723.21 [2,622.28, 2,827.03] | 3,174.13 [3,053.88, 3,294.38] |
| | | Bisexual | 138,475 (2.9) | 4,555 (9.0) | 3,289.40 [3,194.56, 3,386.34] | 2,835.00 [2,745.57, 2,924.43] |
| | | Other | 35,385 (0.7) | 820 (1.6) | 2,317.37 [2,161.45, 2,481.56] | 2,001.60 [1,858.88, 2,144.32] |
| | Day-to-day activities not limited | Total | 32,991,340 (100.0) | 115,280 (100.0) | 349.43 [347.41, 351.45] | 478.54 [475.47, 481.61] |
| | | Heterosexual | 32,019,080 (97.1) | 106,580 (92.5) | 332.86 [330.87, 334.87] | 455.63 [452.57, 458.70] |
| | | LGB+ | 972,260 (2.9) | 8,700 (7.5) | 894.82 [876.12, 913.83] | 909.59 [890.45, 928.73] |
| | | Gay or Lesbian | 517,945 (1.6) | 3,980 (3.5) | 768.42 [744.73, 792.67] | 959.80 [928.62, 990.98] |
| | | Bisexual | 362,385 (1.1) | 4,095 (3.6) | 1,130.01 [1,095.67, 1,165.16] | 929.16 [899.49, 958.83] |
| | | Other | 91,930 (0.3) | 630 (0.5) | 685.30 [632.83, 740.97] | 666.16 [613.69, 718.62] |
| Residence type | Living in a private household | Total | 40,711,850 (100.0) | 209,320 (100.0) | 514.15 [511.95, 516.36] | 723.93 [720.39, 727.47] |
| | | Heterosexual | 39,371,185 (96.7) | 187,075 (89.4) | 475.16 [473.01, 477.32] | 660.66 [657.18, 664.13] |
| | | LGB+ | 1,340,665 (3.3) | 22,245 (10.6) | 1,659.25 [1,637.52, 1,681.20] | 1,701.97 [1,679.58, 1,724.36] |
| | | Gay or Lesbian | 663,745 (1.6) | 8,840 (4.2) | 1,331.84 [1,304.22, 1,359.89] | 1,649.85 [1,613.78, 1,685.92] |
| | | Bisexual | 534,585 (1.3) | 11,245 (5.4) | 2,103.50 [2,064.80, 2,142.75] | 1,866.65 [1,830.95, 1,902.36] |
| | | Other | 142,340 (0.3) | 2,160 (1.0) | 1,517.49 [1,454.17, 1,582.87] | 1,472.69 [1,410.21, 1,535.17] |
| | Living in a communal establishment | Total | 486,475 (100.0) | 6,860 (100.0) | 1,410.14 [1,376.97, 1,443.92] | 2,627.65 [2,551.66, 2,703.64] |
| | | Heterosexual | 446,340 (91.7) | 5,425 (79.1) | 1,215.44 [1,183.31, 1,248.22] | 2,414.70 [2,338.27, 2,491.12] |
| | | LGB+ | 40,135 (8.3) | 1,435 (20.9) | 3,575.43 [3,392.81, 3,765.33] | 4,733.46 [4,339.05, 5,127.88] |
| | | Gay or Lesbian | 10,810 (2.2) | 380 (5.5) | 3,515.26 [3,170.67, 3,887.10] | 4,297.19 [3,732.49, 4,861.88] |
| | | Bisexual | 24,675 (5.1) | 920 (13.4) | 3,728.47 [3,491.40, 3,977.40] | 5,770.71 [5,089.57, 6,451.86] |
| | | Other | 4,650 (1.0) | 135 (2.0) | 2,903.23 [2,434.17, 3,436.32] | 2,540.42 [1,807.95, 3,272.90] |

Notes: Age-standardised rates are expressed per 100,000 people and standardised to the age distribution of the observed study population identifying with an LGB+ orientation in Census 2021. Cells with the value '-' are not applicable for the given characteristic or category. All calculations are based on estimates rounded to the nearest multiple of five for disclosure control reasons. CI: confidence interval.

**Supplementary Table 12: Sensitivity analysis including proxy responses to the sexual orientation question in Census 2021, crude and age-standardised rates of suicide per 100,000 people**

| Characteristic | Category | Sexual orientation | Count (%), Individuals | Count (%), Individuals with death by suicide | Crude rate of suicide [95% CI] | Age-standardised rate of suicide [95% CI] |
|---|---|---|---|---|---|---|
| Total | Total | Total | 41,198,325 (100.0) | 10,910 (100.0) | 26.48 [25.99, 26.98] | 26.09 [25.50, 26.69] |
| | | Heterosexual | 39,817,525 (96.6) | 10,195 (93.4) | 25.60 [25.11, 26.11] | 24.87 [24.28, 25.47] |
| | | LGB+ | 1,380,800 (3.4) | 715 (6.6) | 51.78 [48.06, 55.72] | 51.76 [47.97, 55.56] |
| | | Gay or Lesbian | 674,555 (1.6) | 360 (3.3) | 53.37 [48.00, 59.18] | 50.46 [45.03, 55.89] |

| | | | | | | |
|---|---|---|---|---|---|---|
| | | Bisexual | 559,255 (1.4) | 280 (2.6) | 50.07 [44.37, 56.29] | 54.14 [47.43, 60.86] |
| | | Other | 146,990 (0.4) | 75 (0.7) | 51.02 [40.13, 63.96] | 52.71 [41.33, 66.22] |
| Age on Census Day (10-year bands) | 16 to 24 years | Total | 5,105,765 (100.0) | 1,090 (100.0) | 21.35 [20.08, 22.62] | - |
| | | Heterosexual | 4,719,460 (92.4) | 935 (85.8) | 19.81 [18.54, 21.08] | - |
| | | LGB+ | 386,305 (7.6) | 155 (14.2) | 40.12 [33.81, 46.44] | - |
| | 25 to 34 years | Total | 6,807,450 (100.0) | 1,740 (100.0) | 25.56 [24.36, 26.76] | - |
| | | Heterosexual | 6,398,210 (94.0) | 1,545 (88.8) | 24.15 [22.94, 25.35] | - |
| | | LGB+ | 409,240 (6.0) | 190 (10.9) | 46.43 [39.83, 53.03] | - |
| | 35 to 44 years | Total | 6,608,235 (100.0) | 1,885 (100.0) | 28.53 [27.24, 29.81] | - |
| | | Heterosexual | 6,365,385 (96.3) | 1,745 (92.6) | 27.41 [26.13, 28.70] | - |
| | | LGB+ | 242,850 (3.7) | 140 (7.4) | 57.65 [48.10, 67.20] | - |
| | 45 to 54 years | Total | 6,805,755 (100.0) | 2,315 (100.0) | 34.02 [32.63, 35.40] | - |
| | | Heterosexual | 6,635,310 (97.5) | 2,200 (95.0) | 33.16 [31.77, 34.54] | - |
| | | LGB+ | 170,445 (2.5) | 120 (5.2) | 70.40 [57.81, 83.00] | - |
| | 55 to 64 years | Total | 6,480,905 (100.0) | 1,890 (100.0) | 29.16 [27.85, 30.48] | - |
| | | Heterosexual | 6,372,060 (98.3) | 1,820 (96.3) | 28.56 [27.25, 29.87] | - |
| | | LGB+ | 108,845 (1.7) | 70 (3.7) | 64.31 [50.13, 81.25] | - |
| | 65 years and over | Total | 9,390,215 (100.0) | 1,985 (100.0) | 21.14 [20.21, 22.07] | - |
| | | Heterosexual | 9,327,105 (99.3) | 1,950 (98.2) | 20.91 [19.98, 21.83] | - |
| | | LGB+ | 63,110 (0.7) | 35 (1.8) | 55.46 [38.63, 77.13] | - |
| Sex | Female | Total | 21,319,600 (100.0) | 2,960 (100.0) | 13.88 [13.39, 14.39] | 14.05 [13.43, 14.67] |
| | | Heterosexual | 20,563,625 (96.5) | 2,660 (89.9) | 12.94 [12.45, 13.44] | 12.48 [11.89, 13.08] |
| | | LGB+ | 755,980 (3.5) | 295 (10.0) | 39.02 [34.70, 43.74] | 39.51 [34.94, 44.08] |
| | | Gay or Lesbian | 266,445 (1.2) | 100 (3.4) | 37.53 [30.54, 45.65] | 36.74 [29.47, 44.01] |
| | | Bisexual | 397,245 (1.9) | 160 (5.4) | 40.28 [34.28, 47.02] | 41.30 [34.12, 48.47] |
| | | Other | 92,290 (0.4) | 35 (1.2) | 37.92 [26.42, 52.74] | 38.25 [26.33, 53.61] |
| | Male | Total | 19,878,720 (100.0) | 7,950 (100.0) | 39.99 [39.12, 40.88] | 38.78 [37.75, 39.81] |
| | | Heterosexual | 19,253,900 (96.9) | 7,535 (94.8) | 39.13 [38.26, 40.03] | 37.74 [36.70, 38.78] |
| | | LGB+ | 624,820 (3.1) | 420 (5.3) | 67.22 [60.94, 73.97] | 64.22 [57.90, 70.54] |
| | | Gay or Lesbian | 408,110 (2.1) | 260 (3.3) | 63.71 [56.20, 71.94] | 59.61 [51.79, 67.44] |
| | | Bisexual | 162,010 (0.8) | 120 (1.5) | 74.07 [61.41, 88.57] | 75.03 [61.41, 88.65] |
| | | Other | 54,695 (0.3) | 40 (0.5) | 73.13 [52.25, 99.59] | 75.28 [53.85, 102.43] |
| Region | England: North East | Total | 1,882,045 (100.0) | 625 (100.0) | 33.21 [30.66, 35.92] | 37.76 [34.22, 41.31] |
| | | Heterosexual | 1,823,065 (96.9) | 585 (93.6) | 32.09 [29.54, 34.80] | 36.54 [32.93, 40.15] |
| | | LGB+ | 58,980 (3.1) | 40 (6.4) | 67.82 [48.45, 92.35] | 69.55 [49.70, 94.69] |
| | England: North West | Total | 5,141,245 (100.0) | 1,555 (100.0) | 30.25 [28.76, 31.79] | 31.29 [29.44, 33.15] |
| | | Heterosexual | 4,969,855 (96.7) | 1,435 (92.3) | 28.87 [27.40, 30.41] | 29.15 [27.32, 30.98] |
| | | LGB+ | 171,390 (3.3) | 120 (7.7) | 70.02 [58.05, 83.72] | 70.49 [57.90, 83.07] |
| | England: Yorkshire and The Humber | Total | 3,796,000 (100.0) | 1,115 (100.0) | 29.37 [27.67, 31.15] | 30.03 [27.91, 32.14] |
| | | Heterosexual | 3,672,585 (96.7) | 1,045 (93.7) | 28.45 [26.75, 30.23] | 28.96 [26.82, 31.10] |
| | | LGB+ | 123,415 (3.3) | 65 (5.8) | 52.67 [40.65, 67.13] | 56.35 [43.70, 71.58] |
| | England: East Midlands | Total | 3,407,150 (100.0) | 935 (100.0) | 27.44 [25.71, 29.26] | 27.36 [25.23, 29.48] |
| | | Heterosexual | 3,304,925 (97.0) | 865 (92.5) | 26.17 [24.46, 27.98] | 25.26 [23.18, 27.34] |
| | | LGB+ | 102,225 (3.0) | 70 (7.5) | 68.48 [53.38, 86.52] | 68.32 [53.18, 86.41] |
| | England: West Midlands | Total | 4,059,830 (100.0) | 1,095 (100.0) | 26.97 [25.40, 28.62] | 26.25 [24.38, 28.13] |

| | | | | | | |
|---|---|---|---|---|---|---|
| | | Heterosexual | 3,949,510 (97.3) | 1,045 (95.4) | 26.46 [24.88, 28.11] | 25.77 [23.87, 27.67] |
| | | LGB+ | 110,320 (2.7) | 50 (4.6) | 45.32 [33.64, 59.75] | 44.87 [33.11, 59.40] |
| | England: East of England | Total | 4,428,750 (100.0) | 1,055 (100.0) | 23.82 [22.41, 25.30] | 23.09 [21.33, 24.85] |
| | | Heterosexual | 4,304,030 (97.2) | 990 (93.8) | 23.00 [21.59, 24.48] | 21.73 [19.99, 23.46] |
| | | LGB+ | 124,720 (2.8) | 65 (6.2) | 52.12 [40.22, 66.43] | 51.48 [39.68, 65.67] |
| | England: London | Total | 5,783,795 (100.0) | 1,005 (100.0) | 17.38 [16.32, 18.48] | 16.44 [15.27, 17.61] |
| | | Heterosexual | 5,512,495 (95.3) | 905 (90.0) | 16.42 [15.36, 17.52] | 15.39 [14.22, 16.55] |
| | | LGB+ | 271,300 (4.7) | 95 (9.5) | 35.02 [28.33, 42.81] | 34.88 [28.15, 42.71] |
| | England: South East | Total | 6,487,835 (100.0) | 1,690 (100.0) | 26.05 [24.82, 27.32] | 25.19 [23.67, 26.70] |
| | | Heterosexual | 6,274,920 (96.7) | 1,590 (94.1) | 25.34 [24.11, 26.62] | 24.26 [22.72, 25.79] |
| | | LGB+ | 212,915 (3.3) | 100 (5.9) | 46.97 [38.21, 57.12] | 46.86 [37.65, 56.07] |
| | England: South West | Total | 4,051,285 (100.0) | 1,195 (100.0) | 29.50 [27.85, 31.22] | 29.24 [27.13, 31.35] |
| | | Heterosexual | 3,914,520 (96.6) | 1,130 (94.6) | 28.87 [27.21, 30.60] | 28.48 [26.32, 30.63] |
| | | LGB+ | 136,765 (3.4) | 65 (5.4) | 47.53 [36.68, 60.58] | 48.52 [37.36, 61.94] |
| | Wales | Total | 2,160,385 (100.0) | 645 (100.0) | 29.86 [27.60, 32.25] | 29.34 [26.53, 32.15] |
| | | Heterosexual | 2,091,620 (96.8) | 600 (93.0) | 28.69 [26.44, 31.08] | 28.02 [25.19, 30.84] |
| | | LGB+ | 68,765 (3.2) | 40 (6.2) | 58.17 [41.56, 79.21] | 61.63 [44.25, 83.63] |
| Ethnic group | Asian, Asian British or Asian Welsh | Total | 3,371,415 (100.0) | 415 (100.0) | 12.31 [11.15, 13.55] | 12.02 [10.75, 13.28] |
| | | Heterosexual | 3,302,465 (98.0) | 395 (95.2) | 11.96 [10.81, 13.20] | 11.58 [10.32, 12.83] |
| | | LGB+ | 68,950 (2.0) | 20 (4.8) | 29.01 [17.72, 44.80] | 34.02 [20.90, 52.38] |
| | Black, Black British, Black Welsh, Caribbean or African | Total | 1,432,710 (100.0) | 190 (100.0) | 13.26 [11.44, 15.29] | 14.88 [12.52, 17.24] |
| | | Heterosexual | 1,402,805 (97.9) | 170 (89.5) | 12.12 [10.37, 14.08] | 13.63 [11.35, 15.91] |
| | | LGB+ | 29,910 (2.1) | 15 (7.9) | 50.15 [28.07, 82.72] | 56.42 [32.10, 92.28] |
| | Mixed or Multiple ethnic groups | Total | 782,190 (100.0) | 235 (100.0) | 30.04 [26.33, 34.14] | 30.53 [26.59, 34.46] |
| | | Heterosexual | 727,450 (93.0) | 200 (85.1) | 27.49 [23.81, 31.58] | 28.06 [24.13, 31.98] |
| | | LGB+ | 54,740 (7.0) | 35 (14.9) | 63.94 [44.54, 88.92] | 65.54 [43.89, 93.43] |
| | White | Total | 34,847,830 (100.0) | 9,980 (100.0) | 28.64 [28.08, 29.21] | 28.68 [27.98, 29.38] |
| | | Heterosexual | 33,648,035 (96.6) | 9,340 (93.6) | 27.76 [27.20, 28.33] | 27.45 [26.75, 28.16] |
| | | LGB+ | 1,199,795 (3.4) | 640 (6.4) | 53.34 [49.29, 57.64] | 53.23 [49.10, 57.36] |
| | Other ethnic group | Total | 764,175 (100.0) | 90 (100.0) | 11.78 [9.47, 14.48] | 11.26 [8.75, 14.20] |
| | | Heterosexual | 736,770 (96.4) | 85 (94.4) | 11.54 [9.22, 14.27] | 10.66 [8.22, 13.52] |
| | | LGB+ | 27,405 (3.6) | 5 (5.6) | 18.24 [5.92, 42.58] | 24.01 [6.93, 57.76] |
| Rural/urban classification | Urban | Total | 33,391,510 (100.0) | 8,665 (100.0) | 25.95 [25.41, 26.50] | 25.59 [24.95, 26.22] |
| | | Heterosexual | 32,189,260 (96.4) | 8,055 (93.0) | 25.02 [24.48, 25.58] | 24.35 [23.70, 24.99] |
| | | LGB+ | 1,202,250 (3.6) | 610 (7.0) | 50.74 [46.79, 54.93] | 51.04 [46.99, 55.09] |
| | | Gay or Lesbian | 582,585 (1.7) | 305 (3.5) | 52.35 [46.64, 58.57] | 49.09 [43.37, 54.81] |
| | | Bisexual | 488,625 (1.5) | 240 (2.8) | 49.12 [43.10, 55.74] | 53.15 [45.96, 60.35] |
| | | Other | 131,035 (0.4) | 70 (0.8) | 53.42 [41.64, 67.49] | 54.02 [41.82, 68.60] |
| | Rural | Total | 7,806,815 (100.0) | 2,245 (100.0) | 28.76 [27.58, 29.97] | 28.79 [27.15, 30.42] |
| | | Heterosexual | 7,628,265 (97.7) | 2,140 (95.3) | 28.05 [26.88, 29.27] | 27.64 [26.00, 29.28] |
| | | LGB+ | 178,550 (2.3) | 105 (4.7) | 58.81 [48.10, 71.19] | 57.60 [46.21, 68.98] |
| | | Gay or Lesbian | 91,970 (1.2) | 55 (2.4) | 59.80 [45.05, 77.84] | 60.35 [44.37, 79.89] |
| | | Bisexual | 70,630 (0.9) | 40 (1.8) | 56.63 [40.46, 77.12] | 61.29 [43.44, 83.90] |
| | | Other | 15,950 (0.2) | 5 (0.2) | 31.35 [10.18, 73.16] | 43.86 [17.37, 96.19] |
| Country of birth | Born in the UK | Total | 33,751,745 (100.0) | 9,715 (100.0) | 28.78 [28.21, 29.36] | 29.12 [28.41, 29.82] |

| | | | | | | |
|---|---|---|---|---|---|---|
| | | Heterosexual | 32,638,615 (96.7) | 9,075 (93.4) | 27.80 [27.24, 28.38] | 27.73 [27.02, 28.44] |
| | | LGB+ | 1,113,130 (3.3) | 640 (6.6) | 57.50 [53.13, 62.13] | 58.25 [53.73, 62.77] |
| | | Gay or Lesbian | 563,890 (1.7) | 330 (3.4) | 58.52 [52.38, 65.19] | 55.69 [49.48, 61.90] |
| | | Bisexual | 447,290 (1.3) | 245 (2.5) | 54.77 [48.13, 62.08] | 60.37 [52.24, 68.50] |
| | | Other | 101,950 (0.3) | 65 (0.7) | 63.76 [49.21, 81.26] | 69.20 [53.12, 88.54] |
| | Born outside the UK | Total | 7,446,575 (100.0) | 1,195 (100.0) | 16.05 [15.15, 16.98] | 15.61 [14.51, 16.70] |
| | | Heterosexual | 7,178,910 (96.4) | 1,125 (94.1) | 15.67 [14.77, 16.61] | 14.98 [13.88, 16.08] |
| | | LGB+ | 267,665 (3.6) | 75 (6.3) | 28.02 [22.04, 35.12] | 29.18 [22.65, 36.92] |
| | | Gay or Lesbian | 110,665 (1.5) | 30 (2.5) | 27.11 [18.29, 38.70] | 28.81 [17.23, 44.01] |
| | | Bisexual | 111,965 (1.5) | 35 (2.9) | 31.26 [21.77, 43.47] | 34.69 [23.83, 48.68] |
| | | Other | 45,040 (0.6) | 10 (0.8) | 22.20 [10.65, 40.83] | 23.87 [10.53, 45.38] |
| English language proficiency | Main language is English | Total | 37,542,140 (100.0) | 10,440 (100.0) | 27.81 [27.28, 28.35] | 27.87 [27.21, 28.52] |
| | | Heterosexual | 36,283,540 (96.6) | 9,745 (93.3) | 26.86 [26.33, 27.40] | 26.54 [25.88, 27.19] |
| | | LGB+ | 1,258,600 (3.4) | 695 (6.7) | 55.22 [51.19, 59.48] | 55.22 [51.11, 59.34] |
| | | Gay or Lesbian | 633,445 (1.7) | 350 (3.4) | 55.25 [49.62, 61.36] | 52.71 [46.96, 58.46] |
| | | Bisexual | 505,645 (1.3) | 270 (2.6) | 53.40 [47.22, 60.16] | 58.77 [51.33, 66.21] |
| | | Other | 119,505 (0.3) | 70 (0.7) | 58.57 [45.66, 74.01] | 61.51 [47.68, 78.02] |
| | Main language is not English | Total | 3,656,185 (100.0) | 475 (100.0) | 12.99 [11.85, 14.21] | 11.88 [10.61, 13.15] |
| | | Heterosexual | 3,533,985 (96.7) | 450 (94.7) | 12.73 [11.58, 13.97] | 11.69 [10.40, 12.98] |
| | | LGB+ | 122,200 (3.3) | 20 (4.2) | 16.37 [10.00, 25.28] | 18.48 [10.96, 28.99] |
| | | Gay or Lesbian | 41,110 (1.1) | 10 (2.1) | 24.32 [11.66, 44.73] | 16.24 [6.02, 32.70] |
| | | Bisexual | 53,610 (1.5) | 10 (2.1) | 18.65 [8.94, 34.30] | 16.53 [6.16, 33.26] |
| | | Other | 27,485 (0.8) | 5 (1.1) | 18.19 [5.91, 42.45] | 24.17 [7.36, 57.37] |
| Religion | No religion | Total | 15,126,775 (100.0) | 5,180 (100.0) | 34.24 [33.32, 35.19] | 32.41 [31.46, 33.37] |
| | | Heterosexual | 14,258,160 (94.3) | 4,710 (90.9) | 33.03 [32.10, 33.99] | 30.94 [29.97, 31.91] |
| | | LGB+ | 868,615 (5.7) | 470 (9.1) | 54.11 [49.33, 59.23] | 56.93 [51.65, 62.21] |
| | Christian | Total | 20,881,940 (100.0) | 4,720 (100.0) | 22.60 [21.96, 23.26] | 22.60 [21.67, 23.53] |
| | | Heterosexual | 20,524,925 (98.3) | 4,550 (96.4) | 22.17 [21.53, 22.82] | 21.85 [20.92, 22.77] |
| | | LGB+ | 357,015 (1.7) | 170 (3.6) | 47.62 [40.73, 55.34] | 46.93 [39.42, 54.45] |
| | Buddhist | Total | 202,525 (100.0) | 75 (100.0) | 37.03 [29.13, 46.42] | 29.61 [21.41, 39.33] |
| | | Heterosexual | 186,835 (92.3) | 65 (86.7) | 34.79 [26.85, 44.34] | 26.65 [18.49, 36.47] |
| | | LGB+ | 15,690 (7.7) | 10 (13.3) | 63.73 [30.56, 117.21] | 59.18 [28.57, 108.52] |
| | Hindu | Total | 693,140 (100.0) | 85 (100.0) | 12.26 [9.80, 15.16] | 12.40 [9.39, 15.93] |
| | | Heterosexual | 680,310 (98.1) | 80 (94.1) | 11.76 [9.32, 14.64] | 11.86 [8.91, 15.35] |
| | | LGB+ | 12,830 (1.9) | 5 (5.9) | 38.97 [12.65, 90.95] | 31.03 [6.01, 80.42] |
| | Jewish | Total | 178,245 (100.0) | 45 (100.0) | 25.25 [18.41, 33.78] | 20.99 [14.41, 29.22] |
| | | Heterosexual | 169,850 (95.3) | 40 (88.9) | 23.55 [16.82, 32.07] | 18.98 [12.62, 27.05] |
| | | LGB+ | 8,395 (4.7) | 5 (11.1) | 59.56 [19.34, 138.99] | 55.18 [17.52, 129.56] |
| | Muslim | Total | 2,162,660 (100.0) | 140 (100.0) | 6.47 [5.45, 7.64] | 6.43 [5.30, 7.56] |
| | | Heterosexual | 2,131,220 (98.5) | 135 (96.4) | 6.33 [5.31, 7.50] | 6.20 [5.08, 7.31] |
| | | LGB+ | 31,440 (1.5) | 5 (3.6) | 15.90 [5.16, 37.11] | 26.84 [10.41, 59.28] |
| | Sikh | Total | 354,395 (100.0) | 95 (100.0) | 26.81 [21.69, 32.77] | 23.91 [18.48, 30.23] |
| | | Heterosexual | 348,995 (98.5) | 90 (94.7) | 25.79 [20.74, 31.70] | 22.87 [17.56, 29.08] |
| | | LGB+ | 5,400 (1.5) | 5 (5.3) | 92.59 [30.06, 216.08] | 95.97 [23.99, 238.14] |
| | Other religion | Total | 256,045 (100.0) | 115 (100.0) | 44.91 [37.08, 53.91] | 46.81 [36.76, 56.86] |

| | | | | | | |
|---|---|---|---|---|---|---|
| | | Heterosexual | 214,080 (83.6) | 95 (82.6) | 44.38 [35.90, 54.25] | 44.10 [33.06, 56.97] |
| | | LGB+ | 41,965 (16.4) | 20 (17.4) | 47.66 [29.11, 73.61] | 49.08 [30.53, 75.03] |
| | Not answered | Total | 1,342,600 (100.0) | 455 (100.0) | 33.89 [30.85, 37.15] | 33.67 [29.93, 37.41] |
| | | Heterosexual | 1,303,150 (97.1) | 430 (94.5) | 33.00 [29.95, 36.27] | 32.89 [29.09, 36.70] |
| | | LGB+ | 39,450 (2.9) | 20 (4.4) | 50.70 [30.97, 78.30] | 56.36 [35.47, 85.57] |
| IMD/WIMD decile group | 1 (most deprived) | Total | 3,697,945 (100.0) | 1,230 (100.0) | 33.26 [31.43, 35.17] | 33.09 [31.03, 35.14] |
| | | Heterosexual | 3,552,730 (96.1) | 1,130 (91.9) | 31.81 [29.98, 33.72] | 31.35 [29.30, 33.41] |
| | | LGB+ | 145,215 (3.9) | 100 (8.1) | 68.86 [56.03, 83.76] | 70.61 [56.78, 84.44] |
| | | Gay or Lesbian | 67,800 (1.8) | 45 (3.7) | 66.37 [48.41, 88.81] | 67.57 [49.37, 90.30] |
| | | Bisexual | 60,455 (1.6) | 45 (3.7) | 74.44 [54.29, 99.60] | 81.25 [58.43, 109.76] |
| | | Other | 16,960 (0.5) | 10 (0.8) | 58.96 [28.27, 108.43] | 49.57 [19.07, 98.75] |
| | 2 | Total | 3,911,785 (100.0) | 1,070 (100.0) | 27.35 [25.74, 29.04] | 27.12 [25.28, 28.95] |
| | | Heterosexual | 3,752,330 (95.9) | 990 (92.5) | 26.38 [24.77, 28.08] | 25.85 [24.00, 27.69] |
| | | LGB+ | 159,455 (4.1) | 80 (7.5) | 50.17 [39.78, 62.44] | 50.12 [39.73, 62.39] |
| | | Gay or Lesbian | 75,145 (1.9) | 40 (3.7) | 53.23 [38.03, 72.48] | 51.48 [36.20, 70.83] |
| | | Bisexual | 65,475 (1.7) | 35 (3.3) | 53.46 [37.23, 74.34] | 51.25 [34.29, 73.09] |
| | | Other | 18,835 (0.5) | 10 (0.9) | 53.09 [25.46, 97.64] | 42.25 [18.00, 81.33] |
| | 3 | Total | 4,064,925 (100.0) | 1,085 (100.0) | 26.69 [25.13, 28.33] | 26.70 [24.88, 28.53] |
| | | Heterosexual | 3,893,450 (95.8) | 995 (91.7) | 25.56 [23.99, 27.19] | 25.03 [23.21, 26.84] |
| | | LGB+ | 171,475 (4.2) | 90 (8.3) | 52.49 [42.20, 64.51] | 52.96 [42.63, 65.05] |
| | | Gay or Lesbian | 82,610 (2.0) | 45 (4.1) | 54.47 [39.73, 72.89] | 51.64 [36.89, 70.07] |
| | | Bisexual | 69,120 (1.7) | 35 (3.2) | 50.64 [35.27, 70.42] | 46.63 [31.73, 65.81] |
| | | Other | 19,745 (0.5) | 15 (1.4) | 75.97 [42.52, 125.30] | 66.37 [35.16, 112.39] |
| | 4 | Total | 4,152,080 (100.0) | 1,150 (100.0) | 27.70 [26.12, 29.35] | 26.43 [24.63, 28.23] |
| | | Heterosexual | 3,985,065 (96.0) | 1,070 (93.0) | 26.85 [25.27, 28.51] | 25.52 [23.69, 27.34] |
| | | LGB+ | 167,015 (4.0) | 75 (6.5) | 44.91 [35.32, 56.29] | 45.89 [36.20, 57.41] |
| | | Gay or Lesbian | 81,310 (2.0) | 40 (3.5) | 49.19 [35.15, 66.99] | 46.02 [32.64, 62.97] |
| | | Bisexual | 67,530 (1.6) | 25 (2.2) | 37.02 [23.96, 54.65] | 43.28 [27.71, 64.30] |
| | | Other | 18,175 (0.4) | 5 (0.4) | 27.51 [8.93, 64.20] | 42.57 [18.74, 89.63] |
| | 5 | Total | 4,195,725 (100.0) | 1,195 (100.0) | 28.48 [26.89, 30.14] | 27.18 [25.30, 29.06] |
| | | Heterosexual | 4,046,600 (96.4) | 1,120 (93.7) | 27.68 [26.08, 29.35] | 26.19 [24.29, 28.09] |
| | | LGB+ | 149,120 (3.6) | 70 (5.9) | 46.94 [36.59, 59.31] | 47.94 [37.50, 60.42] |
| | | Gay or Lesbian | 72,645 (1.7) | 40 (3.3) | 55.06 [39.34, 74.98] | 45.81 [32.43, 62.75] |
| | | Bisexual | 60,800 (1.4) | 25 (2.1) | 41.12 [26.61, 60.70] | 38.77 [23.64, 59.18] |
| | | Other | 15,675 (0.4) | 10 (0.8) | 63.80 [30.59, 117.32] | 54.34 [25.22, 101.29] |
| | 6 | Total | 4,290,295 (100.0) | 1,130 (100.0) | 26.34 [24.83, 27.92] | 25.35 [23.53, 27.16] |
| | | Heterosexual | 4,145,555 (96.6) | 1,060 (93.8) | 25.57 [24.05, 27.16] | 24.20 [22.37, 26.02] |
| | | LGB+ | 144,745 (3.4) | 75 (6.6) | 51.82 [40.76, 64.95] | 50.35 [39.46, 63.29] |
| | | Gay or Lesbian | 72,075 (1.7) | 40 (3.5) | 55.50 [39.65, 75.57] | 49.00 [34.32, 67.59] |
| | | Bisexual | 57,895 (1.3) | 30 (2.7) | 51.82 [34.96, 73.97] | 55.46 [35.59, 81.58] |
| | | Other | 14,770 (0.3) | 5 (0.4) | 33.85 [10.99, 79.00] | 42.80 [16.66, 94.43] |
| | 7 | Total | 4,237,365 (100.0) | 1,110 (100.0) | 26.20 [24.68, 27.78] | 25.24 [23.34, 27.14] |
| | | Heterosexual | 4,111,035 (97.0) | 1,040 (93.7) | 25.30 [23.78, 26.88] | 24.19 [22.28, 26.10] |
| | | LGB+ | 126,325 (3.0) | 65 (5.9) | 51.45 [39.71, 65.58] | 52.71 [40.85, 66.99] |
| | | Gay or Lesbian | 63,370 (1.5) | 30 (2.7) | 47.34 [31.94, 67.58] | 45.30 [29.98, 65.44] |

| | | | | | | |
|---|---|---|---|---|---|---|
| | | Bisexual | 50,655 (1.2) | 25 (2.3) | 49.35 [31.94, 72.86] | 65.27 [41.95, 96.74] |
| | | Other | 12,305 (0.3) | 10 (0.9) | 81.27 [38.97, 149.45] | 83.26 [37.81, 156.54] |
| | 8 | Total | 4,258,970 (100.0) | 1,055 (100.0) | 24.77 [23.30, 26.31] | 24.75 [22.83, 26.66] |
| | | Heterosexual | 4,141,200 (97.2) | 1,000 (94.8) | 24.15 [22.67, 25.69] | 23.66 [21.75, 25.58] |
| | | LGB+ | 117,765 (2.8) | 55 (5.2) | 46.70 [35.18, 60.79] | 49.35 [37.39, 63.97] |
| | | Gay or Lesbian | 59,885 (1.4) | 30 (2.8) | 50.10 [33.80, 71.52] | 44.18 [28.64, 64.59] |
| | | Bisexual | 46,730 (1.1) | 25 (2.4) | 53.50 [34.62, 78.97] | 55.70 [35.07, 83.54] |
| | | Other | 11,150 (0.3) | 5 (0.5) | 44.84 [14.56, 104.65] | 45.60 [11.08, 113.77] |
| | 9 | Total | 4,222,750 (100.0) | 970 (100.0) | 22.97 [21.55, 24.46] | 23.13 [21.21, 25.05] |
| | | Heterosexual | 4,115,680 (97.5) | 920 (94.8) | 22.35 [20.93, 23.85] | 22.19 [20.26, 24.13] |
| | | LGB+ | 107,070 (2.5) | 50 (5.2) | 46.70 [34.66, 61.57] | 47.53 [35.37, 62.55] |
| | | Gay or Lesbian | 54,355 (1.3) | 30 (3.1) | 55.19 [37.24, 78.79] | 55.44 [36.20, 80.73] |
| | | Bisexual | 42,540 (1.0) | 20 (2.1) | 47.01 [28.72, 72.61] | 49.49 [28.26, 79.19] |
| | | Other | 10,175 (0.2) | 5 (0.5) | 49.14 [15.96, 114.68] | 32.07 [5.52, 84.50] |
| | 10 (least deprived) | Total | 4,166,490 (100.0) | 920 (100.0) | 22.08 [20.68, 23.56] | 21.40 [19.49, 23.31] |
| | | Heterosexual | 4,073,880 (97.8) | 875 (95.1) | 21.48 [20.08, 22.95] | 20.29 [18.39, 22.19] |
| | | LGB+ | 92,610 (2.2) | 45 (4.9) | 48.59 [35.44, 65.02] | 47.70 [34.45, 64.27] |
| | | Gay or Lesbian | 45,365 (1.1) | 20 (2.2) | 44.09 [26.93, 68.09] | 47.55 [28.15, 74.68] |
| | | Bisexual | 38,050 (0.9) | 20 (2.2) | 52.56 [32.11, 81.18] | 51.90 [28.42, 84.74] |
| | | Other | 9,195 (0.2) | 5 (0.5) | 54.38 [17.66, 126.90] | 79.53 [30.28, 176.82] |
| Highest level of qualification (≥25 years) | No qualifications | Total | 6,640,315 (100.0) | 1,975 (100.0) | 29.74 [28.45, 31.08] | 42.39 [39.59, 45.19] |
| | | Heterosexual | 6,552,600 (98.7) | 1,910 (96.7) | 29.15 [27.86, 30.49] | 41.95 [39.10, 44.80] |
| | | LGB+ | 87,720 (1.3) | 60 (3.0) | 68.40 [52.20, 88.04] | 66.43 [50.06, 86.28] |
| | Level 1 (one to four GCSE passes (grade A* to C or grade 4 and above) and any other GCSEs at other grades, or equivalent qualifications) | Total | 3,547,145 (100.0) | 1,025 (100.0) | 28.90 [27.15, 30.72] | 34.07 [31.35, 36.79] |
| | | Heterosexual | 3,480,285 (98.1) | 980 (95.6) | 28.16 [26.42, 29.98] | 33.28 [30.55, 36.02] |
| | | LGB+ | 66,860 (1.9) | 40 (3.9) | 59.83 [42.74, 81.47] | 62.15 [43.96, 85.19] |
| | Level 2 (five or more GCSE passes (grade A* to C or grade 4 and above) or equivalent qualifications) | Total | 4,308,070 (100.0) | 1,360 (100.0) | 31.57 [29.91, 33.29] | 34.14 [31.99, 36.29] |
| | | Heterosexual | 4,199,885 (97.5) | 1,295 (95.2) | 30.83 [29.18, 32.56] | 33.37 [31.19, 35.54] |
| | | LGB+ | 108,190 (2.5) | 65 (4.8) | 60.08 [46.37, 76.58] | 58.53 [44.85, 74.99] |
| | Apprenticeship | Total | 2,013,560 (100.0) | 740 (100.0) | 36.75 [34.15, 39.50] | 40.01 [35.97, 44.04] |
| | | Heterosexual | 1,986,265 (98.6) | 730 (98.6) | 36.75 [34.13, 39.52] | 40.09 [35.99, 44.20] |
| | | LGB+ | 27,295 (1.4) | 10 (1.4) | 36.64 [17.57, 67.38] | 40.52 [20.62, 72.59] |
| | Level 3 (two or more A Levels or equivalent qualifications) | Total | 5,447,045 (100.0) | 1,545 (100.0) | 28.36 [26.97, 29.81] | 28.94 [27.37, 30.51] |
| | | Heterosexual | 5,279,280 (96.9) | 1,430 (92.6) | 27.09 [25.70, 28.53] | 27.29 [25.74, 28.85] |
| | | LGB+ | 167,765 (3.1) | 115 (7.4) | 68.55 [56.59, 82.28] | 68.40 [55.83, 80.97] |
| | Level 4 and above (Higher National Certificate, Higher National Diploma, Bachelor's degree, or | Total | 13,103,645 (100.0) | 2,935 (100.0) | 22.40 [21.60, 23.22] | 20.64 [19.82, 21.46] |
| | | Heterosexual | 12,583,195 (96.0) | 2,685 (91.5) | 21.34 [20.54, 22.16] | 19.30 [18.49, 20.11] |
| | | LGB+ | 520,445 (4.0) | 255 (8.7) | 49.00 [43.17, 55.39] | 49.54 [43.41, 55.66] |

| | | | | | | |
|---|---|---|---|---|---|---|
| | post-graduate qualifications) | | | | | |
| | Other qualifications | Total | 1,032,785 (100.0) | 245 (100.0) | 23.72 [20.84, 26.89] | 29.65 [24.60, 34.70] |
| | | Heterosexual | 1,016,560 (98.4) | 235 (95.9) | 23.12 [20.26, 26.27] | 28.88 [23.79, 33.96] |
| | | LGB+ | 16,220 (1.6) | 10 (4.1) | 61.65 [29.56, 113.38] | 61.23 [28.66, 113.73] |
| NS-SEC | Class 1 (Higher managerial, administrative and professional occupations) | Total | 5,565,200 (100.0) | 1,190 (100.0) | 21.38 [20.19, 22.63] | 17.25 [14.94, 19.57] |
| | | Heterosexual | 5,373,260 (96.6) | 1,115 (93.7) | 20.75 [19.55, 22.01] | 16.39 [14.03, 18.76] |
| | | LGB+ | 191,940 (3.4) | 75 (6.3) | 39.07 [30.73, 48.98] | 34.63 [26.22, 44.62] |
| | Class 2 (Lower managerial, administrative and professional occupations) | Total | 8,415,665 (100.0) | 1,675 (100.0) | 19.90 [18.96, 20.88] | 17.72 [15.99, 19.45] |
| | | Heterosexual | 8,100,895 (96.3) | 1,540 (91.9) | 19.01 [18.07, 19.98] | 16.47 [14.82, 18.12] |
| | | LGB+ | 314,770 (3.7) | 135 (8.1) | 42.89 [35.96, 50.76] | 42.27 [28.58, 55.96] |
| | Class 3 (Intermediate occupations) | Total | 4,821,360 (100.0) | 940 (100.0) | 19.50 [18.27, 20.78] | 19.56 [17.74, 21.39] |
| | | Heterosexual | 4,682,910 (97.1) | 865 (92.0) | 18.47 [17.26, 19.74] | 18.30 [16.46, 20.14] |
| | | LGB+ | 138,450 (2.9) | 75 (8.0) | 54.17 [42.61, 67.90] | 49.05 [38.37, 61.73] |
| | Class 4 (Small employers and own account workers) | Total | 4,352,905 (100.0) | 1,680 (100.0) | 38.59 [36.77, 40.49] | 39.81 [36.07, 43.55] |
| | | Heterosexual | 4,266,820 (98.0) | 1,630 (97.0) | 38.20 [36.37, 40.10] | 39.77 [35.94, 43.59] |
| | | LGB+ | 86,085 (2.0) | 50 (3.0) | 58.08 [43.11, 76.57] | 44.76 [30.58, 62.27] |
| | Class 5 (Lower supervisory and technical occupations) | Total | 2,251,985 (100.0) | 755 (100.0) | 33.53 [31.18, 36.01] | 33.53 [30.25, 36.82] |
| | | Heterosexual | 2,197,490 (97.6) | 720 (95.4) | 32.76 [30.41, 35.25] | 32.82 [29.50, 36.15] |
| | | LGB+ | 54,500 (2.4) | 30 (4.0) | 55.05 [37.14, 78.58] | 51.90 [35.47, 73.50] |
| | Class 6 (Semi-routine occupations) | Total | 4,745,535 (100.0) | 1,250 (100.0) | 26.34 [24.90, 27.84] | 29.00 [27.01, 31.00] |
| | | Heterosexual | 4,597,870 (96.9) | 1,145 (91.6) | 24.90 [23.48, 26.39] | 26.76 [24.82, 28.71] |
| | | LGB+ | 147,665 (3.1) | 105 (8.4) | 71.11 [58.16, 86.08] | 73.07 [58.52, 87.61] |
| | Class 7 (Routine occupations) | Total | 4,963,690 (100.0) | 1,730 (100.0) | 34.85 [33.23, 36.54] | 38.58 [36.28, 40.88] |
| | | Heterosexual | 4,839,745 (97.5) | 1,635 (94.5) | 33.78 [32.16, 35.46] | 37.20 [34.89, 39.51] |
| | | LGB+ | 123,945 (2.5) | 90 (5.2) | 72.61 [58.39, 89.25] | 74.58 [59.78, 91.89] |
| | Class 8 (Never worked and long-term unemployed) | Total | 3,273,875 (100.0) | 1,215 (100.0) | 37.11 [35.05, 39.26] | 44.60 [41.61, 47.59] |
| | | Heterosexual | 3,177,115 (97.0) | 1,135 (93.4) | 35.72 [33.68, 37.86] | 42.59 [39.59, 45.59] |
| | | LGB+ | 96,765 (3.0) | 80 (6.6) | 82.67 [65.56, 102.90] | 85.80 [67.95, 106.87] |
| | Full-time students | Total | 2,808,105 (100.0) | 485 (100.0) | 17.27 [15.77, 18.88] | 23.68 [19.03, 28.33] |
| | | Heterosexual | 2,581,420 (91.9) | 410 (84.5) | 15.88 [14.38, 17.50] | 21.32 [16.87, 25.76] |
| | | LGB+ | 226,685 (8.1) | 70 (14.4) | 30.88 [24.07, 39.01] | 77.75 [17.35, 149.93] |
| Household NS-SEC | Class 1 (Higher managerial, administrative and professional occupations) | Total | 10,270,425 (100.0) | 2,050 (100.0) | 19.96 [19.11, 20.84] | 18.66 [17.64, 19.68] |
| | | Heterosexual | 9,913,780 (96.5) | 1,915 (93.4) | 19.32 [18.46, 20.20] | 17.77 [16.75, 18.79] |
| | | LGB+ | 356,645 (3.5) | 135 (6.6) | 37.85 [31.74, 44.80] | 38.06 [31.48, 44.65] |
| | Class 2 (Lower managerial, administrative and professional occupations) | Total | 11,489,885 (100.0) | 2,590 (100.0) | 22.54 [21.68, 23.43] | 21.53 [20.52, 22.55] |
| | | Heterosexual | 11,078,640 (96.4) | 2,405 (92.9) | 21.71 [20.85, 22.59] | 20.45 [19.43, 21.46] |
| | | LGB+ | 411,245 (3.6) | 185 (7.1) | 44.99 [38.74, 51.96] | 44.11 [37.73, 50.49] |
| | Class 3 (Intermediate occupations) | Total | 5,198,695 (100.0) | 1,265 (100.0) | 24.33 [23.01, 25.71] | 25.32 [23.64, 26.99] |
| | | Heterosexual | 5,057,880 (97.3) | 1,190 (94.1) | 23.53 [22.21, 24.90] | 24.26 [22.58, 25.94] |
| | | LGB+ | 140,810 (2.7) | 75 (5.9) | 53.26 [41.89, 66.77] | 54.48 [42.87, 68.26] |
| | | Total | 3,976,970 (100.0) | 1,375 (100.0) | 34.57 [32.77, 36.45] | 33.84 [31.52, 36.15] |

| | | | | | | |
|---|---|---|---|---|---|---|
| | Class 4 (Small employers and own account workers) | Heterosexual | 3,885,775 (97.7) | 1,330 (96.7) | 34.23 [32.41, 36.12] | 33.32 [30.99, 35.66] |
| | | LGB+ | 91,200 (2.3) | 45 (3.3) | 49.34 [35.99, 66.02] | 49.21 [35.46, 66.40] |
| | Class 5 (Lower supervisory and technical occupations) | Total | 1,954,560 (100.0) | 605 (100.0) | 30.95 [28.54, 33.52] | 31.22 [28.15, 34.29] |
| | | Heterosexual | 1,900,420 (97.2) | 575 (95.0) | 30.26 [27.83, 32.83] | 30.19 [27.10, 33.28] |
| | | LGB+ | 54,140 (2.8) | 30 (5.0) | 55.41 [37.39, 79.10] | 56.00 [37.88, 79.81] |
| | Class 6 (Semi-routine occupations) | Total | 3,340,505 (100.0) | 1,020 (100.0) | 30.53 [28.69, 32.47] | 33.97 [31.49, 36.45] |
| | | Heterosexual | 3,232,530 (96.8) | 935 (91.7) | 28.92 [27.10, 30.84] | 31.59 [29.14, 34.05] |
| | | LGB+ | 107,975 (3.2) | 85 (8.3) | 78.72 [62.88, 97.34] | 80.02 [63.95, 98.91] |
| | Class 7 (Routine occupations) | Total | 2,439,520 (100.0) | 1,000 (100.0) | 40.99 [38.49, 43.61] | 43.87 [40.50, 47.24] |
| | | Heterosexual | 2,366,810 (97.0) | 935 (93.5) | 39.50 [37.01, 42.12] | 41.90 [38.52, 45.28] |
| | | LGB+ | 72,705 (3.0) | 65 (6.5) | 89.40 [69.00, 113.95] | 89.25 [68.60, 114.08] |
| | Class 8 (Never worked and long-term unemployed) | Total | 1,655,055 (100.0) | 695 (100.0) | 41.99 [38.93, 45.23] | 54.38 [49.15, 59.62] |
| | | Heterosexual | 1,605,800 (97.0) | 655 (94.2) | 40.79 [37.73, 44.04] | 51.80 [46.55, 57.06] |
| | | LGB+ | 49,260 (3.0) | 45 (6.5) | 91.35 [66.63, 122.24] | 93.10 [67.20, 125.47] |
| | Full-time students | Total | 386,235 (100.0) | 95 (100.0) | 24.60 [19.90, 30.07] | 30.59 [21.51, 41.17] |
| | | Heterosexual | 329,545 (85.3) | 70 (73.7) | 21.24 [16.56, 26.84] | 27.23 [18.82, 37.27] |
| | | LGB+ | 56,690 (14.7) | 20 (21.1) | 35.28 [21.55, 54.49] | 91.83 [0.00, 240.36] |
| | Living in a communal establishment | Total | 486,475 (100.0) | 210 (100.0) | 43.17 [37.53, 49.42] | 93.81 [79.03, 108.58] |
| | | Heterosexual | 446,340 (91.7) | 185 (88.1) | 41.45 [35.69, 47.87] | 89.35 [74.45, 104.25] |
| | | LGB+ | 40,135 (8.3) | 25 (11.9) | 62.29 [40.31, 91.95] | 152.45 [84.92, 243.60] |
| Lone person household indicator | Lives alone | Total | 5,954,715 (100.0) | 3,285 (100.0) | 55.17 [53.30, 57.09] | 76.92 [70.25, 83.60] |
| | | Heterosexual | 5,697,830 (95.7) | 3,010 (91.6) | 52.83 [50.96, 54.75] | 70.28 [63.80, 76.77] |
| | | LGB+ | 256,880 (4.3) | 270 (8.2) | 105.11 [92.94, 118.42] | 124.16 [99.11, 149.20] |
| | Does not live alone | Total | 34,757,135 (100.0) | 7,420 (100.0) | 21.35 [20.87, 21.84] | 21.53 [20.96, 22.11] |
| | | Heterosexual | 33,673,350 (96.9) | 7,000 (94.3) | 20.79 [20.30, 21.28] | 20.76 [20.19, 21.34] |
| | | LGB+ | 1,083,785 (3.1) | 420 (5.7) | 38.75 [35.14, 42.64] | 38.90 [35.14, 42.67] |
| | Living in a communal establishment | Total | 486,475 (100.0) | 210 (100.0) | 43.17 [37.53, 49.42] | 93.81 [79.03, 108.58] |
| | | Heterosexual | 446,340 (91.7) | 185 (88.1) | 41.45 [35.69, 47.87] | 89.35 [74.45, 104.25] |
| | | LGB+ | 40,135 (8.3) | 25 (11.9) | 62.29 [40.31, 91.95] | 152.45 [84.92, 243.60] |
| Household size | One person in household | Total | 5,954,715 (100.0) | 3,285 (100.0) | 55.17 [53.30, 57.09] | 76.92 [70.25, 83.60] |
| | | Heterosexual | 5,697,830 (95.7) | 3,010 (91.6) | 52.83 [50.96, 54.75] | 70.28 [63.80, 76.77] |
| | | LGB+ | 256,880 (4.3) | 270 (8.2) | 105.11 [92.94, 118.42] | 124.16 [99.11, 149.20] |
| | Two people in household | Total | 14,026,325 (100.0) | 3,255 (100.0) | 23.21 [22.42, 24.02] | 26.02 [24.65, 27.39] |
| | | Heterosexual | 13,512,625 (96.3) | 3,055 (93.9) | 22.61 [21.81, 23.42] | 25.30 [23.87, 26.72] |
| | | LGB+ | 513,705 (3.7) | 200 (6.1) | 38.93 [33.72, 44.72] | 36.29 [30.90, 41.68] |
| | Three to five people in household | Total | 18,767,340 (100.0) | 3,845 (100.0) | 20.49 [19.85, 21.15] | 20.71 [19.99, 21.43] |
| | | Heterosexual | 18,251,775 (97.3) | 3,640 (94.7) | 19.94 [19.30, 20.60] | 20.01 [19.28, 20.73] |
| | | LGB+ | 515,565 (2.7) | 205 (5.3) | 39.76 [34.51, 45.59] | 42.00 [35.08, 48.91] |
| | Six or more people in household | Total | 1,963,470 (100.0) | 320 (100.0) | 16.30 [14.56, 18.18] | 16.63 [14.72, 18.55] |
| | | Heterosexual | 1,908,955 (97.2) | 305 (95.3) | 15.98 [14.23, 17.87] | 16.25 [14.33, 18.17] |
| | | LGB+ | 54,515 (2.8) | 15 (4.7) | 27.52 [15.40, 45.38] | 28.07 [13.63, 49.37] |
| | Living in a communal establishment | Total | 486,475 (100.0) | 210 (100.0) | 43.17 [37.53, 49.42] | 93.81 [79.03, 108.58] |
| | | Heterosexual | 446,340 (91.7) | 185 (88.1) | 41.45 [35.69, 47.87] | 89.35 [74.45, 104.25] |
| | | LGB+ | 40,135 (8.3) | 25 (11.9) | 62.29 [40.31, 91.95] | 152.45 [84.92, 243.60] |

| | | | | | | |
|---|---|---|---|---|---|---|
| Household tenure | Owned or shared ownership | Total | 27,298,305 (100.0) | 6,100 (100.0) | 22.35 [21.79, 22.91] | 21.21 [20.47, 21.95] |
| | | Heterosexual | 26,653,165 (97.6) | 5,820 (95.4) | 21.84 [21.28, 22.40] | 20.47 [19.73, 21.21] |
| | | LGB+ | 645,140 (2.4) | 280 (4.6) | 43.40 [38.47, 48.79] | 41.54 [36.46, 46.62] |
| | Social rented | Total | 6,069,620 (100.0) | 2,275 (100.0) | 37.48 [35.96, 39.05] | 38.25 [36.47, 40.03] |
| | | Heterosexual | 5,853,620 (96.4) | 2,105 (92.5) | 35.96 [34.44, 37.53] | 36.28 [34.50, 38.05] |
| | | LGB+ | 216,000 (3.6) | 170 (7.5) | 78.70 [67.32, 91.46] | 79.07 [67.06, 91.08] |
| | Private rented or living rent free | Total | 7,343,925 (100.0) | 2,330 (100.0) | 31.73 [30.45, 33.04] | 30.47 [29.19, 31.76] |
| | | Heterosexual | 6,864,400 (93.5) | 2,085 (89.5) | 30.37 [29.08, 31.71] | 28.84 [27.55, 30.13] |
| | | LGB+ | 479,525 (6.5) | 245 (10.5) | 51.09 [44.89, 57.91] | 56.86 [48.96, 64.75] |
| | Living in a communal establishment | Total | 486,475 (100.0) | 210 (100.0) | 43.17 [37.53, 49.42] | 93.81 [79.03, 108.58] |
| | | Heterosexual | 446,340 (91.7) | 185 (88.1) | 41.45 [35.69, 47.87] | 89.35 [74.45, 104.25] |
| | | LGB+ | 40,135 (8.3) | 25 (11.9) | 62.29 [40.31, 91.95] | 152.45 [84.92, 243.60] |
| Economic activity | Economically active | Total | 25,443,735 (100.0) | 5,945 (100.0) | 23.37 [22.78, 23.97] | 22.49 [21.81, 23.18] |
| | | Heterosexual | 24,463,115 (96.1) | 5,515 (92.8) | 22.54 [21.95, 23.15] | 21.52 [20.84, 22.21] |
| | | LGB+ | 980,620 (3.9) | 430 (7.2) | 43.85 [39.80, 48.20] | 43.67 [39.24, 48.10] |
| | | Gay or Lesbian | 510,450 (2.0) | 220 (3.7) | 43.10 [37.59, 49.19] | 42.59 [36.14, 49.05] |
| | | Bisexual | 375,785 (1.5) | 165 (2.8) | 43.91 [37.46, 51.14] | 48.49 [40.24, 56.74] |
| | | Other | 94,385 (0.4) | 45 (0.8) | 47.68 [34.78, 63.80] | 47.52 [33.67, 64.84] |
| | Economically inactive | Total | 15,754,590 (100.0) | 4,965 (100.0) | 31.51 [30.64, 32.40] | 44.61 [42.80, 46.41] |
| | | Heterosexual | 15,354,410 (97.5) | 4,685 (94.4) | 30.51 [29.64, 31.40] | 42.52 [40.70, 44.33] |
| | | LGB+ | 400,180 (2.5) | 285 (5.7) | 71.22 [63.19, 79.98] | 84.00 [73.29, 94.70] |
| | | Gay or Lesbian | 164,110 (1.0) | 140 (2.8) | 85.31 [71.76, 100.67] | 105.74 [85.38, 126.11] |
| | | Bisexual | 183,470 (1.2) | 115 (2.3) | 62.68 [51.75, 75.24] | 73.27 [58.06, 88.48] |
| | | Other | 52,605 (0.3) | 30 (0.6) | 57.03 [38.48, 81.41] | 58.45 [38.19, 85.09] |
| General health (detailed) | Very good health | Total | 16,934,985 (100.0) | 2,840 (100.0) | 16.77 [16.16, 17.40] | 16.54 [15.89, 17.19] |
| | | Heterosexual | 16,399,805 (96.8) | 2,675 (94.2) | 16.31 [15.70, 16.94] | 16.02 [15.37, 16.67] |
| | | LGB+ | 535,180 (3.2) | 165 (5.8) | 30.83 [26.31, 35.91] | 31.68 [26.77, 36.58] |
| | | Gay or Lesbian | 292,080 (1.7) | 95 (3.3) | 32.53 [26.31, 39.76] | 32.79 [26.47, 40.14] |
| | | Bisexual | 194,705 (1.1) | 55 (1.9) | 28.25 [21.28, 36.77] | 35.02 [25.32, 46.88] |
| | | Other | 48,395 (0.3) | 10 (0.4) | 20.66 [9.91, 38.00] | 25.43 [12.68, 45.99] |
| | Good health | Total | 15,579,320 (100.0) | 3,835 (100.0) | 24.62 [23.84, 25.41] | 26.13 [25.07, 27.20] |
| | | Heterosexual | 15,036,325 (96.5) | 3,600 (93.9) | 23.94 [23.17, 24.74] | 25.35 [24.25, 26.45] |
| | | LGB+ | 542,995 (3.5) | 240 (6.3) | 44.20 [38.78, 50.16] | 43.96 [38.37, 49.55] |
| | | Gay or Lesbian | 254,750 (1.6) | 130 (3.4) | 51.03 [42.64, 60.59] | 49.34 [40.47, 58.21] |
| | | Bisexual | 230,325 (1.5) | 85 (2.2) | 36.90 [29.48, 45.63] | 41.60 [32.51, 52.29] |
| | | Other | 57,920 (0.4) | 20 (0.5) | 34.53 [21.09, 53.33] | 39.28 [23.92, 60.77] |
| | Fair health | Total | 6,156,315 (100.0) | 2,495 (100.0) | 40.53 [38.95, 42.15] | 56.64 [53.18, 60.10] |
| | | Heterosexual | 5,946,370 (96.6) | 2,310 (92.6) | 38.85 [37.28, 40.46] | 53.90 [50.27, 57.53] |
| | | LGB+ | 209,945 (3.4) | 180 (7.2) | 85.74 [73.67, 99.22] | 86.91 [74.07, 99.74] |
| | | Gay or Lesbian | 88,170 (1.4) | 75 (3.0) | 85.06 [66.91, 106.63] | 84.44 [64.43, 108.22] |
| | | Bisexual | 95,245 (1.5) | 80 (3.2) | 83.99 [66.60, 104.54] | 87.13 [68.88, 108.68] |
| | | Other | 26,535 (0.4) | 25 (1.0) | 94.22 [60.97, 139.08] | 91.96 [58.74, 136.79] |
| | Bad health | Total | 1,963,555 (100.0) | 1,270 (100.0) | 64.68 [61.17, 68.34] | 97.65 [88.47, 106.83] |
| | | Heterosexual | 1,890,340 (96.3) | 1,175 (92.5) | 62.16 [58.65, 65.82] | 90.96 [81.47, 100.44] |
| | | LGB+ | 73,215 (3.7) | 95 (7.5) | 129.75 [104.98, 158.62] | 136.45 [108.76, 168.72] |

| | | | | | | |
|---|---|---|---|---|---|---|
| | | Gay or Lesbian | 30,720 (1.6) | 40 (3.1) | 130.21 [93.02, 177.31] | 115.14 [75.34, 165.55] |
| | | Bisexual | 31,695 (1.6) | 40 (3.1) | 126.20 [90.16, 171.85] | 136.61 [97.50, 186.16] |
| | | Other | 10,805 (0.6) | 15 (1.2) | 138.82 [77.70, 228.97] | 144.84 [81.05, 238.92] |
| | Very bad health | Total | 564,145 (100.0) | 470 (100.0) | 83.31 [75.95, 91.20] | 132.18 [111.09, 153.26] |
| | | Heterosexual | 544,685 (96.6) | 435 (92.6) | 79.86 [72.53, 87.73] | 122.02 [101.03, 143.01] |
| | | LGB+ | 19,460 (3.4) | 35 (7.4) | 179.86 [125.28, 250.14] | 203.53 [130.44, 297.65] |
| | | Gay or Lesbian | 8,840 (1.6) | 15 (3.2) | 169.68 [94.97, 279.87] | 178.12 [86.29, 313.54] |
| | | Bisexual | 7,290 (1.3) | 15 (3.2) | 205.76 [115.16, 339.37] | 235.93 [120.45, 406.24] |
| | | Other | 3,330 (0.6) | 5 (1.1) | 150.15 [48.75, 350.40] | 140.02 [18.63, 379.75] |
| General health (aggregated) | Good health | Total | 32,514,310 (100.0) | 6,680 (100.0) | 20.54 [20.06, 21.04] | 20.42 [19.86, 20.98] |
| | | Heterosexual | 31,436,130 (96.7) | 6,275 (93.9) | 19.96 [19.47, 20.46] | 19.75 [19.18, 20.31] |
| | | LGB+ | 1,078,175 (3.3) | 405 (6.1) | 37.56 [33.99, 41.41] | 37.91 [34.20, 41.62] |
| | | Gay or Lesbian | 546,830 (1.7) | 230 (3.4) | 42.06 [36.80, 47.86] | 40.54 [35.13, 45.94] |
| | | Bisexual | 425,030 (1.3) | 140 (2.1) | 32.94 [27.71, 38.87] | 38.57 [31.56, 45.59] |
| | | Other | 106,315 (0.3) | 35 (0.5) | 32.92 [22.93, 45.79] | 33.49 [22.50, 47.63] |
| | Not good health | Total | 8,684,015 (100.0) | 4,235 (100.0) | 48.77 [47.31, 50.26] | 68.60 [65.30, 71.89] |
| | | Heterosexual | 8,381,395 (96.5) | 3,920 (92.6) | 46.77 [45.32, 48.26] | 64.64 [61.22, 68.06] |
| | | LGB+ | 302,620 (3.5) | 310 (7.3) | 102.44 [91.35, 114.50] | 103.89 [92.07, 115.72] |
| | | Gay or Lesbian | 127,725 (1.5) | 130 (3.1) | 101.78 [85.04, 120.86] | 97.97 [78.80, 117.15] |
| | | Bisexual | 134,225 (1.5) | 140 (3.3) | 104.30 [87.74, 123.08] | 103.21 [85.88, 120.54] |
| | | Other | 40,670 (0.5) | 45 (1.1) | 110.65 [80.71, 148.05] | 106.19 [76.74, 142.97] |
| Disability | Day-to-day activities limited a lot | Total | 3,441,160 (100.0) | 2,135 (100.0) | 62.04 [59.44, 64.73] | 90.50 [84.98, 96.02] |
| | | Heterosexual | 3,307,285 (96.1) | 1,955 (91.6) | 59.11 [56.52, 61.79] | 84.80 [79.13, 90.47] |
| | | LGB+ | 133,875 (3.9) | 180 (8.4) | 134.45 [115.53, 155.59] | 136.48 [116.20, 156.76] |
| | | Gay or Lesbian | 55,805 (1.6) | 75 (3.5) | 134.40 [105.71, 168.47] | 135.38 [104.09, 172.55] |
| | | Bisexual | 58,395 (1.7) | 80 (3.7) | 137.00 [108.63, 170.51] | 137.14 [108.83, 170.57] |
| | | Other | 19,675 (0.6) | 20 (0.9) | 101.65 [62.09, 156.99] | 107.73 [66.30, 165.69] |
| | Day-to-day activities limited a little | Total | 4,765,825 (100.0) | 2,185 (100.0) | 45.85 [43.94, 47.81] | 56.90 [53.83, 59.96] |
| | | Heterosexual | 4,491,165 (94.2) | 1,975 (90.4) | 43.98 [42.06, 45.96] | 54.92 [51.64, 58.20] |
| | | LGB+ | 274,660 (5.8) | 210 (9.6) | 76.46 [66.47, 87.53] | 79.54 [68.58, 90.50] |
| | | Gay or Lesbian | 100,800 (2.1) | 95 (4.3) | 94.25 [76.25, 115.21] | 96.44 [77.47, 118.53] |
| | | Bisexual | 138,475 (2.9) | 85 (3.9) | 61.38 [49.03, 75.90] | 72.97 [57.21, 91.49] |
| | | Other | 35,385 (0.7) | 25 (1.1) | 70.65 [45.72, 104.30] | 81.31 [51.62, 121.38] |
| | Day-to-day activities not limited | Total | 32,991,340 (100.0) | 6,590 (100.0) | 19.97 [19.50, 20.46] | 19.30 [18.75, 19.85] |
| | | Heterosexual | 32,019,080 (97.1) | 6,265 (95.1) | 19.57 [19.08, 20.06] | 18.84 [18.28, 19.40] |
| | | LGB+ | 972,260 (2.9) | 325 (4.9) | 33.43 [29.89, 37.27] | 33.42 [29.79, 37.05] |
| | | Gay or Lesbian | 517,945 (1.6) | 190 (2.9) | 36.68 [31.65, 42.29] | 33.98 [28.92, 39.04] |
| | | Bisexual | 362,385 (1.1) | 110 (1.7) | 30.35 [24.95, 36.59] | 34.20 [27.40, 40.99] |
| | | Other | 91,930 (0.3) | 30 (0.5) | 32.63 [22.02, 46.59] | 31.10 [20.36, 45.22] |
| Residence type | Living in a private household | Total | 40,711,850 (100.0) | 10,700 (100.0) | 26.28 [25.79, 26.79] | 25.74 [25.15, 26.34] |
| | | Heterosexual | 39,371,185 (96.7) | 10,010 (93.6) | 25.42 [24.93, 25.93] | 24.56 [23.96, 25.15] |
| | | LGB+ | 1,340,665 (3.3) | 690 (6.4) | 51.47 [47.70, 55.45] | 51.13 [47.31, 54.95] |
| | Living in a communal establishment | Total | 486,475 (100.0) | 210 (100.0) | 43.17 [37.53, 49.42] | 93.81 [79.03, 108.58] |
| | | Heterosexual | 446,340 (91.7) | 185 (88.1) | 41.45 [35.69, 47.87] | 89.35 [74.45, 104.25] |
| | | LGB+ | 40,135 (8.3) | 25 (11.9) | 62.29 [40.31, 91.95] | 152.45 [84.92, 243.60] |

Notes: Age-standardised rates are expressed per 100,000 people and standardised to the age distribution of the observed study population identifying with an LGB+ orientation in Census 2021. Cells with the value '-' are not applicable for the given characteristic or category. Sexual orientation groups Gay or Lesbian, Bisexual, and Other are not presented for subcategories where suppression was applied for the purposes of statistical disclosure control. All calculations are based on estimates rounded to the nearest multiple of five for disclosure control reasons. CI: confidence interval.

**Supplementary Table 13: Sensitivity analysis including proxy responses to the sexual orientation question in Census 2021, ratios of age-specific and age-standardised rates of self-harm and suicide per 100,000 people**

| Characteristic | Category | Sexual orientation | Rate ratio denominator | Rate ratio for self-harm [95% CI] | Rate ratio for suicide [95% CI] |
|---|---|---|---|---|---|
| Total | Total | LGB+ | Heterosexual | 2.56 [2.53, 2.60] | 2.08 [1.92, 2.24] |
| | | Gay or Lesbian | Heterosexual | 2.48 [2.42, 2.53] | 2.03 [1.81, 2.25] |
| | | Bisexual | Heterosexual | 2.82 [2.77, 2.88] | 2.18 [1.90, 2.45] |
| | | Other | Heterosexual | 2.21 [2.12, 2.30] | 2.12 [1.63, 2.61] |
| Age on Census Day (10-year bands) | 16 to 24 years | LGB+ | Heterosexual | 2.76 [2.71, 2.82] | 2.03 [1.69, 2.38] |
| | | Gay or Lesbian | Heterosexual | 2.78 [2.69, 2.87] | c |
| | | Bisexual | Heterosexual | 2.83 [2.76, 2.90] | c |
| | | Other | Heterosexual | 2.38 [2.25, 2.52] | c |
| | 25 to 34 years | LGB+ | Heterosexual | 2.47 [2.40, 2.53] | 1.92 [1.64, 2.22] |
| | | Gay or Lesbian | Heterosexual | 2.32 [2.23, 2.42] | c |
| | | Bisexual | Heterosexual | 2.71 [2.61, 2.82] | c |
| | | Other | Heterosexual | 2.15 [1.98, 2.32] | c |
| | 35 to 44 years | LGB+ | Heterosexual | 2.24 [2.14, 2.33] | 2.10 [1.75, 2.47] |
| | | Gay or Lesbian | Heterosexual | 2.00 [1.89, 2.11] | c |
| | | Bisexual | Heterosexual | 2.80 [2.63, 2.97] | c |
| | | Other | Heterosexual | 1.73 [1.50, 1.97] | c |
| | 45 to 54 years | LGB+ | Heterosexual | 2.15 [2.04, 2.26] | 2.12 [1.74, 2.52] |
| | | Gay or Lesbian | Heterosexual | 1.82 [1.70, 1.95] | c |
| | | Bisexual | Heterosexual | 3.09 [2.83, 3.35] | c |
| | | Other | Heterosexual | 1.97 [1.62, 2.32] | c |
| | 55 to 64 years | LGB+ | Heterosexual | 1.95 [1.79, 2.11] | 2.25 [1.72, 2.79] |
| | | Gay or Lesbian | Heterosexual | 1.67 [1.50, 1.84] | c |
| | | Bisexual | Heterosexual | 2.80 [2.41, 3.20] | c |
| | | Other | Heterosexual | 2.00 [1.41, 2.58] | c |
| | 65 years and over | LGB+ | Heterosexual | 1.67 [1.46, 1.88] | 2.65 [1.77, 3.55] |
| | | Gay or Lesbian | Heterosexual | 1.57 [1.33, 1.82] | c |
| | | Bisexual | Heterosexual | 1.92 [1.47, 2.37] | c |
| | | Other | Heterosexual | 1.30 [0.65, 1.96] | c |
| Sex | Female | LGB+ | Heterosexual | 2.78 [2.74, 2.83] | 3.17 [2.78, 3.57] |
| | | Gay or Lesbian | Heterosexual | 3.05 [2.96, 3.14] | 2.94 [2.35, 3.56] |
| | | Bisexual | Heterosexual | 2.85 [2.79, 2.91] | 3.31 [2.72, 3.91] |
| | | Other | Heterosexual | 2.33 [2.22, 2.44] | 3.07 [2.02, 4.13] |
| | Male | LGB+ | Heterosexual | 1.93 [1.88, 1.98] | 1.70 [1.53, 1.88] |
| | | Gay or Lesbian | Heterosexual | 1.89 [1.82, 1.95] | 1.58 [1.37, 1.79] |
| | | Bisexual | Heterosexual | 2.18 [2.08, 2.27] | 1.99 [1.62, 2.36] |
| | | Other | Heterosexual | 1.68 [1.53, 1.82] | 1.99 [1.37, 2.62] |
| Region | England: North East | LGB+ | Heterosexual | 2.20 [2.08, 2.33] | 1.90 [1.30, 2.54] |
| | | Gay or Lesbian | Heterosexual | 2.17 [1.98, 2.35] | c |
| | | Bisexual | Heterosexual | 2.39 [2.17, 2.61] | c |
| | | Other | Heterosexual | 2.13 [1.73, 2.54] | c |
| | England: North West | LGB+ | Heterosexual | 2.73 [2.63, 2.84] | 2.42 [1.97, 2.88] |
| | | Gay or Lesbian | Heterosexual | 2.75 [2.59, 2.90] | c |
| | | Bisexual | Heterosexual | 3.03 [2.86, 3.20] | c |
| | | Other | Heterosexual | 2.07 [1.80, 2.35] | c |
| | England: Yorkshire and The Humber | LGB+ | Heterosexual | 2.25 [2.15, 2.34] | 1.95 [1.47, 2.45] |
| | | Gay or Lesbian | Heterosexual | 2.19 [2.05, 2.33] | c |
| | | Bisexual | Heterosexual | 2.42 [2.28, 2.57] | c |
| | | Other | Heterosexual | 2.02 [1.77, 2.28] | c |
| | England: East Midlands | LGB+ | Heterosexual | 2.65 [2.54, 2.77] | 2.70 [2.05, 3.40] |
| | | Gay or Lesbian | Heterosexual | 2.64 [2.45, 2.83] | c |
| | | Bisexual | Heterosexual | 2.90 [2.72, 3.09] | c |
| | | Other | Heterosexual | 2.08 [1.78, 2.38] | c |
| | England: West Midlands | LGB+ | Heterosexual | 2.92 [2.78, 3.07] | 1.74 [1.24, 2.26] |
| | | Gay or Lesbian | Heterosexual | 2.86 [2.64, 3.07] | c |
| | | Bisexual | Heterosexual | 3.24 [3.02, 3.46] | c |
| | | Other | Heterosexual | 2.15 [1.81, 2.49] | c |
| | England: East of England | LGB+ | Heterosexual | 2.74 [2.63, 2.86] | 2.37 [1.77, 2.99] |
| | | Gay or Lesbian | Heterosexual | 2.56 [2.38, 2.74] | c |
| | | Bisexual | Heterosexual | 3.07 [2.89, 3.25] | c |
| | | Other | Heterosexual | 2.38 [2.08, 2.68] | c |
| | England: London | LGB+ | Heterosexual | 2.36 [2.26, 2.46] | 2.27 [1.79, 2.77] |
| | | Gay or Lesbian | Heterosexual | 2.26 [2.08, 2.43] | c |
| | | Bisexual | Heterosexual | 2.58 [2.43, 2.73] | c |
| | | Other | Heterosexual | 2.32 [2.06, 2.58] | c |

| | | | | | |
|---|---|---|---|---|---|
| | England: South East | LGB+ | Heterosexual | 2.66 [2.57, 2.76] | 1.93 [1.54, 2.34] |
| | | Gay or Lesbian | Heterosexual | 2.50 [2.36, 2.64] | c |
| | | Bisexual | Heterosexual | 2.93 [2.79, 3.06] | c |
| | | Other | Heterosexual | 2.41 [2.18, 2.65] | c |
| | England: South West | LGB+ | Heterosexual | 2.72 [2.61, 2.83] | 1.70 [1.27, 2.16] |
| | | Gay or Lesbian | Heterosexual | 2.67 [2.49, 2.85] | c |
| | | Bisexual | Heterosexual | 2.94 [2.78, 3.11] | c |
| | | Other | Heterosexual | 2.50 [2.21, 2.79] | c |
| | Wales | LGB+ | Heterosexual | 2.46 [2.30, 2.62] | 2.20 [1.52, 2.94] |
| | | Gay or Lesbian | Heterosexual | 2.54 [2.29, 2.80] | c |
| | | Bisexual | Heterosexual | 2.58 [2.33, 2.84] | c |
| | | Other | Heterosexual | 1.86 [1.43, 2.29] | c |
| Ethnic group | Asian, Asian British or Asian Welsh | LGB+ | Heterosexual | 2.32 [2.11, 2.53] | 2.94 [1.64, 4.30] |
| | | Gay or Lesbian | Heterosexual | 2.81 [2.32, 3.30] | c |
| | | Bisexual | Heterosexual | 2.17 [1.91, 2.44] | c |
| | | Other | Heterosexual | 2.20 [1.71, 2.70] | c |
| | Black, Black British, Black Welsh, Caribbean or African | LGB+ | Heterosexual | 3.05 [2.73, 3.37] | 4.14 [2.06, 6.46] |
| | | Gay or Lesbian | Heterosexual | 3.34 [2.70, 3.98] | c |
| | | Bisexual | Heterosexual | 3.07 [2.64, 3.51] | c |
| | | Other | Heterosexual | 2.73 [2.09, 3.38] | c |
| | Mixed or Multiple ethnic groups | LGB+ | Heterosexual | 2.27 [2.13, 2.42] | 2.34 [1.47, 3.30] |
| | | Gay or Lesbian | Heterosexual | 2.07 [1.85, 2.29] | c |
| | | Bisexual | Heterosexual | 2.53 [2.30, 2.75] | c |
| | | Other | Heterosexual | 1.94 [1.60, 2.29] | c |
| | White | LGB+ | Heterosexual | 2.44 [2.41, 2.48] | 1.94 [1.78, 2.10] |
| | | Gay or Lesbian | Heterosexual | 2.32 [2.27, 2.37] | c |
| | | Bisexual | Heterosexual | 2.75 [2.69, 2.81] | c |
| | | Other | Heterosexual | 2.21 [2.11, 2.31] | c |
| | Other ethnic group | LGB+ | Heterosexual | 2.41 [2.06, 2.76] | 2.25 [0.18, 4.62] |
| | | Gay or Lesbian | Heterosexual | 3.20 [2.36, 4.05] | c |
| | | Bisexual | Heterosexual | 2.37 [1.88, 2.87] | c |
| | | Other | Heterosexual | 1.81 [1.29, 2.33] | c |
| Rural/urban classification | Urban | LGB+ | Heterosexual | 2.51 [2.48, 2.55] | 2.10 [1.92, 2.27] |
| | | Gay or Lesbian | Heterosexual | 2.44 [2.38, 2.50] | 2.02 [1.78, 2.26] |
| | | Bisexual | Heterosexual | 2.76 [2.70, 2.82] | 2.18 [1.88, 2.49] |
| | | Other | Heterosexual | 2.17 [2.08, 2.27] | 2.22 [1.69, 2.76] |
| | Rural | LGB+ | Heterosexual | 2.88 [2.77, 2.99] | 2.08 [1.66, 2.52] |
| | | Gay or Lesbian | Heterosexual | 2.72 [2.56, 2.88] | 2.18 [1.56, 2.83] |
| | | Bisexual | Heterosexual | 3.25 [3.08, 3.42] | 2.22 [1.52, 2.94] |
| | | Other | Heterosexual | 2.49 [2.19, 2.79] | 1.59 [0.34, 2.85] |
| Country of birth | Born in the UK | LGB+ | Heterosexual | 2.56 [2.52, 2.60] | 2.10 [1.93, 2.27] |
| | | Gay or Lesbian | Heterosexual | 2.41 [2.36, 2.46] | 2.01 [1.78, 2.24] |
| | | Bisexual | Heterosexual | 2.88 [2.82, 2.94] | 2.18 [1.88, 2.48] |
| | | Other | Heterosexual | 2.38 [2.28, 2.49] | 2.50 [1.88, 3.11] |
| | Born outside the UK | LGB+ | Heterosexual | 2.30 [2.18, 2.42] | 1.95 [1.47, 2.45] |
| | | Gay or Lesbian | Heterosexual | 2.27 [2.04, 2.50] | 1.92 [1.07, 2.79] |
| | | Bisexual | Heterosexual | 2.45 [2.29, 2.62] | 2.32 [1.52, 3.14] |
| | | Other | Heterosexual | 1.99 [1.73, 2.24] | 1.59 [0.53, 2.68] |
| English language proficiency | Main language is English | LGB+ | Heterosexual | 2.55 [2.51, 2.58] | 2.08 [1.92, 2.24] |
| | | Gay or Lesbian | Heterosexual | 2.42 [2.37, 2.47] | 1.99 [1.77, 2.21] |
| | | Bisexual | Heterosexual | 2.84 [2.78, 2.89] | 2.21 [1.93, 2.50] |
| | | Other | Heterosexual | 2.31 [2.21, 2.41] | 2.32 [1.76, 2.88] |
| | Main language is not English | LGB+ | Heterosexual | 2.03 [1.85, 2.22] | 1.58 [0.85, 2.35] |
| | | Gay or Lesbian | Heterosexual | 2.22 [1.80, 2.64] | 1.39 [0.34, 2.47] |
| | | Bisexual | Heterosexual | 2.05 [1.80, 2.30] | 1.41 [0.36, 2.51] |
| | | Other | Heterosexual | 1.86 [1.52, 2.21] | 2.07 [0.19, 3.98] |
| Religion | No religion | LGB+ | Heterosexual | 2.14 [2.11, 2.18] | 1.84 [1.66, 2.02] |
| | | Gay or Lesbian | Heterosexual | 2.06 [2.01, 2.12] | c |
| | | Bisexual | Heterosexual | 2.38 [2.32, 2.43] | c |
| | | Other | Heterosexual | 1.96 [1.85, 2.07] | c |
| | Christian | LGB+ | Heterosexual | 2.98 [2.90, 3.07] | 2.15 [1.80, 2.51] |
| | | Gay or Lesbian | Heterosexual | 2.81 [2.68, 2.94] | c |
| | | Bisexual | Heterosexual | 3.52 [3.37, 3.66] | c |
| | | Other | Heterosexual | 2.18 [1.96, 2.40] | c |
| | Buddhist | LGB+ | Heterosexual | 4.14 [3.49, 4.86] | 2.22 [0.81, 4.16] |
| | | Gay or Lesbian | Heterosexual | 3.61 [2.60, 4.70] | c |
| | | Bisexual | Heterosexual | 4.12 [3.30, 5.02] | c |
| | | Other | Heterosexual | 5.04 [3.48, 6.67] | c |
| | Hindu | LGB+ | Heterosexual | 2.36 [1.78, 2.95] | 2.62 [0.00, 5.73] |
| | | Gay or Lesbian | Heterosexual | 3.47 [1.57, 5.38] | c |
| | | Bisexual | Heterosexual | 2.35 [1.66, 3.05] | c |

| | | | | | |
|---|---|---|---|---|---|
| | | Other | Heterosexual | 1.63 [0.43, 2.85] | c |
| | Jewish | LGB+ | Heterosexual | 3.15 [2.52, 3.85] | 2.91 [0.31, 6.35] |
| | | Gay or Lesbian | Heterosexual | 2.30 [1.47, 3.18] | c |
| | | Bisexual | Heterosexual | 4.04 [3.00, 5.17] | c |
| | | Other | Heterosexual | 2.71 [1.12, 4.37] | c |
| | Muslim | LGB+ | Heterosexual | 2.40 [2.08, 2.72] | 4.33 [0.89, 8.05] |
| | | Gay or Lesbian | Heterosexual | 3.76 [2.75, 4.77] | c |
| | | Bisexual | Heterosexual | 2.41 [1.98, 2.85] | c |
| | | Other | Heterosexual | 1.74 [1.26, 2.21] | c |
| | Sikh | LGB+ | Heterosexual | 2.35 [1.61, 3.12] | 4.20 [0.10, 8.80] |
| | | Gay or Lesbian | Heterosexual | 5.91 [3.04, 8.83] | c |
| | | Bisexual | Heterosexual | 1.21 [0.51, 1.92] | c |
| | | Other | Heterosexual | 2.38 [0.82, 3.95] | c |
| | Other religion | LGB+ | Heterosexual | 2.70 [2.46, 2.97] | 1.11 [0.61, 1.78] |
| | | Gay or Lesbian | Heterosexual | 2.85 [2.41, 3.31] | c |
| | | Bisexual | Heterosexual | 2.75 [2.45, 3.07] | c |
| | | Other | Heterosexual | 2.50 [2.14, 2.89] | c |
| | Not answered | LGB+ | Heterosexual | 2.79 [2.57, 3.00] | 1.71 [0.99, 2.48] |
| | | Gay or Lesbian | Heterosexual | 2.51 [2.19, 2.83] | c |
| | | Bisexual | Heterosexual | 3.38 [3.02, 3.75] | c |
| | | Other | Heterosexual | 2.05 [1.58, 2.53] | c |
| IMD/WIMD decile group | 1 (most deprived) | LGB+ | Heterosexual | 2.45 [2.36, 2.53] | 2.25 [1.80, 2.72] |
| | | Gay or Lesbian | Heterosexual | 2.53 [2.39, 2.66] | 2.15 [1.53, 2.80] |
| | | Bisexual | Heterosexual | 2.66 [2.52, 2.80] | 2.59 [1.80, 3.41] |
| | | Other | Heterosexual | 1.67 [1.47, 1.87] | 1.58 [0.43, 2.75] |
| | 2 | LGB+ | Heterosexual | 2.38 [2.29, 2.47] | 1.94 [1.50, 2.40] |
| | | Gay or Lesbian | Heterosexual | 2.36 [2.23, 2.50] | 1.99 [1.34, 2.66] |
| | | Bisexual | Heterosexual | 2.53 [2.39, 2.66] | 1.98 [1.26, 2.73] |
| | | Other | Heterosexual | 2.00 [1.78, 2.23] | 1.63 [0.52, 2.77] |
| | 3 | LGB+ | Heterosexual | 2.40 [2.31, 2.50] | 2.12 [1.67, 2.59] |
| | | Gay or Lesbian | Heterosexual | 2.45 [2.30, 2.60] | 2.06 [1.42, 2.73] |
| | | Bisexual | Heterosexual | 2.58 [2.43, 2.72] | 1.86 [1.20, 2.53] |
| | | Other | Heterosexual | 1.85 [1.62, 2.07] | 2.65 [1.22, 4.12] |
| | 4 | LGB+ | Heterosexual | 2.45 [2.35, 2.55] | 1.80 [1.38, 2.23] |
| | | Gay or Lesbian | Heterosexual | 2.46 [2.30, 2.62] | 1.80 [1.23, 2.40] |
| | | Bisexual | Heterosexual | 2.67 [2.52, 2.83] | 1.70 [1.02, 2.40] |
| | | Other | Heterosexual | 2.13 [1.88, 2.39] | 1.67 [0.46, 2.90] |
| | 5 | LGB+ | Heterosexual | 2.54 [2.44, 2.66] | 1.83 [1.39, 2.29] |
| | | Gay or Lesbian | Heterosexual | 2.48 [2.31, 2.65] | 1.75 [1.19, 2.33] |
| | | Bisexual | Heterosexual | 2.67 [2.51, 2.83] | 1.48 [0.84, 2.14] |
| | | Other | Heterosexual | 2.41 [2.11, 2.71] | 2.07 [0.74, 3.43] |
| | 6 | LGB+ | Heterosexual | 2.55 [2.44, 2.66] | 2.08 [1.59, 2.60] |
| | | Gay or Lesbian | Heterosexual | 2.31 [2.14, 2.48] | 2.03 [1.36, 2.71] |
| | | Bisexual | Heterosexual | 2.99 [2.80, 3.17] | 2.29 [1.38, 3.23] |
| | | Other | Heterosexual | 2.17 [1.87, 2.47] | 1.77 [0.37, 3.18] |
| | 7 | LGB+ | Heterosexual | 2.62 [2.50, 2.75] | 2.18 [1.64, 2.75] |
| | | Gay or Lesbian | Heterosexual | 2.29 [2.11, 2.48] | 1.87 [1.17, 2.60] |
| | | Bisexual | Heterosexual | 3.04 [2.84, 3.24] | 2.70 [1.63, 3.81] |
| | | Other | Heterosexual | 2.53 [2.17, 2.89] | 3.44 [1.20, 5.72] |
| | 8 | LGB+ | Heterosexual | 2.72 [2.58, 2.85] | 2.09 [1.53, 2.67] |
| | | Gay or Lesbian | Heterosexual | 2.48 [2.28, 2.68] | 1.87 [1.13, 2.62] |
| | | Bisexual | Heterosexual | 3.07 [2.86, 3.28] | 2.35 [1.38, 3.36] |
| | | Other | Heterosexual | 2.52 [2.14, 2.90] | 1.93 [0.03, 3.84] |
| | 9 | LGB+ | Heterosexual | 2.80 [2.65, 2.95] | 2.14 [1.54, 2.78] |
| | | Gay or Lesbian | Heterosexual | 2.59 [2.37, 2.82] | 2.50 [1.53, 3.50] |
| | | Bisexual | Heterosexual | 3.17 [2.93, 3.42] | 2.23 [1.14, 3.35] |
| | | Other | Heterosexual | 2.90 [2.45, 3.35] | 1.45 [0.00, 3.02] |
| | 10 (least deprived) | LGB+ | Heterosexual | 2.85 [2.68, 3.03] | 2.35 [1.63, 3.12] |
| | | Gay or Lesbian | Heterosexual | 2.53 [2.28, 2.78] | 2.34 [1.26, 3.47] |
| | | Bisexual | Heterosexual | 3.21 [2.95, 3.49] | 2.56 [1.25, 3.91] |
| | | Other | Heterosexual | 2.86 [2.37, 3.35] | 3.92 [0.76, 7.14] |
| Highest level of qualification (≥25 years) | No qualifications | LGB+ | Heterosexual | 2.12 [2.00, 2.23] | 1.58 [1.16, 2.02] |
| | | Gay or Lesbian | Heterosexual | 2.89 [2.67, 3.11] | c |
| | | Bisexual | Heterosexual | 2.35 [2.15, 2.54] | c |
| | | Other | Heterosexual | 0.66 [0.54, 0.79] | c |
| | Level 1 (one to four GCSE passes (grade A* to C or grade 4 and above) and any other GCSEs at other grades, or | LGB+ | Heterosexual | 2.54 [2.39, 2.70] | 1.87 [1.26, 2.50] |
| | | Gay or Lesbian | Heterosexual | 2.35 [2.15, 2.56] | c |
| | | Bisexual | Heterosexual | 3.03 [2.77, 3.29] | c |
| | | Other | Heterosexual | 1.78 [1.38, 2.17] | c |

| | | | | | |
|---|---|---|---|---|---|
| | equivalent qualifications) | | | | |
| | Level 2 (five or more GCSE passes (grade A* to C or grade 4 and above) or equivalent qualifications) | LGB+ | Heterosexual | 2.60 [2.47, 2.73] | 1.75 [1.31, 2.22] |
| | | Gay or Lesbian | Heterosexual | 2.22 [2.06, 2.38] | c |
| | | Bisexual | Heterosexual | 3.19 [2.97, 3.42] | c |
| | | Other | Heterosexual | 2.40 [2.00, 2.80] | c |
| | Apprenticeship | LGB+ | Heterosexual | 2.36 [2.11, 2.62] | 1.01 [0.42, 1.62] |
| | | Gay or Lesbian | Heterosexual | 2.28 [1.91, 2.65] | c |
| | | Bisexual | Heterosexual | 2.90 [2.46, 3.35] | c |
| | | Other | Heterosexual | 1.30 [0.81, 1.80] | c |
| | Level 3 (two or more A Levels or equivalent qualifications) | LGB+ | Heterosexual | 2.99 [2.86, 3.12] | 2.51 [2.03, 3.00] |
| | | Gay or Lesbian | Heterosexual | 2.65 [2.49, 2.80] | c |
| | | Bisexual | Heterosexual | 3.55 [3.32, 3.79] | c |
| | | Other | Heterosexual | 3.25 [2.82, 3.69] | c |
| | Level 4 and above (Higher National Certificate, Higher National Diploma, Bachelor's degree, or post-graduate qualifications) | LGB+ | Heterosexual | 2.45 [2.36, 2.53] | 2.57 [2.24, 2.91] |
| | | Gay or Lesbian | Heterosexual | 2.16 [2.05, 2.27] | c |
| | | Bisexual | Heterosexual | 2.87 [2.71, 3.03] | c |
| | | Other | Heterosexual | 2.81 [2.52, 3.09] | c |
| | Other qualifications | LGB+ | Heterosexual | 2.14 [1.81, 2.48] | 2.12 [0.76, 3.59] |
| | | Gay or Lesbian | Heterosexual | 2.38 [1.85, 2.92] | c |
| | | Bisexual | Heterosexual | 2.33 [1.79, 2.88] | c |
| | | Other | Heterosexual | 1.29 [0.70, 1.89] | c |
| NS-SEC | Class 1 (Higher managerial, administrative and professional occupations) | LGB+ | Heterosexual | 3.24 [2.52, 3.99] | 2.11 [1.53, 2.78] |
| | | Gay or Lesbian | Heterosexual | 2.84 [1.74, 3.97] | c |
| | | Bisexual | Heterosexual | 3.78 [2.72, 4.88] | c |
| | | Other | Heterosexual | 2.90 [1.19, 4.64] | c |
| | Class 2 (Lower managerial, administrative and professional occupations) | LGB+ | Heterosexual | 2.96 [2.69, 3.24] | 2.57 [1.72, 3.47] |
| | | Gay or Lesbian | Heterosexual | 2.66 [2.25, 3.08] | c |
| | | Bisexual | Heterosexual | 3.33 [2.94, 3.71] | c |
| | | Other | Heterosexual | 3.04 [2.23, 3.86] | c |
| | Class 3 (Intermediate occupations) | LGB+ | Heterosexual | 2.89 [2.71, 3.06] | 2.68 [2.03, 3.38] |
| | | Gay or Lesbian | Heterosexual | 2.61 [2.36, 2.87] | c |
| | | Bisexual | Heterosexual | 3.28 [3.01, 3.55] | c |
| | | Other | Heterosexual | 2.51 [1.95, 3.08] | c |
| | Class 4 (Small employers and own account workers) | LGB+ | Heterosexual | 2.05 [1.77, 2.33] | 1.13 [0.73, 1.53] |
| | | Gay or Lesbian | Heterosexual | 1.99 [1.47, 2.51] | c |
| | | Bisexual | Heterosexual | 2.30 [1.90, 2.71] | c |
| | | Other | Heterosexual | 1.50 [0.96, 2.04] | c |
| | Class 5 (Lower supervisory and technical occupations) | LGB+ | Heterosexual | 2.91 [2.62, 3.21] | 1.58 [1.02, 2.18] |
| | | Gay or Lesbian | Heterosexual | 2.88 [2.44, 3.31] | c |
| | | Bisexual | Heterosexual | 3.14 [2.71, 3.57] | c |
| | | Other | Heterosexual | 2.53 [1.38, 3.69] | c |
| | Class 6 (Semi-routine occupations) | LGB+ | Heterosexual | 2.64 [2.54, 2.75] | 2.73 [2.16, 3.33] |
| | | Gay or Lesbian | Heterosexual | 2.55 [2.40, 2.70] | c |
| | | Bisexual | Heterosexual | 2.83 [2.68, 2.98] | c |
| | | Other | Heterosexual | 2.32 [2.02, 2.62] | c |
| | Class 7 (Routine occupations) | LGB+ | Heterosexual | 2.47 [2.37, 2.58] | 2.00 [1.58, 2.45] |
| | | Gay or Lesbian | Heterosexual | 2.51 [2.35, 2.67] | c |
| | | Bisexual | Heterosexual | 2.64 [2.49, 2.79] | c |
| | | Other | Heterosexual | 1.77 [1.51, 2.02] | c |
| | Class 8 (Never worked and long-term unemployed) | LGB+ | Heterosexual | 2.45 [2.36, 2.53] | 2.01 [1.56, 2.49] |
| | | Gay or Lesbian | Heterosexual | 2.79 [2.64, 2.95] | c |
| | | Bisexual | Heterosexual | 2.64 [2.51, 2.77] | c |
| | | Other | Heterosexual | 1.37 [1.22, 1.52] | c |
| | Full-time students | LGB+ | Heterosexual | 2.69 [2.47, 2.93] | 3.65 [0.62, 7.03] |
| | | Gay or Lesbian | Heterosexual | 2.89 [2.50, 3.28] | c |
| | | Bisexual | Heterosexual | 2.50 [2.28, 2.72] | c |
| | | Other | Heterosexual | 2.52 [2.03, 3.02] | c |
| Household NS-SEC | Class 1 (Higher managerial, administrative and professional occupations) | LGB+ | Heterosexual | 2.75 [2.65, 2.86] | 2.14 [1.76, 2.54] |
| | | Gay or Lesbian | Heterosexual | 2.58 [2.41, 2.74] | c |
| | | Bisexual | Heterosexual | 3.03 [2.87, 3.18] | c |
| | | Other | Heterosexual | 2.79 [2.49, 3.09] | c |
| | Class 2 (Lower managerial, administrative and | LGB+ | Heterosexual | 2.62 [2.54, 2.69] | 2.16 [1.83, 2.50] |
| | | Gay or Lesbian | Heterosexual | 2.46 [2.35, 2.58] | c |
| | | Bisexual | Heterosexual | 2.84 [2.72, 2.96] | c |

| | | | | | |
|---|---|---|---|---|---|
| | professional occupations) | Other | Heterosexual | 2.62 [2.39, 2.85] | c |
| | Class 3 (Intermediate occupations) | LGB+ | Heterosexual | 2.78 [2.67, 2.89] | 2.25 [1.72, 2.79] |
| | | Gay or Lesbian | Heterosexual | 2.80 [2.63, 2.96] | c |
| | | Bisexual | Heterosexual | 2.89 [2.72, 3.06] | c |
| | | Other | Heterosexual | 2.32 [2.03, 2.62] | c |
| | Class 4 (Small employers and own account workers) | LGB+ | Heterosexual | 2.32 [2.19, 2.45] | 1.48 [1.02, 1.95] |
| | | Gay or Lesbian | Heterosexual | 2.35 [2.13, 2.57] | c |
| | | Bisexual | Heterosexual | 2.53 [2.33, 2.72] | c |
| | | Other | Heterosexual | 1.73 [1.46, 2.00] | c |
| | Class 5 (Lower supervisory and technical occupations) | LGB+ | Heterosexual | 2.56 [2.40, 2.72] | 1.85 [1.18, 2.56] |
| | | Gay or Lesbian | Heterosexual | 2.64 [2.39, 2.89] | c |
| | | Bisexual | Heterosexual | 2.79 [2.55, 3.04] | c |
| | | Other | Heterosexual | 1.52 [1.17, 1.87] | c |
| | Class 6 (Semi-routine occupations) | LGB+ | Heterosexual | 2.56 [2.47, 2.66] | 2.53 [1.97, 3.12] |
| | | Gay or Lesbian | Heterosexual | 2.71 [2.55, 2.86] | c |
| | | Bisexual | Heterosexual | 2.59 [2.46, 2.73] | c |
| | | Other | Heterosexual | 2.10 [1.85, 2.34] | c |
| | Class 7 (Routine occupations) | LGB+ | Heterosexual | 2.43 [2.32, 2.55] | 2.13 [1.59, 2.70] |
| | | Gay or Lesbian | Heterosexual | 2.59 [2.39, 2.78] | c |
| | | Bisexual | Heterosexual | 2.60 [2.44, 2.76] | c |
| | | Other | Heterosexual | 1.54 [1.32, 1.77] | c |
| | Class 8 (Never worked and long-term unemployed) | LGB+ | Heterosexual | 2.27 [2.17, 2.38] | 1.80 [1.24, 2.39] |
| | | Gay or Lesbian | Heterosexual | 2.68 [2.49, 2.88] | c |
| | | Bisexual | Heterosexual | 2.41 [2.27, 2.57] | c |
| | | Other | Heterosexual | 1.20 [1.02, 1.38] | c |
| | Full-time students | LGB+ | Heterosexual | 2.54 [2.18, 2.90] | 3.37 [0.00, 8.58] |
| | | Gay or Lesbian | Heterosexual | 2.87 [2.24, 3.51] | c |
| | | Bisexual | Heterosexual | 2.28 [1.94, 2.62] | c |
| | | Other | Heterosexual | 2.38 [1.49, 3.27] | c |
| | Living in a communal establishment | LGB+ | Heterosexual | 1.96 [1.79, 2.14] | 1.71 [0.86, 2.67] |
| | | Gay or Lesbian | Heterosexual | 1.78 [1.54, 2.02] | c |
| | | Bisexual | Heterosexual | 2.39 [2.10, 2.68] | c |
| | | Other | Heterosexual | 1.05 [0.75, 1.36] | c |
| Lone person household indicator | Lives alone | LGB+ | Heterosexual | 2.04 [1.94, 2.15] | 1.77 [1.39, 2.17] |
| | | Gay or Lesbian | Heterosexual | 2.10 [1.92, 2.28] | c |
| | | Bisexual | Heterosexual | 2.19 [2.06, 2.32] | c |
| | | Other | Heterosexual | 1.69 [1.45, 1.92] | c |
| | Does not live alone | LGB+ | Heterosexual | 2.46 [2.42, 2.50] | 1.87 [1.69, 2.07] |
| | | Gay or Lesbian | Heterosexual | 2.40 [2.34, 2.46] | c |
| | | Bisexual | Heterosexual | 2.68 [2.62, 2.74] | c |
| | | Other | Heterosexual | 2.09 [1.99, 2.19] | c |
| | Living in a communal establishment | LGB+ | Heterosexual | 1.96 [1.79, 2.14] | 1.71 [0.85, 2.66] |
| | | Gay or Lesbian | Heterosexual | 1.78 [1.54, 2.02] | c |
| | | Bisexual | Heterosexual | 2.39 [2.10, 2.68] | c |
| | | Other | Heterosexual | 1.05 [0.75, 1.36] | c |
| Household size | One person in household | LGB+ | Heterosexual | 2.04 [1.94, 2.15] | 1.77 [1.39, 2.18] |
| | | Gay or Lesbian | Heterosexual | 2.10 [1.92, 2.28] | c |
| | | Bisexual | Heterosexual | 2.19 [2.06, 2.32] | c |
| | | Other | Heterosexual | 1.69 [1.45, 1.93] | c |
| | Two people in household | LGB+ | Heterosexual | 2.15 [2.08, 2.22] | 1.43 [1.21, 1.67] |
| | | Gay or Lesbian | Heterosexual | 2.09 [1.97, 2.20] | c |
| | | Bisexual | Heterosexual | 2.38 [2.28, 2.48] | c |
| | | Other | Heterosexual | 2.14 [1.94, 2.34] | c |
| | Three to five people in household | LGB+ | Heterosexual | 2.56 [2.50, 2.62] | 2.10 [1.75, 2.45] |
| | | Gay or Lesbian | Heterosexual | 2.58 [2.48, 2.67] | c |
| | | Bisexual | Heterosexual | 2.72 [2.63, 2.81] | c |
| | | Other | Heterosexual | 1.98 [1.84, 2.11] | c |
| | Six or more people in household | LGB+ | Heterosexual | 2.52 [2.34, 2.69] | 1.73 [0.70, 2.81] |
| | | Gay or Lesbian | Heterosexual | 2.87 [2.51, 3.24] | c |
| | | Bisexual | Heterosexual | 2.64 [2.40, 2.89] | c |
| | | Other | Heterosexual | 1.79 [1.46, 2.11] | c |
| | Living in a communal establishment | LGB+ | Heterosexual | 1.96 [1.79, 2.14] | 1.71 [0.86, 2.65] |
| | | Gay or Lesbian | Heterosexual | 1.78 [1.54, 2.02] | c |
| | | Bisexual | Heterosexual | 2.39 [2.10, 2.68] | c |
| | | Other | Heterosexual | 1.05 [0.75, 1.36] | c |
| Household tenure | Owned or shared ownership | LGB+ | Heterosexual | 2.55 [2.48, 2.61] | 2.03 [1.77, 2.29] |
| | | Gay or Lesbian | Heterosexual | 2.45 [2.35, 2.55] | c |
| | | Bisexual | Heterosexual | 2.73 [2.62, 2.83] | c |
| | | Other | Heterosexual | 2.51 [2.32, 2.70] | c |
| | Social rented | LGB+ | Heterosexual | 2.58 [2.52, 2.65] | 2.18 [1.84, 2.53] |

| | | | | | |
|---|---|---|---|---|---|
| | | Gay or Lesbian | Heterosexual | 2.57 [2.47, 2.66] | c |
| | | Bisexual | Heterosexual | 2.78 [2.69, 2.88] | c |
| | | Other | Heterosexual | 1.96 [1.80, 2.11] | c |
| | Private rented or living rent free | LGB+ | Heterosexual | 2.37 [2.31, 2.43] | 1.97 [1.69, 2.26] |
| | | Gay or Lesbian | Heterosexual | 2.43 [2.33, 2.53] | c |
| | | Bisexual | Heterosexual | 2.46 [2.37, 2.55] | c |
| | | Other | Heterosexual | 1.89 [1.75, 2.03] | c |
| | Living in a communal establishment | LGB+ | Heterosexual | 1.96 [1.79, 2.14] | 1.71 [0.85, 2.66] |
| | | Gay or Lesbian | Heterosexual | 1.78 [1.54, 2.02] | c |
| | | Bisexual | Heterosexual | 2.39 [2.10, 2.68] | c |
| | | Other | Heterosexual | 1.05 [0.75, 1.36] | c |
| Economic activity | Economically active | LGB+ | Heterosexual | 2.43 [2.38, 2.48] | 2.03 [1.81, 2.25] |
| | | Gay or Lesbian | Heterosexual | 2.33 [2.25, 2.41] | 1.98 [1.67, 2.29] |
| | | Bisexual | Heterosexual | 2.58 [2.50, 2.65] | 2.25 [1.86, 2.65] |
| | | Other | Heterosexual | 2.28 [2.13, 2.43] | 2.21 [1.51, 2.91] |
| | Economically inactive | LGB+ | Heterosexual | 2.53 [2.48, 2.59] | 1.98 [1.71, 2.24] |
| | | Gay or Lesbian | Heterosexual | 2.83 [2.73, 2.92] | 2.49 [2.00, 2.98] |
| | | Bisexual | Heterosexual | 2.65 [2.57, 2.73] | 1.72 [1.36, 2.09] |
| | | Other | Heterosexual | 1.72 [1.62, 1.83] | 1.37 [0.86, 1.91] |
| General health (detailed) | Very good health | LGB+ | Heterosexual | 2.17 [2.10, 2.23] | 1.98 [1.66, 2.30] |
| | | Gay or Lesbian | Heterosexual | 2.25 [2.16, 2.35] | 2.05 [1.63, 2.47] |
| | | Bisexual | Heterosexual | 2.27 [2.17, 2.38] | 2.19 [1.53, 2.85] |
| | | Other | Heterosexual | 1.59 [1.42, 1.77] | 1.59 [0.64, 2.54] |
| | Good health | LGB+ | Heterosexual | 2.01 [1.97, 2.06] | 1.73 [1.50, 1.97] |
| | | Gay or Lesbian | Heterosexual | 2.15 [2.07, 2.23] | 1.95 [1.59, 2.31] |
| | | Bisexual | Heterosexual | 2.07 [2.00, 2.14] | 1.64 [1.26, 2.03] |
| | | Other | Heterosexual | 1.51 [1.40, 1.63] | 1.55 [0.86, 2.24] |
| | Fair health | LGB+ | Heterosexual | 1.79 [1.74, 1.84] | 1.61 [1.36, 1.88] |
| | | Gay or Lesbian | Heterosexual | 2.00 [1.91, 2.09] | 1.57 [1.17, 1.98] |
| | | Bisexual | Heterosexual | 1.81 [1.75, 1.88] | 1.62 [1.25, 2.00] |
| | | Other | Heterosexual | 1.36 [1.26, 1.46] | 1.71 [1.02, 2.41] |
| | Bad health | LGB+ | Heterosexual | 1.65 [1.59, 1.73] | 1.50 [1.16, 1.87] |
| | | Gay or Lesbian | Heterosexual | 1.80 [1.68, 1.93] | 1.27 [0.79, 1.77] |
| | | Bisexual | Heterosexual | 1.72 [1.62, 1.81] | 1.50 [1.03, 2.02] |
| | | Other | Heterosexual | 1.28 [1.15, 1.42] | 1.59 [0.78, 2.44] |
| | Very bad health | LGB+ | Heterosexual | 1.95 [1.80, 2.12] | 1.67 [0.99, 2.45] |
| | | Gay or Lesbian | Heterosexual | 2.08 [1.81, 2.37] | 1.46 [0.59, 2.42] |
| | | Bisexual | Heterosexual | 2.09 [1.87, 2.32] | 1.93 [0.83, 3.15] |
| | | Other | Heterosexual | 1.50 [1.22, 1.77] | 1.15 [0.00, 2.51] |
| General health (aggregated) | Good health | LGB+ | Heterosexual | 2.29 [2.25, 2.33] | 1.92 [1.73, 2.12] |
| | | Gay or Lesbian | Heterosexual | 2.33 [2.26, 2.40] | 2.05 [1.77, 2.34] |
| | | Bisexual | Heterosexual | 2.43 [2.36, 2.49] | 1.95 [1.60, 2.31] |
| | | Other | Heterosexual | 1.76 [1.65, 1.86] | 1.70 [1.09, 2.31] |
| | Not good health | LGB+ | Heterosexual | 1.80 [1.76, 1.84] | 1.61 [1.41, 1.81] |
| | | Gay or Lesbian | Heterosexual | 1.99 [1.91, 2.06] | 1.52 [1.21, 1.83] |
| | | Bisexual | Heterosexual | 1.84 [1.78, 1.89] | 1.60 [1.32, 1.88] |
| | | Other | Heterosexual | 1.42 [1.34, 1.50] | 1.64 [1.15, 2.15] |
| Disability | Day-to-day activities limited a lot | LGB+ | Heterosexual | 1.90 [1.85, 1.95] | 1.61 [1.35, 1.88] |
| | | Gay or Lesbian | Heterosexual | 2.04 [1.95, 2.13] | 1.60 [1.20, 2.01] |
| | | Bisexual | Heterosexual | 1.98 [1.91, 2.05] | 1.62 [1.25, 2.00] |
| | | Other | Heterosexual | 1.45 [1.35, 1.55] | 1.27 [0.71, 1.83] |
| | Day-to-day activities limited a little | LGB+ | Heterosexual | 1.47 [1.43, 1.50] | 1.45 [1.24, 1.67] |
| | | Gay or Lesbian | Heterosexual | 1.68 [1.61, 1.75] | 1.76 [1.38, 2.14] |
| | | Bisexual | Heterosexual | 1.50 [1.45, 1.55] | 1.33 [1.02, 1.65] |
| | | Other | Heterosexual | 1.06 [0.98, 1.14] | 1.48 [0.88, 2.09] |
| | Day-to-day activities not limited | LGB+ | Heterosexual | 2.00 [1.95, 2.04] | 1.77 [1.57, 1.98] |
| | | Gay or Lesbian | Heterosexual | 2.11 [2.04, 2.18] | 1.80 [1.53, 2.08] |
| | | Bisexual | Heterosexual | 2.04 [1.97, 2.11] | 1.82 [1.45, 2.18] |
| | | Other | Heterosexual | 1.46 [1.35, 1.58] | 1.65 [1.02, 2.28] |
| Residence type | Living in a private household | LGB+ | Heterosexual | 2.58 [2.54, 2.61] | 2.08 [1.92, 2.25] |
| | | Gay or Lesbian | Heterosexual | 2.50 [2.44, 2.55] | c |
| | | Bisexual | Heterosexual | 2.83 [2.77, 2.88] | c |
| | | Other | Heterosexual | 2.23 [2.13, 2.32] | c |
| | Living in a communal establishment | LGB+ | Heterosexual | 1.96 [1.79, 2.14] | 1.71 [0.85, 2.65] |
| | | Gay or Lesbian | Heterosexual | 1.78 [1.54, 2.02] | c |
| | | Bisexual | Heterosexual | 2.39 [2.10, 2.68] | c |
| | | Other | Heterosexual | 1.05 [0.75, 1.36] | c |

Notes: Rate ratios were calculated using age-specific rates for the age on Census Day variable, and age-standardised rates for all other variables. Cells with the value 'c' have been suppressed for the purposes of statistical disclosure control. All calculations are based on estimates rounded to the nearest multiple of five for disclosure control reasons. CI: confidence interval.

**Supplementary Table 14: Sensitivity analysis restricting to people living in private households only, crude and age-standardised rates of self-harm per 100,000 people**

| Characteristic | Category | Sexual orientation | Count (%), Individuals | Count (%), Individuals with at least one hospital admission or emergency attendance for intentional self-harm | Crude rate of self-harm [95% CI] | Age-standardised rate of self-harm [95% CI] |
|---|---|---|---|---|---|---|
| Total | Total | Total | 28,339,665 (100.0) | 131,605 (100.0) | 464.38 [461.88, 466.90] | 648.49 [644.45, 652.54] |
| | | Heterosexual | 27,270,785 (96.2) | 115,950 (88.1) | 425.18 [422.74, 427.63] | 583.43 [579.46, 587.39] |
| | | LGB+ | 1,068,880 (3.8) | 15,655 (11.9) | 1,464.62 [1,441.76, 1,487.74] | 1,464.40 [1,441.46, 1,487.34] |
| | | Gay or Lesbian | 515,370 (1.8) | 5,965 (4.5) | 1,157.42 [1,128.23, 1,187.17] | 1,411.07 [1,373.48, 1,448.65] |
| | | Bisexual | 434,500 (1.5) | 8,045 (6.1) | 1,851.55 [1,811.31, 1,892.46] | 1,621.89 [1,584.74, 1,659.04] |
| | | Other | 119,005 (0.4) | 1,640 (1.2) | 1,378.09 [1,312.20, 1,446.44] | 1,295.09 [1,231.81, 1,358.38] |
| Age on Census Day (10-year bands) | 16 to 24 years | Total | 2,668,275 (100.0) | 31,900 (100.0) | 1,195.53 [1,182.41, 1,208.65] | - |
| | | Heterosexual | 2,415,345 (90.5) | 24,735 (77.5) | 1,024.08 [1,011.31, 1,036.84] | - |
| | | LGB+ | 252,930 (9.5) | 7,165 (22.5) | 2,832.80 [2,767.21, 2,898.39] | - |
| | | Gay or Lesbian | 71,355 (2.7) | 2,065 (6.5) | 2,893.98 [2,769.16, 3,018.80] | - |
| | | Bisexual | 150,180 (5.6) | 4,315 (13.5) | 2,873.22 [2,787.49, 2,958.95] | - |
| | | Other | 31,390 (1.2) | 785 (2.5) | 2,500.80 [2,325.85, 2,675.74] | - |
| | 25 to 34 years | Total | 5,018,225 (100.0) | 31,060 (100.0) | 618.94 [612.06, 625.83] | - |
| | | Heterosexual | 4,681,300 (93.3) | 26,310 (84.7) | 562.02 [555.23, 568.81] | - |
| | | LGB+ | 336,930 (6.7) | 4,750 (15.3) | 1,409.79 [1,369.70, 1,449.88] | - |
| | | Gay or Lesbian | 148,050 (3.0) | 1,945 (6.3) | 1,313.75 [1,255.36, 1,372.13] | - |
| | | Bisexual | 147,245 (2.9) | 2,285 (7.4) | 1,551.84 [1,488.21, 1,615.46] | - |
| | | Other | 41,635 (0.8) | 525 (1.7) | 1,260.96 [1,153.09, 1,368.82] | - |
| | 35 to 44 years | Total | 4,939,215 (100.0) | 23,000 (100.0) | 465.66 [459.64, 471.68] | - |
| | | Heterosexual | 4,736,350 (95.9) | 20,995 (91.3) | 443.27 [437.28, 449.27] | - |
| | | LGB+ | 202,865 (4.1) | 2,005 (8.7) | 988.34 [945.08, 1,031.60] | - |
| | | Gay or Lesbian | 110,630 (2.2) | 975 (4.2) | 881.32 [826.00, 936.64] | - |
| | | Bisexual | 68,970 (1.4) | 840 (3.7) | 1,217.92 [1,135.56, 1,300.28] | - |
| | | Other | 23,265 (0.5) | 190 (0.8) | 816.68 [700.55, 932.80] | - |
| | 45 to 54 years | Total | 5,078,155 (100.0) | 20,490 (100.0) | 403.49 [397.97, 409.02] | - |
| | | Heterosexual | 4,937,010 (97.2) | 19,355 (94.5) | 392.04 [386.52, 397.56] | - |
| | | LGB+ | 141,145 (2.8) | 1,130 (5.5) | 800.60 [753.92, 847.28] | - |
| | | Gay or Lesbian | 91,630 (1.8) | 610 (3.0) | 665.72 [612.89, 718.55] | - |
| | | Bisexual | 36,830 (0.7) | 425 (2.1) | 1,153.95 [1,044.24, 1,263.66] | - |
| | | Other | 12,685 (0.2) | 95 (0.5) | 748.92 [605.92, 915.51] | - |
| | 55 to 64 years | Total | 4,784,020 (100.0) | 12,745 (100.0) | 266.41 [261.78, 271.03] | - |
| | | Heterosexual | 4,695,320 (98.1) | 12,300 (96.5) | 261.96 [257.33, 266.59] | - |
| | | LGB+ | 88,700 (1.9) | 445 (3.5) | 501.69 [455.08, 548.30] | - |
| | | Gay or Lesbian | 62,205 (1.3) | 270 (2.1) | 434.05 [382.27, 485.82] | - |
| | | Bisexual | 20,005 (0.4) | 135 (1.1) | 674.83 [560.99, 788.67] | - |
| | | Other | 6,490 (0.1) | 35 (0.3) | 539.29 [375.64, 750.02] | - |

| | | | | | | |
|---|---|---|---|---|---|---|
| | 65 years and over | Total | 5,851,775 (100.0) | 12,410 (100.0) | 212.07 [208.34, 215.80] | - |
| | | Heterosexual | 5,805,465 (99.2) | 12,255 (98.8) | 211.09 [207.36, 214.83] | - |
| | | LGB+ | 46,310 (0.8) | 155 (1.2) | 334.70 [282.01, 387.39] | - |
| | | Gay or Lesbian | 31,500 (0.5) | 100 (0.8) | 317.46 [255.24, 379.68] | - |
| | | Bisexual | 11,270 (0.2) | 45 (0.4) | 399.29 [291.25, 534.28] | - |
| | | Other | 3,540 (0.1) | 10 (0.1) | 282.49 [135.46, 519.50] | - |
| Sex | Female | Total | 15,222,250 (100.0) | 78,080 (100.0) | 512.93 [509.34, 516.54] | 732.58 [726.69, 738.48] |
| | | Heterosexual | 14,641,770 (96.2) | 66,840 (85.6) | 456.50 [453.05, 459.98] | 637.08 [631.37, 642.79] |
| | | LGB+ | 580,480 (3.8) | 11,240 (14.4) | 1,936.33 [1,900.70, 1,972.46] | 1,751.12 [1,718.29, 1,783.96] |
| | | Gay or Lesbian | 196,160 (1.3) | 3,320 (4.3) | 1,692.50 [1,635.41, 1,751.07] | 1,924.73 [1,858.03, 1,991.42] |
| | | Bisexual | 309,415 (2.0) | 6,665 (8.5) | 2,154.06 [2,102.66, 2,206.41] | 1,806.79 [1,759.31, 1,854.27] |
| | | Other | 74,905 (0.5) | 1,255 (1.6) | 1,675.46 [1,584.03, 1,770.78] | 1,492.44 [1,407.48, 1,577.41] |
| | Male | Total | 13,117,415 (100.0) | 53,525 (100.0) | 408.05 [404.60, 411.52] | 552.09 [546.62, 557.56] |
| | | Heterosexual | 12,629,015 (96.3) | 49,110 (91.8) | 388.87 [385.43, 392.32] | 525.29 [519.79, 530.79] |
| | | LGB+ | 488,400 (3.7) | 4,415 (8.2) | 903.97 [877.50, 931.04] | 1,000.73 [970.41, 1,031.04] |
| | | Gay or Lesbian | 319,210 (2.4) | 2,645 (4.9) | 828.61 [797.33, 860.80] | 1,001.72 [959.89, 1,043.55] |
| | | Bisexual | 125,085 (1.0) | 1,385 (2.6) | 1,107.25 [1,049.69, 1,167.13] | 1,088.85 [1,031.02, 1,146.68] |
| | | Other | 44,105 (0.3) | 385 (0.7) | 872.92 [787.89, 964.62] | 894.83 [805.09, 984.57] |
| Region | England: North East | Total | 1,314,700 (100.0) | 8,085 (100.0) | 614.97 [601.64, 628.52] | 934.43 [911.21, 957.64] |
| | | Heterosexual | 1,269,590 (96.6) | 7,185 (88.9) | 565.93 [552.92, 579.17] | 860.43 [837.31, 883.55] |
| | | LGB+ | 45,105 (3.4) | 895 (11.1) | 1,984.26 [1,856.37, 2,118.63] | 1,929.70 [1,802.95, 2,056.46] |
| | | Gay or Lesbian | 23,220 (1.8) | 410 (5.1) | 1,765.72 [1,598.92, 1,945.19] | 1,975.63 [1,779.86, 2,171.41] |
| | | Bisexual | 17,835 (1.4) | 400 (4.9) | 2,242.78 [2,028.35, 2,473.71] | 2,015.42 [1,799.20, 2,231.63] |
| | | Other | 4,050 (0.3) | 80 (1.0) | 1,975.31 [1,566.30, 2,458.44] | 1,912.79 [1,511.70, 2,386.58] |
| | England: North West | Total | 3,622,995 (100.0) | 16,080 (100.0) | 443.83 [437.00, 450.75] | 629.24 [618.00, 640.47] |
| | | Heterosexual | 3,489,035 (96.3) | 14,010 (87.1) | 401.54 [394.92, 408.25] | 557.33 [546.42, 568.24] |
| | | LGB+ | 133,960 (3.7) | 2,070 (12.9) | 1,545.24 [1,479.38, 1,613.27] | 1,532.90 [1,466.86, 1,598.93] |
| | | Gay or Lesbian | 70,060 (1.9) | 915 (5.7) | 1,306.02 [1,222.76, 1,393.46] | 1,494.02 [1,393.23, 1,594.82] |
| | | Bisexual | 51,090 (1.4) | 990 (6.2) | 1,937.76 [1,818.92, 2,062.32] | 1,715.56 [1,602.23, 1,828.90] |
| | | Other | 12,805 (0.4) | 165 (1.0) | 1,288.56 [1,099.44, 1,500.87] | 1,204.10 [1,019.56, 1,388.65] |
| | England: Yorkshire and The Humber | Total | 2,594,300 (100.0) | 17,790 (100.0) | 685.73 [675.69, 695.89] | 886.61 [871.37, 901.84] |
| | | Heterosexual | 2,500,105 (96.4) | 16,055 (90.2) | 642.17 [632.28, 652.18] | 818.33 [803.19, 833.48] |
| | | LGB+ | 94,195 (3.6) | 1,735 (9.8) | 1,841.92 [1,756.26, 1,930.68] | 1,779.12 [1,694.98, 1,863.26] |
| | | Gay or Lesbian | 43,275 (1.7) | 685 (3.9) | 1,582.90 [1,466.57, 1,706.01] | 1,766.70 [1,631.31, 1,902.09] |
| | | Bisexual | 40,260 (1.6) | 870 (4.9) | 2,160.95 [2,019.73, 2,309.45] | 1,925.09 [1,786.73, 2,063.44] |
| | | Other | 10,660 (0.4) | 175 (1.0) | 1,641.65 [1,407.43, 1,903.71] | 1,535.15 [1,304.49, 1,765.81] |
| | England: East Midlands | Total | 2,260,080 (100.0) | 11,780 (100.0) | 521.22 [511.85, 530.72] | 753.74 [738.33, 769.16] |
| | | Heterosexual | 2,184,410 (96.7) | 10,365 (88.0) | 474.50 [465.41, 483.72] | 677.41 [662.35, 692.47] |
| | | LGB+ | 75,670 (3.3) | 1,410 (12.0) | 1,863.35 [1,767.35, 1,963.22] | 1,787.80 [1,694.09, 1,881.52] |
| | | Gay or Lesbian | 33,700 (1.5) | 500 (4.2) | 1,483.68 [1,356.46, 1,619.61] | 1,735.17 [1,579.79, 1,890.56] |
| | | Bisexual | 33,525 (1.5) | 770 (6.5) | 2,296.79 [2,137.41, 2,464.92] | 1,992.22 [1,843.06, 2,141.37] |
| | | Other | 8,445 (0.4) | 140 (1.2) | 1,657.79 [1,394.56, 1,956.25] | 1,483.87 [1,235.03, 1,732.70] |
| | England: West Midlands | Total | 2,658,530 (100.0) | 10,160 (100.0) | 382.17 [374.77, 389.67] | 538.03 [526.19, 549.87] |
| | | Heterosexual | 2,576,680 (96.9) | 8,985 (88.4) | 348.70 [341.53, 355.99] | 484.10 [472.58, 495.62] |
| | | LGB+ | 81,855 (3.1) | 1,175 (11.6) | 1,435.47 [1,354.55, 1,519.95] | 1,379.88 [1,300.72, 1,459.05] |

| | | | | | | |
|---|---|---|---|---|---|---|
| | | Gay or Lesbian | 38,105 (1.4) | 455 (4.5) | 1,194.07 [1,086.86, 1,308.99] | 1,366.71 [1,237.66, 1,495.77] |
| | | Bisexual | 34,240 (1.3) | 610 (6.0) | 1,781.54 [1,642.95, 1,928.70] | 1,510.45 [1,385.50, 1,635.40] |
| | | Other | 9,510 (0.4) | 110 (1.1) | 1,156.68 [950.65, 1,394.11] | 1,068.78 [866.04, 1,271.51] |
| | England: East of England | Total | 3,091,550 (100.0) | 14,615 (100.0) | 472.74 [465.11, 480.47] | 682.25 [668.92, 695.58] |
| | | Heterosexual | 2,995,735 (96.9) | 13,030 (89.2) | 434.95 [427.52, 442.49] | 612.31 [599.32, 625.29] |
| | | LGB+ | 95,815 (3.1) | 1,585 (10.8) | 1,654.23 [1,573.78, 1,737.72] | 1,644.60 [1,563.43, 1,725.76] |
| | | Gay or Lesbian | 43,145 (1.4) | 545 (3.7) | 1,263.18 [1,159.34, 1,373.83] | 1,506.05 [1,374.34, 1,637.76] |
| | | Bisexual | 41,305 (1.3) | 860 (5.9) | 2,082.07 [1,945.23, 2,226.01] | 1,849.58 [1,721.34, 1,977.82] |
| | | Other | 11,370 (0.4) | 180 (1.2) | 1,583.11 [1,360.28, 1,832.04] | 1,518.54 [1,294.38, 1,742.70] |
| | England: London | Total | 4,294,885 (100.0) | 16,165 (100.0) | 376.38 [370.60, 382.23] | 447.81 [439.99, 455.63] |
| | | Heterosexual | 4,067,010 (94.7) | 14,330 (88.6) | 352.35 [346.60, 358.16] | 411.23 [403.50, 418.96] |
| | | LGB+ | 227,875 (5.3) | 1,840 (11.4) | 807.46 [770.98, 845.22] | 919.24 [875.57, 962.92] |
| | | Gay or Lesbian | 118,130 (2.8) | 710 (4.4) | 601.03 [557.63, 646.92] | 850.11 [773.45, 926.77] |
| | | Bisexual | 81,400 (1.9) | 875 (5.4) | 1,074.94 [1,004.88, 1,148.59] | 1,028.10 [957.79, 1,098.41] |
| | | Other | 28,350 (0.7) | 250 (1.5) | 881.83 [775.90, 998.19] | 914.58 [798.76, 1,030.40] |
| | England: South East | Total | 4,584,145 (100.0) | 19,865 (100.0) | 433.34 [427.34, 439.41] | 663.66 [652.71, 674.60] |
| | | Heterosexual | 4,418,790 (96.4) | 17,325 (87.2) | 392.08 [386.26, 397.96] | 585.80 [575.16, 596.44] |
| | | LGB+ | 165,355 (3.6) | 2,535 (12.8) | 1,533.07 [1,473.96, 1,593.93] | 1,541.89 [1,481.72, 1,602.05] |
| | | Gay or Lesbian | 78,375 (1.7) | 865 (4.4) | 1,103.67 [1,031.33, 1,179.74] | 1,420.32 [1,321.04, 1,519.59] |
| | | Bisexual | 68,910 (1.5) | 1,395 (7.0) | 2,024.38 [1,919.53, 2,133.47] | 1,721.18 [1,626.96, 1,815.40] |
| | | Other | 18,070 (0.4) | 280 (1.4) | 1,549.53 [1,373.33, 1,742.07] | 1,413.67 [1,245.24, 1,582.10] |
| | England: South West | Total | 2,722,320 (100.0) | 12,445 (100.0) | 457.15 [449.15, 465.25] | 730.31 [715.67, 744.95] |
| | | Heterosexual | 2,619,105 (96.2) | 10,610 (85.3) | 405.10 [397.43, 412.88] | 639.19 [624.93, 653.45] |
| | | LGB+ | 103,215 (3.8) | 1,835 (14.7) | 1,777.84 [1,697.42, 1,861.09] | 1,731.95 [1,652.28, 1,811.62] |
| | | Gay or Lesbian | 45,200 (1.7) | 645 (5.2) | 1,426.99 [1,318.98, 1,541.50] | 1,766.42 [1,626.48, 1,906.36] |
| | | Bisexual | 46,720 (1.7) | 980 (7.9) | 2,097.60 [1,968.31, 2,233.15] | 1,816.08 [1,695.34, 1,936.81] |
| | | Other | 11,295 (0.4) | 215 (1.7) | 1,903.50 [1,657.55, 2,175.66] | 1,687.53 [1,455.94, 1,919.12] |
| | Wales | Total | 1,196,160 (100.0) | 4,615 (100.0) | 385.82 [374.77, 397.11] | 566.21 [548.21, 584.20] |
| | | Heterosexual | 1,150,330 (96.2) | 4,050 (87.8) | 352.07 [341.31, 363.09] | 518.33 [500.46, 536.20] |
| | | LGB+ | 45,835 (3.8) | 565 (12.2) | 1,232.68 [1,133.12, 1,338.65] | 1,171.38 [1,074.37, 1,268.38] |
| | | Gay or Lesbian | 22,165 (1.9) | 235 (5.1) | 1,060.23 [929.00, 1,204.81] | 1,235.52 [1,073.42, 1,397.61] |
| | | Bisexual | 19,220 (1.6) | 290 (6.3) | 1,508.84 [1,340.17, 1,692.88] | 1,300.69 [1,138.26, 1,463.13] |
| | | Other | 4,450 (0.4) | 40 (0.9) | 898.88 [642.17, 1,224.01] | 719.23 [503.92, 991.95] |
| Ethnic group | Asian, Asian British or Asian Welsh | Total | 2,333,750 (100.0) | 6,780 (100.0) | 290.52 [283.64, 297.52] | 331.92 [323.37, 340.47] |
| | | Heterosexual | 2,279,735 (97.7) | 6,420 (94.7) | 281.61 [274.76, 288.59] | 320.86 [312.34, 329.39] |
| | | LGB+ | 54,010 (2.3) | 360 (5.3) | 666.54 [599.46, 739.08] | 718.88 [642.98, 794.79] |
| | | Gay or Lesbian | 16,255 (0.7) | 110 (1.6) | 676.71 [556.18, 815.62] | 819.34 [651.62, 987.07] |
| | | Bisexual | 27,480 (1.2) | 190 (2.8) | 691.41 [596.59, 797.02] | 683.63 [584.71, 782.56] |
| | | Other | 10,275 (0.4) | 60 (0.9) | 583.94 [445.61, 751.65] | 684.55 [517.04, 887.64] |
| | Black, Black British, Black Welsh, Caribbean or African | Total | 1,017,745 (100.0) | 3,590 (100.0) | 352.74 [341.30, 364.47] | 434.82 [419.22, 450.42] |
| | | Heterosexual | 993,825 (97.6) | 3,325 (92.6) | 334.57 [323.29, 346.14] | 408.17 [392.85, 423.48] |
| | | LGB+ | 23,915 (2.3) | 265 (7.4) | 1,108.09 [978.68, 1,249.85] | 1,181.34 [1,038.40, 1,324.27] |
| | | Gay or Lesbian | 7,200 (0.7) | 75 (2.1) | 1,041.67 [819.34, 1,305.74] | 1,287.86 [1,005.97, 1,622.67] |
| | | Bisexual | 10,935 (1.1) | 140 (3.9) | 1,280.29 [1,077.01, 1,510.80] | 1,203.33 [1,000.81, 1,405.85] |
| | | Other | 5,780 (0.6) | 50 (1.4) | 865.05 [642.06, 1,140.46] | 1,068.73 [793.86, 1,408.23] |

| | | | | | | |
|---|---|---|---|---|---|---|
| | Mixed or Multiple ethnic groups | Total | 520,740 (100.0) | 4,020 (100.0) | 771.98 [748.30, 796.22] | 794.72 [769.87, 819.56] |
| | | Heterosexual | 478,925 (92.0) | 3,225 (80.2) | 673.38 [650.34, 697.03] | 697.48 [673.03, 721.94] |
| | | LGB+ | 41,815 (8.0) | 795 (19.8) | 1,901.23 [1,771.35, 2,038.12] | 1,601.94 [1,486.48, 1,717.40] |
| | | Gay or Lesbian | 16,210 (3.1) | 240 (6.0) | 1,480.57 [1,299.16, 1,680.21] | 1,469.93 [1,281.83, 1,658.02] |
| | | Bisexual | 19,785 (3.8) | 460 (11.4) | 2,324.99 [2,117.35, 2,547.49] | 1,746.54 [1,559.80, 1,933.29] |
| | | Other | 5,825 (1.1) | 100 (2.5) | 1,716.74 [1,396.81, 2,088.01] | 1,441.38 [1,145.06, 1,737.70] |
| | White | Total | 23,897,995 (100.0) | 115,290 (100.0) | 482.43 [479.64, 485.22] | 713.70 [708.85, 718.56] |
| | | Heterosexual | 22,971,715 (96.1) | 101,205 (87.8) | 440.56 [437.85, 443.29] | 640.37 [635.59, 645.14] |
| | | LGB+ | 926,280 (3.9) | 14,085 (12.2) | 1,520.60 [1,495.59, 1,545.92] | 1,526.04 [1,500.84, 1,551.25] |
| | | Gay or Lesbian | 468,510 (2.0) | 5,500 (4.8) | 1,173.93 [1,143.11, 1,205.38] | 1,444.24 [1,404.10, 1,484.39] |
| | | Bisexual | 368,005 (1.5) | 7,195 (6.2) | 1,955.14 [1,910.22, 2,000.84] | 1,724.11 [1,681.98, 1,766.24] |
| | | Other | 89,765 (0.4) | 1,390 (1.2) | 1,548.49 [1,468.14, 1,632.09] | 1,425.04 [1,348.62, 1,501.46] |
| | Other ethnic group | Total | 569,435 (100.0) | 1,920 (100.0) | 337.18 [322.26, 352.60] | 384.29 [365.52, 403.06] |
| | | Heterosexual | 546,580 (96.0) | 1,770 (92.2) | 323.83 [308.92, 339.28] | 368.67 [349.80, 387.53] |
| | | LGB+ | 22,855 (4.0) | 150 (7.8) | 656.31 [555.49, 770.15] | 735.14 [613.34, 856.95] |
| | | Gay or Lesbian | 7,200 (1.3) | 40 (2.1) | 555.56 [396.90, 756.51] | 850.53 [579.78, 1,193.46] |
| | | Bisexual | 8,295 (1.5) | 65 (3.4) | 783.60 [604.77, 998.77] | 787.06 [606.73, 1,004.03] |
| | | Other | 7,360 (1.3) | 40 (2.1) | 543.48 [388.27, 740.06] | 583.93 [409.69, 804.61] |
| Rural/urban classification | Urban | Total | 23,090,095 (100.0) | 112,385 (100.0) | 486.72 [483.88, 489.58] | 653.52 [649.17, 657.87] |
| | | Heterosexual | 22,152,635 (95.9) | 98,745 (87.9) | 445.75 [442.97, 448.54] | 588.65 [584.38, 592.91] |
| | | LGB+ | 937,465 (4.1) | 13,645 (12.1) | 1,455.52 [1,431.20, 1,480.15] | 1,449.36 [1,425.02, 1,473.69] |
| | | Gay or Lesbian | 448,580 (1.9) | 5,220 (4.6) | 1,163.67 [1,132.32, 1,195.68] | 1,402.81 [1,362.83, 1,442.79] |
| | | Bisexual | 381,940 (1.7) | 6,975 (6.2) | 1,826.20 [1,783.59, 1,869.57] | 1,599.38 [1,559.86, 1,638.91] |
| | | Other | 106,940 (0.5) | 1,450 (1.3) | 1,355.90 [1,287.00, 1,427.53] | 1,278.91 [1,212.32, 1,345.49] |
| | Rural | Total | 5,249,565 (100.0) | 19,215 (100.0) | 366.03 [360.87, 371.24] | 627.07 [615.92, 638.22] |
| | | Heterosexual | 5,118,155 (97.5) | 17,205 (89.5) | 336.16 [331.15, 341.22] | 561.21 [550.38, 572.05] |
| | | LGB+ | 131,415 (2.5) | 2,010 (10.5) | 1,529.51 [1,463.36, 1,597.87] | 1,601.31 [1,529.56, 1,673.05] |
| | | Gay or Lesbian | 66,790 (1.3) | 745 (3.9) | 1,115.44 [1,036.77, 1,198.49] | 1,492.74 [1,377.86, 1,607.62] |
| | | Bisexual | 52,560 (1.0) | 1,070 (5.6) | 2,035.77 [1,915.60, 2,161.50] | 1,806.59 [1,693.45, 1,919.74] |
| | | Other | 12,065 (0.2) | 190 (1.0) | 1,574.80 [1,358.83, 1,815.35] | 1,478.78 [1,263.10, 1,694.45] |
| Country of birth | Born in the UK | Total | 22,921,610 (100.0) | 116,880 (100.0) | 509.91 [506.99, 512.84] | 738.91 [734.05, 743.78] |
| | | Heterosexual | 22,070,020 (96.3) | 102,595 (87.8) | 464.86 [462.02, 467.71] | 664.31 [659.54, 669.09] |
| | | LGB+ | 851,590 (3.7) | 14,285 (12.2) | 1,677.45 [1,650.05, 1,705.19] | 1,648.70 [1,621.61, 1,675.80] |
| | | Gay or Lesbian | 426,925 (1.9) | 5,515 (4.7) | 1,291.80 [1,257.93, 1,326.35] | 1,550.63 [1,508.17, 1,593.08] |
| | | Bisexual | 344,360 (1.5) | 7,340 (6.3) | 2,131.49 [2,083.00, 2,180.82] | 1,852.99 [1,807.61, 1,898.38] |
| | | Other | 80,305 (0.4) | 1,430 (1.2) | 1,780.71 [1,689.60, 1,875.46] | 1,596.46 [1,511.31, 1,681.61] |
| | Born outside the UK | Total | 5,418,055 (100.0) | 14,720 (100.0) | 271.68 [267.31, 276.11] | 333.07 [326.67, 339.46] |
| | | Heterosexual | 5,200,765 (96.0) | 13,355 (90.7) | 256.79 [252.45, 261.18] | 307.41 [301.13, 313.70] |
| | | LGB+ | 217,290 (4.0) | 1,370 (9.3) | 630.49 [597.55, 664.79] | 724.46 [684.24, 764.67] |
| | | Gay or Lesbian | 88,445 (1.6) | 450 (3.1) | 508.79 [462.86, 558.04] | 706.02 [624.37, 787.68] |
| | | Bisexual | 90,140 (1.7) | 705 (4.8) | 782.12 [725.44, 842.04] | 793.08 [733.06, 853.09] |
| | | Other | 38,700 (0.7) | 215 (1.5) | 555.56 [483.77, 634.99] | 611.83 [526.62, 697.03] |
| English language proficiency | Main language is English | Total | 25,715,175 (100.0) | 125,350 (100.0) | 487.46 [484.76, 490.16] | 698.29 [693.83, 702.75] |
| | | Heterosexual | 24,744,730 (96.2) | 110,150 (87.9) | 445.15 [442.52, 447.78] | 627.69 [623.32, 632.07] |
| | | LGB+ | 970,445 (3.8) | 15,200 (12.1) | 1,566.29 [1,541.49, 1,591.39] | 1,558.03 [1,533.25, 1,582.82] |

| | | | | | | |
|---|---|---|---|---|---|---|
| | | Gay or Lesbian | 483,460 (1.9) | 5,845 (4.7) | 1,208.99 [1,178.20, 1,240.39] | 1,476.16 [1,436.56, 1,515.76] |
| | | Bisexual | 391,770 (1.5) | 7,825 (6.2) | 1,997.35 [1,953.33, 2,042.10] | 1,740.63 [1,699.79, 1,781.47] |
| | | Other | 95,215 (0.4) | 1,535 (1.2) | 1,612.14 [1,532.49, 1,694.86] | 1,466.35 [1,391.64, 1,541.07] |
| | Main language is not English | Total | 2,624,490 (100.0) | 6,255 (100.0) | 238.33 [232.46, 244.31] | 275.60 [267.66, 283.54] |
| | | Heterosexual | 2,526,055 (96.2) | 5,800 (92.7) | 229.61 [223.74, 235.59] | 261.53 [253.64, 269.42] |
| | | LGB+ | 98,435 (3.8) | 455 (7.3) | 462.23 [420.73, 506.72] | 535.19 [482.29, 588.10] |
| | | Gay or Lesbian | 31,910 (1.2) | 125 (2.0) | 391.73 [326.07, 466.72] | 588.45 [455.41, 721.49] |
| | | Bisexual | 42,730 (1.6) | 225 (3.6) | 526.56 [460.00, 600.05] | 554.45 [479.01, 629.89] |
| | | Other | 23,790 (0.9) | 110 (1.8) | 462.38 [380.02, 557.29] | 491.07 [392.86, 589.28] |
| Religion | No religion | Total | 10,589,125 (100.0) | 66,660 (100.0) | 629.51 [624.74, 634.31] | 773.46 [767.20, 779.72] |
| | | Heterosexual | 9,914,285 (93.6) | 56,050 (84.1) | 565.35 [560.68, 570.05] | 693.54 [687.31, 699.77] |
| | | LGB+ | 674,845 (6.4) | 10,610 (15.9) | 1,572.21 [1,542.44, 1,602.42] | 1,445.07 [1,417.26, 1,472.88] |
| | | Gay or Lesbian | 322,090 (3.0) | 3,960 (5.9) | 1,229.47 [1,191.47, 1,268.37] | 1,370.76 [1,327.08, 1,414.45] |
| | | Bisexual | 288,305 (2.7) | 5,620 (8.4) | 1,949.32 [1,898.69, 2,000.97] | 1,621.51 [1,573.75, 1,669.26] |
| | | Other | 64,450 (0.6) | 1,030 (1.5) | 1,598.14 [1,502.02, 1,698.80] | 1,369.61 [1,281.04, 1,458.18] |
| | Christian | Total | 14,170,955 (100.0) | 50,935 (100.0) | 359.43 [356.32, 362.57] | 560.84 [554.21, 567.46] |
| | | Heterosexual | 13,900,760 (98.1) | 47,590 (93.4) | 342.36 [339.29, 345.45] | 520.87 [514.39, 527.35] |
| | | LGB+ | 270,195 (1.9) | 3,345 (6.6) | 1,237.99 [1,196.39, 1,280.67] | 1,527.06 [1,472.48, 1,581.64] |
| | | Gay or Lesbian | 156,185 (1.1) | 1,520 (3.0) | 973.20 [924.89, 1,023.39] | 1,462.11 [1,374.76, 1,549.46] |
| | | Bisexual | 87,620 (0.6) | 1,550 (3.0) | 1,769.00 [1,682.02, 1,859.31] | 1,795.60 [1,705.67, 1,885.52] |
| | | Other | 26,390 (0.2) | 275 (0.5) | 1,042.06 [922.53, 1,172.78] | 1,118.30 [984.43, 1,252.17] |
| | Buddhist | Total | 145,815 (100.0) | 675 (100.0) | 462.92 [428.65, 499.19] | 642.15 [582.74, 701.57] |
| | | Heterosexual | 133,095 (91.3) | 490 (72.6) | 368.16 [336.28, 402.25] | 468.83 [415.46, 522.21] |
| | | LGB+ | 12,720 (8.7) | 185 (27.4) | 1,454.40 [1,252.37, 1,679.75] | 1,795.76 [1,519.51, 2,072.02] |
| | | Gay or Lesbian | 5,515 (3.8) | 60 (8.9) | 1,087.94 [830.21, 1,400.40] | 1,505.16 [1,055.95, 2,049.77] |
| | | Bisexual | 5,460 (3.7) | 95 (14.1) | 1,739.93 [1,407.70, 2,126.97] | 1,846.33 [1,481.57, 2,271.29] |
| | | Other | 1,745 (1.2) | 35 (5.2) | 2,005.73 [1,397.06, 2,789.49] | 2,099.05 [1,438.83, 2,949.22] |
| | Hindu | Total | 500,755 (100.0) | 1,195 (100.0) | 238.64 [225.30, 252.56] | 293.62 [273.96, 313.29] |
| | | Heterosexual | 490,280 (97.9) | 1,140 (95.4) | 232.52 [219.22, 246.42] | 284.69 [265.07, 304.30] |
| | | LGB+ | 10,475 (2.1) | 55 (4.6) | 525.06 [395.55, 683.44] | 622.06 [458.87, 821.63] |
| | | Gay or Lesbian | 1,900 (0.4) | 15 (1.3) | 789.47 [441.86, 1,302.12] | 837.77 [362.81, 1,538.23] |
| | | Bisexual | 6,875 (1.4) | 35 (2.9) | 509.09 [354.60, 708.02] | 615.19 [420.62, 865.74] |
| | | Other | 1,700 (0.3) | 5 (0.4) | 294.12 [95.50, 686.37] | 555.09 [208.32, 1,239.93] |
| | Jewish | Total | 124,415 (100.0) | 535 (100.0) | 430.01 [394.34, 468.04] | 564.53 [498.48, 630.58] |
| | | Heterosexual | 117,720 (94.6) | 455 (85.0) | 386.51 [351.81, 423.71] | 454.03 [391.93, 516.13] |
| | | LGB+ | 6,700 (5.4) | 80 (15.0) | 1,194.03 [946.79, 1,486.07] | 1,342.74 [1,060.07, 1,676.64] |
| | | Gay or Lesbian | 3,305 (2.7) | 25 (4.7) | 756.43 [489.52, 1,116.64] | 1,087.33 [687.90, 1,626.39] |
| | | Bisexual | 2,570 (2.1) | 40 (7.5) | 1,556.42 [1,111.93, 2,119.40] | 1,570.27 [1,125.64, 2,133.44] |
| | | Other | 820 (0.7) | 10 (1.9) | 1,219.51 [584.80, 2,242.73] | 1,265.23 [573.62, 2,380.19] |
| | Muslim | Total | 1,495,785 (100.0) | 5,200 (100.0) | 347.64 [338.26, 357.22] | 374.21 [363.43, 384.98] |
| | | Heterosexual | 1,469,860 (98.3) | 5,025 (96.6) | 341.87 [332.48, 351.45] | 368.32 [357.53, 379.11] |
| | | LGB+ | 25,930 (1.7) | 175 (3.4) | 674.89 [578.60, 782.63] | 780.56 [657.64, 903.47] |
| | | Gay or Lesbian | 3,850 (0.3) | 40 (0.8) | 1,038.96 [742.25, 1,414.77] | 1,217.01 [839.30, 1,695.41] |
| | | Bisexual | 12,495 (0.8) | 90 (1.7) | 720.29 [579.20, 885.36] | 824.51 [656.19, 1,021.45] |
| | | Other | 9,585 (0.6) | 45 (0.9) | 469.48 [342.44, 628.21] | 543.66 [379.12, 749.25] |

| | | | | | | |
|---|---|---|---|---|---|---|
| | Sikh | Total | 248,420 (100.0) | 810 (100.0) | 326.06 [303.99, 349.31] | 365.16 [336.58, 393.74] |
| | | Heterosexual | 244,010 (98.2) | 780 (96.3) | 319.66 [297.62, 342.90] | 355.20 [326.74, 383.66] |
| | | LGB+ | 4,410 (1.8) | 30 (3.7) | 680.27 [458.98, 971.13] | 751.20 [497.45, 1,084.71] |
| | | Gay or Lesbian | 815 (0.3) | 10 (1.2) | 1,226.99 [588.39, 2,256.49] | 1,558.52 [737.76, 2,881.67] |
| | | Bisexual | 2,310 (0.9) | 10 (1.2) | 432.90 [207.59, 796.12] | 413.80 [189.43, 775.52] |
| | | Other | 1,285 (0.5) | 10 (1.2) | 778.21 [373.18, 1,431.16] | 944.46 [391.84, 1,835.35] |
| | Other religion | Total | 198,175 (100.0) | 1,665 (100.0) | 840.17 [800.29, 881.51] | 1,174.63 [1,111.02, 1,238.24] |
| | | Heterosexual | 163,850 (82.7) | 945 (56.8) | 576.75 [540.56, 614.72] | 735.86 [676.05, 795.67] |
| | | LGB+ | 34,325 (17.3) | 720 (43.2) | 2,097.60 [1,947.16, 2,256.57] | 2,041.84 [1,891.83, 2,191.85] |
| | | Gay or Lesbian | 8,220 (4.1) | 160 (9.6) | 1,946.47 [1,656.55, 2,272.53] | 2,190.95 [1,843.95, 2,537.94] |
| | | Bisexual | 17,285 (8.7) | 380 (22.8) | 2,198.44 [1,982.93, 2,430.98] | 2,043.37 [1,833.30, 2,253.44] |
| | | Other | 8,820 (4.5) | 180 (10.8) | 2,040.82 [1,753.56, 2,361.71] | 1,922.87 [1,637.96, 2,207.78] |
| | Not answered | Total | 866,220 (100.0) | 3,920 (100.0) | 452.54 [438.48, 466.93] | 622.04 [599.51, 644.57] |
| | | Heterosexual | 836,935 (96.6) | 3,465 (88.4) | 414.01 [400.34, 428.03] | 558.80 [536.87, 580.73] |
| | | LGB+ | 29,285 (3.4) | 455 (11.6) | 1,553.70 [1,414.20, 1,703.23] | 1,563.10 [1,418.95, 1,707.25] |
| | | Gay or Lesbian | 13,495 (1.6) | 175 (4.5) | 1,296.78 [1,111.76, 1,503.78] | 1,553.32 [1,306.30, 1,800.35] |
| | | Bisexual | 11,575 (1.3) | 225 (5.7) | 1,943.84 [1,698.13, 2,215.12] | 1,753.74 [1,515.72, 1,991.76] |
| | | Other | 4,215 (0.5) | 55 (1.4) | 1,304.86 [983.00, 1,698.46] | 1,244.61 [930.32, 1,628.95] |
| IMD/WIMD decile group | 1 (most deprived) | Total | 2,360,410 (100.0) | 19,200 (100.0) | 813.42 [801.95, 825.01] | 966.70 [952.01, 981.38] |
| | | Heterosexual | 2,252,860 (95.4) | 16,890 (88.0) | 749.71 [738.45, 761.11] | 885.17 [870.68, 899.66] |
| | | LGB+ | 107,550 (4.6) | 2,310 (12.0) | 2,147.84 [2,061.13, 2,237.25] | 2,094.47 [2,008.83, 2,180.10] |
| | | Gay or Lesbian | 48,485 (2.1) | 965 (5.0) | 1,990.31 [1,866.70, 2,119.95] | 2,188.64 [2,047.21, 2,330.07] |
| | | Bisexual | 45,390 (1.9) | 1,155 (6.0) | 2,544.61 [2,399.96, 2,695.71] | 2,302.16 [2,163.13, 2,441.19] |
| | | Other | 13,675 (0.6) | 190 (1.0) | 1,389.40 [1,198.85, 1,601.62] | 1,345.01 [1,149.80, 1,540.21] |
| | 2 | Total | 2,649,785 (100.0) | 17,510 (100.0) | 660.81 [651.06, 670.67] | 810.54 [797.43, 823.65] |
| | | Heterosexual | 2,525,935 (95.3) | 15,425 (88.1) | 610.66 [601.07, 620.38] | 741.90 [728.94, 754.86] |
| | | LGB+ | 123,850 (4.7) | 2,085 (11.9) | 1,683.49 [1,611.99, 1,757.34] | 1,684.78 [1,612.19, 1,757.37] |
| | | Gay or Lesbian | 57,330 (2.2) | 850 (4.9) | 1,482.64 [1,384.63, 1,585.76] | 1,709.09 [1,588.32, 1,829.86] |
| | | Bisexual | 51,035 (1.9) | 1,015 (5.8) | 1,988.83 [1,868.34, 2,115.05] | 1,763.38 [1,650.97, 1,875.79] |
| | | Other | 15,485 (0.6) | 225 (1.3) | 1,453.02 [1,269.35, 1,655.80] | 1,426.86 [1,237.07, 1,616.64] |
| | 3 | Total | 2,821,390 (100.0) | 16,015 (100.0) | 567.63 [558.87, 576.49] | 712.36 [700.17, 724.56] |
| | | Heterosexual | 2,685,000 (95.2) | 14,005 (87.4) | 521.60 [513.00, 530.31] | 646.11 [634.09, 658.12] |
| | | LGB+ | 136,395 (4.8) | 2,015 (12.6) | 1,477.33 [1,413.52, 1,543.27] | 1,505.93 [1,439.72, 1,572.14] |
| | | Gay or Lesbian | 64,690 (2.3) | 830 (5.2) | 1,283.04 [1,197.23, 1,373.38] | 1,562.46 [1,448.26, 1,676.66] |
| | | Bisexual | 55,235 (2.0) | 985 (6.2) | 1,783.29 [1,673.65, 1,898.23] | 1,603.59 [1,498.71, 1,708.47] |
| | | Other | 16,470 (0.6) | 200 (1.2) | 1,214.33 [1,051.86, 1,394.79] | 1,187.00 [1,019.53, 1,354.46] |
| | 4 | Total | 2,880,030 (100.0) | 14,635 (100.0) | 508.15 [499.95, 516.46] | 666.87 [654.64, 679.09] |
| | | Heterosexual | 2,748,740 (95.4) | 12,770 (87.3) | 464.58 [456.55, 472.71] | 597.91 [585.96, 609.87] |
| | | LGB+ | 131,290 (4.6) | 1,865 (12.7) | 1,420.52 [1,356.77, 1,486.49] | 1,451.34 [1,385.24, 1,517.43] |
| | | Gay or Lesbian | 63,465 (2.2) | 720 (4.9) | 1,134.48 [1,053.12, 1,220.47] | 1,433.42 [1,320.87, 1,545.96] |
| | | Bisexual | 52,965 (1.8) | 940 (6.4) | 1,774.76 [1,663.10, 1,891.94] | 1,586.83 [1,480.24, 1,693.42] |
| | | Other | 14,865 (0.5) | 210 (1.4) | 1,412.71 [1,228.09, 1,617.26] | 1,360.13 [1,172.95, 1,547.31] |
| | 5 | Total | 2,915,180 (100.0) | 13,195 (100.0) | 452.63 [444.94, 460.42] | 620.22 [608.00, 632.43] |
| | | Heterosexual | 2,797,960 (96.0) | 11,525 (87.3) | 411.91 [404.42, 419.50] | 551.04 [539.15, 562.93] |
| | | LGB+ | 117,220 (4.0) | 1,670 (12.7) | 1,424.67 [1,357.15, 1,494.68] | 1,439.06 [1,370.03, 1,508.09] |

| | | | | | | |
|---|---|---|---|---|---|---|
| | | Gay or Lesbian | 56,405 (1.9) | 620 (4.7) | 1,099.19 [1,014.36, 1,189.22] | 1,359.54 [1,246.14, 1,472.93] |
| | | Bisexual | 48,025 (1.6) | 870 (6.6) | 1,811.56 [1,693.16, 1,936.04] | 1,549.95 [1,442.07, 1,657.83] |
| | | Other | 12,790 (0.4) | 185 (1.4) | 1,446.44 [1,245.51, 1,670.56] | 1,356.40 [1,158.96, 1,553.85] |
| | 6 | Total | 2,979,555 (100.0) | 12,505 (100.0) | 419.69 [412.37, 427.12] | 590.71 [578.48, 602.93] |
| | | Heterosexual | 2,866,330 (96.2) | 11,025 (88.2) | 384.64 [377.49, 391.89] | 529.49 [517.51, 541.46] |
| | | LGB+ | 113,225 (3.8) | 1,480 (11.8) | 1,307.13 [1,241.38, 1,375.46] | 1,322.65 [1,255.23, 1,390.07] |
| | | Gay or Lesbian | 55,740 (1.9) | 520 (4.2) | 932.90 [854.43, 1,016.64] | 1,164.72 [1,059.06, 1,270.38] |
| | | Bisexual | 45,545 (1.5) | 810 (6.5) | 1,778.46 [1,658.08, 1,905.27] | 1,572.25 [1,458.40, 1,686.10] |
| | | Other | 11,945 (0.4) | 145 (1.2) | 1,213.90 [1,024.36, 1,428.34] | 1,170.65 [979.56, 1,361.74] |
| | 7 | Total | 2,941,315 (100.0) | 11,095 (100.0) | 377.21 [370.23, 384.30] | 558.40 [545.83, 570.97] |
| | | Heterosexual | 2,843,740 (96.7) | 9,840 (88.7) | 346.02 [339.22, 352.93] | 495.56 [483.36, 507.75] |
| | | LGB+ | 97,570 (3.3) | 1,255 (11.3) | 1,286.26 [1,216.07, 1,359.44] | 1,302.86 [1,230.65, 1,375.06] |
| | | Gay or Lesbian | 48,480 (1.6) | 415 (3.7) | 856.02 [775.64, 942.48] | 1,122.47 [1,008.39, 1,236.55] |
| | | Bisexual | 39,240 (1.3) | 700 (6.3) | 1,783.89 [1,654.17, 1,921.08] | 1,518.66 [1,400.30, 1,637.03] |
| | | Other | 9,855 (0.3) | 140 (1.3) | 1,420.60 [1,195.03, 1,676.36] | 1,321.19 [1,101.61, 1,540.76] |
| | 8 | Total | 2,952,670 (100.0) | 10,590 (100.0) | 358.66 [351.86, 365.56] | 538.09 [525.52, 550.65] |
| | | Heterosexual | 2,862,105 (96.9) | 9,425 (89.0) | 329.30 [322.69, 336.02] | 480.53 [468.27, 492.79] |
| | | LGB+ | 90,570 (3.1) | 1,170 (11.0) | 1,291.82 [1,218.85, 1,368.02] | 1,288.06 [1,213.93, 1,362.19] |
| | | Gay or Lesbian | 45,690 (1.5) | 430 (4.1) | 941.12 [854.26, 1,034.42] | 1,166.95 [1,050.78, 1,283.12] |
| | | Bisexual | 36,040 (1.2) | 625 (5.9) | 1,734.18 [1,600.87, 1,875.63] | 1,477.00 [1,354.03, 1,599.97] |
| | | Other | 8,835 (0.3) | 115 (1.1) | 1,301.64 [1,074.64, 1,562.42] | 1,210.82 [984.55, 1,437.10] |
| | 9 | Total | 2,942,955 (100.0) | 9,350 (100.0) | 317.71 [311.30, 324.21] | 494.40 [481.89, 506.90] |
| | | Heterosexual | 2,860,805 (97.2) | 8,350 (89.3) | 291.88 [285.65, 298.20] | 441.53 [429.33, 453.73] |
| | | LGB+ | 82,150 (2.8) | 1,000 (10.7) | 1,217.29 [1,143.00, 1,295.13] | 1,200.70 [1,125.82, 1,275.57] |
| | | Gay or Lesbian | 41,210 (1.4) | 365 (3.9) | 885.71 [797.16, 981.40] | 1,125.01 [1,004.37, 1,245.65] |
| | | Bisexual | 32,940 (1.1) | 510 (5.5) | 1,548.27 [1,416.80, 1,688.66] | 1,344.20 [1,219.45, 1,468.95] |
| | | Other | 8,000 (0.3) | 125 (1.3) | 1,562.50 [1,300.61, 1,861.65] | 1,356.40 [1,112.27, 1,600.53] |
| | 10 (least deprived) | Total | 2,896,370 (100.0) | 7,500 (100.0) | 258.94 [253.12, 264.87] | 435.19 [422.59, 447.79] |
| | | Heterosexual | 2,827,310 (97.6) | 6,700 (89.3) | 236.97 [231.33, 242.72] | 383.99 [371.80, 396.18] |
| | | LGB+ | 69,055 (2.4) | 805 (10.7) | 1,165.74 [1,086.59, 1,249.13] | 1,098.10 [1,020.23, 1,175.98] |
| | | Gay or Lesbian | 33,875 (1.2) | 255 (3.4) | 752.77 [663.20, 851.06] | 926.97 [808.56, 1,045.38] |
| | | Bisexual | 28,090 (1.0) | 440 (5.9) | 1,566.39 [1,423.43, 1,719.82] | 1,266.17 [1,137.54, 1,394.80] |
| | | Other | 7,090 (0.2) | 110 (1.5) | 1,551.48 [1,275.13, 1,869.95] | 1,213.28 [969.53, 1,457.03] |
| Highest level of qualification (≥25 years) | No qualifications | Total | 4,082,460 (100.0) | 22,360 (100.0) | 547.71 [540.55, 554.94] | 973.29 [955.67, 990.91] |
| | | Heterosexual | 4,016,180 (98.4) | 21,325 (95.4) | 530.98 [523.87, 538.15] | 936.49 [918.90, 954.08] |
| | | LGB+ | 66,280 (1.6) | 1,035 (4.6) | 1,561.56 [1,467.86, 1,659.67] | 1,854.87 [1,735.03, 1,974.70] |
| | | Gay or Lesbian | 28,945 (0.7) | 525 (2.3) | 1,813.78 [1,661.93, 1,975.78] | 2,639.76 [2,389.53, 2,889.98] |
| | | Bisexual | 22,360 (0.5) | 415 (1.9) | 1,855.99 [1,681.70, 2,043.44] | 2,043.27 [1,839.44, 2,247.10] |
| | | Other | 14,975 (0.4) | 95 (0.4) | 634.39 [513.26, 775.51] | 657.38 [528.17, 807.91] |
| | Level 1 (one to four GCSE passes (grade A* to C or grade 4 and above) and any other GCSEs at other grades, or equivalent qualifications) | Total | 2,418,070 (100.0) | 12,420 (100.0) | 513.63 [504.64, 522.75] | 839.17 [820.57, 857.76] |
| | | Heterosexual | 2,366,905 (97.9) | 11,530 (92.8) | 487.13 [478.28, 496.11] | 786.93 [768.60, 805.27] |
| | | LGB+ | 51,165 (2.1) | 890 (7.2) | 1,739.47 [1,627.05, 1,857.61] | 1,997.02 [1,861.50, 2,132.55] |
| | | Gay or Lesbian | 28,635 (1.2) | 415 (3.3) | 1,449.28 [1,313.18, 1,595.65] | 1,856.74 [1,663.76, 2,049.73] |
| | | Bisexual | 17,680 (0.7) | 415 (3.3) | 2,347.29 [2,126.85, 2,584.36] | 2,365.22 [2,135.95, 2,594.48] |
| | | Other | 4,850 (0.2) | 60 (0.5) | 1,237.11 [944.05, 1,592.41] | 1,390.01 [1,057.37, 1,793.30] |

| | | | | | | |
|---|---|---|---|---|---|---|
| | Level 2 (five or more GCSE passes (grade A* to C or grade 4 and above) or equivalent qualifications) | Total | 3,077,650 (100.0) | 15,025 (100.0) | 488.20 [480.42, 496.07] | 683.99 [671.27, 696.72] |
| | | Heterosexual | 2,992,585 (97.2) | 13,680 (91.0) | 457.13 [449.50, 464.86] | 632.24 [619.74, 644.74] |
| | | LGB+ | 85,070 (2.8) | 1,345 (9.0) | 1,581.05 [1,497.67, 1,667.86] | 1,630.18 [1,542.62, 1,717.74] |
| | | Gay or Lesbian | 48,610 (1.6) | 585 (3.9) | 1,203.46 [1,107.90, 1,305.05] | 1,367.66 [1,253.06, 1,482.27] |
| | | Bisexual | 29,325 (1.0) | 650 (4.3) | 2,216.54 [2,049.39, 2,393.69] | 2,020.58 [1,863.96, 2,177.20] |
| | | Other | 7,130 (0.2) | 110 (0.7) | 1,542.78 [1,267.98, 1,859.46] | 1,452.61 [1,179.26, 1,725.95] |
| | Apprenticeship | Total | 1,302,750 (100.0) | 5,035 (100.0) | 386.49 [375.89, 397.32] | 602.60 [581.90, 623.30] |
| | | Heterosexual | 1,281,405 (98.4) | 4,780 (94.9) | 373.03 [362.53, 383.76] | 575.03 [554.49, 595.57] |
| | | LGB+ | 21,345 (1.6) | 250 (5.0) | 1,171.23 [1,030.54, 1,325.78] | 1,257.40 [1,100.66, 1,414.15] |
| | | Gay or Lesbian | 10,355 (0.8) | 110 (2.2) | 1,062.29 [873.07, 1,280.35] | 1,254.64 [1,014.60, 1,494.68] |
| | | Bisexual | 7,780 (0.6) | 120 (2.4) | 1,542.42 [1,278.82, 1,844.35] | 1,511.42 [1,240.52, 1,782.32] |
| | | Other | 3,205 (0.2) | 20 (0.4) | 624.02 [381.17, 963.76] | 688.66 [421.24, 1,062.76] |
| | Level 3 (two or more A Levels or equivalent qualifications) | Total | 4,037,275 (100.0) | 16,500 (100.0) | 408.69 [402.48, 414.98] | 488.24 [480.25, 496.23] |
| | | Heterosexual | 3,900,045 (96.6) | 14,575 (88.3) | 373.71 [367.67, 379.83] | 441.48 [433.72, 449.24] |
| | | LGB+ | 137,235 (3.4) | 1,925 (11.7) | 1,402.70 [1,340.74, 1,466.80] | 1,325.74 [1,266.10, 1,385.38] |
| | | Gay or Lesbian | 76,740 (1.9) | 885 (5.4) | 1,153.24 [1,078.51, 1,231.80] | 1,174.04 [1,096.40, 1,251.67] |
| | | Bisexual | 48,230 (1.2) | 840 (5.1) | 1,741.65 [1,625.85, 1,863.53] | 1,561.84 [1,449.31, 1,674.37] |
| | | Other | 12,260 (0.3) | 205 (1.2) | 1,672.10 [1,451.03, 1,917.34] | 1,475.34 [1,262.37, 1,688.31] |
| | Level 4 and above (Higher National Certificate, Higher National Diploma, Bachelor's degree, or post-graduate qualifications) | Total | 10,077,510 (100.0) | 25,810 (100.0) | 256.11 [253.00, 259.26] | 286.11 [282.32, 289.90] |
| | | Heterosexual | 9,635,435 (95.6) | 22,905 (88.7) | 237.72 [234.65, 240.82] | 262.12 [258.39, 265.85] |
| | | LGB+ | 442,075 (4.4) | 2,905 (11.3) | 657.13 [633.45, 681.47] | 643.82 [620.36, 667.29] |
| | | Gay or Lesbian | 244,490 (2.4) | 1,315 (5.1) | 537.85 [509.17, 567.73] | 561.06 [530.35, 591.76] |
| | | Bisexual | 154,580 (1.5) | 1,245 (4.8) | 805.41 [761.29, 851.42] | 753.98 [709.53, 798.43] |
| | | Other | 43,005 (0.4) | 350 (1.4) | 813.86 [730.82, 903.75] | 754.05 [671.76, 836.34] |
| | Other qualifications | Total | 675,670 (100.0) | 2,550 (100.0) | 377.40 [362.90, 392.34] | 556.37 [527.55, 585.20] |
| | | Heterosexual | 662,890 (98.1) | 2,420 (94.9) | 365.07 [350.67, 379.91] | 533.21 [504.41, 562.02] |
| | | LGB+ | 12,780 (1.9) | 130 (5.1) | 1,017.21 [849.88, 1,207.86] | 1,112.72 [916.30, 1,309.15] |
| | | Gay or Lesbian | 6,240 (0.9) | 65 (2.5) | 1,041.67 [803.94, 1,327.69] | 1,317.34 [1,000.10, 1,699.01] |
| | | Bisexual | 4,360 (0.6) | 50 (2.0) | 1,146.79 [851.17, 1,511.90] | 1,158.36 [856.51, 1,531.17] |
| | | Other | 2,185 (0.3) | 15 (0.6) | 686.50 [384.23, 1,132.28] | 625.16 [335.37, 1,052.53] |
| NS-SEC | Class 1 (Higher managerial, administrative and professional occupations) | Total | 4,318,935 (100.0) | 8,080 (100.0) | 187.08 [183.03, 191.21] | 284.19 [264.81, 303.57] |
| | | Heterosexual | 4,152,390 (96.1) | 7,270 (90.0) | 175.08 [171.08, 179.15] | 247.34 [229.85, 264.82] |
| | | LGB+ | 166,550 (3.9) | 810 (10.0) | 486.34 [453.42, 521.02] | 833.83 [659.41, 1,008.26] |
| | | Gay or Lesbian | 100,830 (2.3) | 395 (4.9) | 391.75 [354.06, 432.35] | 723.50 [434.17, 1,012.83] |
| | | Bisexual | 52,810 (1.2) | 330 (4.1) | 624.88 [559.27, 696.07] | 954.35 [708.06, 1,200.64] |
| | | Other | 12,905 (0.3) | 85 (1.1) | 658.66 [526.11, 814.44] | 857.82 [431.46, 1,358.93] |
| | Class 2 (Lower managerial, administrative and professional occupations) | Total | 6,300,945 (100.0) | 17,775 (100.0) | 282.10 [277.97, 286.28] | 437.31 [422.50, 452.12] |
| | | Heterosexual | 6,036,965 (95.8) | 15,715 (88.4) | 260.31 [256.26, 264.42] | 381.91 [367.99, 395.83] |
| | | LGB+ | 263,980 (4.2) | 2,060 (11.6) | 780.36 [747.02, 814.81] | 1,094.18 [1,004.15, 1,184.22] |
| | | Gay or Lesbian | 150,670 (2.4) | 930 (5.2) | 617.24 [578.21, 658.22] | 976.43 [827.86, 1,125.00] |
| | | Bisexual | 90,970 (1.4) | 920 (5.2) | 1,011.32 [947.02, 1,078.84] | 1,241.00 [1,111.74, 1,370.25] |
| | | Other | 22,335 (0.4) | 215 (1.2) | 962.61 [838.24, 1,100.25] | 1,104.10 [853.10, 1,355.09] |
| | Class 3 (Intermediate occupations) | Total | 3,496,100 (100.0) | 13,640 (100.0) | 390.15 [383.63, 396.75] | 592.26 [578.14, 606.38] |
| | | Heterosexual | 3,384,320 (96.8) | 12,100 (88.7) | 357.53 [351.19, 363.96] | 529.72 [515.98, 543.45] |
| | | LGB+ | 111,780 (3.2) | 1,540 (11.3) | 1,377.71 [1,309.75, 1,448.28] | 1,498.54 [1,409.32, 1,587.77] |

| | | | | | | |
|---|---|---|---|---|---|---|
| | | Gay or Lesbian | 55,885 (1.6) | 640 (4.7) | 1,145.21 [1,058.19, 1,237.47] | 1,431.23 [1,284.78, 1,577.68] |
| | | Bisexual | 45,065 (1.3) | 775 (5.7) | 1,719.74 [1,600.78, 1,845.20] | 1,681.78 [1,545.16, 1,818.40] |
| | | Other | 10,830 (0.3) | 125 (0.9) | 1,154.20 [960.75, 1,375.18] | 1,153.51 [917.04, 1,389.98] |
| | Class 4 (Small employers and own account workers) | Total | 2,965,195 (100.0) | 10,485 (100.0) | 353.60 [346.87, 360.44] | 610.62 [588.30, 632.94] |
| | | Heterosexual | 2,895,580 (97.7) | 9,920 (94.6) | 342.59 [335.88, 349.40] | 584.30 [562.11, 606.50] |
| | | LGB+ | 69,615 (2.3) | 565 (5.4) | 811.61 [746.06, 881.37] | 1,197.49 [1,039.11, 1,355.87] |
| | | Gay or Lesbian | 35,085 (1.2) | 250 (2.4) | 712.56 [626.96, 806.58] | 1,183.39 [861.94, 1,504.83] |
| | | Bisexual | 24,175 (0.8) | 245 (2.3) | 1,013.44 [890.50, 1,148.61] | 1,390.51 [1,146.56, 1,634.46] |
| | | Other | 10,355 (0.3) | 70 (0.7) | 676.00 [526.98, 854.09] | 758.30 [538.77, 1,020.65] |
| | Class 5 (Lower supervisory and technical occupations) | Total | 1,472,395 (100.0) | 6,685 (100.0) | 454.02 [443.20, 465.04] | 641.29 [621.22, 661.37] |
| | | Heterosexual | 1,429,330 (97.1) | 6,095 (91.2) | 426.42 [415.78, 437.27] | 590.74 [571.23, 610.25] |
| | | LGB+ | 43,065 (2.9) | 590 (8.8) | 1,370.02 [1,261.69, 1,485.17] | 1,675.40 [1,501.58, 1,849.21] |
| | | Gay or Lesbian | 22,650 (1.5) | 285 (4.3) | 1,258.28 [1,116.42, 1,413.17] | 1,607.73 [1,350.69, 1,864.76] |
| | | Bisexual | 15,880 (1.1) | 260 (3.9) | 1,637.28 [1,444.30, 1,848.87] | 1,826.50 [1,565.80, 2,087.20] |
| | | Other | 4,535 (0.3) | 45 (0.7) | 992.28 [723.78, 1,327.75] | 1,533.60 [921.16, 2,298.78] |
| | Class 6 (Semi-routine occupations) | Total | 3,183,850 (100.0) | 20,720 (100.0) | 650.78 [641.95, 659.71] | 959.66 [944.20, 975.12] |
| | | Heterosexual | 3,070,855 (96.5) | 18,185 (87.8) | 592.18 [583.60, 600.85] | 861.29 [846.17, 876.42] |
| | | LGB+ | 112,990 (3.5) | 2,530 (12.2) | 2,239.14 [2,152.73, 2,328.12] | 2,222.19 [2,132.06, 2,312.31] |
| | | Gay or Lesbian | 49,600 (1.6) | 955 (4.6) | 1,925.40 [1,805.21, 2,051.50] | 2,170.77 [2,025.10, 2,316.43] |
| | | Bisexual | 50,580 (1.6) | 1,325 (6.4) | 2,619.61 [2,480.44, 2,764.56] | 2,386.80 [2,249.32, 2,524.29] |
| | | Other | 12,805 (0.4) | 250 (1.2) | 1,952.36 [1,717.83, 2,209.98] | 1,925.44 [1,664.46, 2,186.43] |
| | Class 7 (Routine occupations) | Total | 3,136,005 (100.0) | 19,990 (100.0) | 637.44 [628.63, 646.33] | 932.02 [916.67, 947.37] |
| | | Heterosexual | 3,041,835 (97.0) | 18,055 (90.3) | 593.56 [584.93, 602.28] | 856.53 [841.41, 871.65] |
| | | LGB+ | 94,170 (3.0) | 1,935 (9.7) | 2,054.79 [1,964.25, 2,148.44] | 2,064.59 [1,970.31, 2,158.88] |
| | | Gay or Lesbian | 38,485 (1.2) | 725 (3.6) | 1,883.85 [1,749.20, 2,026.12] | 2,140.43 [1,977.92, 2,302.95] |
| | | Bisexual | 42,245 (1.3) | 1,015 (5.1) | 2,402.65 [2,257.10, 2,555.13] | 2,193.13 [2,051.62, 2,334.65] |
| | | Other | 13,440 (0.4) | 195 (1.0) | 1,450.89 [1,254.39, 1,669.45] | 1,502.21 [1,276.49, 1,727.93] |
| | Class 8 (Never worked and long-term unemployed) | Total | 2,067,250 (100.0) | 20,395 (100.0) | 986.58 [973.08, 1,000.21] | 1,564.08 [1,540.00, 1,588.15] |
| | | Heterosexual | 1,998,050 (96.7) | 18,175 (89.1) | 909.64 [896.46, 922.96] | 1,435.95 [1,412.14, 1,459.77] |
| | | LGB+ | 69,205 (3.3) | 2,220 (10.9) | 3,207.86 [3,075.79, 3,344.14] | 3,243.52 [3,107.64, 3,379.40] |
| | | Gay or Lesbian | 24,810 (1.2) | 845 (4.1) | 3,405.88 [3,180.08, 3,643.49] | 3,961.43 [3,689.73, 4,233.12] |
| | | Bisexual | 31,205 (1.5) | 1,155 (5.7) | 3,701.33 [3,490.92, 3,921.10] | 3,424.63 [3,220.41, 3,628.84] |
| | | Other | 13,185 (0.6) | 220 (1.1) | 1,668.56 [1,455.35, 1,904.23] | 1,726.00 [1,495.35, 1,956.66] |
| | Full-time students | Total | 1,398,985 (100.0) | 13,835 (100.0) | 988.93 [972.52, 1,005.55] | 602.51 [581.27, 623.74] |
| | | Heterosexual | 1,261,455 (90.2) | 10,435 (75.4) | 827.22 [811.42, 843.25] | 516.77 [495.88, 537.67] |
| | | LGB+ | 137,530 (9.8) | 3,400 (24.6) | 2,472.19 [2,389.78, 2,556.71] | 1,437.37 [1,287.17, 1,587.58] |
| | | Gay or Lesbian | 37,345 (2.7) | 940 (6.8) | 2,517.07 [2,358.71, 2,683.26] | 1,529.17 [1,266.75, 1,791.59] |
| | | Bisexual | 81,570 (5.8) | 2,020 (14.6) | 2,476.40 [2,369.57, 2,586.80] | 1,295.63 [1,153.42, 1,437.84] |
| | | Other | 18,615 (1.3) | 440 (3.2) | 2,363.69 [2,147.96, 2,595.21] | 1,440.25 [1,087.44, 1,793.06] |
| Household NS-SEC | Class 1 (Higher managerial, administrative and professional occupations) | Total | 7,224,420 (100.0) | 17,125 (100.0) | 237.04 [233.51, 240.62] | 347.86 [341.36, 354.36] |
| | | Heterosexual | 6,936,510 (96.0) | 14,925 (87.2) | 215.17 [211.73, 218.65] | 304.18 [297.91, 310.46] |
| | | LGB+ | 287,910 (4.0) | 2,205 (12.9) | 765.86 [734.23, 798.51] | 838.85 [803.05, 874.66] |
| | | Gay or Lesbian | 157,010 (2.2) | 840 (4.9) | 535.00 [499.43, 572.43] | 760.87 [701.59, 820.14] |
| | | Bisexual | 104,825 (1.5) | 1,100 (6.4) | 1,049.37 [988.26, 1,113.26] | 935.12 [877.53, 992.71] |
| | | Other | 26,075 (0.4) | 265 (1.5) | 1,016.30 [897.61, 1,146.32] | 916.30 [802.87, 1,029.72] |

| | | | | | | |
|---|---|---|---|---|---|---|
| | Class 2 (Lower managerial, administrative and professional occupations) | Total | 7,994,445 (100.0) | 28,115 (100.0) | 351.68 [347.58, 355.82] | 495.67 [488.81, 502.54] |
| | | Heterosexual | 7,669,120 (95.9) | 24,560 (87.4) | 320.25 [316.25, 324.28] | 441.21 [434.52, 447.90] |
| | | LGB+ | 325,325 (4.1) | 3,550 (12.6) | 1,091.22 [1,055.61, 1,127.72] | 1,156.13 [1,117.75, 1,194.51] |
| | | Gay or Lesbian | 173,375 (2.2) | 1,445 (5.1) | 833.45 [791.03, 877.56] | 1,073.11 [1,011.96, 1,134.26] |
| | | Bisexual | 121,205 (1.5) | 1,720 (6.1) | 1,419.08 [1,352.80, 1,487.77] | 1,272.50 [1,210.30, 1,334.71] |
| | | Other | 30,745 (0.4) | 390 (1.4) | 1,268.50 [1,145.71, 1,400.86] | 1,200.15 [1,078.23, 1,322.06] |
| | Class 3 (Intermediate occupations) | Total | 3,551,725 (100.0) | 16,550 (100.0) | 465.97 [458.90, 473.12] | 667.65 [656.02, 679.29] |
| | | Heterosexual | 3,443,335 (96.9) | 14,690 (88.8) | 426.62 [419.75, 433.58] | 602.59 [591.22, 613.96] |
| | | LGB+ | 108,390 (3.1) | 1,860 (11.2) | 1,716.03 [1,638.92, 1,795.82] | 1,662.10 [1,586.30, 1,737.89] |
| | | Gay or Lesbian | 51,290 (1.4) | 775 (4.7) | 1,511.02 [1,406.49, 1,621.25] | 1,680.08 [1,559.95, 1,800.22] |
| | | Bisexual | 45,360 (1.3) | 900 (5.4) | 1,984.13 [1,856.60, 2,118.11] | 1,730.39 [1,611.69, 1,849.09] |
| | | Other | 11,740 (0.3) | 180 (1.1) | 1,533.22 [1,317.41, 1,774.30] | 1,403.61 [1,197.17, 1,610.05] |
| | Class 4 (Small employers and own account workers) | Total | 2,794,170 (100.0) | 12,815 (100.0) | 458.63 [450.73, 466.64] | 644.90 [631.15, 658.64] |
| | | Heterosexual | 2,722,625 (97.4) | 11,935 (93.1) | 438.36 [430.53, 446.30] | 610.78 [597.12, 624.44] |
| | | LGB+ | 71,545 (2.6) | 880 (6.9) | 1,230.00 [1,150.06, 1,314.02] | 1,347.93 [1,255.07, 1,440.78] |
| | | Gay or Lesbian | 30,595 (1.1) | 335 (2.6) | 1,094.95 [980.82, 1,218.71] | 1,454.14 [1,283.44, 1,624.83] |
| | | Bisexual | 28,725 (1.0) | 430 (3.4) | 1,496.95 [1,358.79, 1,645.36] | 1,450.30 [1,308.93, 1,591.67] |
| | | Other | 12,225 (0.4) | 110 (0.9) | 899.80 [739.52, 1,084.50] | 970.24 [785.76, 1,154.73] |
| | Class 5 (Lower supervisory and technical occupations) | Total | 1,326,605 (100.0) | 7,555 (100.0) | 569.50 [556.73, 582.49] | 770.17 [750.24, 790.10] |
| | | Heterosexual | 1,284,420 (96.8) | 6,790 (89.9) | 528.64 [516.14, 541.37] | 706.03 [686.46, 725.61] |
| | | LGB+ | 42,185 (3.2) | 765 (10.1) | 1,813.44 [1,687.20, 1,946.63] | 1,808.66 [1,680.30, 1,937.03] |
| | | Gay or Lesbian | 18,910 (1.4) | 315 (4.2) | 1,665.79 [1,486.89, 1,860.28] | 1,890.78 [1,674.02, 2,107.54] |
| | | Bisexual | 18,045 (1.4) | 400 (5.3) | 2,216.68 [2,004.75, 2,444.92] | 1,996.98 [1,795.08, 2,198.88] |
| | | Other | 5,230 (0.4) | 50 (0.7) | 956.02 [709.58, 1,260.40] | 998.40 [741.92, 1,315.17] |
| | Class 6 (Semi-routine occupations) | Total | 2,258,230 (100.0) | 18,110 (100.0) | 801.96 [790.32, 813.72] | 1,133.52 [1,115.16, 1,151.87] |
| | | Heterosexual | 2,175,160 (96.3) | 15,880 (87.7) | 730.06 [718.75, 741.51] | 1,022.19 [1,004.19, 1,040.19] |
| | | LGB+ | 83,070 (3.7) | 2,230 (12.3) | 2,684.48 [2,574.21, 2,798.27] | 2,591.71 [2,483.52, 2,699.90] |
| | | Gay or Lesbian | 34,365 (1.5) | 855 (4.7) | 2,488.00 [2,324.00, 2,660.51] | 2,747.06 [2,561.39, 2,932.73] |
| | | Bisexual | 38,565 (1.7) | 1,155 (6.4) | 2,994.94 [2,824.69, 3,172.78] | 2,670.60 [2,510.01, 2,831.19] |
| | | Other | 10,140 (0.4) | 220 (1.2) | 2,169.63 [1,892.38, 2,476.06] | 2,087.83 [1,811.18, 2,364.49] |
| | Class 7 (Routine occupations) | Total | 1,663,240 (100.0) | 13,945 (100.0) | 838.42 [824.57, 852.46] | 1,156.13 [1,134.23, 1,178.03] |
| | | Heterosexual | 1,605,255 (96.5) | 12,450 (89.3) | 775.58 [762.01, 789.32] | 1,058.49 [1,036.91, 1,080.08] |
| | | LGB+ | 57,990 (3.5) | 1,495 (10.7) | 2,578.03 [2,448.99, 2,712.11] | 2,567.62 [2,436.49, 2,698.75] |
| | | Gay or Lesbian | 21,570 (1.3) | 520 (3.7) | 2,410.76 [2,207.98, 2,627.15] | 2,718.05 [2,478.67, 2,957.43] |
| | | Bisexual | 27,295 (1.6) | 825 (5.9) | 3,022.53 [2,819.77, 3,236.02] | 2,767.50 [2,573.05, 2,961.96] |
| | | Other | 9,120 (0.5) | 150 (1.1) | 1,644.74 [1,392.06, 1,930.01] | 1,654.29 [1,387.75, 1,920.84] |
| | Class 8 (Never worked and long-term unemployed) | Total | 1,178,230 (100.0) | 14,230 (100.0) | 1,207.74 [1,187.98, 1,227.75] | 2,169.63 [2,128.69, 2,210.57] |
| | | Heterosexual | 1,138,610 (96.6) | 12,665 (89.0) | 1,112.32 [1,093.03, 1,131.86] | 1,990.25 [1,949.75, 2,030.75] |
| | | LGB+ | 39,625 (3.4) | 1,565 (11.0) | 3,949.53 [3,756.25, 4,150.17] | 4,334.58 [4,118.49, 4,550.68] |
| | | Gay or Lesbian | 15,185 (1.3) | 610 (4.3) | 4,017.12 [3,704.62, 4,348.94] | 5,387.84 [4,939.72, 5,835.96] |
| | | Bisexual | 17,290 (1.5) | 815 (5.7) | 4,713.71 [4,395.60, 5,048.75] | 4,539.61 [4,223.98, 4,855.24] |
| | | Other | 7,150 (0.6) | 140 (1.0) | 1,958.04 [1,647.14, 2,310.57] | 2,200.70 [1,831.66, 2,569.74] |
| | Full-time students | Total | 348,600 (100.0) | 3,155 (100.0) | 905.05 [873.74, 937.19] | 775.67 [733.17, 818.17] |
| | | Heterosexual | 295,750 (84.8) | 2,050 (65.0) | 693.15 [663.47, 723.82] | 651.52 [609.80, 693.24] |
| | | LGB+ | 52,845 (15.2) | 1,105 (35.0) | 2,091.02 [1,969.53, 2,218.04] | 1,722.13 [1,450.91, 1,993.34] |

| | | | | | | |
|---|---|---|---|---|---|---|
| | | Gay or Lesbian | 13,075 (3.8) | 275 (8.7) | 2,103.25 [1,861.99, 2,367.10] | 1,891.87 [1,408.05, 2,375.68] |
| | | Bisexual | 33,185 (9.5) | 695 (22.0) | 2,094.32 [1,941.49, 2,255.98] | 1,520.48 [1,267.94, 1,773.02] |
| | | Other | 6,585 (1.9) | 135 (4.3) | 2,050.11 [1,718.89, 2,426.56] | 1,744.05 [962.85, 2,525.25] |
| Lone person household indicator | Lives alone | Total | 4,600,305 (100.0) | 35,250 (100.0) | 766.25 [758.28, 774.30] | 1,767.07 [1,733.39, 1,800.74] |
| | | Heterosexual | 4,363,790 (94.9) | 30,480 (86.5) | 698.48 [690.66, 706.36] | 1,542.84 [1,509.40, 1,576.28] |
| | | LGB+ | 236,515 (5.1) | 4,770 (13.5) | 2,016.79 [1,959.95, 2,074.85] | 3,151.90 [3,033.58, 3,270.22] |
| | | Gay or Lesbian | 134,830 (2.9) | 2,115 (6.0) | 1,568.64 [1,502.49, 1,636.95] | 3,192.88 [2,970.92, 3,414.83] |
| | | Bisexual | 78,515 (1.7) | 2,175 (6.2) | 2,770.17 [2,654.96, 2,889.09] | 3,423.13 [3,257.90, 3,588.35] |
| | | Other | 23,170 (0.5) | 475 (1.3) | 2,050.06 [1,869.83, 2,242.99] | 2,665.38 [2,355.67, 2,975.09] |
| | Does not live alone | Total | 23,739,360 (100.0) | 96,350 (100.0) | 405.87 [403.31, 408.44] | 554.94 [551.00, 558.87] |
| | | Heterosexual | 22,906,995 (96.5) | 85,470 (88.7) | 373.12 [370.62, 375.63] | 504.74 [500.87, 508.61] |
| | | LGB+ | 832,365 (3.5) | 10,885 (11.3) | 1,307.72 [1,283.27, 1,332.52] | 1,204.42 [1,181.64, 1,227.21] |
| | | Gay or Lesbian | 380,540 (1.6) | 3,850 (4.0) | 1,011.72 [980.01, 1,044.19] | 1,144.89 [1,108.01, 1,181.78] |
| | | Bisexual | 355,985 (1.5) | 5,870 (6.1) | 1,648.95 [1,607.03, 1,691.68] | 1,340.97 [1,303.59, 1,378.35] |
| | | Other | 95,840 (0.4) | 1,165 (1.2) | 1,215.57 [1,146.76, 1,287.43] | 1,049.77 [988.14, 1,111.40] |
| Household size | One person in household | Total | 4,600,305 (100.0) | 35,250 (100.0) | 766.25 [758.28, 774.30] | 1,767.07 [1,733.39, 1,800.74] |
| | | Heterosexual | 4,363,790 (94.9) | 30,480 (86.5) | 698.48 [690.66, 706.36] | 1,542.84 [1,509.40, 1,576.28] |
| | | LGB+ | 236,515 (5.1) | 4,770 (13.5) | 2,016.79 [1,959.95, 2,074.85] | 3,151.90 [3,033.58, 3,270.22] |
| | | Gay or Lesbian | 134,830 (2.9) | 2,115 (6.0) | 1,568.64 [1,502.49, 1,636.95] | 3,192.88 [2,970.92, 3,414.83] |
| | | Bisexual | 78,515 (1.7) | 2,175 (6.2) | 2,770.17 [2,654.96, 2,889.09] | 3,423.13 [3,257.90, 3,588.35] |
| | | Other | 23,170 (0.5) | 475 (1.3) | 2,050.06 [1,869.83, 2,242.99] | 2,665.38 [2,355.67, 2,975.09] |
| | Two people in household | Total | 10,142,025 (100.0) | 36,480 (100.0) | 359.69 [356.01, 363.40] | 684.10 [674.15, 694.05] |
| | | Heterosexual | 9,726,660 (95.9) | 32,095 (88.0) | 329.97 [326.37, 333.60] | 614.10 [604.19, 624.01] |
| | | LGB+ | 415,365 (4.1) | 4,385 (12.0) | 1,055.70 [1,024.68, 1,087.42] | 1,273.28 [1,231.47, 1,315.09] |
| | | Gay or Lesbian | 239,535 (2.4) | 1,830 (5.0) | 763.98 [729.37, 799.80] | 1,206.62 [1,132.73, 1,280.52] |
| | | Bisexual | 141,265 (1.4) | 2,140 (5.9) | 1,514.88 [1,451.37, 1,580.46] | 1,455.60 [1,388.85, 1,522.35] |
| | | Other | 34,560 (0.3) | 420 (1.2) | 1,215.28 [1,101.82, 1,337.25] | 1,283.06 [1,151.28, 1,414.84] |
| | Three to five people in household | Total | 12,340,900 (100.0) | 53,305 (100.0) | 431.94 [428.28, 435.62] | 530.55 [525.62, 535.48] |
| | | Heterosexual | 11,964,450 (96.9) | 47,495 (89.1) | 396.97 [393.41, 400.55] | 487.20 [482.34, 492.06] |
| | | LGB+ | 376,450 (3.1) | 5,810 (10.9) | 1,543.37 [1,503.93, 1,583.57] | 1,210.13 [1,175.18, 1,245.07] |
| | | Gay or Lesbian | 131,220 (1.1) | 1,840 (3.5) | 1,402.23 [1,338.88, 1,467.79] | 1,181.93 [1,124.51, 1,239.35] |
| | | Bisexual | 192,115 (1.6) | 3,310 (6.2) | 1,722.93 [1,664.73, 1,782.64] | 1,315.00 [1,259.94, 1,370.06] |
| | | Other | 53,115 (0.4) | 660 (1.2) | 1,242.59 [1,149.58, 1,341.11] | 956.29 [879.61, 1,032.97] |
| | Six or more people in household | Total | 1,256,435 (100.0) | 6,565 (100.0) | 522.51 [509.95, 535.31] | 517.14 [503.86, 530.41] |
| | | Heterosexual | 1,215,885 (96.8) | 5,880 (89.6) | 483.60 [471.32, 496.12] | 482.96 [469.83, 496.08] |
| | | LGB+ | 40,550 (3.2) | 685 (10.4) | 1,689.27 [1,565.12, 1,820.65] | 1,194.18 [1,087.00, 1,301.37] |
| | | Gay or Lesbian | 9,785 (0.8) | 180 (2.7) | 1,839.55 [1,580.62, 2,128.80] | 1,287.07 [1,067.33, 1,506.81] |
| | | Bisexual | 22,600 (1.8) | 415 (6.3) | 1,836.28 [1,663.84, 2,021.74] | 1,307.30 [1,150.98, 1,463.62] |
| | | Other | 8,160 (0.6) | 85 (1.3) | 1,041.67 [832.05, 1,288.04] | 841.65 [650.90, 1,065.85] |
| Household tenure | Owned or shared ownership | Total | 18,590,085 (100.0) | 50,600 (100.0) | 272.19 [269.82, 274.57] | 411.94 [407.18, 416.70] |
| | | Heterosexual | 18,092,870 (97.3) | 46,395 (91.7) | 256.43 [254.10, 258.77] | 377.49 [372.81, 382.17] |
| | | LGB+ | 497,215 (2.7) | 4,205 (8.3) | 845.71 [820.34, 871.67] | 931.49 [902.08, 960.90] |
| | | Gay or Lesbian | 284,235 (1.5) | 1,725 (3.4) | 606.89 [578.59, 636.22] | 867.44 [821.03, 913.86] |
| | | Bisexual | 167,880 (0.9) | 1,990 (3.9) | 1,185.37 [1,133.86, 1,238.62] | 1,015.21 [968.33, 1,062.10] |
| | | Other | 45,100 (0.2) | 490 (1.0) | 1,086.47 [992.39, 1,187.07] | 997.42 [906.11, 1,088.74] |

| | | | | | | |
|---|---|---|---|---|---|---|
| | Social rented | Total | 4,048,710 (100.0) | 44,390 (100.0) | 1,096.40 [1,086.22, 1,106.65] | 1,352.60 [1,338.76, 1,366.44] |
| | | Heterosexual | 3,888,600 (96.0) | 39,245 (88.4) | 1,009.23 [999.27, 1,019.27] | 1,232.17 [1,218.61, 1,245.73] |
| | | LGB+ | 160,110 (4.0) | 5,145 (11.6) | 3,213.42 [3,126.20, 3,302.44] | 3,213.51 [3,124.96, 3,302.07] |
| | | Gay or Lesbian | 69,005 (1.7) | 2,020 (4.6) | 2,927.32 [2,801.05, 3,057.83] | 3,273.07 [3,126.77, 3,419.37] |
| | | Bisexual | 71,845 (1.8) | 2,680 (6.0) | 3,730.25 [3,590.35, 3,874.21] | 3,445.69 [3,311.94, 3,579.43] |
| | | Other | 19,260 (0.5) | 445 (1.0) | 2,310.49 [2,100.78, 2,535.47] | 2,347.39 [2,126.71, 2,568.07] |
| | Private rented or living rent free | Total | 5,700,870 (100.0) | 36,615 (100.0) | 642.27 [635.71, 648.88] | 699.37 [691.93, 706.82] |
| | | Heterosexual | 5,289,315 (92.8) | 30,310 (82.8) | 573.04 [566.61, 579.53] | 620.03 [612.72, 627.34] |
| | | LGB+ | 411,550 (7.2) | 6,305 (17.2) | 1,532.01 [1,494.43, 1,570.30] | 1,434.80 [1,396.36, 1,473.23] |
| | | Gay or Lesbian | 162,125 (2.8) | 2,225 (6.1) | 1,372.40 [1,315.96, 1,430.63] | 1,466.17 [1,399.66, 1,532.69] |
| | | Bisexual | 194,775 (3.4) | 3,375 (9.2) | 1,732.77 [1,674.80, 1,792.23] | 1,499.74 [1,439.95, 1,559.53] |
| | | Other | 54,650 (1.0) | 705 (1.9) | 1,290.03 [1,196.55, 1,388.87] | 1,174.99 [1,080.70, 1,269.28] |
| Economic activity | Economically active | Total | 18,496,300 (100.0) | 68,350 (100.0) | 369.53 [366.77, 372.31] | 519.19 [514.57, 523.82] |
| | | Heterosexual | 17,701,905 (95.7) | 60,070 (87.9) | 339.34 [336.63, 342.07] | 468.69 [464.16, 473.23] |
| | | LGB+ | 794,395 (4.3) | 8,280 (12.1) | 1,042.30 [1,019.97, 1,065.00] | 1,126.88 [1,100.90, 1,152.87] |
| | | Gay or Lesbian | 403,140 (2.2) | 3,325 (4.9) | 824.78 [796.98, 853.30] | 1,080.93 [1,038.83, 1,123.04] |
| | | Bisexual | 310,470 (1.7) | 4,125 (6.0) | 1,328.63 [1,288.39, 1,369.81] | 1,209.73 [1,167.51, 1,251.94] |
| | | Other | 80,785 (0.4) | 830 (1.2) | 1,027.42 [958.70, 1,099.76] | 1,062.34 [985.76, 1,138.92] |
| | Economically inactive | Total | 9,843,365 (100.0) | 63,255 (100.0) | 642.62 [637.62, 647.64] | 1,314.63 [1,301.80, 1,327.46] |
| | | Heterosexual | 9,568,880 (97.2) | 55,880 (88.3) | 583.98 [579.14, 588.84] | 1,184.17 [1,171.56, 1,196.77] |
| | | LGB+ | 274,480 (2.8) | 7,375 (11.7) | 2,686.90 [2,625.92, 2,748.93] | 2,970.19 [2,895.99, 3,044.40] |
| | | Gay or Lesbian | 112,230 (1.1) | 2,640 (4.2) | 2,352.31 [2,263.43, 2,443.79] | 3,401.96 [3,259.09, 3,544.83] |
| | | Bisexual | 124,030 (1.3) | 3,920 (6.2) | 3,160.53 [3,062.35, 3,261.04] | 3,093.10 [2,982.72, 3,203.47] |
| | | Other | 38,220 (0.4) | 810 (1.3) | 2,119.31 [1,975.85, 2,270.43] | 2,044.17 [1,893.34, 2,195.00] |
| General health (detailed) | Very good health | Total | 11,788,340 (100.0) | 32,725 (100.0) | 277.60 [274.61, 280.63] | 345.93 [341.96, 349.91] |
| | | Heterosexual | 11,380,650 (96.5) | 29,800 (91.1) | 261.85 [258.88, 264.84] | 326.91 [322.95, 330.87] |
| | | LGB+ | 407,690 (3.5) | 2,925 (8.9) | 717.46 [691.69, 743.94] | 685.86 [660.88, 710.83] |
| | | Gay or Lesbian | 220,835 (1.9) | 1,330 (4.1) | 602.26 [570.32, 635.52] | 721.50 [681.37, 761.62] |
| | | Bisexual | 147,575 (1.3) | 1,375 (4.2) | 931.73 [883.13, 982.31] | 726.43 [684.43, 768.44] |
| | | Other | 39,280 (0.3) | 220 (0.7) | 560.08 [488.51, 639.19] | 515.69 [445.59, 585.78] |
| | Good health | Total | 11,151,785 (100.0) | 45,370 (100.0) | 406.84 [403.11, 410.60] | 690.58 [683.04, 698.11] |
| | | Heterosexual | 10,722,670 (96.2) | 39,875 (87.9) | 371.88 [368.23, 375.54] | 627.75 [620.21, 635.29] |
| | | LGB+ | 429,120 (3.8) | 5,495 (12.1) | 1,280.53 [1,246.89, 1,314.84] | 1,258.63 [1,225.32, 1,291.93] |
| | | Gay or Lesbian | 199,055 (1.8) | 2,050 (4.5) | 1,029.87 [985.76, 1,075.43] | 1,310.12 [1,250.78, 1,369.46] |
| | | Bisexual | 182,550 (1.6) | 2,930 (6.5) | 1,605.04 [1,547.44, 1,664.23] | 1,310.46 [1,260.52, 1,360.40] |
| | | Other | 47,515 (0.4) | 515 (1.1) | 1,083.87 [992.27, 1,181.65] | 971.92 [887.16, 1,056.67] |
| | Fair health | Total | 3,948,530 (100.0) | 31,715 (100.0) | 803.21 [794.39, 812.10] | 1,825.24 [1,798.52, 1,851.95] |
| | | Heterosexual | 3,786,175 (95.9) | 27,325 (86.2) | 721.70 [713.17, 730.31] | 1,633.41 [1,605.80, 1,661.02] |
| | | LGB+ | 162,355 (4.1) | 4,390 (13.8) | 2,703.95 [2,624.55, 2,785.14] | 2,887.60 [2,801.78, 2,973.42] |
| | | Gay or Lesbian | 66,805 (1.7) | 1,550 (4.9) | 2,320.19 [2,206.10, 2,438.64] | 3,303.37 [3,128.25, 3,478.49] |
| | | Bisexual | 74,465 (1.9) | 2,345 (7.4) | 3,149.13 [3,022.95, 3,279.23] | 2,915.83 [2,795.62, 3,036.03] |
| | | Other | 21,085 (0.5) | 490 (1.5) | 2,323.93 [2,122.69, 2,539.10] | 2,217.71 [2,020.47, 2,414.95] |
| | Bad health | Total | 1,152,955 (100.0) | 16,470 (100.0) | 1,428.50 [1,406.77, 1,450.49] | 3,252.95 [3,176.84, 3,329.06] |
| | | Heterosexual | 1,097,345 (95.2) | 14,295 (86.8) | 1,302.69 [1,281.42, 1,324.22] | 2,940.36 [2,859.21, 3,021.51] |
| | | LGB+ | 55,610 (4.8) | 2,175 (13.2) | 3,911.17 [3,748.51, 4,079.07] | 4,626.61 [4,423.14, 4,830.07] |

| | | | | | | |
|---|---|---|---|---|---|---|
| | | Gay or Lesbian | 22,495 (2.0) | 775 (4.7) | 3,445.21 [3,206.89, 3,696.55] | 5,129.93 [4,711.25, 5,548.62] |
| | | Bisexual | 24,540 (2.1) | 1,090 (6.6) | 4,441.73 [4,181.92, 4,713.45] | 4,686.00 [4,403.73, 4,968.27] |
| | | Other | 8,575 (0.7) | 305 (1.9) | 3,556.85 [3,168.83, 3,979.27] | 3,842.28 [3,404.81, 4,279.74] |
| | Very bad health | Total | 298,055 (100.0) | 5,325 (100.0) | 1,786.58 [1,738.92, 1,835.23] | 4,183.86 [3,996.91, 4,370.82] |
| | | Heterosexual | 283,950 (95.3) | 4,655 (87.4) | 1,639.37 [1,592.61, 1,687.16] | 3,734.70 [3,542.12, 3,927.28] |
| | | LGB+ | 14,105 (4.7) | 670 (12.6) | 4,750.09 [4,397.17, 5,123.79] | 6,377.49 [5,822.76, 6,932.22] |
| | | Gay or Lesbian | 6,180 (2.1) | 260 (4.9) | 4,207.12 [3,711.23, 4,750.82] | 7,029.86 [5,921.35, 8,138.37] |
| | | Bisexual | 5,370 (1.8) | 305 (5.7) | 5,679.70 [5,060.10, 6,354.24] | 6,625.14 [5,820.49, 7,429.80] |
| | | Other | 2,555 (0.9) | 105 (2.0) | 4,109.59 [3,361.24, 4,974.91] | 5,032.75 [3,973.77, 6,091.72] |
| General health (aggregated) | Good health | Total | 22,940,125 (100.0) | 78,095 (100.0) | 340.43 [338.05, 342.83] | 475.84 [472.14, 479.54] |
| | | Heterosexual | 22,103,315 (96.4) | 69,675 (89.2) | 315.22 [312.89, 317.57] | 438.53 [434.87, 442.19] |
| | | LGB+ | 836,810 (3.6) | 8,420 (10.8) | 1,006.20 [984.82, 1,027.93] | 975.33 [954.47, 996.18] |
| | | Gay or Lesbian | 419,890 (1.8) | 3,380 (4.3) | 804.97 [778.06, 832.58] | 985.33 [950.79, 1,019.87] |
| | | Bisexual | 330,125 (1.4) | 4,305 (5.5) | 1,304.05 [1,265.38, 1,343.60] | 1,044.78 [1,011.52, 1,078.05] |
| | | Other | 86,795 (0.4) | 735 (0.9) | 846.82 [786.70, 910.32] | 763.68 [707.70, 819.67] |
| | Not good health | Total | 5,399,540 (100.0) | 53,505 (100.0) | 990.92 [982.54, 999.35] | 2,182.74 [2,157.07, 2,208.41] |
| | | Heterosexual | 5,167,470 (95.7) | 46,275 (86.5) | 895.51 [887.37, 903.70] | 1,951.16 [1,924.57, 1,977.74] |
| | | LGB+ | 232,070 (4.3) | 7,230 (13.5) | 3,115.44 [3,044.04, 3,188.09] | 3,448.98 [3,368.61, 3,529.35] |
| | | Gay or Lesbian | 95,480 (1.8) | 2,585 (4.8) | 2,707.37 [2,604.00, 2,813.80] | 3,876.88 [3,713.34, 4,040.42] |
| | | Bisexual | 104,375 (1.9) | 3,740 (7.0) | 3,583.23 [3,469.31, 3,699.95] | 3,484.26 [3,371.90, 3,596.62] |
| | | Other | 32,210 (0.6) | 905 (1.7) | 2,809.69 [2,629.59, 2,998.87] | 2,806.37 [2,623.14, 2,989.60] |
| Disability | Day-to-day activities limited a lot | Total | 1,904,385 (100.0) | 30,355 (100.0) | 1,593.95 [1,576.07, 1,611.99] | 3,395.89 [3,345.58, 3,446.19] |
| | | Heterosexual | 1,807,315 (94.9) | 25,710 (84.7) | 1,422.55 [1,405.22, 1,440.05] | 3,024.14 [2,971.81, 3,076.48] |
| | | LGB+ | 97,075 (5.1) | 4,645 (15.3) | 4,784.96 [4,648.33, 4,924.58] | 5,187.21 [5,036.66, 5,337.76] |
| | | Gay or Lesbian | 39,370 (2.1) | 1,645 (5.4) | 4,178.31 [3,978.81, 4,385.22] | 5,663.27 [5,369.21, 5,957.33] |
| | | Bisexual | 43,105 (2.3) | 2,385 (7.9) | 5,533.00 [5,313.15, 5,759.61] | 5,320.20 [5,105.31, 5,535.09] |
| | | Other | 14,595 (0.8) | 615 (2.0) | 4,213.77 [3,887.28, 4,560.36] | 4,169.00 [3,839.31, 4,498.70] |
| | Day-to-day activities limited a little | Total | 3,184,715 (100.0) | 31,655 (100.0) | 993.97 [983.05, 1,004.98] | 1,803.33 [1,780.38, 1,826.29] |
| | | Heterosexual | 2,970,515 (93.3) | 26,195 (82.8) | 881.83 [871.19, 892.58] | 1,678.11 [1,653.24, 1,702.97] |
| | | LGB+ | 214,195 (6.7) | 5,460 (17.2) | 2,549.08 [2,481.91, 2,617.61] | 2,369.47 [2,305.38, 2,433.56] |
| | | Gay or Lesbian | 76,965 (2.4) | 1,815 (5.7) | 2,358.21 [2,250.96, 2,469.26] | 2,749.78 [2,621.51, 2,878.04] |
| | | Bisexual | 108,705 (3.4) | 3,055 (9.7) | 2,810.36 [2,711.58, 2,911.82] | 2,407.08 [2,313.44, 2,500.71] |
| | | Other | 28,525 (0.9) | 590 (1.9) | 2,068.36 [1,904.81, 2,242.20] | 1,764.19 [1,614.80, 1,913.59] |
| | Day-to-day activities not limited | Total | 23,250,565 (100.0) | 69,590 (100.0) | 299.30 [297.08, 301.54] | 411.07 [407.60, 414.55] |
| | | Heterosexual | 22,492,955 (96.7) | 64,045 (92.0) | 284.73 [282.53, 286.95] | 389.82 [386.34, 393.30] |
| | | LGB+ | 757,610 (3.3) | 5,545 (8.0) | 731.91 [712.77, 751.43] | 747.04 [727.35, 766.73] |
| | | Gay or Lesbian | 399,030 (1.7) | 2,510 (3.6) | 629.03 [604.66, 654.13] | 786.38 [753.79, 818.97] |
| | | Bisexual | 282,690 (1.2) | 2,605 (3.7) | 921.50 [886.45, 957.59] | 775.75 [744.67, 806.82] |
| | | Other | 75,885 (0.3) | 435 (0.6) | 573.24 [520.63, 629.72] | 549.08 [496.94, 601.22] |

Notes: Age-standardised rates are expressed per 100,000 people and standardised to the age distribution of the observed study population identifying with an LGB+ orientation in Census 2021. Cells with the value '-' are not applicable for the given characteristic or category. All calculations are based on estimates rounded to the nearest multiple of five for disclosure control reasons. CI: confidence interval.

**Supplementary Table 15: Sensitivity analysis restricting to people living in private households only, crude and age-standardised rates of suicide per 100,000 people**

| Characteristic | Category | Sexual orientation | Count (%), Individuals | Count (%), Individuals with death by suicide | Crude rate of suicide [95% CI] | Age-standardised rate of suicide [95% CI] |
|---|---|---|---|---|---|---|
| Total | Total | Total | 28,339,665 (100.0) | 7,050 (100.0) | 24.88 [24.30, 25.46] | 24.49 [23.79, 25.19] |
| | | Heterosexual | 27,270,785 (96.2) | 6,510 (92.3) | 23.87 [23.30, 24.46] | 23.11 [22.42, 23.81] |
| | | LGB+ | 1,068,880 (3.8) | 540 (7.7) | 50.52 [46.35, 54.97] | 50.68 [46.41, 54.95] |
| | | Gay or Lesbian | 515,370 (1.8) | 275 (3.9) | 53.36 [47.24, 60.05] | 51.21 [44.90, 57.53] |
| | | Bisexual | 434,500 (1.5) | 200 (2.8) | 46.03 [39.87, 52.87] | 50.86 [43.44, 58.29] |
| | | Other | 119,005 (0.4) | 65 (0.9) | 54.62 [42.15, 69.62] | 54.63 [41.73, 70.16] |
| Age on Census Day (10-year bands) | 16 to 24 years | Total | 2,668,275 (100.0) | 555 (100.0) | 20.80 [19.07, 22.53] | - |
| | | Heterosexual | 2,415,345 (90.5) | 460 (82.9) | 19.04 [17.30, 20.79] | - |
| | | LGB+ | 252,930 (9.5) | 95 (17.1) | 37.56 [30.39, 45.91] | - |
| | 25 to 34 years | Total | 5,018,225 (100.0) | 1,155 (100.0) | 23.02 [21.69, 24.34] | - |
| | | Heterosexual | 4,681,300 (93.3) | 1,005 (87.0) | 21.47 [20.14, 22.80] | - |
| | | LGB+ | 336,930 (6.7) | 150 (13.0) | 44.52 [37.40, 51.64] | - |
| | 35 to 44 years | Total | 4,939,215 (100.0) | 1,320 (100.0) | 26.72 [25.28, 28.17] | - |
| | | Heterosexual | 4,736,350 (95.9) | 1,205 (91.3) | 25.44 [24.01, 26.88] | - |
| | | LGB+ | 202,865 (4.1) | 115 (8.7) | 56.69 [46.33, 67.05] | - |
| | 45 to 54 years | Total | 5,078,155 (100.0) | 1,570 (100.0) | 30.92 [29.39, 32.45] | - |
| | | Heterosexual | 4,937,010 (97.2) | 1,475 (93.9) | 29.88 [28.35, 31.40] | - |
| | | LGB+ | 141,145 (2.8) | 95 (6.1) | 67.31 [54.46, 82.28] | - |
| | 55 to 64 years | Total | 4,784,020 (100.0) | 1,310 (100.0) | 27.38 [25.90, 28.87] | - |
| | | Heterosexual | 4,695,320 (98.1) | 1,250 (95.4) | 26.62 [25.15, 28.10] | - |
| | | LGB+ | 88,700 (1.9) | 60 (4.6) | 67.64 [51.62, 87.07] | - |
| | 65 years and over | Total | 5,851,775 (100.0) | 1,140 (100.0) | 19.48 [18.35, 20.61] | - |
| | | Heterosexual | 5,805,465 (99.2) | 1,115 (97.8) | 19.21 [18.08, 20.33] | - |
| | | LGB+ | 46,310 (0.8) | 25 (2.2) | 53.98 [34.94, 79.69] | - |
| Sex | Female | Total | 15,222,250 (100.0) | 2,005 (100.0) | 13.17 [12.60, 13.76] | 13.41 [12.70, 14.12] |
| | | Heterosexual | 14,641,770 (96.2) | 1,790 (89.3) | 12.23 [11.67, 12.81] | 11.86 [11.18, 12.54] |
| | | LGB+ | 580,480 (3.8) | 215 (10.7) | 37.04 [32.25, 42.33] | 37.32 [32.25, 42.38] |
| | | Gay or Lesbian | 196,160 (1.3) | 75 (3.7) | 38.23 [30.07, 47.93] | 35.83 [27.90, 45.25] |
| | | Bisexual | 309,415 (2.0) | 115 (5.7) | 37.17 [30.69, 44.61] | 37.87 [30.03, 45.71] |
| | | Other | 74,905 (0.5) | 30 (1.5) | 40.05 [27.02, 57.17] | 39.21 [25.88, 56.73] |
| | Male | Total | 13,117,415 (100.0) | 5,050 (100.0) | 38.50 [37.44, 39.58] | 37.82 [36.54, 39.11] |
| | | Heterosexual | 12,629,015 (96.3) | 4,720 (93.5) | 37.37 [36.32, 38.46] | 36.55 [35.25, 37.85] |
| | | LGB+ | 488,400 (3.7) | 325 (6.4) | 66.54 [59.51, 74.19] | 64.01 [56.88, 71.13] |
| | | Gay or Lesbian | 319,210 (2.4) | 205 (4.1) | 64.22 [55.73, 73.64] | 61.03 [51.97, 70.08] |
| | | Bisexual | 125,085 (1.0) | 90 (1.8) | 71.95 [57.86, 88.44] | 71.32 [57.05, 88.01] |
| | | Other | 44,105 (0.3) | 35 (0.7) | 79.36 [55.27, 110.37] | 77.53 [53.52, 108.44] |
| Region | England: North East | Total | 1,314,700 (100.0) | 420 (100.0) | 31.95 [28.96, 35.15] | 36.12 [31.88, 40.36] |
| | | Heterosexual | 1,269,590 (96.6) | 385 (91.7) | 30.32 [27.37, 33.51] | 34.79 [30.44, 39.15] |
| | | LGB+ | 45,105 (3.4) | 30 (7.1) | 66.51 [44.88, 94.95] | 72.05 [49.20, 102.09] |
| | England: North West | Total | 3,622,995 (100.0) | 1,055 (100.0) | 29.12 [27.39, 30.93] | 30.30 [28.10, 32.50] |
| | | Heterosexual | 3,489,035 (96.3) | 955 (90.5) | 27.37 [25.66, 29.16] | 27.59 [25.44, 29.74] |

| | | | | | | |
|---|---|---|---|---|---|---|
| | | LGB+ | 133,960 (3.7) | 100 (9.5) | 74.65 [60.74, 90.79] | 75.33 [60.62, 90.04] |
| | England: Yorkshire and The Humber | Total | 2,594,300 (100.0) | 735 (100.0) | 28.33 [26.32, 30.46] | 28.21 [25.75, 30.67] |
| | | Heterosexual | 2,500,105 (96.4) | 680 (92.5) | 27.20 [25.19, 29.32] | 26.69 [24.22, 29.15] |
| | | LGB+ | 94,195 (3.6) | 55 (7.5) | 58.39 [43.99, 76.00] | 60.02 [45.17, 78.19] |
| | England: East Midlands | Total | 2,260,080 (100.0) | 600 (100.0) | 26.55 [24.47, 28.76] | 26.17 [23.63, 28.70] |
| | | Heterosexual | 2,184,410 (96.7) | 555 (92.5) | 25.41 [23.34, 27.61] | 24.16 [21.69, 26.64] |
| | | LGB+ | 75,670 (3.3) | 45 (7.5) | 59.47 [43.38, 79.57] | 60.43 [44.16, 80.75] |
| | England: West Midlands | Total | 2,658,530 (100.0) | 670 (100.0) | 25.20 [23.33, 27.18] | 24.25 [22.04, 26.45] |
| | | Heterosexual | 2,576,680 (96.9) | 635 (94.8) | 24.64 [22.76, 26.64] | 23.97 [21.72, 26.23] |
| | | LGB+ | 81,855 (3.1) | 30 (4.5) | 36.65 [24.73, 52.32] | 40.56 [27.71, 57.45] |
| | England: East of England | Total | 3,091,550 (100.0) | 665 (100.0) | 21.51 [19.91, 23.21] | 21.31 [19.21, 23.41] |
| | | Heterosexual | 2,995,735 (96.9) | 620 (93.2) | 20.70 [19.10, 22.39] | 19.93 [17.86, 22.00] |
| | | LGB+ | 95,815 (3.1) | 45 (6.8) | 46.97 [34.26, 62.84] | 47.77 [34.96, 63.78] |
| | England: London | Total | 4,294,885 (100.0) | 715 (100.0) | 16.65 [15.45, 17.91] | 16.09 [14.75, 17.44] |
| | | Heterosexual | 4,067,010 (94.7) | 635 (88.8) | 15.61 [14.42, 16.88] | 14.93 [13.59, 16.27] |
| | | LGB+ | 227,875 (5.3) | 80 (11.2) | 35.11 [27.84, 43.69] | 34.73 [27.38, 43.42] |
| | England: South East | Total | 4,584,145 (100.0) | 1,155 (100.0) | 25.20 [23.76, 26.69] | 24.36 [22.54, 26.18] |
| | | Heterosexual | 4,418,790 (96.4) | 1,075 (93.1) | 24.33 [22.90, 25.83] | 23.13 [21.29, 24.98] |
| | | LGB+ | 165,355 (3.6) | 80 (6.9) | 48.38 [38.36, 60.21] | 47.53 [37.59, 59.28] |
| | England: South West | Total | 2,722,320 (100.0) | 730 (100.0) | 26.82 [24.91, 28.83] | 26.53 [24.09, 28.98] |
| | | Heterosexual | 2,619,105 (96.2) | 685 (93.8) | 26.15 [24.23, 28.19] | 25.75 [23.23, 28.27] |
| | | LGB+ | 103,215 (3.8) | 45 (6.2) | 43.60 [31.80, 58.34] | 46.23 [33.82, 61.73] |
| | Wales | Total | 1,196,160 (100.0) | 310 (100.0) | 25.92 [23.11, 28.97] | 25.04 [21.69, 28.38] |
| | | Heterosexual | 1,150,330 (96.2) | 285 (91.9) | 24.78 [21.98, 27.83] | 23.37 [20.06, 26.69] |
| | | LGB+ | 45,835 (3.8) | 25 (8.1) | 54.54 [35.30, 80.52] | 59.19 [38.76, 86.77] |
| Ethnic group | Asian, Asian British or Asian Welsh | Total | 2,333,750 (100.0) | 280 (100.0) | 12.00 [10.63, 13.49] | 11.82 [10.32, 13.33] |
| | | Heterosexual | 2,279,735 (97.7) | 265 (94.6) | 11.62 [10.27, 13.11] | 11.46 [9.97, 12.96] |
| | | LGB+ | 54,010 (2.3) | 15 (5.4) | 27.77 [15.54, 45.81] | 27.93 [14.19, 48.19] |
| | Black, Black British, Black Welsh, Caribbean or African | Total | 1,017,745 (100.0) | 125 (100.0) | 12.28 [10.22, 14.63] | 14.87 [11.98, 17.77] |
| | | Heterosexual | 993,825 (97.6) | 110 (88.0) | 11.07 [9.10, 13.34] | 13.07 [10.34, 15.80] |
| | | LGB+ | 23,915 (2.3) | 15 (12.0) | 62.72 [35.11, 103.45] | 64.75 [35.71, 107.56] |
| | Mixed or Multiple ethnic groups | Total | 520,740 (100.0) | 160 (100.0) | 30.73 [26.15, 35.87] | 30.82 [25.96, 35.67] |
| | | Heterosexual | 478,925 (92.0) | 135 (84.4) | 28.19 [23.63, 33.36] | 28.47 [23.56, 33.38] |
| | | LGB+ | 41,815 (8.0) | 25 (15.6) | 59.79 [38.69, 88.26] | 64.26 [39.17, 98.11] |
| | White | Total | 23,897,995 (100.0) | 6,420 (100.0) | 26.86 [26.21, 27.53] | 26.88 [26.06, 27.71] |
| | | Heterosexual | 22,971,715 (96.1) | 5,935 (92.4) | 25.84 [25.18, 26.50] | 25.49 [24.66, 26.32] |
| | | LGB+ | 926,280 (3.9) | 485 (7.6) | 52.36 [47.80, 57.23] | 52.37 [47.71, 57.04] |
| | Other ethnic group | Total | 569,435 (100.0) | 65 (100.0) | 11.41 [8.81, 14.55] | 11.65 [8.68, 15.22] |
| | | Heterosexual | 546,580 (96.0) | 60 (92.3) | 10.98 [8.38, 14.13] | 10.69 [7.88, 14.09] |
| | | LGB+ | 22,855 (4.0) | 5 (7.7) | 21.88 [7.10, 51.05] | 29.91 [8.98, 71.25] |
| Rural/urban classification | Urban | Total | 23,090,095 (100.0) | 5,675 (100.0) | 24.58 [23.94, 25.23] | 24.28 [23.53, 25.03] |
| | | Heterosexual | 22,152,635 (95.9) | 5,205 (91.7) | 23.50 [22.86, 24.14] | 22.89 [22.14, 23.64] |
| | | LGB+ | 937,465 (4.1) | 470 (8.3) | 50.14 [45.70, 54.88] | 50.21 [45.67, 54.76] |
| | | Gay or Lesbian | 448,580 (1.9) | 235 (4.1) | 52.39 [45.90, 59.53] | 49.52 [42.92, 56.11] |
| | | Bisexual | 381,940 (1.7) | 175 (3.1) | 45.82 [39.28, 53.13] | 50.64 [42.67, 58.62] |

| | | | | | | |
|---|---|---|---|---|---|---|
| | | Other | 106,940 (0.5) | 60 (1.1) | 56.11 [42.81, 72.22] | 55.65 [41.92, 72.30] |
| | Rural | Total | 5,249,565 (100.0) | 1,380 (100.0) | 26.29 [24.92, 27.71] | 25.56 [23.64, 27.48] |
| | | Heterosexual | 5,118,155 (97.5) | 1,305 (94.6) | 25.50 [24.13, 26.92] | 24.24 [22.33, 26.16] |
| | | LGB+ | 131,415 (2.5) | 75 (5.4) | 57.07 [44.89, 71.54] | 54.68 [42.52, 69.12] |
| | | Gay or Lesbian | 66,790 (1.3) | 40 (2.9) | 59.89 [42.79, 81.55] | 64.61 [45.22, 89.17] |
| | | Bisexual | 52,560 (1.0) | 25 (1.8) | 47.56 [30.78, 70.21] | 51.39 [32.95, 76.27] |
| | | Other | 12,065 (0.2) | 5 (0.4) | 41.44 [13.46, 96.71] | 45.95 [15.20, 106.69] |
| Country of birth | Born in the UK | Total | 22,921,610 (100.0) | 6,185 (100.0) | 26.98 [26.31, 27.66] | 27.27 [26.44, 28.11] |
| | | Heterosexual | 22,070,020 (96.3) | 5,705 (92.2) | 25.85 [25.18, 26.53] | 25.67 [24.83, 26.50] |
| | | LGB+ | 851,590 (3.7) | 480 (7.8) | 56.37 [51.43, 61.64] | 57.27 [52.15, 62.39] |
| | | Gay or Lesbian | 426,925 (1.9) | 255 (4.1) | 59.73 [52.62, 67.53] | 56.69 [49.43, 63.94] |
| | | Bisexual | 344,360 (1.5) | 175 (2.8) | 50.82 [43.57, 58.93] | 56.78 [47.77, 65.78] |
| | | Other | 80,305 (0.4) | 55 (0.9) | 68.49 [51.60, 89.15] | 72.57 [54.11, 95.15] |
| | Born outside the UK | Total | 5,418,055 (100.0) | 865 (100.0) | 15.97 [14.92, 17.07] | 15.68 [14.44, 16.92] |
| | | Heterosexual | 5,200,765 (96.0) | 805 (93.1) | 15.48 [14.43, 16.59] | 15.04 [13.79, 16.28] |
| | | LGB+ | 217,290 (4.0) | 60 (6.9) | 27.61 [21.07, 35.54] | 29.26 [22.01, 38.06] |
| | | Gay or Lesbian | 88,445 (1.6) | 25 (2.9) | 28.27 [18.29, 41.73] | 30.75 [17.04, 49.26] |
| | | Bisexual | 90,140 (1.7) | 30 (3.5) | 33.28 [22.45, 47.51] | 33.02 [21.32, 48.40] |
| | | Other | 38,700 (0.7) | 10 (1.2) | 25.84 [12.39, 47.52] | 24.49 [10.08, 47.72] |
| English language proficiency | Main language is English | Total | 25,715,175 (100.0) | 6,710 (100.0) | 26.09 [25.47, 26.73] | 26.17 [25.40, 26.94] |
| | | Heterosexual | 24,744,730 (96.2) | 6,185 (92.2) | 25.00 [24.38, 25.63] | 24.66 [23.88, 25.43] |
| | | LGB+ | 970,445 (3.8) | 525 (7.8) | 54.10 [49.57, 58.93] | 54.20 [49.56, 58.84] |
| | | Gay or Lesbian | 483,460 (1.9) | 270 (4.0) | 55.85 [49.38, 62.92] | 53.58 [46.89, 60.28] |
| | | Bisexual | 391,770 (1.5) | 195 (2.9) | 49.77 [43.03, 57.27] | 55.64 [47.38, 63.90] |
| | | Other | 95,215 (0.4) | 60 (0.9) | 63.02 [48.09, 81.11] | 63.82 [48.04, 82.95] |
| | Main language is not English | Total | 2,624,490 (100.0) | 340 (100.0) | 12.95 [11.61, 14.41] | 12.08 [10.64, 13.51] |
| | | Heterosexual | 2,526,055 (96.2) | 325 (95.6) | 12.87 [11.51, 14.34] | 11.85 [10.39, 13.30] |
| | | LGB+ | 98,435 (3.8) | 15 (4.4) | 15.24 [8.53, 25.13] | 18.68 [10.19, 31.19] |
| | | Gay or Lesbian | 31,910 (1.2) | 5 (1.5) | 15.67 [5.09, 36.57] | 18.00 [5.68, 42.34] |
| | | Bisexual | 42,730 (1.6) | 5 (1.5) | 11.70 [3.80, 27.31] | 12.86 [3.51, 31.30] |
| | | Other | 23,790 (0.9) | 5 (1.5) | 21.02 [6.82, 49.05] | 26.42 [8.74, 61.35] |
| Religion | No religion | Total | 10,589,125 (100.0) | 3,465 (100.0) | 32.72 [31.64, 33.83] | 31.05 [29.93, 32.18] |
| | | Heterosexual | 9,914,285 (93.6) | 3,100 (89.5) | 31.27 [30.18, 32.39] | 29.34 [28.20, 30.48] |
| | | LGB+ | 674,845 (6.4) | 360 (10.4) | 53.35 [47.98, 59.15] | 56.67 [50.71, 62.63] |
| | Christian | Total | 14,170,955 (100.0) | 2,925 (100.0) | 20.64 [19.90, 21.40] | 20.41 [19.32, 21.51] |
| | | Heterosexual | 13,900,760 (98.1) | 2,800 (95.7) | 20.14 [19.40, 20.90] | 19.67 [18.58, 20.77] |
| | | LGB+ | 270,195 (1.9) | 125 (4.3) | 46.26 [38.51, 55.12] | 43.27 [35.03, 51.51] |
| | Buddhist | Total | 145,815 (100.0) | 50 (100.0) | 34.29 [25.45, 45.21] | 29.07 [20.01, 40.26] |
| | | Heterosexual | 133,095 (91.3) | 45 (90.0) | 33.81 [24.66, 45.24] | 24.45 [16.01, 35.00] |
| | | LGB+ | 12,720 (8.7) | 10 (20.0) | 78.62 [37.70, 144.58] | 65.41 [28.15, 125.49] |
| | Hindu | Total | 500,755 (100.0) | 65 (100.0) | 12.98 [10.02, 16.54] | 13.05 [9.56, 17.24] |
| | | Heterosexual | 490,280 (97.9) | 65 (100.0) | 13.26 [10.23, 16.90] | 12.36 [8.99, 16.43] |
| | | LGB+ | 10,475 (2.1) | 5 (7.7) | 47.73 [15.50, 111.39] | 33.38 [2.63, 94.11] |
| | Jewish | Total | 124,415 (100.0) | 30 (100.0) | 24.11 [16.27, 34.42] | 19.01 [11.67, 28.65] |
| | | Heterosexual | 117,720 (94.6) | 25 (83.3) | 21.24 [13.74, 31.35] | 16.75 [9.95, 25.92] |

| | | | | | | |
|---|---|---|---|---|---|---|
| | | LGB+ | 6,700 (5.4) | 5 (16.7) | 74.63 [24.23, 174.15] | 43.11 [5.25, 117.88] |
| | Muslim | Total | 1,495,785 (100.0) | 85 (100.0) | 5.68 [4.54, 7.03] | 5.68 [4.46, 7.11] |
| | | Heterosexual | 1,469,860 (98.3) | 80 (94.1) | 5.44 [4.32, 6.77] | 5.49 [4.29, 6.91] |
| | | LGB+ | 25,930 (1.7) | 5 (5.9) | 19.28 [6.26, 45.00] | 15.75 [2.18, 42.54] |
| | Sikh | Total | 248,420 (100.0) | 70 (100.0) | 28.18 [21.97, 35.60] | 25.97 [19.32, 33.91] |
| | | Heterosexual | 244,010 (98.2) | 65 (92.9) | 26.64 [20.56, 33.95] | 24.74 [18.27, 32.52] |
| | | LGB+ | 4,410 (1.8) | 5 (7.1) | 113.38 [36.81, 264.59] | 90.26 [12.67, 243.48] |
| | Other religion | Total | 198,175 (100.0) | 90 (100.0) | 45.41 [36.52, 55.82] | 46.95 [36.24, 59.49] |
| | | Heterosexual | 163,850 (82.7) | 75 (83.3) | 45.77 [36.00, 57.38] | 44.08 [31.52, 59.00] |
| | | LGB+ | 34,325 (17.3) | 20 (22.2) | 58.27 [35.59, 89.99] | 51.22 [30.26, 80.54] |
| | Not answered | Total | 866,220 (100.0) | 275 (100.0) | 31.75 [28.11, 35.73] | 31.43 [26.86, 36.00] |
| | | Heterosexual | 836,935 (96.6) | 260 (94.5) | 31.07 [27.40, 35.08] | 30.57 [25.91, 35.24] |
| | | LGB+ | 29,285 (3.4) | 15 (5.5) | 51.22 [28.67, 84.48] | 51.16 [28.53, 84.53] |
| IMD/WIMD decile group | 1 (most deprived) | Total | 2,360,410 (100.0) | 765 (100.0) | 32.41 [30.15, 34.79] | 32.15 [29.66, 34.65] |
| | | Heterosexual | 2,252,860 (95.4) | 695 (90.8) | 30.85 [28.60, 33.23] | 30.43 [27.92, 32.93] |
| | | LGB+ | 107,550 (4.6) | 70 (9.2) | 65.09 [50.74, 82.23] | 66.54 [51.79, 84.16] |
| | | Gay or Lesbian | 48,485 (2.1) | 35 (4.6) | 72.19 [50.28, 100.40] | 65.91 [44.96, 92.89] |
| | | Bisexual | 45,390 (1.9) | 30 (3.9) | 66.09 [44.59, 94.35] | 80.39 [53.66, 115.53] |
| | | Other | 13,675 (0.6) | 5 (0.7) | 36.56 [11.87, 85.33] | 30.77 [8.85, 74.07] |
| | 2 | Total | 2,649,785 (100.0) | 655 (100.0) | 24.72 [22.86, 26.69] | 24.36 [22.27, 26.45] |
| | | Heterosexual | 2,525,935 (95.3) | 600 (91.6) | 23.75 [21.89, 25.73] | 23.00 [20.91, 25.09] |
| | | LGB+ | 123,850 (4.7) | 55 (8.4) | 44.41 [33.45, 57.80] | 43.92 [32.99, 57.28] |
| | | Gay or Lesbian | 57,330 (2.2) | 25 (3.8) | 43.61 [28.22, 64.37] | 42.76 [26.63, 64.53] |
| | | Bisexual | 51,035 (1.9) | 20 (3.1) | 39.19 [23.94, 60.52] | 42.21 [25.75, 65.23] |
| | | Other | 15,485 (0.6) | 10 (1.5) | 64.58 [30.97, 118.76] | 52.38 [22.34, 100.81] |
| | 3 | Total | 2,821,390 (100.0) | 710 (100.0) | 25.16 [23.35, 27.09] | 24.76 [22.69, 26.83] |
| | | Heterosexual | 2,685,000 (95.2) | 630 (88.7) | 23.46 [21.67, 25.37] | 22.62 [20.58, 24.66] |
| | | LGB+ | 136,395 (4.8) | 80 (11.3) | 58.65 [46.51, 73.00] | 56.68 [44.73, 70.81] |
| | | Gay or Lesbian | 64,690 (2.3) | 40 (5.6) | 61.83 [44.17, 84.20] | 60.83 [42.51, 84.03] |
| | | Bisexual | 55,235 (2.0) | 30 (4.2) | 54.31 [36.65, 77.54] | 48.26 [31.15, 70.74] |
| | | Other | 16,470 (0.6) | 10 (1.4) | 60.72 [29.12, 111.66] | 60.86 [28.77, 112.61] |
| | 4 | Total | 2,880,030 (100.0) | 790 (100.0) | 27.43 [25.55, 29.41] | 26.01 [23.88, 28.13] |
| | | Heterosexual | 2,748,740 (95.4) | 725 (91.8) | 26.38 [24.49, 28.37] | 24.87 [22.72, 27.02] |
| | | LGB+ | 131,290 (4.6) | 65 (8.2) | 49.51 [38.21, 63.10] | 48.46 [37.23, 61.96] |
| | | Gay or Lesbian | 63,465 (2.2) | 35 (4.4) | 55.15 [38.41, 76.70] | 51.83 [35.50, 72.87] |
| | | Bisexual | 52,965 (1.8) | 20 (2.5) | 37.76 [23.07, 58.32] | 41.01 [23.91, 64.93] |
| | | Other | 14,865 (0.5) | 5 (0.6) | 33.64 [10.92, 78.50] | 52.06 [22.88, 109.68] |
| | 5 | Total | 2,915,180 (100.0) | 785 (100.0) | 26.93 [25.08, 28.88] | 26.11 [23.89, 28.33] |
| | | Heterosexual | 2,797,960 (96.0) | 730 (93.0) | 26.09 [24.23, 28.05] | 25.06 [22.80, 27.32] |
| | | LGB+ | 117,220 (4.0) | 55 (7.0) | 46.92 [35.35, 61.07] | 47.02 [35.43, 61.19] |
| | | Gay or Lesbian | 56,405 (1.9) | 30 (3.8) | 53.19 [35.88, 75.93] | 47.18 [31.84, 67.35] |
| | | Bisexual | 48,025 (1.6) | 20 (2.5) | 41.64 [25.44, 64.32] | 39.26 [22.44, 62.79] |
| | | Other | 12,790 (0.4) | 5 (0.6) | 39.09 [12.69, 91.23] | 36.50 [12.41, 84.07] |
| | 6 | Total | 2,979,555 (100.0) | 735 (100.0) | 24.67 [22.92, 26.52] | 24.03 [21.89, 26.17] |
| | | Heterosexual | 2,866,330 (96.2) | 675 (91.8) | 23.55 [21.81, 25.39] | 22.54 [20.40, 24.68] |

| | | | | | | |
|---|---|---|---|---|---|---|
| | | LGB+ | 113,225 (3.8) | 60 (8.2) | 52.99 [40.44, 68.21] | 53.28 [40.71, 68.52] |
| | | Gay or Lesbian | 55,740 (1.9) | 30 (4.1) | 53.82 [36.31, 76.83] | 50.50 [33.94, 72.27] |
| | | Bisexual | 45,545 (1.5) | 20 (2.7) | 43.91 [26.82, 67.82] | 57.03 [34.93, 87.95] |
| | | Other | 11,945 (0.4) | 5 (0.7) | 41.86 [13.59, 97.68] | 53.03 [20.93, 116.44] |
| | 7 | Total | 2,941,315 (100.0) | 705 (100.0) | 23.97 [22.23, 25.81] | 22.90 [20.73, 25.08] |
| | | Heterosexual | 2,843,740 (96.7) | 660 (93.6) | 23.21 [21.47, 25.05] | 21.84 [19.64, 24.03] |
| | | LGB+ | 97,570 (3.3) | 50 (7.1) | 51.25 [38.04, 67.56] | 47.96 [35.24, 63.66] |
| | | Gay or Lesbian | 48,480 (1.6) | 20 (2.8) | 41.25 [25.20, 63.71] | 41.90 [25.76, 64.47] |
| | | Bisexual | 39,240 (1.3) | 15 (2.1) | 38.23 [21.39, 63.05] | 50.42 [28.31, 83.03] |
| | | Other | 9,855 (0.3) | 10 (1.4) | 101.47 [48.66, 186.61] | 91.00 [37.73, 176.87] |
| | 8 | Total | 2,952,670 (100.0) | 680 (100.0) | 23.03 [21.33, 24.83] | 22.89 [20.64, 25.14] |
| | | Heterosexual | 2,862,105 (96.9) | 635 (93.4) | 22.19 [20.49, 23.98] | 21.37 [19.15, 23.59] |
| | | LGB+ | 90,570 (3.1) | 45 (6.6) | 49.69 [36.24, 66.48] | 51.46 [37.64, 68.73] |
| | | Gay or Lesbian | 45,690 (1.5) | 25 (3.7) | 54.72 [35.41, 80.77] | 48.01 [29.46, 73.06] |
| | | Bisexual | 36,040 (1.2) | 20 (2.9) | 55.49 [33.90, 85.71] | 54.40 [31.66, 86.21] |
| | | Other | 8,835 (0.3) | 5 (0.7) | 56.59 [18.38, 132.07] | 56.73 [13.68, 141.74] |
| | 9 | Total | 2,942,955 (100.0) | 625 (100.0) | 21.24 [19.60, 22.97] | 21.34 [19.05, 23.62] |
| | | Heterosexual | 2,860,805 (97.2) | 590 (94.4) | 20.62 [18.99, 22.36] | 20.44 [18.11, 22.77] |
| | | LGB+ | 82,150 (2.8) | 35 (5.6) | 42.60 [29.68, 59.25] | 42.91 [29.89, 59.67] |
| | | Gay or Lesbian | 41,210 (1.4) | 20 (3.2) | 48.53 [29.64, 74.95] | 48.76 [27.97, 77.84] |
| | | Bisexual | 32,940 (1.1) | 15 (2.4) | 45.54 [25.49, 75.11] | 49.09 [25.22, 84.29] |
| | | Other | 8,000 (0.3) | 5 (0.8) | 62.50 [20.29, 145.85] | 40.71 [7.26, 106.76] |
| | 10 (least deprived) | Total | 2,896,370 (100.0) | 600 (100.0) | 20.72 [19.09, 22.44] | 19.74 [17.46, 22.02] |
| | | Heterosexual | 2,827,310 (97.6) | 570 (95.0) | 20.16 [18.54, 21.89] | 18.68 [16.40, 20.96] |
| | | LGB+ | 69,055 (2.4) | 30 (5.0) | 43.44 [29.31, 62.02] | 45.69 [30.93, 65.08] |
| | | Gay or Lesbian | 33,875 (1.2) | 15 (2.5) | 44.28 [24.78, 73.03] | 56.03 [31.37, 92.41] |
| | | Bisexual | 28,090 (1.0) | 10 (1.7) | 35.60 [17.07, 65.47] | 42.72 [18.67, 81.49] |
| | | Other | 7,090 (0.2) | 5 (0.8) | 70.52 [22.90, 164.57] | 84.91 [27.43, 198.43] |
| Highest level of qualification (≥25 years) | No qualifications | Total | 4,082,460 (100.0) | 1,180 (100.0) | 28.90 [27.28, 30.60] | 38.38 [35.23, 41.52] |
| | | Heterosexual | 4,016,180 (98.4) | 1,125 (95.3) | 28.01 [26.40, 29.70] | 37.41 [34.22, 40.60] |
| | | LGB+ | 66,280 (1.6) | 50 (4.2) | 75.44 [55.99, 99.45] | 74.84 [55.07, 99.26] |
| | Level 1 (one to four GCSE passes (grade A* to C or grade 4 and above) and any other GCSEs at other grades, or equivalent qualifications) | Total | 2,418,070 (100.0) | 635 (100.0) | 26.26 [24.26, 28.38] | 32.13 [28.82, 35.44] |
| | | Heterosexual | 2,366,905 (97.9) | 605 (95.3) | 25.56 [23.56, 27.68] | 31.33 [28.00, 34.67] |
| | | LGB+ | 51,165 (2.1) | 25 (3.9) | 48.86 [31.62, 72.13] | 54.56 [35.41, 80.41] |
| | Level 2 (five or more GCSE passes (grade A* to C or grade 4 and above) or equivalent qualifications) | Total | 3,077,650 (100.0) | 910 (100.0) | 29.57 [27.68, 31.55] | 31.43 [28.99, 33.88] |
| | | Heterosexual | 2,992,585 (97.2) | 865 (95.1) | 28.90 [27.01, 30.90] | 30.72 [28.24, 33.20] |
| | | LGB+ | 85,070 (2.8) | 45 (4.9) | 52.90 [38.58, 70.78] | 53.34 [38.88, 71.41] |
| | Apprenticeship | Total | 1,302,750 (100.0) | 425 (100.0) | 32.62 [29.60, 35.88] | 33.85 [29.38, 38.31] |
| | | Heterosexual | 1,281,405 (98.4) | 420 (98.8) | 32.78 [29.72, 36.07] | 34.02 [29.45, 38.59] |
| | | LGB+ | 21,345 (1.6) | 5 (1.2) | 23.42 [7.61, 54.67] | 32.09 [13.42, 68.95] |
| | | Total | 4,037,275 (100.0) | 1,035 (100.0) | 25.64 [24.10, 27.25] | 26.11 [24.37, 27.85] |

| | | | | | | |
|---|---|---|---|---|---|---|
| | Level 3 (two or more A Levels or equivalent qualifications) | Heterosexual | 3,900,045 (96.6) | 940 (90.8) | 24.10 [22.59, 25.69] | 24.15 [22.44, 25.86] |
| | | LGB+ | 137,235 (3.4) | 95 (9.2) | 69.22 [56.01, 84.62] | 69.53 [56.19, 85.08] |
| | Level 4 and above (Higher National Certificate, Higher National Diploma, Bachelor's degree, or post-graduate qualifications) | Total | 10,077,510 (100.0) | 2,165 (100.0) | 21.48 [20.59, 22.41] | 20.03 [19.10, 20.95] |
| | | Heterosexual | 9,635,435 (95.6) | 1,955 (90.3) | 20.29 [19.40, 21.21] | 18.59 [17.67, 19.50] |
| | | LGB+ | 442,075 (4.4) | 210 (9.7) | 47.50 [41.30, 54.38] | 48.08 [41.54, 54.62] |
| | Other qualifications | Total | 675,670 (100.0) | 150 (100.0) | 22.20 [18.79, 26.05] | 26.79 [20.93, 32.65] |
| | | Heterosexual | 662,890 (98.1) | 140 (93.3) | 21.12 [17.77, 24.92] | 25.88 [19.98, 31.77] |
| | | LGB+ | 12,780 (1.9) | 10 (6.7) | 78.25 [37.52, 143.90] | 61.42 [24.61, 120.75] |
| NS-SEC | Class 1 (Higher managerial, administrative and professional occupations) | Total | 4,318,935 (100.0) | 895 (100.0) | 20.72 [19.39, 22.13] | 16.39 [14.82, 17.96] |
| | | Heterosexual | 4,152,390 (96.1) | 830 (92.7) | 19.99 [18.65, 21.40] | 15.43 [13.85, 17.01] |
| | | LGB+ | 166,550 (3.9) | 65 (7.3) | 39.03 [30.12, 49.74] | 34.74 [26.25, 44.95] |
| | Class 2 (Lower managerial, administrative and professional occupations) | Total | 6,300,945 (100.0) | 1,170 (100.0) | 18.57 [17.52, 19.66] | 17.09 [15.03, 19.16] |
| | | Heterosexual | 6,036,965 (95.8) | 1,060 (90.6) | 17.56 [16.52, 18.65] | 15.64 [13.71, 17.57] |
| | | LGB+ | 263,980 (4.2) | 105 (9.0) | 39.78 [32.53, 48.15] | 41.35 [27.95, 54.74] |
| | Class 3 (Intermediate occupations) | Total | 3,496,100 (100.0) | 670 (100.0) | 19.16 [17.74, 20.67] | 19.65 [17.58, 21.72] |
| | | Heterosexual | 3,384,320 (96.8) | 615 (91.8) | 18.17 [16.76, 19.67] | 18.56 [16.45, 20.67] |
| | | LGB+ | 111,780 (3.2) | 50 (7.5) | 44.73 [33.20, 58.97] | 45.07 [33.64, 59.18] |
| | Class 4 (Small employers and own account workers) | Total | 2,965,195 (100.0) | 1,080 (100.0) | 36.42 [34.28, 38.66] | 36.68 [32.95, 40.41] |
| | | Heterosexual | 2,895,580 (97.7) | 1,040 (96.3) | 35.92 [33.77, 38.17] | 36.55 [32.71, 40.39] |
| | | LGB+ | 69,615 (2.3) | 40 (3.7) | 57.46 [41.05, 78.24] | 45.90 [30.64, 65.23] |
| | Class 5 (Lower supervisory and technical occupations) | Total | 1,472,395 (100.0) | 445 (100.0) | 30.22 [27.48, 33.17] | 29.26 [25.74, 32.77] |
| | | Heterosexual | 1,429,330 (97.1) | 420 (94.4) | 29.38 [26.64, 32.33] | 28.25 [24.70, 31.81] |
| | | LGB+ | 43,065 (2.9) | 25 (5.6) | 58.05 [37.57, 85.70] | 54.67 [35.57, 80.45] |
| | Class 6 (Semi-routine occupations) | Total | 3,183,850 (100.0) | 785 (100.0) | 24.66 [22.96, 26.44] | 27.00 [24.72, 29.28] |
| | | Heterosexual | 3,070,855 (96.5) | 705 (89.8) | 22.96 [21.29, 24.72] | 24.78 [22.54, 27.01] |
| | | LGB+ | 112,990 (3.5) | 75 (9.6) | 66.38 [52.21, 83.20] | 66.47 [52.03, 83.61] |
| | Class 7 (Routine occupations) | Total | 3,136,005 (100.0) | 1,035 (100.0) | 33.00 [31.02, 35.08] | 36.37 [33.67, 39.07] |
| | | Heterosexual | 3,041,835 (97.0) | 965 (93.2) | 31.72 [29.75, 33.79] | 34.57 [31.87, 37.28] |
| | | LGB+ | 94,170 (3.0) | 75 (7.2) | 79.64 [62.64, 99.83] | 77.00 [60.13, 97.04] |
| | Class 8 (Never worked and long-term unemployed) | Total | 2,067,250 (100.0) | 745 (100.0) | 36.04 [33.50, 38.72] | 43.07 [39.34, 46.80] |
| | | Heterosexual | 1,998,050 (96.7) | 685 (91.9) | 34.28 [31.76, 36.95] | 40.53 [36.81, 44.26] |
| | | LGB+ | 69,205 (3.3) | 60 (8.1) | 86.70 [66.16, 111.60] | 87.34 [66.34, 112.80] |
| | Full-time students | Total | 1,398,985 (100.0) | 230 (100.0) | 16.44 [14.38, 18.71] | 21.76 [16.48, 27.04] |
| | | Heterosexual | 1,261,455 (90.2) | 185 (80.4) | 14.67 [12.63, 16.94] | 19.17 [14.17, 24.17] |
| | | LGB+ | 137,530 (9.8) | 45 (19.6) | 32.72 [23.87, 43.78] | 74.68 [6.79, 159.50] |
| Household NS-SEC | Class 1 (Higher managerial, administrative and professional occupations) | Total | 7,224,420 (100.0) | 1,375 (100.0) | 19.03 [18.04, 20.07] | 17.69 [16.47, 18.91] |
| | | Heterosexual | 6,936,510 (96.0) | 1,265 (92.0) | 18.24 [17.25, 19.27] | 16.52 [15.31, 17.74] |
| | | LGB+ | 287,910 (4.0) | 110 (8.0) | 38.21 [31.40, 46.05] | 37.87 [30.62, 45.11] |
| | Class 2 (Lower managerial, administrative and professional occupations) | Total | 7,994,445 (100.0) | 1,635 (100.0) | 20.45 [19.47, 21.47] | 19.48 [18.30, 20.66] |
| | | Heterosexual | 7,669,120 (95.9) | 1,495 (91.4) | 19.49 [18.52, 20.51] | 18.32 [17.13, 19.51] |
| | | LGB+ | 325,325 (4.1) | 140 (8.6) | 43.03 [36.20, 50.78] | 41.78 [34.82, 48.73] |

| | | | | | | |
|---|---|---|---|---|---|---|
| | Class 3 (Intermediate occupations) | Total | 3,551,725 (100.0) | 810 (100.0) | 22.81 [21.26, 24.43] | 23.77 [21.78, 25.76] |
| | | Heterosexual | 3,443,335 (96.9) | 755 (93.2) | 21.93 [20.39, 23.55] | 22.79 [20.78, 24.79] |
| | | LGB+ | 108,390 (3.1) | 55 (6.8) | 50.74 [38.23, 66.05] | 50.79 [38.12, 66.28] |
| | Class 4 (Small employers and own account workers) | Total | 2,794,170 (100.0) | 920 (100.0) | 32.93 [30.83, 35.12] | 31.44 [28.78, 34.11] |
| | | Heterosexual | 2,722,625 (97.4) | 880 (95.7) | 32.32 [30.22, 34.53] | 30.81 [28.12, 33.49] |
| | | LGB+ | 71,545 (2.6) | 35 (3.8) | 48.92 [34.07, 68.04] | 47.78 [32.70, 67.21] |
| | Class 5 (Lower supervisory and technical occupations) | Total | 1,326,605 (100.0) | 400 (100.0) | 30.15 [27.27, 33.26] | 30.74 [27.12, 34.37] |
| | | Heterosexual | 1,284,420 (96.8) | 375 (93.8) | 29.20 [26.32, 32.31] | 29.29 [25.68, 32.90] |
| | | LGB+ | 42,185 (3.2) | 25 (6.2) | 59.26 [38.35, 87.48] | 62.59 [40.97, 91.77] |
| | Class 6 (Semi-routine occupations) | Total | 2,258,230 (100.0) | 675 (100.0) | 29.89 [27.68, 32.23] | 32.65 [29.77, 35.54] |
| | | Heterosexual | 2,175,160 (96.3) | 615 (91.1) | 28.27 [26.08, 30.60] | 30.39 [27.51, 33.27] |
| | | LGB+ | 83,070 (3.7) | 65 (9.6) | 78.25 [60.39, 99.73] | 76.34 [58.55, 97.75] |
| | Class 7 (Routine occupations) | Total | 1,663,240 (100.0) | 670 (100.0) | 40.28 [37.29, 43.45] | 43.06 [39.22, 46.90] |
| | | Heterosexual | 1,605,255 (96.5) | 615 (91.8) | 38.31 [35.34, 41.46] | 40.45 [36.62, 44.28] |
| | | LGB+ | 57,990 (3.5) | 55 (8.2) | 94.84 [71.45, 123.45] | 96.06 [72.20, 125.22] |
| | Class 8 (Never worked and long-term unemployed) | Total | 1,178,230 (100.0) | 485 (100.0) | 41.16 [37.58, 45.00] | 53.85 [47.82, 59.87] |
| | | Heterosexual | 1,138,610 (96.6) | 445 (91.8) | 39.08 [35.54, 42.89] | 50.04 [44.07, 56.02] |
| | | LGB+ | 39,625 (3.4) | 40 (8.2) | 100.95 [72.12, 137.46] | 105.68 [75.21, 144.27] |
| | Full-time students | Total | 348,600 (100.0) | 80 (100.0) | 22.95 [18.20, 28.56] | 28.28 [18.94, 39.32] |
| | | Heterosexual | 295,750 (84.8) | 60 (75.0) | 20.29 [15.48, 26.11] | 24.94 [16.68, 34.96] |
| | | LGB+ | 52,845 (15.2) | 20 (25.0) | 37.85 [23.12, 58.45] | 97.81 [0.00, 270.93] |
| Lone person household indicator | Lives alone | Total | 4,600,305 (100.0) | 2,555 (100.0) | 55.54 [53.41, 57.74] | 71.60 [66.47, 76.73] |
| | | Heterosexual | 4,363,790 (94.9) | 2,315 (90.6) | 53.05 [50.91, 55.26] | 65.93 [60.89, 70.97] |
| | | LGB+ | 236,515 (5.1) | 245 (9.6) | 103.59 [91.02, 117.40] | 113.50 [94.50, 132.50] |
| | Does not live alone | Total | 23,739,360 (100.0) | 4,495 (100.0) | 18.93 [18.39, 19.50] | 19.05 [18.39, 19.71] |
| | | Heterosexual | 22,906,995 (96.5) | 4,200 (93.4) | 18.34 [17.78, 18.90] | 18.22 [17.55, 18.88] |
| | | LGB+ | 832,365 (3.5) | 300 (6.7) | 36.04 [32.08, 40.36] | 36.54 [32.35, 40.74] |
| Household size | One person in household | Total | 4,600,305 (100.0) | 2,555 (100.0) | 55.54 [53.41, 57.74] | 71.60 [66.47, 76.73] |
| | | Heterosexual | 4,363,790 (94.9) | 2,315 (90.6) | 53.05 [50.91, 55.26] | 65.93 [60.89, 70.97] |
| | | LGB+ | 236,515 (5.1) | 245 (9.6) | 103.59 [91.02, 117.40] | 113.50 [94.50, 132.50] |
| | Two people in household | Total | 10,142,025 (100.0) | 2,105 (100.0) | 20.76 [19.88, 21.66] | 22.36 [20.99, 23.73] |
| | | Heterosexual | 9,726,660 (95.9) | 1,950 (92.6) | 20.05 [19.17, 20.96] | 21.27 [19.86, 22.69] |
| | | LGB+ | 415,365 (4.1) | 160 (7.6) | 38.52 [32.78, 44.97] | 36.18 [30.40, 41.95] |
| | Three to five people in household | Total | 12,340,900 (100.0) | 2,215 (100.0) | 17.95 [17.21, 18.71] | 18.32 [17.46, 19.17] |
| | | Heterosexual | 11,964,450 (96.9) | 2,085 (94.1) | 17.43 [16.69, 18.19] | 17.61 [16.75, 18.47] |
| | | LGB+ | 376,450 (3.1) | 135 (6.1) | 35.86 [30.07, 42.45] | 39.78 [31.30, 48.27] |
| | Six or more people in household | Total | 1,256,435 (100.0) | 175 (100.0) | 13.93 [11.94, 16.15] | 14.11 [11.90, 16.33] |
| | | Heterosexual | 1,215,885 (96.8) | 165 (94.3) | 13.57 [11.58, 15.81] | 13.96 [11.71, 16.20] |
| | | LGB+ | 40,550 (3.2) | 5 (2.9) | 12.33 [4.00, 28.78] | 18.94 [5.87, 44.76] |
| Household tenure | Owned or shared ownership | Total | 18,590,085 (100.0) | 3,820 (100.0) | 20.55 [19.90, 21.21] | 19.44 [18.54, 20.33] |
| | | Heterosexual | 18,092,870 (97.3) | 3,625 (94.9) | 20.04 [19.39, 20.70] | 18.72 [17.81, 19.62] |
| | | LGB+ | 497,215 (2.7) | 195 (5.1) | 39.22 [33.91, 45.13] | 37.52 [32.02, 43.02] |
| | Social rented | Total | 4,048,710 (100.0) | 1,500 (100.0) | 37.05 [35.20, 38.97] | 37.36 [35.22, 39.50] |
| | | Heterosexual | 3,888,600 (96.0) | 1,370 (91.3) | 35.23 [33.39, 37.15] | 35.15 [33.02, 37.28] |
| | | LGB+ | 160,110 (4.0) | 130 (8.7) | 81.19 [67.84, 96.41] | 81.17 [67.11, 95.22] |

| | | | | | | |
|---|---|---|---|---|---|---|
| | Private rented or living rent free | Total | 5,700,870 (100.0) | 1,735 (100.0) | 30.43 [29.02, 31.90] | 29.67 [28.24, 31.10] |
| | | Heterosexual | 5,289,315 (92.8) | 1,515 (87.3) | 28.64 [27.22, 30.12] | 27.54 [26.12, 28.97] |
| | | LGB+ | 411,550 (7.2) | 215 (12.4) | 52.24 [45.49, 59.71] | 60.55 [51.56, 69.54] |
| Economic activity | Economically active | Total | 18,496,300 (100.0) | 4,055 (100.0) | 21.92 [21.25, 22.61] | 21.07 [20.30, 21.85] |
| | | Heterosexual | 17,701,905 (95.7) | 3,710 (91.5) | 20.96 [20.29, 21.64] | 19.95 [19.17, 20.73] |
| | | LGB+ | 794,395 (4.3) | 345 (8.5) | 43.43 [38.97, 48.26] | 43.86 [38.88, 48.83] |
| | | Gay or Lesbian | 403,140 (2.2) | 180 (4.4) | 44.65 [38.36, 51.67] | 44.59 [37.12, 52.06] |
| | | Bisexual | 310,470 (1.7) | 125 (3.1) | 40.26 [33.51, 47.97] | 45.35 [36.34, 54.37] |
| | | Other | 80,785 (0.4) | 40 (1.0) | 49.51 [35.37, 67.42] | 53.07 [36.81, 73.68] |
| | Economically inactive | Total | 9,843,365 (100.0) | 3,000 (100.0) | 30.48 [29.40, 31.59] | 41.88 [39.74, 44.03] |
| | | Heterosexual | 9,568,880 (97.2) | 2,800 (93.3) | 29.26 [28.19, 30.37] | 39.35 [37.20, 41.49] |
| | | LGB+ | 274,480 (2.8) | 200 (6.7) | 72.87 [63.12, 83.69] | 84.10 [71.49, 96.72] |
| | | Gay or Lesbian | 112,230 (1.1) | 100 (3.3) | 89.10 [72.50, 108.37] | 109.34 [84.22, 134.46] |
| | | Bisexual | 124,030 (1.3) | 80 (2.7) | 64.50 [51.14, 80.28] | 74.35 [57.28, 94.51] |
| | | Other | 38,220 (0.4) | 20 (0.7) | 52.33 [31.96, 80.82] | 56.22 [33.70, 87.73] |
| General health (detailed) | Very good health | Total | 11,788,340 (100.0) | 1,860 (100.0) | 15.78 [15.07, 16.51] | 15.44 [14.67, 16.20] |
| | | Heterosexual | 11,380,650 (96.5) | 1,735 (93.3) | 15.25 [14.54, 15.98] | 14.80 [14.04, 15.57] |
| | | LGB+ | 407,690 (3.5) | 125 (6.7) | 30.66 [25.52, 36.53] | 31.38 [25.86, 36.89] |
| | Good health | Total | 11,151,785 (100.0) | 2,575 (100.0) | 23.09 [22.21, 24.00] | 24.60 [23.38, 25.82] |
| | | Heterosexual | 10,722,670 (96.2) | 2,390 (92.8) | 22.29 [21.40, 23.20] | 23.85 [22.59, 25.11] |
| | | LGB+ | 429,120 (3.8) | 180 (7.0) | 41.95 [36.04, 48.54] | 42.63 [36.42, 48.84] |
| | Fair health | Total | 3,948,530 (100.0) | 1,545 (100.0) | 39.13 [37.20, 41.13] | 52.17 [48.22, 56.11] |
| | | Heterosexual | 3,786,175 (95.9) | 1,405 (90.9) | 37.11 [35.19, 39.10] | 48.49 [44.38, 52.61] |
| | | LGB+ | 162,355 (4.1) | 140 (9.1) | 86.23 [72.54, 101.76] | 85.93 [71.46, 100.40] |
| | Bad health | Total | 1,152,955 (100.0) | 780 (100.0) | 67.65 [62.99, 72.57] | 92.52 [81.33, 103.72] |
| | | Heterosexual | 1,097,345 (95.2) | 710 (91.0) | 64.70 [60.03, 69.64] | 86.14 [74.23, 98.06] |
| | | LGB+ | 55,610 (4.8) | 70 (9.0) | 125.88 [98.13, 159.04] | 127.16 [97.34, 162.81] |
| | Very bad health | Total | 298,055 (100.0) | 295 (100.0) | 98.98 [88.00, 110.94] | 143.15 [114.72, 171.59] |
| | | Heterosexual | 283,950 (95.3) | 270 (91.5) | 95.09 [84.08, 107.13] | 133.84 [104.21, 163.46] |
| | | LGB+ | 14,105 (4.7) | 25 (8.5) | 177.24 [114.70, 261.64] | 194.51 [119.47, 295.78] |
| General health (aggregated) | Good health | Total | 22,940,125 (100.0) | 4,435 (100.0) | 19.33 [18.77, 19.91] | 19.22 [18.56, 19.88] |
| | | Heterosexual | 22,103,315 (96.4) | 4,125 (93.0) | 18.66 [18.10, 19.24] | 18.46 [17.79, 19.13] |
| | | LGB+ | 836,810 (3.6) | 310 (7.0) | 37.05 [33.04, 41.41] | 37.26 [33.09, 41.44] |
| | | Gay or Lesbian | 419,890 (1.8) | 180 (4.1) | 42.87 [36.83, 49.61] | 41.66 [35.39, 47.94] |
| | | Bisexual | 330,125 (1.4) | 100 (2.3) | 30.29 [24.65, 36.84] | 35.17 [27.63, 42.71] |
| | | Other | 86,795 (0.4) | 25 (0.6) | 28.80 [18.64, 42.52] | 33.02 [21.16, 49.02] |
| | Not good health | Total | 5,399,540 (100.0) | 2,620 (100.0) | 48.52 [46.68, 50.42] | 63.94 [60.12, 67.76] |
| | | Heterosexual | 5,167,470 (95.7) | 2,385 (91.0) | 46.15 [44.32, 48.04] | 59.28 [55.30, 63.25] |
| | | LGB+ | 232,070 (4.3) | 235 (9.0) | 101.26 [88.73, 115.07] | 101.22 [88.00, 114.44] |
| | | Gay or Lesbian | 95,480 (1.8) | 95 (3.6) | 99.50 [80.50, 121.63] | 99.64 [78.00, 124.84] |
| | | Bisexual | 104,375 (1.9) | 100 (3.8) | 95.81 [77.95, 116.53] | 97.30 [78.20, 116.39] |
| | | Other | 32,210 (0.6) | 35 (1.3) | 108.66 [75.69, 151.12] | 115.14 [81.06, 159.02] |
| Disability | Day-to-day activities limited a lot | Total | 1,904,385 (100.0) | 1,325 (100.0) | 69.58 [65.88, 73.43] | 96.14 [88.50, 103.77] |
| | | Heterosexual | 1,807,315 (94.9) | 1,185 (89.4) | 65.57 [61.89, 69.41] | 88.83 [80.82, 96.85] |
| | | LGB+ | 97,075 (5.1) | 140 (10.6) | 144.22 [121.32, 170.18] | 144.56 [120.19, 168.93] |

| | | | | | | |
|---|---|---|---|---|---|---|
| | | Gay or Lesbian | 39,370 (2.1) | 60 (4.5) | 152.40 [116.30, 196.17] | 154.46 [114.04, 203.47] |
| | | Bisexual | 43,105 (2.3) | 60 (4.5) | 139.19 [106.22, 179.17] | 137.53 [104.85, 177.15] |
| | | Other | 14,595 (0.8) | 20 (1.5) | 137.03 [83.70, 211.64] | 129.96 [77.65, 203.14] |
| | Day-to-day activities limited a little | Total | 3,184,715 (100.0) | 1,380 (100.0) | 43.33 [41.08, 45.68] | 52.85 [49.33, 56.38] |
| | | Heterosexual | 2,970,515 (93.3) | 1,230 (89.1) | 41.41 [39.12, 43.79] | 50.98 [47.14, 54.81] |
| | | LGB+ | 214,195 (6.7) | 150 (10.9) | 70.03 [59.27, 82.18] | 72.81 [60.93, 84.69] |
| | | Gay or Lesbian | 76,965 (2.4) | 70 (5.1) | 90.95 [70.90, 114.91] | 92.57 [71.32, 117.96] |
| | | Bisexual | 108,705 (3.4) | 60 (4.3) | 55.20 [42.12, 71.05] | 65.13 [48.44, 85.36] |
| | | Other | 28,525 (0.9) | 20 (1.4) | 70.11 [42.83, 108.29] | 79.07 [45.46, 126.09] |
| | Day-to-day activities not limited | Total | 23,250,565 (100.0) | 4,350 (100.0) | 18.71 [18.16, 19.27] | 18.03 [17.39, 18.68] |
| | | Heterosexual | 22,492,955 (96.7) | 4,095 (94.1) | 18.21 [17.65, 18.77] | 17.50 [16.85, 18.15] |
| | | LGB+ | 757,610 (3.3) | 255 (5.9) | 33.66 [29.65, 38.05] | 33.20 [29.11, 37.29] |
| | | Gay or Lesbian | 399,030 (1.7) | 150 (3.4) | 37.59 [31.82, 44.11] | 34.64 [28.83, 40.45] |
| | | Bisexual | 282,690 (1.2) | 80 (1.8) | 28.30 [22.44, 35.22] | 31.95 [24.94, 40.23] |
| | | Other | 75,885 (0.3) | 25 (0.6) | 32.94 [21.32, 48.63] | 32.74 [20.81, 48.85] |

Notes: Age-standardised rates are expressed per 100,000 people and standardised to the age distribution of the observed study population identifying with an LGB+ orientation in Census 2021. Cells with the value '-' are not applicable for the given characteristic or category. Sexual orientation groups Gay or Lesbian, Bisexual, and Other are not presented for subcategories where suppression was applied for the purposes of statistical disclosure control. All calculations are based on estimates rounded to the nearest multiple of five for disclosure control reasons. CI: confidence interval.

**Supplementary Table 16: Sensitivity analysis restricting to people living in private households only, ratios of age-specific and age-standardised rates of self-harm and suicide per 100,000 people**

| Characteristic | Category | Sexual orientation | Rate ratio denominator | Rate ratio for self-harm [95% CI] | Rate ratio for suicide [95% CI] |
|---|---|---|---|---|---|
| Total | Total | LGB+ | Heterosexual | 2.51 [2.47, 2.55] | 2.19 [2.00, 2.39] |
| | | Gay or Lesbian | Heterosexual | 2.42 [2.35, 2.49] | 2.22 [1.94, 2.50] |
| | | Bisexual | Heterosexual | 2.78 [2.71, 2.85] | 2.20 [1.87, 2.53] |
| | | Other | Heterosexual | 2.22 [2.11, 2.33] | 2.36 [1.77, 2.97] |
| Age on Census Day (10-year bands) | 16 to 24 years | LGB+ | Heterosexual | 2.77 [2.69, 2.84] | 1.97 [1.55, 2.43] |
| | | Gay or Lesbian | Heterosexual | 2.83 [2.70, 2.95] | c |
| | | Bisexual | Heterosexual | 2.81 [2.72, 2.90] | c |
| | | Other | Heterosexual | 2.44 [2.27, 2.62] | c |
| | 25 to 34 years | LGB+ | Heterosexual | 2.51 [2.43, 2.59] | 2.07 [1.72, 2.44] |
| | | Gay or Lesbian | Heterosexual | 2.34 [2.23, 2.45] | c |
| | | Bisexual | Heterosexual | 2.76 [2.64, 2.88] | c |
| | | Other | Heterosexual | 2.24 [2.05, 2.44] | c |
| | 35 to 44 years | LGB+ | Heterosexual | 2.23 [2.13, 2.33] | 2.23 [1.81, 2.66] |
| | | Gay or Lesbian | Heterosexual | 1.99 [1.86, 2.12] | c |
| | | Bisexual | Heterosexual | 2.75 [2.56, 2.94] | c |
| | | Other | Heterosexual | 1.84 [1.58, 2.11] | c |
| | 45 to 54 years | LGB+ | Heterosexual | 2.04 [1.92, 2.17] | 2.25 [1.79, 2.73] |
| | | Gay or Lesbian | Heterosexual | 1.70 [1.56, 1.83] | c |
| | | Bisexual | Heterosexual | 2.94 [2.66, 3.23] | c |
| | | Other | Heterosexual | 1.91 [1.53, 2.30] | c |
| | 55 to 64 years | LGB+ | Heterosexual | 1.92 [1.73, 2.10] | 2.54 [1.89, 3.21] |
| | | Gay or Lesbian | Heterosexual | 1.66 [1.46, 1.86] | c |
| | | Bisexual | Heterosexual | 2.58 [2.14, 3.01] | c |
| | | Other | Heterosexual | 2.06 [1.37, 2.74] | c |
| | 65 years and over | LGB+ | Heterosexual | 1.59 [1.33, 1.84] | 2.81 [1.71, 3.93] |
| | | Gay or Lesbian | Heterosexual | 1.50 [1.21, 1.80] | c |
| | | Bisexual | Heterosexual | 1.89 [1.34, 2.45] | c |
| | | Other | Heterosexual | 1.34 [0.51, 2.16] | c |
| Sex | Female | LGB+ | Heterosexual | 2.75 [2.69, 2.81] | 3.15 [2.69, 3.62] |
| | | Gay or Lesbian | Heterosexual | 3.02 [2.91, 3.13] | 3.02 [2.30, 3.77] |
| | | Bisexual | Heterosexual | 2.84 [2.76, 2.91] | 3.19 [2.52, 3.89] |
| | | Other | Heterosexual | 2.34 [2.21, 2.48] | 3.31 [2.06, 4.56] |
| | Male | LGB+ | Heterosexual | 1.91 [1.84, 1.97] | 1.75 [1.55, 1.96] |
| | | Gay or Lesbian | Heterosexual | 1.91 [1.83, 1.99] | 1.67 [1.42, 1.93] |
| | | Bisexual | Heterosexual | 2.07 [1.96, 2.18] | 1.95 [1.53, 2.37] |
| | | Other | Heterosexual | 1.70 [1.53, 1.88] | 2.12 [1.40, 2.85] |
| Region | England: North East | LGB+ | Heterosexual | 2.24 [2.09, 2.40] | 2.07 [1.33, 2.87] |
| | | Gay or Lesbian | Heterosexual | 2.30 [2.06, 2.53] | c |
| | | Bisexual | Heterosexual | 2.34 [2.08, 2.60] | c |
| | | Other | Heterosexual | 2.22 [1.73, 2.72] | c |
| | England: North West | LGB+ | Heterosexual | 2.75 [2.62, 2.88] | 2.73 [2.17, 3.32] |
| | | Gay or Lesbian | Heterosexual | 2.68 [2.49, 2.87] | c |
| | | Bisexual | Heterosexual | 3.08 [2.87, 3.29] | c |
| | | Other | Heterosexual | 2.16 [1.83, 2.50] | c |
| | England: Yorkshire and The Humber | LGB+ | Heterosexual | 2.17 [2.06, 2.29] | 2.25 [1.64, 2.91] |
| | | Gay or Lesbian | Heterosexual | 2.16 [1.99, 2.33] | c |
| | | Bisexual | Heterosexual | 2.35 [2.18, 2.53] | c |
| | | Other | Heterosexual | 1.88 [1.59, 2.16] | c |
| | England: East Midlands | LGB+ | Heterosexual | 2.64 [2.49, 2.79] | 2.50 [1.75, 3.31] |
| | | Gay or Lesbian | Heterosexual | 2.56 [2.33, 2.80] | c |
| | | Bisexual | Heterosexual | 2.94 [2.71, 3.17] | c |
| | | Other | Heterosexual | 2.19 [1.82, 2.56] | c |
| | England: West Midlands | LGB+ | Heterosexual | 2.85 [2.67, 3.03] | 1.69 [1.09, 2.32] |
| | | Gay or Lesbian | Heterosexual | 2.82 [2.55, 3.10] | c |
| | | Bisexual | Heterosexual | 3.12 [2.85, 3.39] | c |
| | | Other | Heterosexual | 2.21 [1.79, 2.63] | c |
| | England: East of England | LGB+ | Heterosexual | 2.69 [2.54, 2.83] | 2.40 [1.68, 3.16] |
| | | Gay or Lesbian | Heterosexual | 2.46 [2.24, 2.68] | c |
| | | Bisexual | Heterosexual | 3.02 [2.80, 3.24] | c |
| | | Other | Heterosexual | 2.48 [2.11, 2.85] | c |
| | England: London | LGB+ | Heterosexual | 2.24 [2.12, 2.35] | 2.33 [1.78, 2.91] |
| | | Gay or Lesbian | Heterosexual | 2.07 [1.88, 2.26] | c |
| | | Bisexual | Heterosexual | 2.50 [2.32, 2.68] | c |
| | | Other | Heterosexual | 2.22 [1.94, 2.51] | c |
| | | LGB+ | Heterosexual | 2.63 [2.52, 2.75] | 2.05 [1.58, 2.55] |

| | | | | | |
|---|---|---|---|---|---|
| | England: South East | Gay or Lesbian | Heterosexual | 2.42 [2.25, 2.60] | c |
| | | Bisexual | Heterosexual | 2.94 [2.77, 3.11] | c |
| | | Other | Heterosexual | 2.41 [2.12, 2.71] | c |
| | England: South West | LGB+ | Heterosexual | 2.71 [2.57, 2.85] | 1.80 [1.26, 2.37] |
| | | Gay or Lesbian | Heterosexual | 2.76 [2.54, 2.99] | c |
| | | Bisexual | Heterosexual | 2.84 [2.64, 3.04] | c |
| | | Other | Heterosexual | 2.64 [2.27, 3.01] | c |
| | Wales | LGB+ | Heterosexual | 2.26 [2.06, 2.46] | 2.53 [1.55, 3.64] |
| | | Gay or Lesbian | Heterosexual | 2.38 [2.06, 2.71] | c |
| | | Bisexual | Heterosexual | 2.51 [2.19, 2.84] | c |
| | | Other | Heterosexual | 1.39 [0.94, 1.84] | c |
| Ethnic group | Asian, Asian British or Asian Welsh | LGB+ | Heterosexual | 2.24 [2.00, 2.49] | 2.44 [1.06, 3.90] |
| | | Gay or Lesbian | Heterosexual | 2.55 [2.02, 3.08] | c |
| | | Bisexual | Heterosexual | 2.13 [1.82, 2.45] | c |
| | | Other | Heterosexual | 2.13 [1.57, 2.69] | c |
| | Black, Black British, Black Welsh, Caribbean or African | LGB+ | Heterosexual | 2.89 [2.53, 3.26] | 4.95 [2.33, 8.00] |
| | | Gay or Lesbian | Heterosexual | 3.16 [2.42, 3.90] | c |
| | | Bisexual | Heterosexual | 2.95 [2.44, 3.46] | c |
| | | Other | Heterosexual | 2.62 [1.89, 3.35] | c |
| | Mixed or Multiple ethnic groups | LGB+ | Heterosexual | 2.30 [2.11, 2.48] | 2.26 [1.25, 3.40] |
| | | Gay or Lesbian | Heterosexual | 2.11 [1.83, 2.39] | c |
| | | Bisexual | Heterosexual | 2.50 [2.22, 2.79] | c |
| | | Other | Heterosexual | 2.07 [1.64, 2.50] | c |
| | White | LGB+ | Heterosexual | 2.38 [2.34, 2.43] | 2.05 [1.86, 2.25] |
| | | Gay or Lesbian | Heterosexual | 2.26 [2.19, 2.32] | c |
| | | Bisexual | Heterosexual | 2.69 [2.62, 2.76] | c |
| | | Other | Heterosexual | 2.23 [2.10, 2.35] | c |
| | Other ethnic group | LGB+ | Heterosexual | 1.99 [1.65, 2.34] | 2.80 [0.24, 5.86] |
| | | Gay or Lesbian | Heterosexual | 2.31 [1.51, 3.12] | c |
| | | Bisexual | Heterosexual | 2.13 [1.61, 2.68] | c |
| | | Other | Heterosexual | 1.58 [1.07, 2.11] | c |
| Rural/urban classification | Urban | LGB+ | Heterosexual | 2.46 [2.42, 2.51] | 2.19 [1.98, 2.41] |
| | | Gay or Lesbian | Heterosexual | 2.38 [2.31, 2.45] | 2.16 [1.87, 2.46] |
| | | Bisexual | Heterosexual | 2.72 [2.65, 2.79] | 2.21 [1.86, 2.57] |
| | | Other | Heterosexual | 2.17 [2.06, 2.29] | 2.43 [1.79, 3.08] |
| | Rural | LGB+ | Heterosexual | 2.85 [2.72, 2.99] | 2.26 [1.71, 2.83] |
| | | Gay or Lesbian | Heterosexual | 2.66 [2.45, 2.87] | 2.67 [1.79, 3.58] |
| | | Bisexual | Heterosexual | 3.22 [3.01, 3.43] | 2.12 [1.27, 3.00] |
| | | Other | Heterosexual | 2.63 [2.25, 3.02] | 1.90 [0.25, 3.57] |
| Country of birth | Born in the UK | LGB+ | Heterosexual | 2.48 [2.44, 2.53] | 2.23 [2.02, 2.45] |
| | | Gay or Lesbian | Heterosexual | 2.33 [2.27, 2.40] | 2.21 [1.92, 2.50] |
| | | Bisexual | Heterosexual | 2.79 [2.72, 2.86] | 2.21 [1.86, 2.57] |
| | | Other | Heterosexual | 2.40 [2.27, 2.53] | 2.83 [2.05, 3.60] |
| | Born outside the UK | LGB+ | Heterosexual | 2.36 [2.22, 2.50] | 1.95 [1.42, 2.50] |
| | | Gay or Lesbian | Heterosexual | 2.30 [2.03, 2.57] | 2.05 [1.03, 3.09] |
| | | Bisexual | Heterosexual | 2.58 [2.38, 2.78] | 2.20 [1.34, 3.09] |
| | | Other | Heterosexual | 1.99 [1.71, 2.27] | 1.63 [0.48, 2.78] |
| English language proficiency | Main language is English | LGB+ | Heterosexual | 2.48 [2.44, 2.53] | 2.20 [2.00, 2.40] |
| | | Gay or Lesbian | Heterosexual | 2.35 [2.29, 2.42] | 2.17 [1.90, 2.46] |
| | | Bisexual | Heterosexual | 2.77 [2.71, 2.84] | 2.26 [1.92, 2.60] |
| | | Other | Heterosexual | 2.34 [2.22, 2.46] | 2.59 [1.91, 3.28] |
| | Main language is not English | LGB+ | Heterosexual | 2.05 [1.84, 2.26] | 1.58 [0.75, 2.45] |
| | | Gay or Lesbian | Heterosexual | 2.25 [1.74, 2.77] | 1.52 [0.17, 2.92] |
| | | Bisexual | Heterosexual | 2.12 [1.83, 2.42] | 1.09 [0.06, 2.14] |
| | | Other | Heterosexual | 1.88 [1.50, 2.26] | 2.23 [0.29, 4.23] |
| Religion | No religion | LGB+ | Heterosexual | 2.08 [2.04, 2.13] | 1.93 [1.72, 2.15] |
| | | Gay or Lesbian | Heterosexual | 1.98 [1.91, 2.04] | c |
| | | Bisexual | Heterosexual | 2.34 [2.27, 2.41] | c |
| | | Other | Heterosexual | 1.97 [1.85, 2.10] | c |
| | Christian | LGB+ | Heterosexual | 2.93 [2.82, 3.04] | 2.20 [1.77, 2.64] |
| | | Gay or Lesbian | Heterosexual | 2.81 [2.64, 2.98] | c |
| | | Bisexual | Heterosexual | 3.45 [3.27, 3.63] | c |
| | | Other | Heterosexual | 2.15 [1.89, 2.41] | c |
| | Buddhist | LGB+ | Heterosexual | 3.83 [3.14, 4.62] | 2.67 [0.83, 5.37] |
| | | Gay or Lesbian | Heterosexual | 3.21 [2.16, 4.35] | c |
| | | Bisexual | Heterosexual | 3.94 [3.04, 4.93] | c |
| | | Other | Heterosexual | 4.48 [2.89, 6.18] | c |
| | Hindu | LGB+ | Heterosexual | 2.19 [1.56, 2.83] | 2.70 [0.00, 6.41] |
| | | Gay or Lesbian | Heterosexual | 2.94 [1.02, 4.90] | c |
| | | Bisexual | Heterosexual | 2.16 [1.41, 2.94] | c |
| | | Other | Heterosexual | 1.95 [0.37, 3.55] | c |

| | | | | | |
|---|---|---|---|---|---|
| | Jewish | LGB+ | Heterosexual | 2.96 [2.23, 3.79] | 2.57 [0.00, 6.82] |
| | | Gay or Lesbian | Heterosexual | 2.39 [1.40, 3.48] | c |
| | | Bisexual | Heterosexual | 3.46 [2.34, 4.70] | c |
| | | Other | Heterosexual | 2.79 [0.98, 4.71] | c |
| | Muslim | LGB+ | Heterosexual | 2.12 [1.78, 2.46] | 2.87 [0.00, 6.38] |
| | | Gay or Lesbian | Heterosexual | 3.30 [2.18, 4.42] | c |
| | | Bisexual | Heterosexual | 2.24 [1.75, 2.73] | c |
| | | Other | Heterosexual | 1.48 [0.99, 1.96] | c |
| | Sikh | LGB+ | Heterosexual | 2.11 [1.32, 2.93] | 3.65 [0.00, 8.30] |
| | | Gay or Lesbian | Heterosexual | 4.39 [1.64, 7.18] | c |
| | | Bisexual | Heterosexual | 1.16 [0.41, 1.93] | c |
| | | Other | Heterosexual | 2.66 [0.80, 4.56] | c |
| | Other religion | LGB+ | Heterosexual | 2.77 [2.49, 3.10] | 1.16 [0.60, 1.95] |
| | | Gay or Lesbian | Heterosexual | 2.98 [2.47, 3.53] | c |
| | | Bisexual | Heterosexual | 2.78 [2.43, 3.16] | c |
| | | Other | Heterosexual | 2.61 [2.18, 3.07] | c |
| | Not answered | LGB+ | Heterosexual | 2.80 [2.52, 3.08] | 1.67 [0.82, 2.61] |
| | | Gay or Lesbian | Heterosexual | 2.78 [2.33, 3.24] | c |
| | | Bisexual | Heterosexual | 3.14 [2.70, 3.59] | c |
| | | Other | Heterosexual | 2.23 [1.62, 2.84] | c |
| IMD/WIMD decile group | 1 (most deprived) | LGB+ | Heterosexual | 2.37 [2.26, 2.47] | 2.19 [1.65, 2.75] |
| | | Gay or Lesbian | Heterosexual | 2.47 [2.31, 2.64] | 2.17 [1.41, 2.96] |
| | | Bisexual | Heterosexual | 2.60 [2.44, 2.76] | 2.64 [1.66, 3.65] |
| | | Other | Heterosexual | 1.52 [1.30, 1.74] | 1.01 [0.07, 1.97] |
| | 2 | LGB+ | Heterosexual | 2.27 [2.17, 2.38] | 1.91 [1.39, 2.47] |
| | | Gay or Lesbian | Heterosexual | 2.30 [2.14, 2.47] | 1.86 [1.07, 2.67] |
| | | Bisexual | Heterosexual | 2.38 [2.22, 2.54] | 1.83 [1.02, 2.67] |
| | | Other | Heterosexual | 1.92 [1.67, 2.18] | 2.28 [0.72, 3.88] |
| | 3 | LGB+ | Heterosexual | 2.33 [2.22, 2.44] | 2.51 [1.92, 3.13] |
| | | Gay or Lesbian | Heterosexual | 2.42 [2.24, 2.60] | 2.69 [1.80, 3.63] |
| | | Bisexual | Heterosexual | 2.48 [2.31, 2.65] | 2.13 [1.29, 3.01] |
| | | Other | Heterosexual | 1.84 [1.58, 2.10] | 2.69 [1.00, 4.43] |
| | 4 | LGB+ | Heterosexual | 2.43 [2.31, 2.55] | 1.95 [1.45, 2.47] |
| | | Gay or Lesbian | Heterosexual | 2.40 [2.20, 2.59] | 2.08 [1.35, 2.85] |
| | | Bisexual | Heterosexual | 2.65 [2.47, 2.84] | 1.65 [0.87, 2.46] |
| | | Other | Heterosexual | 2.27 [1.96, 2.59] | 2.09 [0.56, 3.65] |
| | 5 | LGB+ | Heterosexual | 2.61 [2.48, 2.75] | 1.88 [1.37, 2.42] |
| | | Gay or Lesbian | Heterosexual | 2.47 [2.26, 2.68] | 1.88 [1.19, 2.60] |
| | | Bisexual | Heterosexual | 2.81 [2.61, 3.02] | 1.57 [0.80, 2.35] |
| | | Other | Heterosexual | 2.46 [2.10, 2.82] | 1.46 [0.20, 2.73] |
| | 6 | LGB+ | Heterosexual | 2.50 [2.36, 2.64] | 2.36 [1.75, 3.02] |
| | | Gay or Lesbian | Heterosexual | 2.20 [2.00, 2.41] | 2.24 [1.42, 3.09] |
| | | Bisexual | Heterosexual | 2.97 [2.74, 3.20] | 2.53 [1.42, 3.69] |
| | | Other | Heterosexual | 2.21 [1.84, 2.58] | 2.35 [0.51, 4.25] |
| | 7 | LGB+ | Heterosexual | 2.63 [2.47, 2.79] | 2.20 [1.55, 2.89] |
| | | Gay or Lesbian | Heterosexual | 2.27 [2.03, 2.50] | 1.92 [1.09, 2.80] |
| | | Bisexual | Heterosexual | 3.06 [2.81, 3.32] | 2.31 [1.15, 3.53] |
| | | Other | Heterosexual | 2.67 [2.22, 3.11] | 4.17 [1.26, 7.17] |
| | 8 | LGB+ | Heterosexual | 2.68 [2.51, 2.85] | 2.41 [1.69, 3.18] |
| | | Gay or Lesbian | Heterosexual | 2.43 [2.18, 2.68] | 2.25 [1.27, 3.27] |
| | | Bisexual | Heterosexual | 3.07 [2.81, 3.34] | 2.55 [1.34, 3.81] |
| | | Other | Heterosexual | 2.52 [2.05, 3.00] | 2.65 [0.04, 5.32] |
| | 9 | LGB+ | Heterosexual | 2.72 [2.54, 2.91] | 2.10 [1.39, 2.87] |
| | | Gay or Lesbian | Heterosexual | 2.55 [2.27, 2.83] | 2.39 [1.23, 3.60] |
| | | Bisexual | Heterosexual | 3.04 [2.75, 3.34] | 2.40 [1.06, 3.81] |
| | | Other | Heterosexual | 3.07 [2.51, 3.64] | 1.99 [0.00, 4.15] |
| | 10 (least deprived) | LGB+ | Heterosexual | 2.86 [2.64, 3.09] | 2.45 [1.57, 3.41] |
| | | Gay or Lesbian | Heterosexual | 2.41 [2.10, 2.73] | 3.00 [1.47, 4.64] |
| | | Bisexual | Heterosexual | 3.30 [2.95, 3.65] | 2.29 [0.76, 3.90] |
| | | Other | Heterosexual | 3.16 [2.52, 3.81] | 4.55 [0.55, 8.65] |
| Highest level of qualification (≥25 years) | No qualifications | LGB+ | Heterosexual | 1.98 [1.85, 2.11] | 2.00 [1.42, 2.62] |
| | | Gay or Lesbian | Heterosexual | 2.82 [2.55, 3.09] | c |
| | | Bisexual | Heterosexual | 2.18 [1.96, 2.40] | c |
| | | Other | Heterosexual | 0.70 [0.56, 0.85] | c |
| | Level 1 (one to four GCSE passes (grade A* to C or grade 4 and above) and any other GCSEs at other grades, or | LGB+ | Heterosexual | 2.54 [2.36, 2.72] | 1.74 [1.05, 2.47] |
| | | Gay or Lesbian | Heterosexual | 2.36 [2.11, 2.61] | c |
| | | Bisexual | Heterosexual | 3.01 [2.71, 3.31] | c |
| | | Other | Heterosexual | 1.77 [1.32, 2.22] | c |

| | | | | | |
|---|---|---|---|---|---|
| | equivalent qualifications) | | | | |
| | Level 2 (five or more GCSE passes (grade A* to C or grade 4 and above) or equivalent qualifications) | LGB+ | Heterosexual | 2.58 [2.43, 2.73] | 1.74 [1.22, 2.28] |
| | | Gay or Lesbian | Heterosexual | 2.16 [1.98, 2.35] | c |
| | | Bisexual | Heterosexual | 3.20 [2.94, 3.45] | c |
| | | Other | Heterosexual | 2.30 [1.86, 2.73] | c |
| | Apprenticeship | LGB+ | Heterosexual | 2.19 [1.91, 2.47] | 0.94 [0.23, 1.69] |
| | | Gay or Lesbian | Heterosexual | 2.18 [1.76, 2.61] | c |
| | | Bisexual | Heterosexual | 2.63 [2.15, 3.11] | c |
| | | Other | Heterosexual | 1.20 [0.67, 1.72] | c |
| | Level 3 (two or more A Levels or equivalent qualifications) | LGB+ | Heterosexual | 3.00 [2.86, 3.15] | 2.88 [2.28, 3.51] |
| | | Gay or Lesbian | Heterosexual | 2.66 [2.48, 2.84] | c |
| | | Bisexual | Heterosexual | 3.54 [3.28, 3.80] | c |
| | | Other | Heterosexual | 3.34 [2.86, 3.83] | c |
| | Level 4 and above (Higher National Certificate, Higher National Diploma, Bachelor's degree, or post-graduate qualifications) | LGB+ | Heterosexual | 2.46 [2.36, 2.55] | 2.59 [2.22, 2.97] |
| | | Gay or Lesbian | Heterosexual | 2.14 [2.02, 2.26] | c |
| | | Bisexual | Heterosexual | 2.88 [2.70, 3.05] | c |
| | | Other | Heterosexual | 2.88 [2.56, 3.19] | c |
| | Other qualifications | LGB+ | Heterosexual | 2.09 [1.71, 2.48] | 2.37 [0.68, 4.33] |
| | | Gay or Lesbian | Heterosexual | 2.47 [1.83, 3.13] | c |
| | | Bisexual | Heterosexual | 2.17 [1.56, 2.80] | c |
| | | Other | Heterosexual | 1.17 [0.55, 1.80] | c |
| NS-SEC | Class 1 (Higher managerial, administrative and professional occupations) | LGB+ | Heterosexual | 3.37 [2.65, 4.14] | 2.25 [1.64, 2.90] |
| | | Gay or Lesbian | Heterosexual | 2.93 [1.76, 4.12] | c |
| | | Bisexual | Heterosexual | 3.86 [2.85, 4.92] | c |
| | | Other | Heterosexual | 3.47 [1.64, 5.34] | c |
| | Class 2 (Lower managerial, administrative and professional occupations) | LGB+ | Heterosexual | 2.87 [2.61, 3.13] | 2.64 [1.76, 3.61] |
| | | Gay or Lesbian | Heterosexual | 2.56 [2.16, 2.96] | c |
| | | Bisexual | Heterosexual | 3.25 [2.90, 3.61] | c |
| | | Other | Heterosexual | 2.89 [2.23, 3.56] | c |
| | Class 3 (Intermediate occupations) | LGB+ | Heterosexual | 2.83 [2.65, 3.02] | 2.43 [1.74, 3.18] |
| | | Gay or Lesbian | Heterosexual | 2.70 [2.42, 2.99] | c |
| | | Bisexual | Heterosexual | 3.17 [2.90, 3.45] | c |
| | | Other | Heterosexual | 2.18 [1.73, 2.63] | c |
| | Class 4 (Small employers and own account workers) | LGB+ | Heterosexual | 2.05 [1.77, 2.34] | 1.26 [0.80, 1.74] |
| | | Gay or Lesbian | Heterosexual | 2.03 [1.47, 2.58] | c |
| | | Bisexual | Heterosexual | 2.38 [1.96, 2.81] | c |
| | | Other | Heterosexual | 1.30 [0.90, 1.71] | c |
| | Class 5 (Lower supervisory and technical occupations) | LGB+ | Heterosexual | 2.84 [2.53, 3.15] | 1.93 [1.17, 2.76] |
| | | Gay or Lesbian | Heterosexual | 2.72 [2.28, 3.17] | c |
| | | Bisexual | Heterosexual | 3.09 [2.64, 3.55] | c |
| | | Other | Heterosexual | 2.60 [1.47, 3.72] | c |
| | Class 6 (Semi-routine occupations) | LGB+ | Heterosexual | 2.58 [2.47, 2.69] | 2.68 [2.04, 3.37] |
| | | Gay or Lesbian | Heterosexual | 2.52 [2.34, 2.70] | c |
| | | Bisexual | Heterosexual | 2.77 [2.61, 2.94] | c |
| | | Other | Heterosexual | 2.24 [1.93, 2.54] | c |
| | Class 7 (Routine occupations) | LGB+ | Heterosexual | 2.41 [2.29, 2.53] | 2.23 [1.70, 2.79] |
| | | Gay or Lesbian | Heterosexual | 2.50 [2.30, 2.70] | c |
| | | Bisexual | Heterosexual | 2.56 [2.39, 2.73] | c |
| | | Other | Heterosexual | 1.75 [1.49, 2.02] | c |
| | Class 8 (Never worked and long-term unemployed) | LGB+ | Heterosexual | 2.26 [2.16, 2.36] | 2.15 [1.59, 2.76] |
| | | Gay or Lesbian | Heterosexual | 2.76 [2.57, 2.95] | c |
| | | Bisexual | Heterosexual | 2.38 [2.24, 2.53] | c |
| | | Other | Heterosexual | 1.20 [1.04, 1.36] | c |
| | Full-time students | LGB+ | Heterosexual | 2.78 [2.47, 3.10] | 3.90 [0.07, 8.26] |
| | | Gay or Lesbian | Heterosexual | 2.96 [2.44, 3.48] | c |
| | | Bisexual | Heterosexual | 2.51 [2.22, 2.81] | c |
| | | Other | Heterosexual | 2.79 [2.10, 3.49] | c |
| Household NS-SEC | Class 1 (Higher managerial, administrative | LGB+ | Heterosexual | 2.76 [2.63, 2.89] | 2.29 [1.83, 2.78] |
| | | Gay or Lesbian | Heterosexual | 2.50 [2.30, 2.71] | c |
| | | Bisexual | Heterosexual | 3.07 [2.88, 3.28] | c |
| | | Other | Heterosexual | 3.01 [2.64, 3.39] | c |

| | | | | | |
|---|---|---|---|---|---|
| | and professional occupations) | | | | |
| | Class 2 (Lower managerial, administrative and professional occupations) | LGB+ | Heterosexual | 2.62 [2.52, 2.72] | 2.28 [1.88, 2.70] |
| | | Gay or Lesbian | Heterosexual | 2.43 [2.29, 2.58] | c |
| | | Bisexual | Heterosexual | 2.88 [2.74, 3.03] | c |
| | | Other | Heterosexual | 2.72 [2.44, 3.00] | c |
| | Class 3 (Intermediate occupations) | LGB+ | Heterosexual | 2.76 [2.62, 2.90] | 2.23 [1.62, 2.87] |
| | | Gay or Lesbian | Heterosexual | 2.79 [2.58, 3.00] | c |
| | | Bisexual | Heterosexual | 2.87 [2.67, 3.08] | c |
| | | Other | Heterosexual | 2.33 [1.98, 2.68] | c |
| | Class 4 (Small employers and own account workers) | LGB+ | Heterosexual | 2.21 [2.05, 2.37] | 1.55 [1.01, 2.12] |
| | | Gay or Lesbian | Heterosexual | 2.38 [2.10, 2.67] | c |
| | | Bisexual | Heterosexual | 2.37 [2.14, 2.61] | c |
| | | Other | Heterosexual | 1.59 [1.29, 1.89] | c |
| | Class 5 (Lower supervisory and technical occupations) | LGB+ | Heterosexual | 2.56 [2.37, 2.76] | 2.14 [1.30, 3.03] |
| | | Gay or Lesbian | Heterosexual | 2.68 [2.36, 3.00] | c |
| | | Bisexual | Heterosexual | 2.83 [2.53, 3.13] | c |
| | | Other | Heterosexual | 1.41 [1.02, 1.81] | c |
| | Class 6 (Semi-routine occupations) | LGB+ | Heterosexual | 2.54 [2.42, 2.65] | 2.51 [1.87, 3.21] |
| | | Gay or Lesbian | Heterosexual | 2.69 [2.50, 2.87] | c |
| | | Bisexual | Heterosexual | 2.61 [2.45, 2.78] | c |
| | | Other | Heterosexual | 2.04 [1.77, 2.32] | c |
| | Class 7 (Routine occupations) | LGB+ | Heterosexual | 2.43 [2.29, 2.56] | 2.37 [1.72, 3.07] |
| | | Gay or Lesbian | Heterosexual | 2.57 [2.34, 2.80] | c |
| | | Bisexual | Heterosexual | 2.61 [2.42, 2.81] | c |
| | | Other | Heterosexual | 1.56 [1.31, 1.82] | c |
| | Class 8 (Never worked and long-term unemployed) | LGB+ | Heterosexual | 2.18 [2.06, 2.30] | 2.11 [1.43, 2.86] |
| | | Gay or Lesbian | Heterosexual | 2.71 [2.48, 2.94] | c |
| | | Bisexual | Heterosexual | 2.28 [2.12, 2.45] | c |
| | | Other | Heterosexual | 1.11 [0.92, 1.29] | c |
| | Full-time students | LGB+ | Heterosexual | 2.64 [2.20, 3.10] | 3.92 [0.00, 10.72] |
| | | Gay or Lesbian | Heterosexual | 2.90 [2.15, 3.68] | c |
| | | Bisexual | Heterosexual | 2.33 [1.93, 2.76] | c |
| | | Other | Heterosexual | 2.68 [1.47, 3.90] | c |
| Lone person household indicator | Lives alone | LGB+ | Heterosexual | 2.04 [1.95, 2.13] | 1.72 [1.42, 2.05] |
| | | Gay or Lesbian | Heterosexual | 2.07 [1.92, 2.22] | c |
| | | Bisexual | Heterosexual | 2.22 [2.10, 2.34] | c |
| | | Other | Heterosexual | 1.73 [1.52, 1.93] | c |
| | Does not live alone | LGB+ | Heterosexual | 2.39 [2.34, 2.44] | 2.01 [1.77, 2.25] |
| | | Gay or Lesbian | Heterosexual | 2.27 [2.19, 2.34] | c |
| | | Bisexual | Heterosexual | 2.66 [2.58, 2.73] | c |
| | | Other | Heterosexual | 2.08 [1.96, 2.20] | c |
| Household size | One person in household | LGB+ | Heterosexual | 2.04 [1.95, 2.13] | 1.72 [1.42, 2.05] |
| | | Gay or Lesbian | Heterosexual | 2.07 [1.92, 2.22] | c |
| | | Bisexual | Heterosexual | 2.22 [2.10, 2.34] | c |
| | | Other | Heterosexual | 1.73 [1.52, 1.93] | c |
| | Two people in household | LGB+ | Heterosexual | 2.07 [2.00, 2.15] | 1.70 [1.41, 2.00] |
| | | Gay or Lesbian | Heterosexual | 1.96 [1.84, 2.09] | c |
| | | Bisexual | Heterosexual | 2.37 [2.26, 2.49] | c |
| | | Other | Heterosexual | 2.09 [1.87, 2.31] | c |
| | Three to five people in household | LGB+ | Heterosexual | 2.48 [2.41, 2.56] | 2.26 [1.77, 2.76] |
| | | Gay or Lesbian | Heterosexual | 2.43 [2.31, 2.55] | c |
| | | Bisexual | Heterosexual | 2.70 [2.58, 2.82] | c |
| | | Other | Heterosexual | 1.96 [1.80, 2.12] | c |
| | Six or more people in household | LGB+ | Heterosexual | 2.47 [2.24, 2.71] | 1.36 [0.14, 2.65] |
| | | Gay or Lesbian | Heterosexual | 2.66 [2.21, 3.13] | c |
| | | Bisexual | Heterosexual | 2.71 [2.38, 3.04] | c |
| | | Other | Heterosexual | 1.74 [1.32, 2.16] | c |
| Household tenure | Owned or shared ownership | LGB+ | Heterosexual | 2.47 [2.38, 2.55] | 2.00 [1.70, 2.32] |
| | | Gay or Lesbian | Heterosexual | 2.30 [2.17, 2.42] | c |
| | | Bisexual | Heterosexual | 2.69 [2.56, 2.82] | c |
| | | Other | Heterosexual | 2.64 [2.40, 2.89] | c |
| | Social rented | LGB+ | Heterosexual | 2.61 [2.53, 2.69] | 2.31 [1.89, 2.74] |
| | | Gay or Lesbian | Heterosexual | 2.66 [2.53, 2.78] | c |
| | | Bisexual | Heterosexual | 2.80 [2.68, 2.91] | c |
| | | Other | Heterosexual | 1.91 [1.72, 2.09] | c |
| | Private rented or living rent free | LGB+ | Heterosexual | 2.31 [2.25, 2.38] | 2.20 [1.86, 2.55] |
| | | Gay or Lesbian | Heterosexual | 2.36 [2.25, 2.48] | c |
| | | Bisexual | Heterosexual | 2.42 [2.32, 2.52] | c |
| | | Other | Heterosexual | 1.90 [1.74, 2.05] | c |
| | | LGB+ | Heterosexual | 2.40 [2.34, 2.46] | 2.20 [1.94, 2.47] |

| | | | | | |
|---|---|---|---|---|---|
| Economic activity | Economically active | Gay or Lesbian | Heterosexual | 2.31 [2.21, 2.40] | 2.23 [1.86, 2.62] |
| | | Bisexual | Heterosexual | 2.58 [2.49, 2.67] | 2.27 [1.81, 2.74] |
| | | Other | Heterosexual | 2.27 [2.10, 2.43] | 2.66 [1.77, 3.55] |
| | Economically inactive | LGB+ | Heterosexual | 2.51 [2.44, 2.58] | 2.14 [1.80, 2.49] |
| | | Gay or Lesbian | Heterosexual | 2.87 [2.75, 3.00] | 2.78 [2.13, 3.44] |
| | | Bisexual | Heterosexual | 2.61 [2.51, 2.71] | 1.89 [1.42, 2.37] |
| | | Other | Heterosexual | 1.73 [1.60, 1.86] | 1.43 [0.78, 2.08] |
| General health (detailed) | Very good health | LGB+ | Heterosexual | 2.10 [2.02, 2.18] | 2.12 [1.73, 2.52] |
| | | Gay or Lesbian | Heterosexual | 2.21 [2.08, 2.33] | c |
| | | Bisexual | Heterosexual | 2.22 [2.09, 2.35] | c |
| | | Other | Heterosexual | 1.58 [1.36, 1.79] | c |
| | Good health | LGB+ | Heterosexual | 2.00 [1.95, 2.06] | 1.79 [1.52, 2.07] |
| | | Gay or Lesbian | Heterosexual | 2.09 [1.99, 2.18] | c |
| | | Bisexual | Heterosexual | 2.09 [2.00, 2.17] | c |
| | | Other | Heterosexual | 1.55 [1.41, 1.68] | c |
| | Fair health | LGB+ | Heterosexual | 1.77 [1.71, 1.83] | 1.77 [1.45, 2.12] |
| | | Gay or Lesbian | Heterosexual | 2.02 [1.91, 2.14] | c |
| | | Bisexual | Heterosexual | 1.79 [1.71, 1.86] | c |
| | | Other | Heterosexual | 1.36 [1.24, 1.48] | c |
| | Bad health | LGB+ | Heterosexual | 1.57 [1.49, 1.66] | 1.48 [1.08, 1.93] |
| | | Gay or Lesbian | Heterosexual | 1.74 [1.60, 1.90] | c |
| | | Bisexual | Heterosexual | 1.59 [1.49, 1.70] | c |
| | | Other | Heterosexual | 1.31 [1.15, 1.46] | c |
| | Very bad health | LGB+ | Heterosexual | 1.71 [1.54, 1.88] | 1.45 [0.80, 2.25] |
| | | Gay or Lesbian | Heterosexual | 1.88 [1.57, 2.20] | c |
| | | Bisexual | Heterosexual | 1.77 [1.54, 2.01] | c |
| | | Other | Heterosexual | 1.35 [1.06, 1.64] | c |
| General health (aggregated) | Good health | LGB+ | Heterosexual | 2.22 [2.17, 2.28] | 2.02 [1.78, 2.26] |
| | | Gay or Lesbian | Heterosexual | 2.25 [2.17, 2.33] | 2.26 [1.91, 2.60] |
| | | Bisexual | Heterosexual | 2.38 [2.30, 2.46] | 1.90 [1.49, 2.32] |
| | | Other | Heterosexual | 1.74 [1.61, 1.87] | 1.79 [1.07, 2.51] |
| | Not good health | LGB+ | Heterosexual | 1.77 [1.72, 1.82] | 1.71 [1.46, 1.97] |
| | | Gay or Lesbian | Heterosexual | 1.99 [1.90, 2.07] | 1.68 [1.29, 2.09] |
| | | Bisexual | Heterosexual | 1.79 [1.72, 1.85] | 1.64 [1.31, 1.99] |
| | | Other | Heterosexual | 1.44 [1.34, 1.53] | 1.94 [1.31, 2.59] |
| Disability | Day-to-day activities limited a lot | LGB+ | Heterosexual | 1.72 [1.66, 1.77] | 1.63 [1.33, 1.95] |
| | | Gay or Lesbian | Heterosexual | 1.87 [1.77, 1.98] | 1.74 [1.24, 2.26] |
| | | Bisexual | Heterosexual | 1.76 [1.68, 1.84] | 1.55 [1.14, 1.98] |
| | | Other | Heterosexual | 1.38 [1.27, 1.49] | 1.46 [0.80, 2.16] |
| | Day-to-day activities limited a little | LGB+ | Heterosexual | 1.41 [1.37, 1.46] | 1.43 [1.18, 1.69] |
| | | Gay or Lesbian | Heterosexual | 1.64 [1.56, 1.72] | 1.82 [1.36, 2.29] |
| | | Bisexual | Heterosexual | 1.43 [1.38, 1.49] | 1.28 [0.92, 1.65] |
| | | Other | Heterosexual | 1.05 [0.96, 1.14] | 1.55 [0.81, 2.31] |
| | Day-to-day activities not limited | LGB+ | Heterosexual | 1.92 [1.86, 1.97] | 1.90 [1.66, 2.15] |
| | | Gay or Lesbian | Heterosexual | 2.02 [1.93, 2.10] | 1.98 [1.64, 2.32] |
| | | Bisexual | Heterosexual | 1.99 [1.91, 2.07] | 1.83 [1.40, 2.26] |
| | | Other | Heterosexual | 1.41 [1.28, 1.54] | 1.87 [1.11, 2.63] |

Notes: Rate ratios were calculated using age-specific rates for the age on Census Day variable, and age-standardised rates for all other variables. Cells with the value 'c' have been suppressed for the purposes of statistical disclosure control. All calculations are based on estimates rounded to the nearest multiple of five for disclosure control reasons. CI: confidence interval.

**Supplementary Table 17: Sensitivity analysis including in-patient admissions from HES and PEDW for self-harm with undetermined intent, crude and age-standardised rates of self-harm per 100,000 people**

| Characteristic | Category | Sexual orientation | Count (%), Individuals | Count (%), Individuals with at least one hospital admission or emergency attendance for intentional self-harm | Crude rate of self-harm [95% CI] | Age-standardised rate of self-harm [95% CI] |
|---|---|---|---|---|---|---|
| Total | Total | Total | 28,659,465 (100.0) | 136,370 (100.0) | 475.83 [473.31, 478.36] | 673.71 [669.56, 677.87] |
| | | Heterosexual | 27,554,145 (96.1) | 119,625 (87.7) | 434.15 [431.69, 436.61] | 602.79 [598.72, 606.85] |
| | | LGB+ | 1,105,325 (3.9) | 16,745 (12.3) | 1,514.94 [1,492.08, 1,538.06] | 1,515.08 [1,492.14, 1,538.03] |
| | | Gay or Lesbian | 524,585 (1.8) | 6,225 (4.6) | 1,186.65 [1,157.36, 1,216.50] | 1,457.59 [1,419.43, 1,495.75] |
| | | Bisexual | 457,520 (1.6) | 8,770 (6.4) | 1,916.86 [1,876.95, 1,957.40] | 1,676.46 [1,639.62, 1,713.31] |
| | | Other | 123,220 (0.4) | 1,755 (1.3) | 1,424.28 [1,358.42, 1,492.51] | 1,343.14 [1,279.72, 1,406.55] |
| Age on Census Day (10-year bands) | 16 to 24 years | Total | 2,849,490 (100.0) | 34,345 (100.0) | 1,205.30 [1,192.56, 1,218.05] | - |
| | | Heterosexual | 2,564,750 (90.0) | 26,275 (76.5) | 1,024.47 [1,012.08, 1,036.85] | - |
| | | LGB+ | 284,740 (10.0) | 8,070 (23.5) | 2,834.16 [2,772.33, 2,896.00] | - |
| | | Gay or Lesbian | 78,410 (2.8) | 2,240 (6.5) | 2,856.78 [2,738.47, 2,975.08] | - |
| | | Bisexual | 171,285 (6.0) | 4,945 (14.4) | 2,887.00 [2,806.53, 2,967.47] | - |
| | | Other | 35,045 (1.2) | 890 (2.6) | 2,539.59 [2,372.74, 2,706.44] | - |
| | 25 to 34 years | Total | 5,049,945 (100.0) | 31,730 (100.0) | 628.32 [621.41, 635.24] | - |
| | | Heterosexual | 4,710,035 (93.3) | 26,865 (84.7) | 570.38 [563.56, 577.20] | - |
| | | LGB+ | 339,910 (6.7) | 4,865 (15.3) | 1,431.26 [1,391.04, 1,471.48] | - |
| | | Gay or Lesbian | 149,275 (3.0) | 2,000 (6.3) | 1,339.81 [1,281.09, 1,398.53] | - |
| | | Bisexual | 148,645 (2.9) | 2,335 (7.4) | 1,570.86 [1,507.14, 1,634.57] | - |
| | | Other | 41,990 (0.8) | 530 (1.7) | 1,262.21 [1,154.74, 1,369.67] | - |
| | 35 to 44 years | Total | 4,949,735 (100.0) | 23,450 (100.0) | 473.76 [467.70, 479.83] | - |
| | | Heterosexual | 4,746,265 (95.9) | 21,415 (91.3) | 451.20 [445.15, 457.24] | - |
| | | LGB+ | 203,465 (4.1) | 2,035 (8.7) | 1,000.17 [956.72, 1,043.63] | - |
| | | Gay or Lesbian | 110,930 (2.2) | 985 (4.2) | 887.95 [832.49, 943.40] | - |
| | | Bisexual | 69,180 (1.4) | 860 (3.7) | 1,243.13 [1,160.05, 1,326.22] | - |
| | | Other | 23,355 (0.5) | 190 (0.8) | 813.53 [697.85, 929.21] | - |
| | 45 to 54 years | Total | 5,086,860 (100.0) | 20,855 (100.0) | 409.98 [404.41, 415.54] | - |
| | | Heterosexual | 4,945,310 (97.2) | 19,695 (94.4) | 398.26 [392.69, 403.82] | - |
| | | LGB+ | 141,550 (2.8) | 1,160 (5.6) | 819.50 [772.34, 866.66] | - |
| | | Gay or Lesbian | 91,870 (1.8) | 620 (3.0) | 674.87 [621.74, 727.99] | - |
| | | Bisexual | 36,955 (0.7) | 440 (2.1) | 1,190.64 [1,079.38, 1,301.89] | - |
| | | Other | 12,725 (0.3) | 100 (0.5) | 785.85 [631.83, 939.88] | - |
| | 55 to 64 years | Total | 4,793,020 (100.0) | 13,015 (100.0) | 271.54 [266.88, 276.21] | - |
| | | Heterosexual | 4,704,060 (98.1) | 12,560 (96.5) | 267.00 [262.33, 271.67] | - |
| | | LGB+ | 88,960 (1.9) | 455 (3.5) | 511.47 [464.47, 558.46] | - |
| | | Gay or Lesbian | 62,355 (1.3) | 275 (2.1) | 441.02 [388.90, 493.15] | - |
| | | Bisexual | 20,095 (0.4) | 145 (1.1) | 721.57 [604.12, 839.02] | - |
| | | Other | 6,510 (0.1) | 35 (0.3) | 537.63 [374.48, 747.72] | - |

| | | | | | | |
|---|---|---|---|---|---|---|
| | 65 years and over | Total | 5,930,420 (100.0) | 12,970 (100.0) | 218.70 [214.94, 222.47] | - |
| | | Heterosexual | 5,883,725 (99.2) | 12,815 (98.8) | 217.80 [214.03, 221.58] | - |
| | | LGB+ | 46,695 (0.8) | 160 (1.2) | 342.65 [289.56, 395.74] | - |
| | | Gay or Lesbian | 31,745 (0.5) | 100 (0.8) | 315.01 [253.27, 376.75] | - |
| | | Bisexual | 11,365 (0.2) | 45 (0.3) | 395.95 [288.81, 529.82] | - |
| | | Other | 3,590 (0.1) | 10 (0.1) | 278.55 [133.58, 512.27] | - |
| Sex | Female | Total | 15,400,570 (100.0) | 80,700 (100.0) | 524.01 [520.40, 527.63] | 762.60 [756.52, 768.68] |
| | | Heterosexual | 14,795,975 (96.1) | 68,620 (85.0) | 463.77 [460.31, 467.26] | 658.24 [652.36, 664.12] |
| | | LGB+ | 604,590 (3.9) | 12,080 (15.0) | 1,998.05 [1,962.57, 2,034.00] | 1,807.45 [1,774.75, 1,840.14] |
| | | Gay or Lesbian | 199,830 (1.3) | 3,470 (4.3) | 1,736.48 [1,679.17, 1,795.23] | 1,996.42 [1,928.63, 2,064.21] |
| | | Bisexual | 326,810 (2.1) | 7,265 (9.0) | 2,223.00 [2,172.18, 2,274.72] | 1,864.71 [1,817.65, 1,911.76] |
| | | Other | 77,950 (0.5) | 1,345 (1.7) | 1,725.47 [1,634.47, 1,820.20] | 1,544.65 [1,459.91, 1,629.39] |
| | Male | Total | 13,258,895 (100.0) | 55,670 (100.0) | 419.87 [416.39, 423.37] | 571.80 [566.21, 577.38] |
| | | Heterosexual | 12,758,165 (96.2) | 51,000 (91.6) | 399.74 [396.28, 403.23] | 543.14 [537.52, 548.75] |
| | | LGB+ | 500,730 (3.8) | 4,665 (8.4) | 931.64 [905.10, 958.77] | 1,033.36 [1,002.89, 1,063.84] |
| | | Gay or Lesbian | 324,755 (2.4) | 2,755 (4.9) | 848.33 [816.95, 880.61] | 1,026.53 [984.36, 1,068.70] |
| | | Bisexual | 130,710 (1.0) | 1,505 (2.7) | 1,151.40 [1,093.96, 1,211.08] | 1,131.66 [1,074.01, 1,189.31] |
| | | Other | 45,265 (0.3) | 405 (0.7) | 894.73 [809.70, 986.26] | 926.29 [835.66, 1,016.91] |
| Region | England: North East | Total | 1,330,595 (100.0) | 8,295 (100.0) | 623.41 [610.06, 636.97] | 950.51 [927.05, 973.98] |
| | | Heterosexual | 1,283,770 (96.5) | 7,360 (88.7) | 573.31 [560.29, 586.56] | 872.72 [849.36, 896.09] |
| | | LGB+ | 46,825 (3.5) | 935 (11.3) | 1,996.80 [1,870.84, 2,129.00] | 1,941.10 [1,816.35, 2,065.85] |
| | | Gay or Lesbian | 23,665 (1.8) | 425 (5.1) | 1,795.90 [1,629.20, 1,975.03] | 2,023.30 [1,825.67, 2,220.93] |
| | | Bisexual | 18,945 (1.4) | 420 (5.1) | 2,216.94 [2,009.97, 2,439.45] | 1,983.61 [1,774.05, 2,193.18] |
| | | Other | 4,215 (0.3) | 90 (1.1) | 2,135.23 [1,716.98, 2,624.56] | 2,031.35 [1,626.28, 2,505.26] |
| | England: North West | Total | 3,659,340 (100.0) | 16,645 (100.0) | 454.86 [447.98, 461.83] | 652.76 [641.21, 664.31] |
| | | Heterosexual | 3,521,115 (96.2) | 14,450 (86.8) | 410.38 [403.72, 417.13] | 575.20 [564.00, 586.40] |
| | | LGB+ | 138,225 (3.8) | 2,195 (13.2) | 1,587.99 [1,522.25, 1,655.84] | 1,577.18 [1,511.20, 1,643.16] |
| | | Gay or Lesbian | 71,195 (1.9) | 955 (5.7) | 1,341.39 [1,257.65, 1,429.23] | 1,557.02 [1,453.74, 1,660.30] |
| | | Bisexual | 53,820 (1.5) | 1,065 (6.4) | 1,978.82 [1,861.74, 2,101.33] | 1,753.04 [1,641.08, 1,865.00] |
| | | Other | 13,210 (0.4) | 175 (1.1) | 1,324.75 [1,135.74, 1,536.23] | 1,240.13 [1,055.34, 1,424.92] |
| | England: Yorkshire and The Humber | Total | 2,627,670 (100.0) | 18,300 (100.0) | 696.43 [686.38, 706.60] | 906.63 [891.20, 922.07] |
| | | Heterosexual | 2,529,495 (96.3) | 16,460 (89.9) | 650.72 [640.82, 660.74] | 833.69 [818.35, 849.02] |
| | | LGB+ | 98,175 (3.7) | 1,840 (10.1) | 1,874.20 [1,789.54, 1,961.84] | 1,804.01 [1,721.15, 1,886.88] |
| | | Gay or Lesbian | 44,200 (1.7) | 700 (3.8) | 1,583.71 [1,468.55, 1,705.51] | 1,767.74 [1,633.45, 1,902.02] |
| | | Bisexual | 42,885 (1.6) | 950 (5.2) | 2,215.23 [2,076.58, 2,360.69] | 1,951.15 [1,816.01, 2,086.29] |
| | | Other | 11,085 (0.4) | 190 (1.0) | 1,714.03 [1,478.96, 1,975.84] | 1,598.79 [1,367.50, 1,830.08] |
| | England: East Midlands | Total | 2,290,650 (100.0) | 12,310 (100.0) | 537.40 [527.95, 546.98] | 783.12 [767.42, 798.82] |
| | | Heterosexual | 2,211,910 (96.6) | 10,800 (87.7) | 488.27 [479.10, 497.56] | 701.76 [686.42, 717.11] |
| | | LGB+ | 78,740 (3.4) | 1,510 (12.3) | 1,917.70 [1,822.19, 2,016.93] | 1,835.20 [1,742.18, 1,928.23] |
| | | Gay or Lesbian | 34,430 (1.5) | 525 (4.3) | 1,524.83 [1,397.17, 1,661.02] | 1,780.99 [1,624.97, 1,937.01] |
| | | Bisexual | 35,525 (1.6) | 840 (6.8) | 2,364.53 [2,207.31, 2,529.99] | 2,038.71 [1,891.95, 2,185.48] |
| | | Other | 8,785 (0.4) | 150 (1.2) | 1,707.46 [1,445.15, 2,003.61] | 1,510.46 [1,264.07, 1,756.85] |
| | England: West Midlands | Total | 2,684,810 (100.0) | 10,500 (100.0) | 391.09 [383.64, 398.64] | 558.97 [546.80, 571.14] |
| | | Heterosexual | 2,600,505 (96.9) | 9,255 (88.1) | 355.89 [348.68, 363.22] | 500.45 [488.62, 512.27] |
| | | LGB+ | 84,310 (3.1) | 1,245 (11.9) | 1,476.69 [1,395.80, 1,561.06] | 1,429.27 [1,349.66, 1,508.88] |

| | | | | | | |
|---|---|---|---|---|---|---|
| | | Gay or Lesbian | 38,695 (1.4) | 470 (4.5) | 1,214.63 [1,107.28, 1,329.57] | 1,428.48 [1,295.70, 1,561.26] |
| | | Bisexual | 35,760 (1.3) | 655 (6.2) | 1,831.66 [1,694.05, 1,977.46] | 1,569.03 [1,443.81, 1,694.25] |
| | | Other | 9,855 (0.4) | 115 (1.1) | 1,166.92 [963.41, 1,400.71] | 1,085.39 [884.12, 1,286.66] |
| | England: East of England | Total | 3,121,770 (100.0) | 15,115 (100.0) | 484.18 [476.49, 491.96] | 711.89 [698.11, 725.66] |
| | | Heterosexual | 3,022,385 (96.8) | 13,415 (88.8) | 443.85 [436.38, 451.43] | 634.88 [621.47, 648.30] |
| | | LGB+ | 99,385 (3.2) | 1,705 (11.3) | 1,715.55 [1,635.08, 1,798.96] | 1,704.58 [1,623.48, 1,785.67] |
| | | Gay or Lesbian | 44,105 (1.4) | 575 (3.8) | 1,303.71 [1,199.31, 1,414.76] | 1,578.32 [1,444.30, 1,712.33] |
| | | Bisexual | 43,470 (1.4) | 935 (6.2) | 2,150.91 [2,015.23, 2,293.32] | 1,911.49 [1,784.17, 2,038.82] |
| | | Other | 11,810 (0.4) | 190 (1.3) | 1,608.81 [1,388.17, 1,854.54] | 1,546.23 [1,324.43, 1,768.04] |
| | England: London | Total | 4,329,065 (100.0) | 16,800 (100.0) | 388.07 [382.23, 393.99] | 472.21 [464.01, 480.41] |
| | | Heterosexual | 4,096,810 (94.6) | 14,815 (88.2) | 361.62 [355.82, 367.49] | 430.22 [422.15, 438.29] |
| | | LGB+ | 232,255 (5.4) | 1,980 (11.8) | 852.51 [815.37, 890.91] | 986.53 [941.17, 1,031.90] |
| | | Gay or Lesbian | 119,415 (2.8) | 745 (4.4) | 623.87 [579.87, 670.33] | 900.51 [820.28, 980.74] |
| | | Bisexual | 83,985 (1.9) | 960 (5.7) | 1,143.06 [1,071.89, 1,217.72] | 1,104.06 [1,032.11, 1,176.01] |
| | | Other | 28,855 (0.7) | 275 (1.6) | 953.04 [843.72, 1,072.60] | 1,013.90 [890.57, 1,137.23] |
| | England: South East | Total | 4,642,855 (100.0) | 20,680 (100.0) | 445.42 [439.37, 451.53] | 690.58 [679.38, 701.77] |
| | | Heterosexual | 4,470,840 (96.3) | 17,935 (86.7) | 401.16 [395.31, 407.07] | 605.56 [594.69, 616.44] |
| | | LGB+ | 172,015 (3.7) | 2,745 (13.3) | 1,595.79 [1,536.65, 1,656.63] | 1,598.27 [1,538.34, 1,658.21] |
| | | Gay or Lesbian | 80,120 (1.7) | 905 (4.4) | 1,129.56 [1,057.15, 1,205.61] | 1,453.43 [1,354.05, 1,552.81] |
| | | Bisexual | 73,040 (1.6) | 1,540 (7.4) | 2,108.43 [2,004.43, 2,216.43] | 1,785.34 [1,692.01, 1,878.66] |
| | | Other | 18,850 (0.4) | 305 (1.5) | 1,618.04 [1,441.53, 1,810.20] | 1,474.36 [1,306.11, 1,642.61] |
| | England: South West | Total | 2,760,745 (100.0) | 12,960 (100.0) | 469.44 [461.39, 477.59] | 755.65 [740.77, 770.53] |
| | | Heterosexual | 2,653,295 (96.1) | 10,980 (84.7) | 413.83 [406.12, 421.64] | 658.03 [643.54, 672.52] |
| | | LGB+ | 107,450 (3.9) | 1,980 (15.3) | 1,842.72 [1,762.44, 1,925.71] | 1,783.91 [1,704.76, 1,863.05] |
| | | Gay or Lesbian | 46,135 (1.7) | 670 (5.2) | 1,452.26 [1,344.36, 1,566.51] | 1,808.56 [1,668.15, 1,948.96] |
| | | Bisexual | 49,505 (1.8) | 1,085 (8.4) | 2,191.70 [2,063.21, 2,326.09] | 1,889.08 [1,769.27, 2,008.89] |
| | | Other | 11,805 (0.4) | 220 (1.7) | 1,863.62 [1,625.48, 2,126.83] | 1,675.30 [1,448.98, 1,901.62] |
| | Wales | Total | 1,211,965 (100.0) | 4,770 (100.0) | 393.58 [382.49, 404.91] | 584.28 [565.95, 602.61] |
| | | Heterosexual | 1,164,015 (96.0) | 4,160 (87.2) | 357.38 [346.61, 368.41] | 532.06 [513.86, 550.25] |
| | | LGB+ | 47,950 (4.0) | 610 (12.8) | 1,272.16 [1,173.19, 1,377.24] | 1,199.23 [1,103.40, 1,295.06] |
| | | Gay or Lesbian | 22,625 (1.9) | 245 (5.1) | 1,082.87 [951.51, 1,227.30] | 1,263.93 [1,101.24, 1,426.61] |
| | | Bisexual | 20,585 (1.7) | 320 (6.7) | 1,554.53 [1,388.85, 1,734.53] | 1,325.93 [1,167.09, 1,484.77] |
| | | Other | 4,745 (0.4) | 40 (0.8) | 842.99 [602.25, 1,147.92] | 731.29 [521.15, 997.46] |
| Ethnic group | Asian, Asian British or Asian Welsh | Total | 2,386,835 (100.0) | 7,105 (100.0) | 297.67 [290.79, 304.68] | 340.88 [332.37, 349.38] |
| | | Heterosexual | 2,328,580 (97.6) | 6,685 (94.1) | 287.08 [280.24, 294.05] | 327.98 [319.51, 336.44] |
| | | LGB+ | 58,255 (2.4) | 415 (5.8) | 712.39 [645.49, 784.33] | 738.09 [666.03, 810.14] |
| | | Gay or Lesbian | 17,395 (0.7) | 125 (1.8) | 718.60 [598.15, 856.18] | 855.58 [695.01, 1,016.15] |
| | | Bisexual | 30,175 (1.3) | 225 (3.2) | 745.65 [651.40, 849.71] | 694.49 [601.28, 787.70] |
| | | Other | 10,690 (0.4) | 70 (1.0) | 654.82 [510.46, 827.32] | 740.09 [569.94, 943.42] |
| | Black, Black British, Black Welsh, Caribbean or African | Total | 1,035,960 (100.0) | 3,810 (100.0) | 367.77 [356.19, 379.64] | 457.28 [441.42, 473.15] |
| | | Heterosexual | 1,011,020 (97.6) | 3,515 (92.3) | 347.67 [336.27, 359.36] | 427.35 [411.81, 442.89] |
| | | LGB+ | 24,940 (2.4) | 295 (7.7) | 1,182.84 [1,051.70, 1,325.81] | 1,249.92 [1,106.75, 1,393.10] |
| | | Gay or Lesbian | 7,395 (0.7) | 80 (2.1) | 1,081.81 [857.81, 1,346.41] | 1,314.41 [1,032.15, 1,647.83] |
| | | Bisexual | 11,590 (1.1) | 160 (4.2) | 1,380.50 [1,174.88, 1,611.75] | 1,268.74 [1,068.53, 1,468.95] |
| | | Other | 5,955 (0.6) | 60 (1.6) | 1,007.56 [768.87, 1,296.93] | 1,195.95 [906.40, 1,547.00] |

| | | | | | | |
|---|---|---|---|---|---|---|
| | Mixed or Multiple ethnic groups | Total | 531,980 (100.0) | 4,245 (100.0) | 797.96 [774.14, 822.34] | 828.93 [803.71, 854.14] |
| | | Heterosexual | 488,015 (91.7) | 3,365 (79.3) | 689.53 [666.43, 713.23] | 721.57 [696.80, 746.34] |
| | | LGB+ | 43,965 (8.3) | 875 (20.6) | 1,990.22 [1,860.52, 2,126.58] | 1,674.58 [1,559.59, 1,789.58] |
| | | Gay or Lesbian | 16,670 (3.1) | 255 (6.0) | 1,529.69 [1,347.69, 1,729.43] | 1,549.82 [1,358.43, 1,741.21] |
| | | Bisexual | 21,210 (4.0) | 515 (12.1) | 2,428.10 [2,222.89, 2,647.15] | 1,838.51 [1,652.96, 2,024.06] |
| | | Other | 6,085 (1.1) | 105 (2.5) | 1,725.55 [1,411.33, 2,088.89] | 1,447.42 [1,156.59, 1,738.25] |
| | White | Total | 24,127,370 (100.0) | 119,210 (100.0) | 494.09 [491.29, 496.90] | 743.63 [738.60, 748.66] |
| | | Heterosexual | 23,172,720 (96.0) | 104,225 (87.4) | 449.77 [447.05, 452.51] | 663.75 [658.81, 668.70] |
| | | LGB+ | 954,650 (4.0) | 14,990 (12.6) | 1,570.21 [1,545.17, 1,595.55] | 1,578.76 [1,553.48, 1,604.04] |
| | | Gay or Lesbian | 475,725 (2.0) | 5,715 (4.8) | 1,201.32 [1,170.38, 1,232.88] | 1,490.91 [1,450.05, 1,531.78] |
| | | Bisexual | 385,885 (1.6) | 7,795 (6.5) | 2,020.03 [1,975.43, 2,065.38] | 1,781.15 [1,739.32, 1,822.99] |
| | | Other | 93,035 (0.4) | 1,480 (1.2) | 1,590.80 [1,510.78, 1,673.96] | 1,472.14 [1,395.81, 1,548.46] |
| | Other ethnic group | Total | 577,320 (100.0) | 2,000 (100.0) | 346.43 [331.41, 361.95] | 399.87 [380.74, 418.99] |
| | | Heterosexual | 553,810 (95.9) | 1,830 (91.5) | 330.44 [315.47, 345.93] | 379.68 [360.58, 398.79] |
| | | LGB+ | 23,510 (4.1) | 170 (8.5) | 723.10 [618.48, 840.34] | 820.35 [693.04, 947.66] |
| | | Gay or Lesbian | 7,395 (1.3) | 50 (2.5) | 676.13 [501.84, 891.40] | 1,005.10 [711.00, 1,368.34] |
| | | Bisexual | 8,655 (1.5) | 75 (3.8) | 866.55 [681.60, 1,086.23] | 873.40 [687.30, 1,094.44] |
| | | Other | 7,460 (1.3) | 45 (2.2) | 603.22 [439.99, 807.15] | 609.05 [429.71, 833.14] |
| Rural/urban classification | Urban | Total | 23,366,335 (100.0) | 116,635 (100.0) | 499.16 [496.30, 502.03] | 679.08 [674.61, 683.54] |
| | | Heterosexual | 22,395,100 (95.8) | 101,970 (87.4) | 455.32 [452.53, 458.13] | 608.05 [603.67, 612.42] |
| | | LGB+ | 971,235 (4.2) | 14,665 (12.6) | 1,509.93 [1,485.59, 1,534.57] | 1,500.38 [1,476.08, 1,524.67] |
| | | Gay or Lesbian | 457,000 (2.0) | 5,455 (4.7) | 1,193.65 [1,162.19, 1,225.76] | 1,447.95 [1,407.46, 1,488.44] |
| | | Bisexual | 403,390 (1.7) | 7,655 (6.6) | 1,897.67 [1,855.39, 1,940.66] | 1,655.38 [1,616.21, 1,694.54] |
| | | Other | 110,845 (0.5) | 1,560 (1.3) | 1,407.37 [1,338.39, 1,478.98] | 1,329.48 [1,262.77, 1,396.18] |
| | Rural | Total | 5,293,135 (100.0) | 19,735 (100.0) | 372.84 [367.66, 378.08] | 650.08 [638.58, 661.58] |
| | | Heterosexual | 5,159,045 (97.5) | 17,655 (89.5) | 342.21 [337.18, 347.30] | 580.36 [569.18, 591.53] |
| | | LGB+ | 134,090 (2.5) | 2,080 (10.5) | 1,551.20 [1,485.24, 1,619.33] | 1,648.05 [1,575.74, 1,720.36] |
| | | Gay or Lesbian | 67,585 (1.3) | 770 (3.9) | 1,139.31 [1,060.24, 1,222.70] | 1,549.10 [1,431.04, 1,667.15] |
| | | Bisexual | 54,130 (1.0) | 1,115 (5.6) | 2,059.86 [1,940.71, 2,184.40] | 1,851.86 [1,739.11, 1,964.62] |
| | | Other | 12,370 (0.2) | 195 (1.0) | 1,576.39 [1,362.89, 1,813.86] | 1,497.06 [1,282.64, 1,711.47] |
| Country of birth | Born in the UK | Total | 23,138,360 (100.0) | 120,950 (100.0) | 522.73 [519.78, 525.68] | 770.65 [765.63, 775.67] |
| | | Heterosexual | 22,260,930 (96.2) | 105,720 (87.4) | 474.91 [472.05, 477.78] | 689.11 [684.18, 694.03] |
| | | LGB+ | 877,430 (3.8) | 15,230 (12.6) | 1,735.75 [1,708.29, 1,763.54] | 1,711.88 [1,684.64, 1,739.12] |
| | | Gay or Lesbian | 433,385 (1.9) | 5,740 (4.7) | 1,324.46 [1,290.41, 1,359.17] | 1,608.65 [1,565.28, 1,652.02] |
| | | Bisexual | 360,775 (1.6) | 7,970 (6.6) | 2,209.13 [2,160.90, 2,258.18] | 1,921.87 [1,876.72, 1,967.02] |
| | | Other | 83,265 (0.4) | 1,520 (1.3) | 1,825.50 [1,734.87, 1,919.63] | 1,647.62 [1,562.68, 1,732.55] |
| | Born outside the UK | Total | 5,521,105 (100.0) | 15,420 (100.0) | 279.29 [274.90, 283.74] | 343.34 [336.91, 349.76] |
| | | Heterosexual | 5,293,215 (95.9) | 13,905 (90.2) | 262.69 [258.35, 267.10] | 314.86 [308.56, 321.16] |
| | | LGB+ | 227,895 (4.1) | 1,515 (9.8) | 664.78 [631.72, 699.12] | 740.36 [701.98, 778.74] |
| | | Gay or Lesbian | 91,200 (1.7) | 480 (3.1) | 526.32 [480.28, 575.58] | 704.70 [629.42, 779.99] |
| | | Bisexual | 96,745 (1.8) | 800 (5.2) | 826.92 [770.60, 886.26] | 803.52 [746.56, 860.49] |
| | | Other | 39,950 (0.7) | 235 (1.5) | 588.24 [515.43, 668.45] | 654.88 [568.16, 741.60] |
| English language proficiency | Main language is English | Total | 25,978,125 (100.0) | 129,835 (100.0) | 499.79 [497.07, 502.51] | 727.38 [722.78, 731.97] |
| | | Heterosexual | 24,976,765 (96.1) | 113,595 (87.5) | 454.80 [452.16, 457.46] | 650.24 [645.74, 654.75] |
| | | LGB+ | 1,001,360 (3.9) | 16,240 (12.5) | 1,621.79 [1,596.95, 1,646.93] | 1,616.30 [1,591.43, 1,641.17] |

| | | | | | | |
|---|---|---|---|---|---|---|
| | | Gay or Lesbian | 491,205 (1.9) | 6,085 (4.7) | 1,238.79 [1,207.86, 1,270.31] | 1,528.72 [1,488.35, 1,569.08] |
| | | Bisexual | 411,395 (1.6) | 8,515 (6.6) | 2,069.79 [2,026.06, 2,114.22] | 1,803.36 [1,762.78, 1,843.94] |
| | | Other | 98,760 (0.4) | 1,640 (1.3) | 1,660.59 [1,581.19, 1,742.95] | 1,519.66 [1,444.95, 1,594.37] |
| | Main language is not English | Total | 2,681,340 (100.0) | 6,535 (100.0) | 243.72 [237.85, 249.70] | 281.47 [273.55, 289.38] |
| | | Heterosexual | 2,577,380 (96.1) | 6,030 (92.3) | 233.96 [228.09, 239.94] | 265.79 [257.93, 273.64] |
| | | LGB+ | 103,965 (3.9) | 505 (7.7) | 485.74 [444.29, 530.01] | 541.77 [491.83, 591.72] |
| | | Gay or Lesbian | 33,380 (1.2) | 140 (2.1) | 419.41 [352.82, 494.92] | 585.61 [466.59, 704.63] |
| | | Bisexual | 46,125 (1.7) | 255 (3.9) | 552.85 [487.07, 625.03] | 554.25 [483.91, 624.60] |
| | | Other | 24,460 (0.9) | 115 (1.8) | 470.16 [388.16, 564.35] | 504.71 [406.28, 603.14] |
| Religion | No religion | Total | 10,732,605 (100.0) | 69,240 (100.0) | 645.14 [640.34, 649.96] | 802.70 [796.29, 809.12] |
| | | Heterosexual | 10,031,320 (93.5) | 57,855 (83.6) | 576.74 [572.05, 581.46] | 715.75 [709.36, 722.14] |
| | | LGB+ | 701,285 (6.5) | 11,385 (16.4) | 1,623.45 [1,593.76, 1,653.55] | 1,492.79 [1,465.06, 1,520.52] |
| | | Gay or Lesbian | 328,620 (3.1) | 4,140 (6.0) | 1,259.81 [1,221.73, 1,298.79] | 1,415.59 [1,371.34, 1,459.84] |
| | | Bisexual | 305,285 (2.8) | 6,135 (8.9) | 2,009.60 [1,959.62, 2,060.53] | 1,670.85 [1,623.67, 1,718.02] |
| | | Other | 67,380 (0.6) | 1,105 (1.6) | 1,639.95 [1,544.67, 1,739.57] | 1,411.26 [1,323.52, 1,498.99] |
| | Christian | Total | 14,299,140 (100.0) | 52,600 (100.0) | 367.85 [364.72, 371.01] | 582.25 [575.42, 589.07] |
| | | Heterosexual | 14,022,860 (98.1) | 49,055 (93.3) | 349.82 [346.73, 352.93] | 538.83 [532.16, 545.49] |
| | | LGB+ | 276,275 (1.9) | 3,550 (6.7) | 1,284.95 [1,243.03, 1,327.93] | 1,586.57 [1,531.53, 1,641.61] |
| | | Gay or Lesbian | 157,940 (1.1) | 1,580 (3.0) | 1,000.38 [951.66, 1,050.95] | 1,528.12 [1,437.78, 1,618.46] |
| | | Bisexual | 91,305 (0.6) | 1,670 (3.2) | 1,829.03 [1,742.35, 1,918.91] | 1,853.57 [1,764.20, 1,942.94] |
| | | Other | 27,035 (0.2) | 295 (0.6) | 1,091.18 [970.20, 1,223.07] | 1,179.38 [1,042.60, 1,316.16] |
| | Buddhist | Total | 149,690 (100.0) | 695 (100.0) | 464.29 [430.41, 500.13] | 620.81 [565.26, 676.37] |
| | | Heterosexual | 136,525 (91.2) | 500 (71.9) | 366.23 [334.83, 399.79] | 448.01 [398.72, 497.30] |
| | | LGB+ | 13,165 (8.8) | 195 (28.1) | 1,481.20 [1,280.59, 1,704.32] | 1,794.28 [1,527.59, 2,060.98] |
| | | Gay or Lesbian | 5,640 (3.8) | 60 (8.6) | 1,063.83 [811.81, 1,369.36] | 1,416.08 [1,004.54, 1,915.01] |
| | | Bisexual | 5,725 (3.8) | 100 (14.4) | 1,746.72 [1,421.21, 2,124.49] | 1,898.34 [1,523.59, 2,273.09] |
| | | Other | 1,800 (1.2) | 35 (5.0) | 1,944.44 [1,354.38, 2,704.25] | 1,998.98 [1,371.42, 2,807.08] |
| | Hindu | Total | 509,375 (100.0) | 1,240 (100.0) | 243.44 [230.07, 257.37] | 300.57 [280.92, 320.21] |
| | | Heterosexual | 498,540 (97.9) | 1,180 (95.2) | 236.69 [223.38, 250.59] | 289.21 [269.72, 308.71] |
| | | LGB+ | 10,835 (2.1) | 60 (4.8) | 553.76 [422.58, 712.80] | 688.92 [519.88, 893.87] |
| | | Gay or Lesbian | 1,945 (0.4) | 15 (1.2) | 771.21 [431.64, 1,271.99] | 910.61 [426.43, 1,624.65] |
| | | Bisexual | 7,135 (1.4) | 40 (3.2) | 560.62 [400.51, 763.40] | 696.73 [493.31, 954.37] |
| | | Other | 1,750 (0.3) | 5 (0.4) | 285.71 [92.77, 666.76] | 524.28 [198.57, 1,167.53] |
| | Jewish | Total | 126,115 (100.0) | 565 (100.0) | 448.00 [411.82, 486.52] | 597.04 [529.23, 664.86] |
| | | Heterosexual | 119,135 (94.5) | 475 (84.1) | 398.71 [363.65, 436.23] | 469.98 [406.83, 533.13] |
| | | LGB+ | 6,975 (5.5) | 90 (15.9) | 1,290.32 [1,037.57, 1,586.03] | 1,433.35 [1,149.41, 1,765.55] |
| | | Gay or Lesbian | 3,385 (2.7) | 30 (5.3) | 886.26 [597.96, 1,265.20] | 1,189.85 [768.24, 1,743.98] |
| | | Bisexual | 2,735 (2.2) | 50 (8.8) | 1,828.15 [1,356.89, 2,410.19] | 1,711.37 [1,260.44, 2,268.30] |
| | | Other | 860 (0.7) | 10 (1.8) | 1,162.79 [557.60, 2,138.41] | 1,182.54 [538.04, 2,221.55] |
| | Muslim | Total | 1,509,160 (100.0) | 5,395 (100.0) | 357.48 [348.01, 367.15] | 392.08 [381.04, 403.12] |
| | | Heterosexual | 1,482,735 (98.2) | 5,200 (96.4) | 350.70 [341.24, 360.37] | 384.76 [373.73, 395.79] |
| | | LGB+ | 26,425 (1.8) | 195 (3.6) | 737.94 [637.99, 849.10] | 879.36 [748.65, 1,010.07] |
| | | Gay or Lesbian | 3,975 (0.3) | 45 (0.8) | 1,132.08 [825.74, 1,514.81] | 1,309.97 [921.50, 1,795.34] |
| | | Bisexual | 12,780 (0.8) | 105 (1.9) | 821.60 [671.98, 994.59] | 923.45 [737.11, 1,109.79] |
| | | Other | 9,670 (0.6) | 50 (0.9) | 517.06 [383.77, 681.68] | 636.12 [452.16, 863.33] |

| | | | | | | |
|---|---|---|---|---|---|---|
| | Sikh | Total | 249,655 (100.0) | 830 (100.0) | 332.46 [310.22, 355.87] | 375.85 [346.63, 405.07] |
| | | Heterosexual | 245,185 (98.2) | 800 (96.4) | 326.28 [304.06, 349.70] | 365.24 [336.14, 394.34] |
| | | LGB+ | 4,475 (1.8) | 35 (4.2) | 782.12 [544.78, 1,087.74] | 793.17 [530.15, 1,131.85] |
| | | Gay or Lesbian | 835 (0.3) | 10 (1.2) | 1,197.60 [574.30, 2,202.44] | 1,508.04 [713.63, 2,788.71] |
| | | Bisexual | 2,355 (0.9) | 10 (1.2) | 424.63 [203.63, 780.91] | 487.49 [244.28, 879.56] |
| | | Other | 1,285 (0.5) | 10 (1.2) | 778.21 [373.18, 1,431.16] | 959.24 [390.16, 1,876.67] |
| | Other religion | Total | 201,225 (100.0) | 1,745 (100.0) | 867.19 [826.97, 908.85] | 1,221.90 [1,157.10, 1,286.71] |
| | | Heterosexual | 165,730 (82.4) | 975 (55.9) | 588.31 [551.95, 626.42] | 753.83 [693.16, 814.51] |
| | | LGB+ | 35,495 (17.6) | 770 (44.1) | 2,169.32 [2,018.78, 2,328.11] | 2,106.58 [1,956.65, 2,256.52] |
| | | Gay or Lesbian | 8,455 (4.2) | 170 (9.7) | 2,010.64 [1,719.75, 2,336.65] | 2,223.87 [1,879.25, 2,568.49] |
| | | Bisexual | 17,955 (8.9) | 410 (23.5) | 2,283.49 [2,067.78, 2,515.58] | 2,125.78 [1,915.63, 2,335.93] |
| | | Other | 9,090 (4.5) | 190 (10.9) | 2,090.21 [1,803.55, 2,409.48] | 1,981.65 [1,695.83, 2,267.47] |
| | Not answered | Total | 882,505 (100.0) | 4,060 (100.0) | 460.05 [446.01, 474.43] | 640.28 [617.33, 663.23] |
| | | Heterosexual | 852,110 (96.6) | 3,585 (88.3) | 420.72 [407.06, 434.72] | 574.38 [552.02, 596.75] |
| | | LGB+ | 30,395 (3.4) | 475 (11.7) | 1,562.76 [1,425.36, 1,709.82] | 1,573.57 [1,431.95, 1,715.19] |
| | | Gay or Lesbian | 13,790 (1.6) | 175 (4.3) | 1,269.04 [1,087.98, 1,471.61] | 1,520.05 [1,278.74, 1,761.36] |
| | | Bisexual | 12,255 (1.4) | 245 (6.0) | 1,999.18 [1,756.67, 2,265.82] | 1,789.59 [1,554.76, 2,024.41] |
| | | Other | 4,350 (0.5) | 55 (1.4) | 1,264.37 [952.49, 1,645.75] | 1,281.58 [967.44, 1,665.73] |
| IMD/WIMD decile group | 1 (most deprived) | Total | 2,378,710 (100.0) | 19,735 (100.0) | 829.65 [818.12, 841.31] | 1,001.76 [986.63, 1,016.90] |
| | | Heterosexual | 2,269,820 (95.4) | 17,345 (87.9) | 764.16 [752.83, 775.62] | 914.77 [899.86, 929.68] |
| | | LGB+ | 108,885 (4.6) | 2,390 (12.1) | 2,194.98 [2,107.85, 2,284.78] | 2,180.56 [2,092.88, 2,268.25] |
| | | Gay or Lesbian | 48,905 (2.1) | 985 (5.0) | 2,014.11 [1,890.28, 2,143.92] | 2,271.32 [2,124.49, 2,418.15] |
| | | Bisexual | 46,150 (1.9) | 1,205 (6.1) | 2,611.05 [2,465.69, 2,762.74] | 2,393.09 [2,252.71, 2,533.47] |
| | | Other | 13,830 (0.6) | 200 (1.0) | 1,446.13 [1,252.65, 1,661.04] | 1,426.56 [1,224.43, 1,628.69] |
| | 2 | Total | 2,677,675 (100.0) | 18,015 (100.0) | 672.79 [663.00, 682.68] | 837.79 [824.32, 851.26] |
| | | Heterosexual | 2,550,445 (95.2) | 15,805 (87.7) | 619.70 [610.07, 629.43] | 762.76 [749.48, 776.05] |
| | | LGB+ | 127,225 (4.8) | 2,210 (12.3) | 1,737.08 [1,665.41, 1,811.05] | 1,750.98 [1,677.56, 1,824.41] |
| | | Gay or Lesbian | 58,190 (2.2) | 880 (4.9) | 1,512.29 [1,414.01, 1,615.60] | 1,771.90 [1,647.63, 1,896.17] |
| | | Bisexual | 53,140 (2.0) | 1,095 (6.1) | 2,060.59 [1,940.34, 2,186.35] | 1,837.38 [1,724.73, 1,950.04] |
| | | Other | 15,895 (0.6) | 235 (1.3) | 1,478.45 [1,295.45, 1,680.06] | 1,485.07 [1,292.55, 1,677.58] |
| | 3 | Total | 2,851,655 (100.0) | 16,605 (100.0) | 582.29 [573.47, 591.22] | 743.64 [731.02, 756.26] |
| | | Heterosexual | 2,711,710 (95.1) | 14,455 (87.1) | 533.06 [524.40, 541.82] | 670.09 [657.70, 682.49] |
| | | LGB+ | 139,940 (4.9) | 2,150 (12.9) | 1,536.37 [1,472.11, 1,602.72] | 1,580.41 [1,513.10, 1,647.72] |
| | | Gay or Lesbian | 65,585 (2.3) | 860 (5.2) | 1,311.28 [1,225.09, 1,401.92] | 1,629.38 [1,511.02, 1,747.75] |
| | | Bisexual | 57,480 (2.0) | 1,080 (6.5) | 1,878.91 [1,768.51, 1,994.40] | 1,694.27 [1,588.53, 1,800.02] |
| | | Other | 16,880 (0.6) | 215 (1.3) | 1,273.70 [1,109.12, 1,455.81] | 1,252.13 [1,081.65, 1,422.61] |
| | 4 | Total | 2,920,575 (100.0) | 15,235 (100.0) | 521.64 [513.39, 529.99] | 691.90 [679.40, 704.39] |
| | | Heterosexual | 2,784,255 (95.3) | 13,230 (86.8) | 475.17 [467.11, 483.34] | 617.49 [605.27, 629.71] |
| | | LGB+ | 136,320 (4.7) | 2,005 (13.2) | 1,470.80 [1,407.12, 1,536.62] | 1,490.48 [1,425.06, 1,555.91] |
| | | Gay or Lesbian | 64,705 (2.2) | 760 (5.0) | 1,174.56 [1,092.53, 1,261.12] | 1,494.77 [1,380.42, 1,609.13] |
| | | Bisexual | 56,185 (1.9) | 1,020 (6.7) | 1,815.43 [1,705.72, 1,930.35] | 1,621.78 [1,516.94, 1,726.62] |
| | | Other | 15,430 (0.5) | 225 (1.5) | 1,458.20 [1,273.87, 1,661.70] | 1,403.65 [1,217.26, 1,590.04] |
| | 5 | Total | 2,951,235 (100.0) | 13,690 (100.0) | 463.87 [456.14, 471.71] | 643.06 [630.55, 655.58] |
| | | Heterosexual | 2,830,040 (95.9) | 11,915 (87.0) | 421.02 [413.49, 428.65] | 569.19 [557.00, 581.37] |
| | | LGB+ | 121,195 (4.1) | 1,775 (13.0) | 1,464.58 [1,397.23, 1,534.34] | 1,472.63 [1,404.03, 1,541.22] |

| | | | | | | |
|---|---|---|---|---|---|---|
| | | Gay or Lesbian | 57,450 (1.9) | 645 (4.7) | 1,122.72 [1,037.73, 1,212.80] | 1,395.72 [1,281.50, 1,509.94] |
| | | Bisexual | 50,520 (1.7) | 930 (6.8) | 1,840.86 [1,724.43, 1,963.07] | 1,580.31 [1,473.89, 1,686.72] |
| | | Other | 13,225 (0.4) | 195 (1.4) | 1,474.48 [1,274.78, 1,696.59] | 1,400.09 [1,202.48, 1,597.70] |
| | 6 | Total | 3,012,900 (100.0) | 12,940 (100.0) | 429.49 [422.12, 436.95] | 613.41 [600.86, 625.97] |
| | | Heterosexual | 2,895,560 (96.1) | 11,365 (87.8) | 392.50 [385.31, 399.78] | 547.56 [535.26, 559.86] |
| | | LGB+ | 117,340 (3.9) | 1,580 (12.2) | 1,346.51 [1,280.93, 1,414.59] | 1,354.63 [1,287.78, 1,421.48] |
| | | Gay or Lesbian | 56,785 (1.9) | 545 (4.2) | 959.76 [880.86, 1,043.83] | 1,194.08 [1,087.75, 1,300.42] |
| | | Bisexual | 48,150 (1.6) | 880 (6.8) | 1,827.62 [1,708.85, 1,952.47] | 1,605.70 [1,493.79, 1,717.61] |
| | | Other | 12,405 (0.4) | 155 (1.2) | 1,249.50 [1,060.53, 1,462.42] | 1,179.10 [990.95, 1,367.25] |
| | 7 | Total | 2,975,620 (100.0) | 11,530 (100.0) | 387.48 [380.44, 394.62] | 581.88 [568.97, 594.79] |
| | | Heterosexual | 2,874,130 (96.6) | 10,165 (88.2) | 353.67 [346.83, 360.62] | 512.59 [500.08, 525.10] |
| | | LGB+ | 101,485 (3.4) | 1,365 (11.8) | 1,345.03 [1,274.61, 1,418.32] | 1,355.52 [1,283.51, 1,427.52] |
| | | Gay or Lesbian | 49,510 (1.7) | 435 (3.8) | 878.61 [797.97, 965.19] | 1,145.50 [1,031.63, 1,259.37] |
| | | Bisexual | 41,695 (1.4) | 780 (6.8) | 1,870.73 [1,741.73, 2,006.75] | 1,581.93 [1,464.57, 1,699.30] |
| | | Other | 10,285 (0.3) | 155 (1.3) | 1,507.05 [1,279.14, 1,763.86] | 1,387.04 [1,166.47, 1,607.61] |
| | 8 | Total | 2,988,555 (100.0) | 11,020 (100.0) | 368.74 [361.89, 375.69] | 559.27 [546.43, 572.11] |
| | | Heterosexual | 2,894,235 (96.8) | 9,745 (88.4) | 336.70 [330.05, 343.46] | 495.45 [482.95, 507.95] |
| | | LGB+ | 94,325 (3.2) | 1,275 (11.6) | 1,351.71 [1,278.52, 1,427.99] | 1,338.35 [1,264.50, 1,412.19] |
| | | Gay or Lesbian | 46,645 (1.6) | 450 (4.1) | 964.73 [877.65, 1,058.12] | 1,207.18 [1,090.03, 1,324.34] |
| | | Bisexual | 38,410 (1.3) | 695 (6.3) | 1,809.42 [1,677.38, 1,949.10] | 1,528.24 [1,406.72, 1,649.76] |
| | | Other | 9,265 (0.3) | 130 (1.2) | 1,403.13 [1,172.31, 1,666.10] | 1,281.50 [1,055.21, 1,507.80] |
| | 9 | Total | 2,970,040 (100.0) | 9,695 (100.0) | 326.43 [319.96, 332.99] | 519.65 [506.67, 532.64] |
| | | Heterosexual | 2,884,990 (97.1) | 8,615 (88.9) | 298.61 [292.34, 304.99] | 460.37 [447.73, 473.01] |
| | | LGB+ | 85,050 (2.9) | 1,085 (11.2) | 1,275.72 [1,200.93, 1,353.95] | 1,258.54 [1,183.22, 1,333.87] |
| | | Gay or Lesbian | 41,945 (1.4) | 390 (4.0) | 929.79 [839.79, 1,026.81] | 1,193.62 [1,069.49, 1,317.75] |
| | | Bisexual | 34,720 (1.2) | 560 (5.8) | 1,612.90 [1,482.07, 1,752.20] | 1,392.29 [1,268.92, 1,515.66] |
| | | Other | 8,385 (0.3) | 135 (1.4) | 1,610.02 [1,349.90, 1,905.65] | 1,389.98 [1,148.12, 1,631.84] |
| | 10 (least deprived) | Total | 2,932,505 (100.0) | 7,895 (100.0) | 269.22 [263.32, 275.23] | 455.37 [442.57, 468.18] |
| | | Heterosexual | 2,858,950 (97.5) | 6,980 (88.4) | 244.15 [238.45, 249.94] | 397.34 [384.99, 409.68] |
| | | LGB+ | 73,555 (2.5) | 915 (11.6) | 1,243.97 [1,164.66, 1,327.25] | 1,148.48 [1,071.52, 1,225.45] |
| | | Gay or Lesbian | 34,865 (1.2) | 275 (3.5) | 788.76 [698.28, 887.70] | 964.97 [846.49, 1,083.46] |
| | | Bisexual | 31,070 (1.1) | 525 (6.6) | 1,689.73 [1,548.27, 1,840.65] | 1,324.28 [1,197.93, 1,450.62] |
| | | Other | 7,620 (0.3) | 115 (1.5) | 1,509.19 [1,245.99, 1,811.55] | 1,222.43 [985.34, 1,459.51] |
| Highest level of qualification (≥25 years) | No qualifications | Total | 4,136,345 (100.0) | 23,120 (100.0) | 558.95 [551.77, 566.20] | 997.40 [979.58, 1,015.21] |
| | | Heterosexual | 4,069,475 (98.4) | 22,040 (95.3) | 541.59 [534.47, 548.79] | 958.74 [940.96, 976.52] |
| | | LGB+ | 66,870 (1.6) | 1,080 (4.7) | 1,615.07 [1,520.18, 1,714.35] | 1,921.75 [1,800.10, 2,043.40] |
| | | Gay or Lesbian | 29,215 (0.7) | 540 (2.3) | 1,848.37 [1,695.73, 2,011.05] | 2,716.39 [2,463.13, 2,969.64] |
| | | Bisexual | 22,570 (0.5) | 440 (1.9) | 1,949.49 [1,771.57, 2,140.44] | 2,148.09 [1,939.99, 2,356.20] |
| | | Other | 15,085 (0.4) | 95 (0.4) | 629.76 [509.52, 769.85] | 659.52 [530.63, 809.67] |
| | Level 1 (one to four GCSE passes (grade A* to C or grade 4 and above) and any other GCSEs at other | Total | 2,425,320 (100.0) | 12,700 (100.0) | 523.64 [514.57, 532.83] | 855.95 [837.17, 874.74] |
| | | Heterosexual | 2,373,955 (97.9) | 11,790 (92.8) | 496.64 [487.71, 505.69] | 802.46 [783.94, 820.98] |
| | | LGB+ | 51,365 (2.1) | 915 (7.2) | 1,781.37 [1,667.80, 1,900.63] | 2,036.89 [1,900.18, 2,173.61] |
| | | Gay or Lesbian | 28,730 (1.2) | 425 (3.3) | 1,479.29 [1,341.98, 1,626.84] | 1,885.05 [1,690.48, 2,079.61] |
| | | Bisexual | 17,765 (0.7) | 425 (3.3) | 2,392.34 [2,170.28, 2,630.97] | 2,427.06 [2,195.43, 2,658.68] |
| | | Other | 4,870 (0.2) | 65 (0.5) | 1,334.70 [1,030.10, 1,701.19] | 1,413.63 [1,077.06, 1,818.57] |

| | | | | | | |
|---|---|---|---|---|---|---|
| | grades, or equivalent qualifications) | | | | | |
| | Level 2 (five or more GCSE passes (grade A* to C or grade 4 and above) or equivalent qualifications) | Total | 3,085,135 (100.0) | 15,305 (100.0) | 496.09 [488.26, 504.01] | 695.38 [682.54, 708.21] |
| | | Heterosexual | 2,999,755 (97.2) | 13,930 (91.0) | 464.37 [456.69, 472.15] | 642.23 [629.63, 654.83] |
| | | LGB+ | 85,380 (2.8) | 1,375 (9.0) | 1,610.45 [1,526.44, 1,697.88] | 1,663.29 [1,574.95, 1,751.63] |
| | | Gay or Lesbian | 48,750 (1.6) | 590 (3.9) | 1,210.26 [1,114.56, 1,311.97] | 1,388.12 [1,272.68, 1,503.56] |
| | | Bisexual | 29,450 (1.0) | 670 (4.4) | 2,275.04 [2,106.01, 2,454.03] | 2,079.81 [1,921.00, 2,238.62] |
| | | Other | 7,180 (0.2) | 110 (0.7) | 1,532.03 [1,259.15, 1,846.51] | 1,458.17 [1,185.12, 1,731.22] |
| | Apprenticeship | Total | 1,309,970 (100.0) | 5,175 (100.0) | 395.05 [384.36, 405.96] | 615.52 [594.62, 636.42] |
| | | Heterosexual | 1,288,495 (98.4) | 4,915 (95.0) | 381.45 [370.86, 392.27] | 586.57 [565.85, 607.29] |
| | | LGB+ | 21,475 (1.6) | 260 (5.0) | 1,210.71 [1,068.01, 1,367.17] | 1,302.30 [1,143.20, 1,461.40] |
| | | Gay or Lesbian | 10,410 (0.8) | 110 (2.1) | 1,056.68 [868.46, 1,273.58] | 1,274.95 [1,033.47, 1,516.43] |
| | | Bisexual | 7,835 (0.6) | 130 (2.5) | 1,659.22 [1,386.28, 1,970.19] | 1,607.00 [1,328.03, 1,885.96] |
| | | Other | 3,230 (0.2) | 20 (0.4) | 619.20 [378.22, 956.30] | 683.92 [418.43, 1,055.32] |
| | Level 3 (two or more A Levels or equivalent qualifications) | Total | 4,046,195 (100.0) | 16,755 (100.0) | 414.09 [407.85, 420.41] | 494.63 [486.59, 502.67] |
| | | Heterosexual | 3,908,325 (96.6) | 14,800 (88.3) | 378.68 [372.60, 384.83] | 447.21 [439.39, 455.02] |
| | | LGB+ | 137,875 (3.4) | 1,960 (11.7) | 1,421.58 [1,359.33, 1,485.94] | 1,341.57 [1,281.70, 1,401.45] |
| | | Gay or Lesbian | 77,020 (1.9) | 900 (5.4) | 1,168.53 [1,093.42, 1,247.43] | 1,188.55 [1,110.54, 1,266.56] |
| | | Bisexual | 48,515 (1.2) | 850 (5.1) | 1,752.04 [1,636.22, 1,873.89] | 1,574.89 [1,462.11, 1,687.67] |
| | | Other | 12,335 (0.3) | 210 (1.3) | 1,702.47 [1,479.98, 1,948.97] | 1,512.33 [1,297.35, 1,727.30] |
| | Level 4 and above (Higher National Certificate, Higher National Diploma, Bachelor's degree, or post-graduate qualifications) | Total | 10,118,375 (100.0) | 26,300 (100.0) | 259.92 [256.79, 263.08] | 289.78 [285.97, 293.58] |
| | | Heterosexual | 9,673,645 (95.6) | 23,350 (88.8) | 241.38 [238.29, 244.49] | 265.58 [261.84, 269.32] |
| | | LGB+ | 444,730 (4.4) | 2,950 (11.2) | 663.32 [639.60, 687.70] | 650.04 [626.53, 673.55] |
| | | Gay or Lesbian | 245,755 (2.4) | 1,345 (5.1) | 547.29 [518.43, 577.34] | 571.74 [540.82, 602.65] |
| | | Bisexual | 155,690 (1.5) | 1,255 (4.8) | 806.09 [762.10, 851.95] | 756.43 [712.02, 800.83] |
| | | Other | 43,285 (0.4) | 350 (1.3) | 808.59 [726.09, 897.90] | 756.01 [673.85, 838.16] |
| | Other qualifications | Total | 688,635 (100.0) | 2,665 (100.0) | 387.00 [372.44, 401.98] | 573.90 [544.68, 603.12] |
| | | Heterosexual | 675,740 (98.1) | 2,530 (94.9) | 374.40 [359.96, 389.28] | 549.35 [520.16, 578.53] |
| | | LGB+ | 12,895 (1.9) | 135 (5.1) | 1,046.92 [877.77, 1,239.15] | 1,158.69 [959.13, 1,358.25] |
| | | Gay or Lesbian | 6,295 (0.9) | 70 (2.6) | 1,111.99 [866.85, 1,404.94] | 1,383.67 [1,058.00, 1,772.84] |
| | | Bisexual | 4,410 (0.6) | 55 (2.1) | 1,247.17 [939.54, 1,623.36] | 1,215.58 [907.51, 1,592.33] |
| | | Other | 2,190 (0.3) | 15 (0.6) | 684.93 [383.35, 1,129.69] | 621.92 [333.38, 1,047.44] |
| NS-SEC | Class 1 (Higher managerial, administrative and professional occupations) | Total | 4,328,200 (100.0) | 8,250 (100.0) | 190.61 [186.52, 194.77] | 300.89 [278.00, 323.78] |
| | | Heterosexual | 4,161,235 (96.1) | 7,420 (89.9) | 178.31 [174.28, 182.42] | 258.83 [238.36, 279.30] |
| | | LGB+ | 166,965 (3.9) | 830 (10.1) | 497.11 [463.86, 532.11] | 926.49 [716.66, 1,136.33] |
| | | Gay or Lesbian | 101,055 (2.3) | 405 (4.9) | 400.77 [362.69, 441.77] | 775.51 [445.82, 1,105.20] |
| | | Bisexual | 52,965 (1.2) | 335 (4.1) | 632.49 [566.57, 703.98] | 1,076.45 [774.82, 1,378.07] |
| | | Other | 12,945 (0.3) | 85 (1.0) | 656.62 [524.49, 811.93] | 899.79 [394.44, 1,493.74] |
| | Class 2 (Lower managerial, administrative and professional occupations) | Total | 6,323,480 (100.0) | 18,100 (100.0) | 286.23 [282.08, 290.44] | 461.25 [443.92, 478.58] |
| | | Heterosexual | 6,058,685 (95.8) | 16,005 (88.4) | 264.17 [260.09, 268.29] | 400.48 [384.25, 416.70] |
| | | LGB+ | 264,795 (4.2) | 2,095 (11.6) | 791.18 [757.66, 825.80] | 1,176.10 [1,069.33, 1,282.87] |
| | | Gay or Lesbian | 151,070 (2.4) | 945 (5.2) | 625.54 [586.29, 666.73] | 1,044.69 [869.48, 1,219.91] |
| | | Bisexual | 91,305 (1.4) | 930 (5.1) | 1,018.56 [954.14, 1,086.19] | 1,337.57 [1,184.81, 1,490.33] |
| | | Other | 22,420 (0.4) | 220 (1.2) | 981.27 [855.88, 1,119.86] | 1,139.42 [854.30, 1,424.55] |

| | | | | | | |
|---|---|---|---|---|---|---|
| | Class 3 (Intermediate occupations) | Total | 3,512,855 (100.0) | 13,880 (100.0) | 395.12 [388.57, 401.75] | 617.52 [601.94, 633.11] |
| | | Heterosexual | 3,400,585 (96.8) | 12,310 (88.7) | 362.00 [355.63, 368.45] | 551.14 [536.00, 566.28] |
| | | LGB+ | 112,275 (3.2) | 1,570 (11.3) | 1,398.35 [1,330.03, 1,469.27] | 1,570.82 [1,472.68, 1,668.97] |
| | | Gay or Lesbian | 56,090 (1.6) | 650 (4.7) | 1,158.85 [1,071.46, 1,251.47] | 1,501.08 [1,337.78, 1,664.38] |
| | | Bisexual | 45,315 (1.3) | 785 (5.7) | 1,732.32 [1,613.24, 1,857.86] | 1,757.17 [1,610.39, 1,903.96] |
| | | Other | 10,865 (0.3) | 130 (0.9) | 1,196.50 [999.67, 1,420.75] | 1,197.82 [942.30, 1,453.33] |
| | Class 4 (Small employers and own account workers) | Total | 2,973,310 (100.0) | 10,695 (100.0) | 359.70 [352.91, 366.58] | 637.84 [612.77, 662.91] |
| | | Heterosexual | 2,903,450 (97.7) | 10,115 (94.6) | 348.38 [341.62, 355.24] | 608.40 [583.52, 633.28] |
| | | LGB+ | 69,860 (2.3) | 580 (5.4) | 830.23 [764.03, 900.63] | 1,281.99 [1,100.83, 1,463.14] |
| | | Gay or Lesbian | 35,235 (1.2) | 255 (2.4) | 723.71 [637.60, 818.21] | 1,260.34 [885.53, 1,635.15] |
| | | Bisexual | 24,240 (0.8) | 255 (2.4) | 1,051.98 [926.81, 1,189.34] | 1,496.96 [1,219.28, 1,774.64] |
| | | Other | 10,385 (0.3) | 70 (0.7) | 674.05 [525.45, 851.62] | 795.26 [553.21, 1,084.50] |
| | Class 5 (Lower supervisory and technical occupations) | Total | 1,479,235 (100.0) | 6,835 (100.0) | 462.06 [451.17, 473.15] | 666.61 [644.75, 688.46] |
| | | Heterosexual | 1,435,915 (97.1) | 6,235 (91.2) | 434.22 [423.51, 445.13] | 613.35 [592.16, 634.53] |
| | | LGB+ | 43,320 (2.9) | 600 (8.8) | 1,385.04 [1,276.42, 1,500.44] | 1,756.73 [1,562.28, 1,951.18] |
| | | Gay or Lesbian | 22,760 (1.5) | 285 (4.2) | 1,252.20 [1,111.02, 1,406.34] | 1,670.86 [1,382.83, 1,958.89] |
| | | Bisexual | 16,000 (1.1) | 270 (4.0) | 1,687.50 [1,492.20, 1,901.26] | 1,920.15 [1,633.70, 2,206.60] |
| | | Other | 4,555 (0.3) | 45 (0.7) | 987.93 [720.60, 1,321.92] | 1,639.04 [937.74, 2,515.26] |
| | Class 6 (Semi-routine occupations) | Total | 3,198,160 (100.0) | 21,070 (100.0) | 658.82 [649.95, 667.77] | 996.67 [980.04, 1,013.29] |
| | | Heterosexual | 3,084,725 (96.5) | 18,495 (87.8) | 599.57 [590.96, 608.27] | 892.59 [876.33, 908.85] |
| | | LGB+ | 113,435 (3.5) | 2,575 (12.2) | 2,270.02 [2,183.18, 2,359.43] | 2,306.77 [2,211.76, 2,401.79] |
| | | Gay or Lesbian | 49,760 (1.6) | 970 (4.6) | 1,949.36 [1,828.60, 2,076.00] | 2,250.58 [2,095.09, 2,406.07] |
| | | Bisexual | 50,810 (1.6) | 1,350 (6.4) | 2,656.96 [2,517.10, 2,802.56] | 2,474.91 [2,332.57, 2,617.25] |
| | | Other | 12,865 (0.4) | 255 (1.2) | 1,982.12 [1,746.28, 2,240.93] | 2,005.09 [1,724.97, 2,285.21] |
| | Class 7 (Routine occupations) | Total | 3,152,270 (100.0) | 20,455 (100.0) | 648.90 [640.04, 657.85] | 972.09 [955.71, 988.48] |
| | | Heterosexual | 3,057,560 (97.0) | 18,475 (90.3) | 604.24 [595.56, 613.02] | 891.41 [875.28, 907.54] |
| | | LGB+ | 94,715 (3.0) | 1,980 (9.7) | 2,090.48 [1,999.41, 2,184.64] | 2,158.79 [2,059.82, 2,257.76] |
| | | Gay or Lesbian | 38,680 (1.2) | 735 (3.6) | 1,900.21 [1,765.30, 2,042.69] | 2,201.99 [2,031.01, 2,372.98] |
| | | Bisexual | 42,530 (1.3) | 1,050 (5.1) | 2,468.85 [2,321.75, 2,622.81] | 2,316.24 [2,169.21, 2,463.27] |
| | | Other | 13,505 (0.4) | 195 (1.0) | 1,443.91 [1,248.35, 1,661.41] | 1,577.03 [1,334.08, 1,819.99] |
| | Class 8 (Never worked and long-term unemployed) | Total | 2,106,980 (100.0) | 21,285 (100.0) | 1,010.21 [996.69, 1,023.88] | 1,641.43 [1,616.34, 1,666.51] |
| | | Heterosexual | 2,036,855 (96.7) | 18,940 (89.0) | 929.86 [916.67, 943.20] | 1,499.65 [1,474.91, 1,524.40] |
| | | LGB+ | 70,125 (3.3) | 2,345 (11.0) | 3,344.03 [3,210.04, 3,482.18] | 3,460.37 [3,319.71, 3,601.04] |
| | | Gay or Lesbian | 25,165 (1.2) | 890 (4.2) | 3,536.66 [3,308.09, 3,776.85] | 4,208.08 [3,926.39, 4,489.76] |
| | | Bisexual | 31,620 (1.5) | 1,225 (5.8) | 3,874.13 [3,660.19, 4,097.31] | 3,660.35 [3,450.22, 3,870.47] |
| | | Other | 13,340 (0.6) | 230 (1.1) | 1,724.14 [1,508.50, 1,961.95] | 1,871.60 [1,627.91, 2,115.29] |
| | Full-time students | Total | 1,584,975 (100.0) | 15,800 (100.0) | 996.86 [981.38, 1,012.53] | 616.35 [595.90, 636.80] |
| | | Heterosexual | 1,415,135 (89.3) | 11,630 (73.6) | 821.83 [806.96, 836.90] | 525.27 [505.12, 545.42] |
| | | LGB+ | 169,840 (10.7) | 4,170 (26.4) | 2,455.25 [2,381.29, 2,530.93] | 1,447.62 [1,305.41, 1,589.84] |
| | | Gay or Lesbian | 44,770 (2.8) | 1,085 (6.9) | 2,423.50 [2,281.42, 2,572.11] | 1,530.05 [1,280.15, 1,779.95] |
| | | Bisexual | 102,735 (6.5) | 2,565 (16.2) | 2,496.71 [2,401.02, 2,595.25] | 1,313.85 [1,179.41, 1,448.29] |
| | | Other | 22,335 (1.4) | 525 (3.3) | 2,350.57 [2,153.78, 2,560.51] | 1,454.33 [1,123.19, 1,785.47] |
| Household NS-SEC | Class 1 (Higher managerial, administrative and | Total | 7,224,420 (100.0) | 17,350 (100.0) | 240.16 [236.60, 243.76] | 367.23 [360.21, 374.25] |
| | | Heterosexual | 6,936,510 (96.0) | 15,130 (87.2) | 218.12 [214.66, 221.62] | 319.90 [313.14, 326.66] |
| | | LGB+ | 287,910 (4.0) | 2,215 (12.8) | 769.34 [737.63, 802.06] | 889.58 [851.33, 927.83] |

| | | | | | | |
|---|---|---|---|---|---|---|
| | professional occupations) | Gay or Lesbian | 157,010 (2.2) | 850 (4.9) | 541.37 [505.58, 579.02] | 814.39 [749.23, 879.55] |
| | | Bisexual | 104,825 (1.5) | 1,105 (6.4) | 1,054.14 [992.89, 1,118.17] | 981.30 [921.69, 1,040.90] |
| | | Other | 26,075 (0.4) | 265 (1.5) | 1,016.30 [897.61, 1,146.32] | 962.84 [844.33, 1,081.36] |
| | Class 2 (Lower managerial, administrative and professional occupations) | Total | 7,994,445 (100.0) | 28,405 (100.0) | 355.31 [351.19, 359.47] | 516.35 [509.06, 523.64] |
| | | Heterosexual | 7,669,120 (95.9) | 24,830 (87.4) | 323.77 [319.75, 327.82] | 458.63 [451.54, 465.73] |
| | | LGB+ | 325,325 (4.1) | 3,570 (12.6) | 1,097.36 [1,061.66, 1,133.96] | 1,209.16 [1,168.67, 1,249.66] |
| | | Gay or Lesbian | 173,375 (2.2) | 1,455 (5.1) | 839.22 [796.65, 883.48] | 1,130.55 [1,064.15, 1,196.95] |
| | | Bisexual | 121,205 (1.5) | 1,730 (6.1) | 1,427.33 [1,360.86, 1,496.22] | 1,325.11 [1,261.05, 1,389.18] |
| | | Other | 30,745 (0.4) | 390 (1.4) | 1,268.50 [1,145.71, 1,400.86] | 1,242.66 [1,116.67, 1,368.65] |
| | Class 3 (Intermediate occupations) | Total | 3,551,725 (100.0) | 16,690 (100.0) | 469.91 [462.81, 477.10] | 688.55 [676.42, 700.67] |
| | | Heterosexual | 3,443,335 (96.9) | 14,825 (88.8) | 430.54 [423.64, 437.53] | 620.58 [608.74, 632.42] |
| | | LGB+ | 108,390 (3.1) | 1,865 (11.2) | 1,720.64 [1,643.43, 1,800.54] | 1,714.27 [1,636.22, 1,792.32] |
| | | Gay or Lesbian | 51,290 (1.4) | 780 (4.7) | 1,520.76 [1,415.90, 1,631.34] | 1,732.90 [1,607.37, 1,858.42] |
| | | Bisexual | 45,360 (1.3) | 905 (5.4) | 1,995.15 [1,867.26, 2,129.49] | 1,779.31 [1,658.96, 1,899.65] |
| | | Other | 11,740 (0.3) | 180 (1.1) | 1,533.22 [1,317.41, 1,774.30] | 1,468.96 [1,254.03, 1,683.89] |
| | Class 4 (Small employers and own account workers) | Total | 2,794,170 (100.0) | 12,955 (100.0) | 463.64 [455.69, 471.70] | 664.74 [650.52, 678.96] |
| | | Heterosexual | 2,722,625 (97.4) | 12,075 (93.2) | 443.51 [435.63, 451.49] | 628.58 [614.47, 642.70] |
| | | LGB+ | 71,545 (2.6) | 880 (6.8) | 1,230.00 [1,150.06, 1,314.02] | 1,402.04 [1,305.95, 1,498.14] |
| | | Gay or Lesbian | 30,595 (1.1) | 340 (2.6) | 1,111.29 [996.29, 1,235.92] | 1,516.11 [1,337.90, 1,694.32] |
| | | Bisexual | 28,725 (1.0) | 430 (3.3) | 1,496.95 [1,358.79, 1,645.36] | 1,498.58 [1,354.21, 1,642.95] |
| | | Other | 12,225 (0.4) | 110 (0.8) | 899.80 [739.52, 1,084.50] | 1,026.79 [831.59, 1,221.99] |
| | Class 5 (Lower supervisory and technical occupations) | Total | 1,326,605 (100.0) | 7,620 (100.0) | 574.40 [561.57, 587.44] | 790.36 [769.71, 811.02] |
| | | Heterosexual | 1,284,420 (96.8) | 6,855 (90.0) | 533.70 [521.14, 546.49] | 723.06 [702.80, 743.33] |
| | | LGB+ | 42,185 (3.2) | 770 (10.1) | 1,825.29 [1,698.63, 1,958.90] | 1,870.31 [1,737.64, 2,002.98] |
| | | Gay or Lesbian | 18,910 (1.4) | 315 (4.1) | 1,665.79 [1,486.89, 1,860.28] | 1,938.04 [1,713.02, 2,163.06] |
| | | Bisexual | 18,045 (1.4) | 400 (5.2) | 2,216.68 [2,004.75, 2,444.92] | 2,069.77 [1,863.38, 2,276.17] |
| | | Other | 5,230 (0.4) | 55 (0.7) | 1,051.63 [792.23, 1,368.83] | 1,043.29 [777.06, 1,368.85] |
| | Class 6 (Semi-routine occupations) | Total | 2,258,230 (100.0) | 18,260 (100.0) | 808.60 [796.91, 820.41] | 1,159.98 [1,141.10, 1,178.86] |
| | | Heterosexual | 2,175,160 (96.3) | 16,025 (87.8) | 736.73 [725.36, 748.22] | 1,044.79 [1,026.29, 1,063.29] |
| | | LGB+ | 83,070 (3.7) | 2,235 (12.2) | 2,690.50 [2,580.10, 2,804.41] | 2,653.21 [2,542.96, 2,763.46] |
| | | Gay or Lesbian | 34,365 (1.5) | 855 (4.7) | 2,488.00 [2,324.00, 2,660.51] | 2,820.33 [2,629.00, 3,011.66] |
| | | Bisexual | 38,565 (1.7) | 1,160 (6.4) | 3,007.91 [2,837.28, 3,186.11] | 2,725.72 [2,563.86, 2,887.59] |
| | | Other | 10,140 (0.4) | 220 (1.2) | 2,169.63 [1,892.38, 2,476.06] | 2,137.56 [1,855.02, 2,420.09] |
| | Class 7 (Routine occupations) | Total | 1,663,240 (100.0) | 14,065 (100.0) | 845.64 [831.72, 859.73] | 1,179.92 [1,157.48, 1,202.36] |
| | | Heterosexual | 1,605,255 (96.5) | 12,565 (89.3) | 782.74 [769.11, 796.55] | 1,078.98 [1,056.87, 1,101.09] |
| | | LGB+ | 57,990 (3.5) | 1,500 (10.7) | 2,586.65 [2,457.39, 2,720.95] | 2,627.49 [2,494.10, 2,760.89] |
| | | Gay or Lesbian | 21,570 (1.3) | 520 (3.7) | 2,410.76 [2,207.98, 2,627.15] | 2,767.79 [2,523.36, 3,012.22] |
| | | Bisexual | 27,295 (1.6) | 830 (5.9) | 3,040.85 [2,837.47, 3,254.96] | 2,835.22 [2,638.70, 3,031.75] |
| | | Other | 9,120 (0.5) | 150 (1.1) | 1,644.74 [1,392.06, 1,930.01] | 1,706.68 [1,432.58, 1,980.78] |
| | Class 8 (Never worked and long-term unemployed) | Total | 1,178,230 (100.0) | 14,385 (100.0) | 1,220.90 [1,201.03, 1,241.02] | 2,205.68 [2,163.93, 2,247.43] |
| | | Heterosexual | 1,138,610 (96.6) | 12,810 (89.1) | 1,125.06 [1,105.66, 1,144.71] | 2,020.02 [1,978.77, 2,061.27] |
| | | LGB+ | 39,625 (3.4) | 1,575 (10.9) | 3,974.76 [3,780.87, 4,176.03] | 4,441.73 [4,220.70, 4,662.76] |
| | | Gay or Lesbian | 15,185 (1.3) | 615 (4.3) | 4,050.05 [3,736.24, 4,383.18] | 5,536.63 [5,075.51, 5,997.76] |
| | | Bisexual | 17,290 (1.5) | 815 (5.7) | 4,713.71 [4,395.60, 5,048.75] | 4,630.27 [4,310.72, 4,949.83] |
| | | Other | 7,150 (0.6) | 145 (1.0) | 2,027.97 [1,711.33, 2,386.22] | 2,266.94 [1,886.00, 2,647.88] |

| | | | | | | |
|---|---|---|---|---|---|---|
| | Full-time students | Total | 348,600 (100.0) | 3,175 (100.0) | 910.79 [879.38, 943.03] | 796.70 [754.54, 838.86] |
| | | Heterosexual | 295,750 (84.8) | 2,065 (65.0) | 698.22 [668.43, 729.00] | 667.04 [625.65, 708.44] |
| | | LGB+ | 52,845 (15.2) | 1,110 (35.0) | 2,100.48 [1,978.72, 2,227.78] | 1,754.03 [1,489.68, 2,018.38] |
| | | Gay or Lesbian | 13,075 (3.8) | 275 (8.7) | 2,103.25 [1,861.99, 2,367.10] | 1,920.29 [1,447.94, 2,392.63] |
| | | Bisexual | 33,185 (9.5) | 700 (22.0) | 2,109.39 [1,956.00, 2,271.61] | 1,559.22 [1,312.16, 1,806.28] |
| | | Other | 6,585 (1.9) | 135 (4.3) | 2,050.11 [1,718.89, 2,426.56] | 1,768.11 [1,007.43, 2,528.80] |
| | Living in a communal establishment | Total | 319,805 (100.0) | 3,465 (100.0) | 1,083.47 [1,047.69, 1,120.16] | 1,560.90 [1,478.29, 1,643.51] |
| | | Heterosexual | 283,355 (88.6) | 2,440 (70.4) | 861.11 [827.28, 895.97] | 1,390.50 [1,308.06, 1,472.94] |
| | | LGB+ | 36,445 (11.4) | 1,025 (29.6) | 2,812.46 [2,642.89, 2,990.04] | 3,135.22 [2,662.89, 3,607.55] |
| | | Gay or Lesbian | 9,215 (2.9) | 225 (6.5) | 2,441.67 [2,133.03, 2,782.42] | 2,481.32 [1,928.66, 3,033.98] |
| | | Bisexual | 23,020 (7.2) | 695 (20.1) | 3,019.11 [2,798.80, 3,252.16] | 4,730.07 [3,639.57, 5,820.57] |
| | | Other | 4,210 (1.3) | 105 (3.0) | 2,494.06 [2,039.90, 3,019.21] | 1,361.57 [868.38, 1,854.75] |
| Lone person household indicator | Lives alone | Total | 4,600,305 (100.0) | 35,640 (100.0) | 774.73 [766.71, 782.82] | 1,865.65 [1,827.04, 1,904.26] |
| | | Heterosexual | 4,363,790 (94.9) | 30,845 (86.5) | 706.84 [698.97, 714.77] | 1,622.30 [1,583.99, 1,660.60] |
| | | LGB+ | 236,515 (5.1) | 4,795 (13.5) | 2,027.36 [1,970.37, 2,085.57] | 3,343.08 [3,208.43, 3,477.73] |
| | | Gay or Lesbian | 134,830 (2.9) | 2,130 (6.0) | 1,579.77 [1,513.38, 1,648.31] | 3,411.68 [3,155.53, 3,667.83] |
| | | Bisexual | 78,515 (1.7) | 2,190 (6.1) | 2,789.28 [2,673.67, 2,908.60] | 3,609.62 [3,425.42, 3,793.82] |
| | | Other | 23,170 (0.5) | 475 (1.3) | 2,050.06 [1,869.83, 2,242.99] | 2,796.16 [2,446.18, 3,146.14] |
| | Does not live alone | Total | 23,739,360 (100.0) | 97,265 (100.0) | 409.72 [407.15, 412.30] | 576.67 [572.54, 580.79] |
| | | Heterosexual | 22,906,995 (96.5) | 86,340 (88.8) | 376.92 [374.41, 379.44] | 523.33 [519.28, 527.39] |
| | | LGB+ | 832,365 (3.5) | 10,925 (11.2) | 1,312.53 [1,288.03, 1,337.37] | 1,257.88 [1,234.22, 1,281.53] |
| | | Gay or Lesbian | 380,540 (1.6) | 3,870 (4.0) | 1,016.98 [985.18, 1,049.53] | 1,203.39 [1,164.15, 1,242.63] |
| | | Bisexual | 355,985 (1.5) | 5,885 (6.1) | 1,653.16 [1,611.19, 1,695.95] | 1,389.95 [1,352.03, 1,427.86] |
| | | Other | 95,840 (0.4) | 1,170 (1.2) | 1,220.78 [1,151.83, 1,292.79] | 1,100.47 [1,036.56, 1,164.37] |
| | Living in a communal establishment | Total | 319,805 (100.0) | 3,465 (100.0) | 1,083.47 [1,047.69, 1,120.16] | 1,560.90 [1,478.29, 1,643.51] |
| | | Heterosexual | 283,355 (88.6) | 2,440 (70.4) | 861.11 [827.28, 895.97] | 1,390.50 [1,308.06, 1,472.94] |
| | | LGB+ | 36,445 (11.4) | 1,025 (29.6) | 2,812.46 [2,642.89, 2,990.04] | 3,135.22 [2,662.89, 3,607.55] |
| | | Gay or Lesbian | 9,215 (2.9) | 225 (6.5) | 2,441.67 [2,133.03, 2,782.42] | 2,481.32 [1,928.66, 3,033.98] |
| | | Bisexual | 23,020 (7.2) | 695 (20.1) | 3,019.11 [2,798.80, 3,252.16] | 4,730.07 [3,639.57, 5,820.57] |
| | | Other | 4,210 (1.3) | 105 (3.0) | 2,494.06 [2,039.90, 3,019.21] | 1,361.57 [868.38, 1,854.75] |
| Household size | One person in household | Total | 4,600,305 (100.0) | 35,640 (100.0) | 774.73 [766.71, 782.82] | 1,865.65 [1,827.04, 1,904.26] |
| | | Heterosexual | 4,363,790 (94.9) | 30,845 (86.5) | 706.84 [698.97, 714.77] | 1,622.30 [1,583.99, 1,660.60] |
| | | LGB+ | 236,515 (5.1) | 4,795 (13.5) | 2,027.36 [1,970.37, 2,085.57] | 3,343.08 [3,208.43, 3,477.73] |
| | | Gay or Lesbian | 134,830 (2.9) | 2,130 (6.0) | 1,579.77 [1,513.38, 1,648.31] | 3,411.68 [3,155.53, 3,667.83] |
| | | Bisexual | 78,515 (1.7) | 2,190 (6.1) | 2,789.28 [2,673.67, 2,908.60] | 3,609.62 [3,425.42, 3,793.82] |
| | | Other | 23,170 (0.5) | 475 (1.3) | 2,050.06 [1,869.83, 2,242.99] | 2,796.16 [2,446.18, 3,146.14] |
| | Two people in household | Total | 10,142,025 (100.0) | 36,905 (100.0) | 363.88 [360.18, 367.61] | 718.94 [707.85, 730.04] |
| | | Heterosexual | 9,726,660 (95.9) | 32,495 (88.1) | 334.08 [330.46, 337.73] | 644.52 [633.46, 655.58] |
| | | LGB+ | 415,365 (4.1) | 4,410 (11.9) | 1,061.72 [1,030.61, 1,093.52] | 1,339.10 [1,293.04, 1,385.17] |
| | | Gay or Lesbian | 239,535 (2.4) | 1,845 (5.0) | 770.24 [735.49, 806.21] | 1,279.04 [1,195.61, 1,362.47] |
| | | Bisexual | 141,265 (1.4) | 2,145 (5.8) | 1,518.42 [1,454.84, 1,584.07] | 1,511.79 [1,441.38, 1,582.19] |
| | | Other | 34,560 (0.3) | 425 (1.2) | 1,229.75 [1,115.60, 1,352.41] | 1,354.91 [1,211.40, 1,498.42] |
| | Three to five people in household | Total | 12,340,900 (100.0) | 53,740 (100.0) | 435.46 [431.79, 439.16] | 551.29 [546.17, 556.41] |
| | | Heterosexual | 11,964,450 (96.9) | 47,910 (89.2) | 400.44 [396.86, 404.04] | 504.88 [499.83, 509.92] |
| | | LGB+ | 376,450 (3.1) | 5,830 (10.8) | 1,548.68 [1,509.18, 1,588.95] | 1,259.16 [1,223.80, 1,294.53] |

| | | | | | | |
|---|---|---|---|---|---|---|
| | | Gay or Lesbian | 131,220 (1.1) | 1,845 (3.4) | 1,406.04 [1,342.60, 1,471.69] | 1,233.34 [1,174.55, 1,292.12] |
| | | Bisexual | 192,115 (1.6) | 3,325 (6.2) | 1,730.73 [1,672.40, 1,790.58] | 1,362.68 [1,307.54, 1,417.82] |
| | | Other | 53,115 (0.4) | 660 (1.2) | 1,242.59 [1,149.58, 1,341.11] | 1,003.22 [924.46, 1,081.98] |
| | Six or more people in household | Total | 1,256,435 (100.0) | 6,620 (100.0) | 526.89 [514.27, 539.74] | 534.47 [520.96, 547.99] |
| | | Heterosexual | 1,215,885 (96.8) | 5,935 (89.7) | 488.12 [475.78, 500.70] | 498.33 [484.98, 511.68] |
| | | LGB+ | 40,550 (3.2) | 685 (10.3) | 1,689.27 [1,565.12, 1,820.65] | 1,240.30 [1,132.69, 1,347.91] |
| | | Gay or Lesbian | 9,785 (0.8) | 180 (2.7) | 1,839.55 [1,580.62, 2,128.80] | 1,335.64 [1,115.15, 1,556.14] |
| | | Bisexual | 22,600 (1.8) | 415 (6.3) | 1,836.28 [1,663.84, 2,021.74] | 1,351.16 [1,195.20, 1,507.11] |
| | | Other | 8,160 (0.6) | 85 (1.3) | 1,041.67 [832.05, 1,288.04] | 885.45 [690.18, 1,114.95] |
| | Living in a communal establishment | Total | 319,805 (100.0) | 3,465 (100.0) | 1,083.47 [1,047.69, 1,120.16] | 1,560.90 [1,478.29, 1,643.51] |
| | | Heterosexual | 283,355 (88.6) | 2,440 (70.4) | 861.11 [827.28, 895.97] | 1,390.50 [1,308.06, 1,472.94] |
| | | LGB+ | 36,445 (11.4) | 1,025 (29.6) | 2,812.46 [2,642.89, 2,990.04] | 3,135.22 [2,662.89, 3,607.55] |
| | | Gay or Lesbian | 9,215 (2.9) | 225 (6.5) | 2,441.67 [2,133.03, 2,782.42] | 2,481.32 [1,928.66, 3,033.98] |
| | | Bisexual | 23,020 (7.2) | 695 (20.1) | 3,019.11 [2,798.80, 3,252.16] | 4,730.07 [3,639.57, 5,820.57] |
| | | Other | 4,210 (1.3) | 105 (3.0) | 2,494.06 [2,039.90, 3,019.21] | 1,361.57 [868.38, 1,854.75] |
| Household tenure | Owned or shared ownership | Total | 18,590,085 (100.0) | 51,250 (100.0) | 275.68 [273.30, 278.08] | 430.78 [425.76, 435.81] |
| | | Heterosexual | 18,092,870 (97.3) | 47,020 (91.7) | 259.88 [257.54, 262.24] | 393.60 [388.67, 398.53] |
| | | LGB+ | 497,215 (2.7) | 4,225 (8.2) | 849.73 [824.30, 875.75] | 980.96 [950.18, 1,011.73] |
| | | Gay or Lesbian | 284,235 (1.5) | 1,735 (3.4) | 610.41 [582.02, 639.82] | 917.92 [868.29, 967.56] |
| | | Bisexual | 167,880 (0.9) | 2,000 (3.9) | 1,191.33 [1,139.68, 1,244.71] | 1,063.93 [1,015.82, 1,112.04] |
| | | Other | 45,100 (0.2) | 490 (1.0) | 1,086.47 [992.39, 1,187.07] | 1,041.34 [947.25, 1,135.44] |
| | Social rented | Total | 4,048,710 (100.0) | 44,785 (100.0) | 1,106.15 [1,095.93, 1,116.45] | 1,381.68 [1,367.51, 1,395.86] |
| | | Heterosexual | 3,888,600 (96.0) | 39,625 (88.5) | 1,019.00 [1,009.00, 1,029.09] | 1,257.39 [1,243.51, 1,271.26] |
| | | LGB+ | 160,110 (4.0) | 5,160 (11.5) | 3,222.78 [3,135.44, 3,311.94] | 3,283.08 [3,193.12, 3,373.04] |
| | | Gay or Lesbian | 69,005 (1.7) | 2,025 (4.5) | 2,934.57 [2,808.13, 3,065.23] | 3,351.81 [3,201.75, 3,501.86] |
| | | Bisexual | 71,845 (1.8) | 2,690 (6.0) | 3,744.17 [3,604.00, 3,888.39] | 3,512.27 [3,377.59, 3,646.94] |
| | | Other | 19,260 (0.5) | 445 (1.0) | 2,310.49 [2,100.78, 2,535.47] | 2,398.58 [2,173.94, 2,623.22] |
| | Private rented or living rent free | Total | 5,700,870 (100.0) | 36,870 (100.0) | 646.74 [640.16, 653.38] | 721.36 [713.50, 729.22] |
| | | Heterosexual | 5,289,315 (92.8) | 30,535 (82.8) | 577.30 [570.84, 583.81] | 637.42 [629.72, 645.11] |
| | | LGB+ | 411,550 (7.2) | 6,335 (17.2) | 1,539.30 [1,501.63, 1,577.68] | 1,493.34 [1,452.96, 1,533.72] |
| | | Gay or Lesbian | 162,125 (2.8) | 2,235 (6.1) | 1,378.57 [1,322.00, 1,436.93] | 1,531.87 [1,459.72, 1,604.03] |
| | | Bisexual | 194,775 (3.4) | 3,390 (9.2) | 1,740.47 [1,682.37, 1,800.07] | 1,555.05 [1,493.74, 1,616.36] |
| | | Other | 54,650 (1.0) | 710 (1.9) | 1,299.18 [1,205.36, 1,398.36] | 1,231.46 [1,131.99, 1,330.93] |
| | Living in a communal establishment | Total | 319,805 (100.0) | 3,465 (100.0) | 1,083.47 [1,047.69, 1,120.16] | 1,560.90 [1,478.29, 1,643.51] |
| | | Heterosexual | 283,355 (88.6) | 2,440 (70.4) | 861.11 [827.28, 895.97] | 1,390.50 [1,308.06, 1,472.94] |
| | | LGB+ | 36,445 (11.4) | 1,025 (29.6) | 2,812.46 [2,642.89, 2,990.04] | 3,135.22 [2,662.89, 3,607.55] |
| | | Gay or Lesbian | 9,215 (2.9) | 225 (6.5) | 2,441.67 [2,133.03, 2,782.42] | 2,481.32 [1,928.66, 3,033.98] |
| | | Bisexual | 23,020 (7.2) | 695 (20.1) | 3,019.11 [2,798.80, 3,252.16] | 4,730.07 [3,639.57, 5,820.57] |
| | | Other | 4,210 (1.3) | 105 (3.0) | 2,494.06 [2,039.90, 3,019.21] | 1,361.57 [868.38, 1,854.75] |
| Economic activity | Economically active | Total | 18,575,200 (100.0) | 69,875 (100.0) | 376.17 [373.39, 378.97] | 547.81 [542.82, 552.79] |
| | | Heterosexual | 17,769,215 (95.7) | 61,235 (87.6) | 344.61 [341.89, 347.35] | 491.75 [486.88, 496.63] |
| | | LGB+ | 805,985 (4.3) | 8,640 (12.4) | 1,071.98 [1,049.49, 1,094.83] | 1,188.31 [1,161.27, 1,215.34] |
| | | Gay or Lesbian | 406,400 (2.2) | 3,420 (4.9) | 841.54 [813.57, 870.22] | 1,136.95 [1,092.31, 1,181.59] |
| | | Bisexual | 317,585 (1.7) | 4,360 (6.2) | 1,372.86 [1,332.41, 1,414.23] | 1,273.63 [1,230.54, 1,316.71] |
| | | Other | 81,995 (0.4) | 860 (1.2) | 1,048.84 [979.91, 1,121.35] | 1,115.52 [1,035.50, 1,195.55] |

| | | | | | | |
|---|---|---|---|---|---|---|
| | Economically inactive | Total | 10,084,270 (100.0) | 66,495 (100.0) | 659.39 [654.39, 664.42] | 1,326.31 [1,313.79, 1,338.82] |
| | | Heterosexual | 9,784,925 (97.0) | 58,390 (87.8) | 596.73 [591.90, 601.59] | 1,190.57 [1,178.27, 1,202.87] |
| | | LGB+ | 299,340 (3.0) | 8,105 (12.2) | 2,707.62 [2,648.99, 2,767.22] | 2,981.35 [2,909.44, 3,053.26] |
| | | Gay or Lesbian | 118,185 (1.2) | 2,800 (4.2) | 2,369.17 [2,282.22, 2,458.58] | 3,371.40 [3,234.06, 3,508.74] |
| | | Bisexual | 139,935 (1.4) | 4,410 (6.6) | 3,151.46 [3,059.13, 3,245.88] | 3,121.99 [3,014.45, 3,229.52] |
| | | Other | 41,225 (0.4) | 895 (1.3) | 2,171.01 [2,031.09, 2,318.03] | 2,083.83 [1,936.75, 2,230.92] |
| General health (detailed) | Very good health | Total | 11,919,820 (100.0) | 33,790 (100.0) | 283.48 [280.46, 286.52] | 360.08 [356.00, 364.15] |
| | | Heterosexual | 11,498,565 (96.5) | 30,665 (90.8) | 266.69 [263.71, 269.69] | 339.28 [335.21, 343.34] |
| | | LGB+ | 421,255 (3.5) | 3,125 (9.2) | 741.83 [716.05, 768.31] | 712.32 [687.23, 737.41] |
| | | Gay or Lesbian | 224,690 (1.9) | 1,385 (4.1) | 616.40 [584.37, 649.74] | 748.65 [707.62, 789.69] |
| | | Bisexual | 156,015 (1.3) | 1,505 (4.5) | 964.65 [916.52, 1,014.65] | 752.69 [711.14, 794.25] |
| | | Other | 40,545 (0.3) | 235 (0.7) | 579.60 [507.86, 658.64] | 531.61 [461.41, 601.82] |
| | Good health | Total | 11,249,390 (100.0) | 47,100 (100.0) | 418.69 [414.92, 422.49] | 721.66 [713.89, 729.44] |
| | | Heterosexual | 10,803,965 (96.0) | 41,165 (87.4) | 381.02 [377.35, 384.72] | 653.07 [645.28, 660.87] |
| | | LGB+ | 445,425 (4.0) | 5,935 (12.6) | 1,332.44 [1,298.75, 1,366.77] | 1,303.20 [1,270.02, 1,336.39] |
| | | Gay or Lesbian | 202,910 (1.8) | 2,160 (4.6) | 1,064.51 [1,020.09, 1,110.37] | 1,361.52 [1,301.42, 1,421.62] |
| | | Bisexual | 193,105 (1.7) | 3,215 (6.8) | 1,664.90 [1,607.84, 1,723.46] | 1,354.28 [1,304.89, 1,403.66] |
| | | Other | 49,415 (0.4) | 555 (1.2) | 1,123.14 [1,031.63, 1,220.59] | 1,007.53 [922.89, 1,092.16] |
| | Fair health | Total | 4,008,345 (100.0) | 33,030 (100.0) | 824.03 [815.17, 832.97] | 1,912.87 [1,885.17, 1,940.56] |
| | | Heterosexual | 3,840,595 (95.8) | 28,315 (85.7) | 737.26 [728.69, 745.89] | 1,702.00 [1,673.29, 1,730.71] |
| | | LGB+ | 167,750 (4.2) | 4,715 (14.3) | 2,810.73 [2,731.07, 2,892.13] | 2,997.48 [2,911.56, 3,083.39] |
| | | Gay or Lesbian | 67,995 (1.7) | 1,610 (4.9) | 2,367.82 [2,253.56, 2,486.37] | 3,380.83 [3,204.61, 3,557.05] |
| | | Bisexual | 77,840 (1.9) | 2,570 (7.8) | 3,301.64 [3,175.22, 3,431.81] | 3,047.88 [2,927.80, 3,167.95] |
| | | Other | 21,915 (0.5) | 535 (1.6) | 2,441.25 [2,238.74, 2,657.16] | 2,323.27 [2,125.16, 2,521.37] |
| | Bad health | Total | 1,175,695 (100.0) | 16,965 (100.0) | 1,442.98 [1,421.34, 1,464.86] | 3,394.35 [3,314.21, 3,474.50] |
| | | Heterosexual | 1,119,055 (95.2) | 14,690 (86.6) | 1,312.71 [1,291.57, 1,334.12] | 3,053.44 [2,967.71, 3,139.17] |
| | | LGB+ | 56,640 (4.8) | 2,275 (13.4) | 4,016.60 [3,853.22, 4,185.11] | 4,804.07 [4,596.01, 5,012.13] |
| | | Gay or Lesbian | 22,750 (1.9) | 795 (4.7) | 3,494.51 [3,255.78, 3,746.10] | 5,266.12 [4,836.77, 5,695.47] |
| | | Bisexual | 25,130 (2.1) | 1,155 (6.8) | 4,596.10 [4,334.83, 4,869.00] | 4,890.15 [4,602.45, 5,177.85] |
| | | Other | 8,760 (0.7) | 325 (1.9) | 3,710.05 [3,317.61, 4,136.14] | 4,009.22 [3,561.40, 4,457.05] |
| | Very bad health | Total | 306,215 (100.0) | 5,485 (100.0) | 1,791.23 [1,744.13, 1,839.27] | 4,340.40 [4,143.83, 4,536.98] |
| | | Heterosexual | 291,960 (95.3) | 4,790 (87.3) | 1,640.64 [1,594.50, 1,687.77] | 3,841.32 [3,639.64, 4,043.01] |
| | | LGB+ | 14,255 (4.7) | 700 (12.8) | 4,910.56 [4,553.47, 5,288.20] | 6,708.01 [6,125.82, 7,290.20] |
| | | Gay or Lesbian | 6,240 (2.0) | 270 (4.9) | 4,326.92 [3,826.15, 4,875.02] | 7,471.47 [6,278.30, 8,664.63] |
| | | Bisexual | 5,425 (1.8) | 320 (5.8) | 5,898.62 [5,269.97, 6,581.62] | 7,018.61 [6,173.21, 7,864.01] |
| | | Other | 2,585 (0.8) | 105 (1.9) | 4,061.90 [3,322.23, 4,917.17] | 5,170.46 [4,087.42, 6,253.51] |
| General health (aggregated) | Good health | Total | 23,169,210 (100.0) | 80,885 (100.0) | 349.11 [346.70, 351.52] | 495.94 [492.13, 499.74] |
| | | Heterosexual | 22,302,530 (96.3) | 71,830 (88.8) | 322.07 [319.72, 324.44] | 454.99 [451.22, 458.75] |
| | | LGB+ | 866,680 (3.7) | 9,055 (11.2) | 1,044.79 [1,023.38, 1,066.54] | 1,012.75 [991.87, 1,033.63] |
| | | Gay or Lesbian | 427,600 (1.8) | 3,545 (4.4) | 829.05 [801.98, 856.80] | 1,025.06 [989.86, 1,060.26] |
| | | Bisexual | 349,125 (1.5) | 4,720 (5.8) | 1,351.95 [1,313.65, 1,391.08] | 1,081.81 [1,048.87, 1,114.75] |
| | | Other | 89,960 (0.4) | 790 (1.0) | 878.17 [817.99, 941.60] | 792.62 [736.50, 848.73] |
| | Not good health | Total | 5,490,255 (100.0) | 55,480 (100.0) | 1,010.52 [1,002.13, 1,018.96] | 2,276.87 [2,250.23, 2,303.50] |
| | | Heterosexual | 5,251,610 (95.7) | 47,790 (86.1) | 910.01 [901.87, 918.20] | 2,024.22 [1,996.55, 2,051.88] |
| | | LGB+ | 238,645 (4.3) | 7,690 (13.9) | 3,222.36 [3,150.74, 3,295.20] | 3,568.76 [3,488.09, 3,649.43] |

| | | | | | | |
|---|---|---|---|---|---|---|
| | | Gay or Lesbian | 96,985 (1.8) | 2,675 (4.8) | 2,758.16 [2,654.62, 2,864.70] | 3,969.38 [3,804.15, 4,134.61] |
| | | Bisexual | 108,395 (2.0) | 4,045 (7.3) | 3,731.72 [3,617.60, 3,848.53] | 3,625.64 [3,513.22, 3,738.05] |
| | | Other | 33,260 (0.6) | 965 (1.7) | 2,901.38 [2,721.19, 3,090.37] | 2,917.44 [2,733.15, 3,101.72] |
| Disability | Day-to-day activities limited a lot | Total | 1,976,155 (100.0) | 31,535 (100.0) | 1,595.78 [1,578.21, 1,613.49] | 3,527.82 [3,475.96, 3,579.68] |
| | | Heterosexual | 1,876,155 (94.9) | 26,620 (84.4) | 1,418.86 [1,401.87, 1,436.01] | 3,124.66 [3,070.62, 3,178.70] |
| | | LGB+ | 100,005 (5.1) | 4,915 (15.6) | 4,914.75 [4,778.30, 5,054.11] | 5,352.52 [5,201.44, 5,503.60] |
| | | Gay or Lesbian | 40,150 (2.0) | 1,695 (5.4) | 4,221.67 [4,023.06, 4,427.54] | 5,765.07 [5,469.21, 6,060.93] |
| | | Bisexual | 44,795 (2.3) | 2,555 (8.1) | 5,703.76 [5,484.72, 5,929.31] | 5,504.47 [5,289.79, 5,719.15] |
| | | Other | 15,060 (0.8) | 660 (2.1) | 4,382.47 [4,054.46, 4,729.95] | 4,378.50 [4,044.51, 4,712.50] |
| | Day-to-day activities limited a little | Total | 3,226,275 (100.0) | 33,095 (100.0) | 1,025.80 [1,014.77, 1,036.91] | 1,880.85 [1,857.33, 1,904.37] |
| | | Heterosexual | 3,002,375 (93.1) | 27,170 (82.1) | 904.95 [894.22, 915.78] | 1,745.09 [1,719.45, 1,770.74] |
| | | LGB+ | 223,905 (6.9) | 5,925 (17.9) | 2,646.21 [2,579.26, 2,714.47] | 2,452.77 [2,389.05, 2,516.48] |
| | | Gay or Lesbian | 78,960 (2.4) | 1,920 (5.8) | 2,431.61 [2,324.05, 2,542.87] | 2,841.54 [2,712.73, 2,970.35] |
| | | Bisexual | 115,035 (3.6) | 3,375 (10.2) | 2,933.89 [2,835.73, 3,034.58] | 2,505.72 [2,412.75, 2,598.69] |
| | | Other | 29,905 (0.9) | 630 (1.9) | 2,106.67 [1,945.36, 2,277.79] | 1,799.41 [1,652.05, 1,946.77] |
| | Day-to-day activities not limited | Total | 23,457,035 (100.0) | 71,735 (100.0) | 305.81 [303.58, 308.06] | 426.38 [422.81, 429.95] |
| | | Heterosexual | 22,675,615 (96.7) | 65,835 (91.8) | 290.33 [288.12, 292.56] | 403.35 [399.78, 406.92] |
| | | LGB+ | 781,420 (3.3) | 5,905 (8.2) | 755.68 [736.52, 775.20] | 770.98 [751.29, 790.68] |
| | | Gay or Lesbian | 405,475 (1.7) | 2,605 (3.6) | 642.46 [618.02, 667.61] | 811.03 [777.89, 844.16] |
| | | Bisexual | 297,690 (1.3) | 2,835 (4.0) | 952.33 [917.60, 988.05] | 799.62 [768.88, 830.37] |
| | | Other | 78,255 (0.3) | 460 (0.6) | 587.82 [535.32, 644.08] | 569.24 [516.86, 621.62] |
| Residence type | Living in a private household | Total | 28,339,665 (100.0) | 132,905 (100.0) | 468.97 [466.45, 471.50] | 671.26 [667.01, 675.50] |
| | | Heterosexual | 27,270,785 (96.2) | 117,185 (88.2) | 429.71 [427.25, 432.18] | 602.65 [598.49, 606.80] |
| | | LGB+ | 1,068,880 (3.8) | 15,720 (11.8) | 1,470.70 [1,447.80, 1,493.87] | 1,521.57 [1,497.71, 1,545.44] |
| | | Gay or Lesbian | 515,370 (1.8) | 6,000 (4.5) | 1,164.21 [1,134.94, 1,194.05] | 1,474.64 [1,434.59, 1,514.70] |
| | | Bisexual | 434,500 (1.5) | 8,075 (6.1) | 1,858.46 [1,818.14, 1,899.44] | 1,675.26 [1,637.46, 1,713.06] |
| | | Other | 119,005 (0.4) | 1,645 (1.2) | 1,382.29 [1,316.30, 1,450.75] | 1,345.25 [1,279.81, 1,410.69] |
| | Living in a communal establishment | Total | 319,805 (100.0) | 3,465 (100.0) | 1,083.47 [1,047.69, 1,120.16] | 1,560.90 [1,478.29, 1,643.51] |
| | | Heterosexual | 283,355 (88.6) | 2,440 (70.4) | 861.11 [827.28, 895.97] | 1,390.50 [1,308.06, 1,472.94] |
| | | LGB+ | 36,445 (11.4) | 1,025 (29.6) | 2,812.46 [2,642.89, 2,990.04] | 3,135.22 [2,662.89, 3,607.55] |
| | | Gay or Lesbian | 9,215 (2.9) | 225 (6.5) | 2,441.67 [2,133.03, 2,782.42] | 2,481.32 [1,928.66, 3,033.98] |
| | | Bisexual | 23,020 (7.2) | 695 (20.1) | 3,019.11 [2,798.80, 3,252.16] | 4,730.07 [3,639.57, 5,820.57] |
| | | Other | 4,210 (1.3) | 105 (3.0) | 2,494.06 [2,039.90, 3,019.21] | 1,361.57 [868.38, 1,854.75] |

Notes: Age-standardised rates are expressed per 100,000 people and standardised to the age distribution of the observed study population identifying with an LGB+ orientation in Census 2021. Cells with the value '-' are not applicable for the given characteristic or category. All calculations are based on estimates rounded to the nearest multiple of five for disclosure control reasons. CI: confidence interval.

**Supplementary Table 18: Sensitivity analysis including in-patient admissions from HES and PEDW for self-harm with undetermined intent, ratios of age-specific and age-standardised rates of self-harm per 100,000 people**

| Characteristic | Category | Sexual orientation | Rate ratio denominator | Rate ratio for self-harm [95% CI] |
|---|---|---|---|---|
| Total | Total | LGB+ | Heterosexual | 2.51 [2.47, 2.55] |
| | | Gay or Lesbian | Heterosexual | 2.42 [2.35, 2.48] |
| | | Bisexual | Heterosexual | 2.78 [2.72, 2.84] |
| | | Other | Heterosexual | 2.23 [2.12, 2.34] |
| Age on Census Day (10-year bands) | 16 to 24 years | LGB+ | Heterosexual | 2.77 [2.70, 2.84] |
| | | Gay or Lesbian | Heterosexual | 2.79 [2.67, 2.91] |
| | | Bisexual | Heterosexual | 2.82 [2.73, 2.90] |
| | | Other | Heterosexual | 2.48 [2.31, 2.65] |
| | 25 to 34 years | LGB+ | Heterosexual | 2.51 [2.43, 2.59] |
| | | Gay or Lesbian | Heterosexual | 2.35 [2.24, 2.46] |
| | | Bisexual | Heterosexual | 2.75 [2.64, 2.87] |
| | | Other | Heterosexual | 2.21 [2.02, 2.40] |
| | 35 to 44 years | LGB+ | Heterosexual | 2.22 [2.12, 2.32] |
| | | Gay or Lesbian | Heterosexual | 1.97 [1.84, 2.09] |
| | | Bisexual | Heterosexual | 2.76 [2.57, 2.94] |
| | | Other | Heterosexual | 1.80 [1.55, 2.06] |
| | 45 to 54 years | LGB+ | Heterosexual | 2.06 [1.94, 2.18] |
| | | Gay or Lesbian | Heterosexual | 1.69 [1.56, 1.83] |
| | | Bisexual | Heterosexual | 2.99 [2.71, 3.27] |
| | | Other | Heterosexual | 1.97 [1.59, 2.36] |
| | 55 to 64 years | LGB+ | Heterosexual | 1.92 [1.74, 2.10] |
| | | Gay or Lesbian | Heterosexual | 1.65 [1.46, 1.85] |
| | | Bisexual | Heterosexual | 2.70 [2.26, 3.15] |
| | | Other | Heterosexual | 2.01 [1.34, 2.69] |
| | 65 years and over | LGB+ | Heterosexual | 1.57 [1.33, 1.82] |
| | | Gay or Lesbian | Heterosexual | 1.45 [1.16, 1.73] |
| | | Bisexual | Heterosexual | 1.82 [1.29, 2.35] |
| | | Other | Heterosexual | 1.28 [0.49, 2.07] |
| Sex | Female | LGB+ | Heterosexual | 2.75 [2.69, 2.80] |
| | | Gay or Lesbian | Heterosexual | 3.03 [2.93, 3.14] |
| | | Bisexual | Heterosexual | 2.83 [2.76, 2.91] |
| | | Other | Heterosexual | 2.35 [2.22, 2.48] |
| | Male | LGB+ | Heterosexual | 1.90 [1.84, 1.96] |
| | | Gay or Lesbian | Heterosexual | 1.89 [1.81, 1.97] |
| | | Bisexual | Heterosexual | 2.08 [1.97, 2.19] |
| | | Other | Heterosexual | 1.71 [1.54, 1.87] |
| Region | England: North East | LGB+ | Heterosexual | 2.22 [2.07, 2.38] |
| | | Gay or Lesbian | Heterosexual | 2.32 [2.09, 2.55] |
| | | Bisexual | Heterosexual | 2.27 [2.03, 2.52] |
| | | Other | Heterosexual | 2.33 [1.83, 2.82] |
| | England: North West | LGB+ | Heterosexual | 2.74 [2.62, 2.87] |
| | | Gay or Lesbian | Heterosexual | 2.71 [2.52, 2.89] |
| | | Bisexual | Heterosexual | 3.05 [2.85, 3.25] |
| | | Other | Heterosexual | 2.16 [1.83, 2.48] |
| | England: Yorkshire and The Humber | LGB+ | Heterosexual | 2.16 [2.06, 2.27] |
| | | Gay or Lesbian | Heterosexual | 2.12 [1.96, 2.29] |
| | | Bisexual | Heterosexual | 2.34 [2.17, 2.51] |
| | | Other | Heterosexual | 1.92 [1.64, 2.20] |
| | England: East Midlands | LGB+ | Heterosexual | 2.62 [2.47, 2.76] |
| | | Gay or Lesbian | Heterosexual | 2.54 [2.31, 2.77] |
| | | Bisexual | Heterosexual | 2.91 [2.69, 3.12] |
| | | Other | Heterosexual | 2.15 [1.80, 2.51] |
| | England: West Midlands | LGB+ | Heterosexual | 2.86 [2.68, 3.03] |
| | | Gay or Lesbian | Heterosexual | 2.85 [2.58, 3.13] |
| | | Bisexual | Heterosexual | 3.14 [2.87, 3.40] |
| | | Other | Heterosexual | 2.17 [1.76, 2.58] |
| | England: East of England | LGB+ | Heterosexual | 2.68 [2.55, 2.83] |
| | | Gay or Lesbian | Heterosexual | 2.49 [2.27, 2.70] |
| | | Bisexual | Heterosexual | 3.01 [2.80, 3.22] |
| | | Other | Heterosexual | 2.44 [2.08, 2.79] |
| | England: London | LGB+ | Heterosexual | 2.29 [2.18, 2.41] |
| | | Gay or Lesbian | Heterosexual | 2.09 [1.90, 2.28] |
| | | Bisexual | Heterosexual | 2.57 [2.39, 2.74] |
| | | Other | Heterosexual | 2.36 [2.07, 2.65] |
| | England: South East | LGB+ | Heterosexual | 2.64 [2.53, 2.75] |

| | | | | |
|---|---|---|---|---|
| | | Gay or Lesbian | Heterosexual | 2.40 [2.23, 2.57] |
| | | Bisexual | Heterosexual | 2.95 [2.79, 3.11] |
| | | Other | Heterosexual | 2.43 [2.15, 2.71] |
| | England: South West | LGB+ | Heterosexual | 2.71 [2.58, 2.85] |
| | | Gay or Lesbian | Heterosexual | 2.75 [2.53, 2.97] |
| | | Bisexual | Heterosexual | 2.87 [2.68, 3.07] |
| | | Other | Heterosexual | 2.55 [2.20, 2.90] |
| | Wales | LGB+ | Heterosexual | 2.25 [2.06, 2.45] |
| | | Gay or Lesbian | Heterosexual | 2.38 [2.06, 2.69] |
| | | Bisexual | Heterosexual | 2.49 [2.18, 2.81] |
| | | Other | Heterosexual | 1.37 [0.95, 1.81] |
| Ethnic group | Asian, Asian British or Asian Welsh | LGB+ | Heterosexual | 2.25 [2.02, 2.48] |
| | | Gay or Lesbian | Heterosexual | 2.61 [2.11, 3.10] |
| | | Bisexual | Heterosexual | 2.12 [1.83, 2.41] |
| | | Other | Heterosexual | 2.26 [1.70, 2.81] |
| | Black, Black British, Black Welsh, Caribbean or African | LGB+ | Heterosexual | 2.92 [2.58, 3.28] |
| | | Gay or Lesbian | Heterosexual | 3.08 [2.37, 3.79] |
| | | Bisexual | Heterosexual | 2.97 [2.49, 3.45] |
| | | Other | Heterosexual | 2.80 [2.07, 3.54] |
| | Mixed or Multiple ethnic groups | LGB+ | Heterosexual | 2.32 [2.14, 2.50] |
| | | Gay or Lesbian | Heterosexual | 2.15 [1.88, 2.42] |
| | | Bisexual | Heterosexual | 2.55 [2.28, 2.82] |
| | | Other | Heterosexual | 2.01 [1.60, 2.42] |
| | White | LGB+ | Heterosexual | 2.38 [2.34, 2.42] |
| | | Gay or Lesbian | Heterosexual | 2.25 [2.18, 2.31] |
| | | Bisexual | Heterosexual | 2.68 [2.62, 2.75] |
| | | Other | Heterosexual | 2.22 [2.10, 2.33] |
| | Other ethnic group | LGB+ | Heterosexual | 2.16 [1.81, 2.52] |
| | | Gay or Lesbian | Heterosexual | 2.65 [1.81, 3.49] |
| | | Bisexual | Heterosexual | 2.30 [1.78, 2.84] |
| | | Other | Heterosexual | 1.60 [1.09, 2.13] |
| Rural/urban classification | Urban | LGB+ | Heterosexual | 2.47 [2.42, 2.51] |
| | | Gay or Lesbian | Heterosexual | 2.38 [2.31, 2.45] |
| | | Bisexual | Heterosexual | 2.72 [2.66, 2.79] |
| | | Other | Heterosexual | 2.19 [2.08, 2.30] |
| | Rural | LGB+ | Heterosexual | 2.84 [2.70, 2.98] |
| | | Gay or Lesbian | Heterosexual | 2.67 [2.46, 2.88] |
| | | Bisexual | Heterosexual | 3.19 [2.99, 3.40] |
| | | Other | Heterosexual | 2.58 [2.21, 2.95] |
| Country of birth | Born in the UK | LGB+ | Heterosexual | 2.48 [2.44, 2.53] |
| | | Gay or Lesbian | Heterosexual | 2.33 [2.27, 2.40] |
| | | Bisexual | Heterosexual | 2.79 [2.72, 2.86] |
| | | Other | Heterosexual | 2.39 [2.27, 2.52] |
| | Born outside the UK | LGB+ | Heterosexual | 2.35 [2.22, 2.48] |
| | | Gay or Lesbian | Heterosexual | 2.24 [2.00, 2.48] |
| | | Bisexual | Heterosexual | 2.55 [2.37, 2.74] |
| | | Other | Heterosexual | 2.08 [1.80, 2.36] |
| English language proficiency | Main language is English | LGB+ | Heterosexual | 2.49 [2.44, 2.53] |
| | | Gay or Lesbian | Heterosexual | 2.35 [2.29, 2.41] |
| | | Bisexual | Heterosexual | 2.77 [2.71, 2.84] |
| | | Other | Heterosexual | 2.34 [2.22, 2.45] |
| | Main language is not English | LGB+ | Heterosexual | 2.04 [1.84, 2.24] |
| | | Gay or Lesbian | Heterosexual | 2.20 [1.75, 2.66] |
| | | Bisexual | Heterosexual | 2.09 [1.81, 2.36] |
| | | Other | Heterosexual | 1.90 [1.52, 2.28] |
| Religion | No religion | LGB+ | Heterosexual | 2.09 [2.04, 2.13] |
| | | Gay or Lesbian | Heterosexual | 1.98 [1.91, 2.04] |
| | | Bisexual | Heterosexual | 2.33 [2.27, 2.40] |
| | | Other | Heterosexual | 1.97 [1.85, 2.10] |
| | Christian | LGB+ | Heterosexual | 2.94 [2.84, 3.05] |
| | | Gay or Lesbian | Heterosexual | 2.84 [2.67, 3.01] |
| | | Bisexual | Heterosexual | 3.44 [3.27, 3.61] |
| | | Other | Heterosexual | 2.19 [1.93, 2.44] |
| | Buddhist | LGB+ | Heterosexual | 4.00 [3.31, 4.80] |
| | | Gay or Lesbian | Heterosexual | 3.16 [2.15, 4.25] |
| | | Bisexual | Heterosexual | 4.24 [3.32, 5.25] |
| | | Other | Heterosexual | 4.46 [2.89, 6.12] |
| | Hindu | LGB+ | Heterosexual | 2.38 [1.74, 3.04] |
| | | Gay or Lesbian | Heterosexual | 3.15 [1.23, 5.09] |
| | | Bisexual | Heterosexual | 2.41 [1.64, 3.21] |
| | | Other | Heterosexual | 1.81 [0.35, 3.28] |

| | | | | |
|---|---|---|---|---|
| | Jewish | LGB+ | Heterosexual | 3.05 [2.33, 3.87] |
| | | Gay or Lesbian | Heterosexual | 2.53 [1.52, 3.63] |
| | | Bisexual | Heterosexual | 3.64 [2.55, 4.87] |
| | | Other | Heterosexual | 2.52 [0.87, 4.25] |
| | Muslim | LGB+ | Heterosexual | 2.29 [1.94, 2.63] |
| | | Gay or Lesbian | Heterosexual | 3.40 [2.31, 4.51] |
| | | Bisexual | Heterosexual | 2.40 [1.92, 2.89] |
| | | Other | Heterosexual | 1.65 [1.14, 2.17] |
| | Sikh | LGB+ | Heterosexual | 2.17 [1.38, 2.99] |
| | | Gay or Lesbian | Heterosexual | 4.13 [1.55, 6.77] |
| | | Bisexual | Heterosexual | 1.33 [0.54, 2.14] |
| | | Other | Heterosexual | 2.63 [0.77, 4.53] |
| | Other religion | LGB+ | Heterosexual | 2.79 [2.51, 3.12] |
| | | Gay or Lesbian | Heterosexual | 2.95 [2.45, 3.49] |
| | | Bisexual | Heterosexual | 2.82 [2.48, 3.20] |
| | | Other | Heterosexual | 2.63 [2.21, 3.08] |
| | Not answered | LGB+ | Heterosexual | 2.74 [2.47, 3.01] |
| | | Gay or Lesbian | Heterosexual | 2.65 [2.22, 3.09] |
| | | Bisexual | Heterosexual | 3.12 [2.69, 3.55] |
| | | Other | Heterosexual | 2.23 [1.64, 2.83] |
| IMD/WIMD decile group | 1 (most deprived) | LGB+ | Heterosexual | 2.38 [2.28, 2.49] |
| | | Gay or Lesbian | Heterosexual | 2.48 [2.32, 2.65] |
| | | Bisexual | Heterosexual | 2.62 [2.46, 2.78] |
| | | Other | Heterosexual | 1.56 [1.34, 1.78] |
| | 2 | LGB+ | Heterosexual | 2.30 [2.19, 2.40] |
| | | Gay or Lesbian | Heterosexual | 2.32 [2.16, 2.49] |
| | | Bisexual | Heterosexual | 2.41 [2.26, 2.56] |
| | | Other | Heterosexual | 1.95 [1.70, 2.20] |
| | 3 | LGB+ | Heterosexual | 2.36 [2.25, 2.47] |
| | | Gay or Lesbian | Heterosexual | 2.43 [2.25, 2.61] |
| | | Bisexual | Heterosexual | 2.53 [2.36, 2.69] |
| | | Other | Heterosexual | 1.87 [1.61, 2.12] |
| | 4 | LGB+ | Heterosexual | 2.41 [2.30, 2.53] |
| | | Gay or Lesbian | Heterosexual | 2.42 [2.23, 2.61] |
| | | Bisexual | Heterosexual | 2.63 [2.45, 2.81] |
| | | Other | Heterosexual | 2.27 [1.97, 2.58] |
| | 5 | LGB+ | Heterosexual | 2.59 [2.46, 2.72] |
| | | Gay or Lesbian | Heterosexual | 2.45 [2.25, 2.66] |
| | | Bisexual | Heterosexual | 2.78 [2.58, 2.98] |
| | | Other | Heterosexual | 2.46 [2.11, 2.81] |
| | 6 | LGB+ | Heterosexual | 2.47 [2.34, 2.61] |
| | | Gay or Lesbian | Heterosexual | 2.18 [1.98, 2.38] |
| | | Bisexual | Heterosexual | 2.93 [2.72, 3.15] |
| | | Other | Heterosexual | 2.15 [1.81, 2.50] |
| | 7 | LGB+ | Heterosexual | 2.64 [2.49, 2.80] |
| | | Gay or Lesbian | Heterosexual | 2.23 [2.01, 2.46] |
| | | Bisexual | Heterosexual | 3.09 [2.85, 3.33] |
| | | Other | Heterosexual | 2.71 [2.27, 3.14] |
| | 8 | LGB+ | Heterosexual | 2.70 [2.54, 2.87] |
| | | Gay or Lesbian | Heterosexual | 2.44 [2.19, 2.68] |
| | | Bisexual | Heterosexual | 3.08 [2.83, 3.34] |
| | | Other | Heterosexual | 2.59 [2.13, 3.05] |
| | 9 | LGB+ | Heterosexual | 2.73 [2.56, 2.92] |
| | | Gay or Lesbian | Heterosexual | 2.59 [2.32, 2.87] |
| | | Bisexual | Heterosexual | 3.02 [2.75, 3.31] |
| | | Other | Heterosexual | 3.02 [2.49, 3.56] |
| | 10 (least deprived) | LGB+ | Heterosexual | 2.89 [2.68, 3.11] |
| | | Gay or Lesbian | Heterosexual | 2.43 [2.12, 2.74] |
| | | Bisexual | Heterosexual | 3.33 [3.00, 3.67] |
| | | Other | Heterosexual | 3.08 [2.47, 3.68] |
| Highest level of qualification (≥25 years) | No qualifications | LGB+ | Heterosexual | 2.00 [1.87, 2.14] |
| | | Gay or Lesbian | Heterosexual | 2.83 [2.56, 3.10] |
| | | Bisexual | Heterosexual | 2.24 [2.02, 2.46] |
| | | Other | Heterosexual | 0.69 [0.55, 0.83] |
| | Level 1 (one to four GCSE passes (grade A* to C or grade 4 and above) and any other GCSEs at other grades, or equivalent qualifications) | LGB+ | Heterosexual | 2.54 [2.36, 2.72] |
| | | Gay or Lesbian | Heterosexual | 2.35 [2.10, 2.60] |
| | | Bisexual | Heterosexual | 3.02 [2.73, 3.32] |
| | | Other | Heterosexual | 1.76 [1.31, 2.21] |
| | | LGB+ | Heterosexual | 2.59 [2.44, 2.74] |

| | | | | |
|---|---|---|---|---|
| | Level 2 (five or more GCSE passes (grade A* to C or grade 4 and above) or equivalent qualifications) | Gay or Lesbian | Heterosexual | 2.16 [1.98, 2.35] |
| | | Bisexual | Heterosexual | 3.24 [2.98, 3.49] |
| | | Other | Heterosexual | 2.27 [1.84, 2.70] |
| | Apprenticeship | LGB+ | Heterosexual | 2.22 [1.94, 2.51] |
| | | Gay or Lesbian | Heterosexual | 2.17 [1.76, 2.60] |
| | | Bisexual | Heterosexual | 2.74 [2.26, 3.23] |
| | | Other | Heterosexual | 1.17 [0.66, 1.68] |
| | Level 3 (two or more A Levels or equivalent qualifications) | LGB+ | Heterosexual | 3.00 [2.86, 3.14] |
| | | Gay or Lesbian | Heterosexual | 2.66 [2.48, 2.84] |
| | | Bisexual | Heterosexual | 3.52 [3.26, 3.78] |
| | | Other | Heterosexual | 3.38 [2.89, 3.87] |
| | Level 4 and above (Higher National Certificate, Higher National Diploma, Bachelor's degree, or post-graduate qualifications) | LGB+ | Heterosexual | 2.45 [2.35, 2.54] |
| | | Gay or Lesbian | Heterosexual | 2.15 [2.03, 2.27] |
| | | Bisexual | Heterosexual | 2.85 [2.68, 3.02] |
| | | Other | Heterosexual | 2.85 [2.54, 3.16] |
| | Other qualifications | LGB+ | Heterosexual | 2.11 [1.73, 2.50] |
| | | Gay or Lesbian | Heterosexual | 2.52 [1.88, 3.17] |
| | | Bisexual | Heterosexual | 2.21 [1.61, 2.83] |
| | | Other | Heterosexual | 1.13 [0.53, 1.74] |
| NS-SEC | Class 1 (Higher managerial, administrative and professional occupations) | LGB+ | Heterosexual | 3.58 [2.74, 4.47] |
| | | Gay or Lesbian | Heterosexual | 3.00 [1.72, 4.31] |
| | | Bisexual | Heterosexual | 4.16 [2.96, 5.40] |
| | | Other | Heterosexual | 3.48 [1.43, 5.59] |
| | Class 2 (Lower managerial, administrative and professional occupations) | LGB+ | Heterosexual | 2.94 [2.65, 3.23] |
| | | Gay or Lesbian | Heterosexual | 2.61 [2.16, 3.06] |
| | | Bisexual | Heterosexual | 3.34 [2.94, 3.75] |
| | | Other | Heterosexual | 2.85 [2.13, 3.57] |
| | Class 3 (Intermediate occupations) | LGB+ | Heterosexual | 2.85 [2.66, 3.05] |
| | | Gay or Lesbian | Heterosexual | 2.72 [2.42, 3.03] |
| | | Bisexual | Heterosexual | 3.19 [2.91, 3.47] |
| | | Other | Heterosexual | 2.17 [1.71, 2.64] |
| | Class 4 (Small employers and own account workers) | LGB+ | Heterosexual | 2.11 [1.80, 2.42] |
| | | Gay or Lesbian | Heterosexual | 2.07 [1.45, 2.69] |
| | | Bisexual | Heterosexual | 2.46 [2.00, 2.93] |
| | | Other | Heterosexual | 1.31 [0.88, 1.74] |
| | Class 5 (Lower supervisory and technical occupations) | LGB+ | Heterosexual | 2.86 [2.54, 3.20] |
| | | Gay or Lesbian | Heterosexual | 2.72 [2.25, 3.21] |
| | | Bisexual | Heterosexual | 3.13 [2.65, 3.61] |
| | | Other | Heterosexual | 2.67 [1.43, 3.91] |
| | Class 6 (Semi-routine occupations) | LGB+ | Heterosexual | 2.58 [2.47, 2.70] |
| | | Gay or Lesbian | Heterosexual | 2.52 [2.34, 2.70] |
| | | Bisexual | Heterosexual | 2.77 [2.61, 2.94] |
| | | Other | Heterosexual | 2.25 [1.93, 2.56] |
| | Class 7 (Routine occupations) | LGB+ | Heterosexual | 2.42 [2.30, 2.54] |
| | | Gay or Lesbian | Heterosexual | 2.47 [2.27, 2.67] |
| | | Bisexual | Heterosexual | 2.60 [2.43, 2.77] |
| | | Other | Heterosexual | 1.77 [1.50, 2.05] |
| | Class 8 (Never worked and long-term unemployed) | LGB+ | Heterosexual | 2.31 [2.21, 2.41] |
| | | Gay or Lesbian | Heterosexual | 2.81 [2.61, 3.00] |
| | | Bisexual | Heterosexual | 2.44 [2.30, 2.59] |
| | | Other | Heterosexual | 1.25 [1.08, 1.41] |
| | Full-time students | LGB+ | Heterosexual | 2.76 [2.47, 3.05] |
| | | Gay or Lesbian | Heterosexual | 2.91 [2.43, 3.40] |
| | | Bisexual | Heterosexual | 2.50 [2.23, 2.78] |
| | | Other | Heterosexual | 2.77 [2.13, 3.41] |
| Household NS-SEC | Class 1 (Higher managerial, administrative and professional occupations) | LGB+ | Heterosexual | 2.78 [2.65, 2.92] |
| | | Gay or Lesbian | Heterosexual | 2.55 [2.34, 2.76] |
| | | Bisexual | Heterosexual | 3.07 [2.87, 3.27] |
| | | Other | Heterosexual | 3.01 [2.64, 3.39] |
| | Class 2 (Lower managerial, administrative and professional occupations) | LGB+ | Heterosexual | 2.64 [2.54, 2.73] |
| | | Gay or Lesbian | Heterosexual | 2.47 [2.31, 2.62] |
| | | Bisexual | Heterosexual | 2.89 [2.74, 3.04] |
| | | Other | Heterosexual | 2.71 [2.43, 2.99] |
| | | LGB+ | Heterosexual | 2.76 [2.63, 2.90] |

| | | | | |
|---|---|---|---|---|
| | Class 3 (Intermediate occupations) | Gay or Lesbian | Heterosexual | 2.79 [2.59, 3.00] |
| | | Bisexual | Heterosexual | 2.87 [2.67, 3.07] |
| | | Other | Heterosexual | 2.37 [2.02, 2.72] |
| | Class 4 (Small employers and own account workers) | LGB+ | Heterosexual | 2.23 [2.07, 2.39] |
| | | Gay or Lesbian | Heterosexual | 2.41 [2.13, 2.70] |
| | | Bisexual | Heterosexual | 2.38 [2.15, 2.62] |
| | | Other | Heterosexual | 1.63 [1.32, 1.95] |
| | Class 5 (Lower supervisory and technical occupations) | LGB+ | Heterosexual | 2.59 [2.39, 2.79] |
| | | Gay or Lesbian | Heterosexual | 2.68 [2.36, 3.00] |
| | | Bisexual | Heterosexual | 2.86 [2.57, 3.16] |
| | | Other | Heterosexual | 1.44 [1.05, 1.84] |
| | Class 6 (Semi-routine occupations) | LGB+ | Heterosexual | 2.54 [2.42, 2.66] |
| | | Gay or Lesbian | Heterosexual | 2.70 [2.51, 2.89] |
| | | Bisexual | Heterosexual | 2.61 [2.45, 2.77] |
| | | Other | Heterosexual | 2.05 [1.77, 2.32] |
| | Class 7 (Routine occupations) | LGB+ | Heterosexual | 2.44 [2.30, 2.57] |
| | | Gay or Lesbian | Heterosexual | 2.57 [2.33, 2.80] |
| | | Bisexual | Heterosexual | 2.63 [2.44, 2.82] |
| | | Other | Heterosexual | 1.58 [1.33, 1.84] |
| | Class 8 (Never worked and long-term unemployed) | LGB+ | Heterosexual | 2.20 [2.08, 2.32] |
| | | Gay or Lesbian | Heterosexual | 2.74 [2.51, 2.98] |
| | | Bisexual | Heterosexual | 2.29 [2.13, 2.46] |
| | | Other | Heterosexual | 1.12 [0.93, 1.31] |
| | Full-time students | LGB+ | Heterosexual | 2.63 [2.21, 3.07] |
| | | Gay or Lesbian | Heterosexual | 2.88 [2.15, 3.62] |
| | | Bisexual | Heterosexual | 2.34 [1.95, 2.74] |
| | | Other | Heterosexual | 2.65 [1.51, 3.82] |
| | Living in a communal establishment | LGB+ | Heterosexual | 2.25 [1.90, 2.63] |
| | | Gay or Lesbian | Heterosexual | 1.78 [1.38, 2.20] |
| | | Bisexual | Heterosexual | 3.40 [2.60, 4.23] |
| | | Other | Heterosexual | 0.98 [0.62, 1.34] |
| Lone person household indicator | Lives alone | LGB+ | Heterosexual | 2.06 [1.97, 2.16] |
| | | Gay or Lesbian | Heterosexual | 2.10 [1.94, 2.27] |
| | | Bisexual | Heterosexual | 2.23 [2.10, 2.35] |
| | | Other | Heterosexual | 1.72 [1.50, 1.94] |
| | Does not live alone | LGB+ | Heterosexual | 2.40 [2.36, 2.45] |
| | | Gay or Lesbian | Heterosexual | 2.30 [2.22, 2.38] |
| | | Bisexual | Heterosexual | 2.66 [2.58, 2.73] |
| | | Other | Heterosexual | 2.10 [1.98, 2.23] |
| | Living in a communal establishment | LGB+ | Heterosexual | 2.25 [1.90, 2.63] |
| | | Gay or Lesbian | Heterosexual | 1.78 [1.38, 2.20] |
| | | Bisexual | Heterosexual | 3.40 [2.61, 4.23] |
| | | Other | Heterosexual | 0.98 [0.62, 1.35] |
| Household size | One person in household | LGB+ | Heterosexual | 2.06 [1.96, 2.16] |
| | | Gay or Lesbian | Heterosexual | 2.10 [1.94, 2.27] |
| | | Bisexual | Heterosexual | 2.23 [2.10, 2.35] |
| | | Other | Heterosexual | 1.72 [1.51, 1.94] |
| | Two people in household | LGB+ | Heterosexual | 2.08 [2.00, 2.16] |
| | | Gay or Lesbian | Heterosexual | 1.98 [1.85, 2.12] |
| | | Bisexual | Heterosexual | 2.35 [2.23, 2.46] |
| | | Other | Heterosexual | 2.10 [1.88, 2.33] |
| | Three to five people in household | LGB+ | Heterosexual | 2.49 [2.42, 2.57] |
| | | Gay or Lesbian | Heterosexual | 2.44 [2.32, 2.56] |
| | | Bisexual | Heterosexual | 2.70 [2.59, 2.81] |
| | | Other | Heterosexual | 1.99 [1.83, 2.14] |
| | Six or more people in household | LGB+ | Heterosexual | 2.49 [2.26, 2.72] |
| | | Gay or Lesbian | Heterosexual | 2.68 [2.23, 3.13] |
| | | Bisexual | Heterosexual | 2.71 [2.39, 3.03] |
| | | Other | Heterosexual | 1.78 [1.36, 2.20] |
| | Living in a communal establishment | LGB+ | Heterosexual | 2.25 [1.90, 2.63] |
| | | Gay or Lesbian | Heterosexual | 1.78 [1.38, 2.20] |
| | | Bisexual | Heterosexual | 3.40 [2.60, 4.23] |
| | | Other | Heterosexual | 0.98 [0.62, 1.34] |
| Household tenure | Owned or shared ownership | LGB+ | Heterosexual | 2.49 [2.41, 2.58] |
| | | Gay or Lesbian | Heterosexual | 2.33 [2.20, 2.46] |
| | | Bisexual | Heterosexual | 2.70 [2.58, 2.83] |
| | | Other | Heterosexual | 2.65 [2.40, 2.89] |
| | Social rented | LGB+ | Heterosexual | 2.61 [2.53, 2.69] |
| | | Gay or Lesbian | Heterosexual | 2.67 [2.54, 2.79] |
| | | Bisexual | Heterosexual | 2.79 [2.68, 2.91] |
| | | Other | Heterosexual | 1.91 [1.73, 2.09] |

| | | | | |
|---|---|---|---|---|
| | Private rented or living rent free | LGB+ | Heterosexual | 2.34 [2.27, 2.41] |
| | | Gay or Lesbian | Heterosexual | 2.40 [2.29, 2.52] |
| | | Bisexual | Heterosexual | 2.44 [2.34, 2.54] |
| | | Other | Heterosexual | 1.93 [1.77, 2.09] |
| | Living in a communal establishment | LGB+ | Heterosexual | 2.25 [1.90, 2.63] |
| | | Gay or Lesbian | Heterosexual | 1.78 [1.38, 2.21] |
| | | Bisexual | Heterosexual | 3.40 [2.60, 4.23] |
| | | Other | Heterosexual | 0.98 [0.62, 1.34] |
| Economic activity | Economically active | LGB+ | Heterosexual | 2.42 [2.36, 2.48] |
| | | Gay or Lesbian | Heterosexual | 2.31 [2.22, 2.41] |
| | | Bisexual | Heterosexual | 2.59 [2.50, 2.68] |
| | | Other | Heterosexual | 2.27 [2.10, 2.43] |
| | Economically inactive | LGB+ | Heterosexual | 2.50 [2.44, 2.57] |
| | | Gay or Lesbian | Heterosexual | 2.83 [2.71, 2.95] |
| | | Bisexual | Heterosexual | 2.62 [2.53, 2.72] |
| | | Other | Heterosexual | 1.75 [1.63, 1.88] |
| General health (detailed) | Very good health | LGB+ | Heterosexual | 2.10 [2.02, 2.18] |
| | | Gay or Lesbian | Heterosexual | 2.21 [2.08, 2.33] |
| | | Bisexual | Heterosexual | 2.22 [2.09, 2.34] |
| | | Other | Heterosexual | 1.57 [1.36, 1.77] |
| | Good health | LGB+ | Heterosexual | 2.00 [1.94, 2.05] |
| | | Gay or Lesbian | Heterosexual | 2.08 [1.99, 2.18] |
| | | Bisexual | Heterosexual | 2.07 [1.99, 2.15] |
| | | Other | Heterosexual | 1.54 [1.41, 1.67] |
| | Fair health | LGB+ | Heterosexual | 1.76 [1.70, 1.82] |
| | | Gay or Lesbian | Heterosexual | 1.99 [1.88, 2.10] |
| | | Bisexual | Heterosexual | 1.79 [1.71, 1.87] |
| | | Other | Heterosexual | 1.37 [1.25, 1.48] |
| | Bad health | LGB+ | Heterosexual | 1.57 [1.49, 1.66] |
| | | Gay or Lesbian | Heterosexual | 1.72 [1.58, 1.87] |
| | | Bisexual | Heterosexual | 1.60 [1.50, 1.71] |
| | | Other | Heterosexual | 1.31 [1.16, 1.47] |
| | Very bad health | LGB+ | Heterosexual | 1.75 [1.57, 1.93] |
| | | Gay or Lesbian | Heterosexual | 1.95 [1.62, 2.28] |
| | | Bisexual | Heterosexual | 1.83 [1.59, 2.07] |
| | | Other | Heterosexual | 1.35 [1.06, 1.64] |
| General health (aggregated) | Good health | LGB+ | Heterosexual | 2.23 [2.18, 2.28] |
| | | Gay or Lesbian | Heterosexual | 2.25 [2.17, 2.33] |
| | | Bisexual | Heterosexual | 2.38 [2.30, 2.45] |
| | | Other | Heterosexual | 1.74 [1.62, 1.87] |
| | Not good health | LGB+ | Heterosexual | 1.76 [1.72, 1.81] |
| | | Gay or Lesbian | Heterosexual | 1.96 [1.88, 2.05] |
| | | Bisexual | Heterosexual | 1.79 [1.73, 1.85] |
| | | Other | Heterosexual | 1.44 [1.35, 1.53] |
| Disability | Day-to-day activities limited a lot | LGB+ | Heterosexual | 1.71 [1.66, 1.77] |
| | | Gay or Lesbian | Heterosexual | 1.85 [1.75, 1.95] |
| | | Bisexual | Heterosexual | 1.76 [1.69, 1.84] |
| | | Other | Heterosexual | 1.40 [1.29, 1.51] |
| | Day-to-day activities limited a little | LGB+ | Heterosexual | 1.41 [1.36, 1.45] |
| | | Gay or Lesbian | Heterosexual | 1.63 [1.55, 1.71] |
| | | Bisexual | Heterosexual | 1.44 [1.38, 1.49] |
| | | Other | Heterosexual | 1.03 [0.94, 1.12] |
| | Day-to-day activities not limited | LGB+ | Heterosexual | 1.91 [1.86, 1.96] |
| | | Gay or Lesbian | Heterosexual | 2.01 [1.93, 2.09] |
| | | Bisexual | Heterosexual | 1.98 [1.90, 2.06] |
| | | Other | Heterosexual | 1.41 [1.28, 1.54] |
| Residence type | Living in a private household | LGB+ | Heterosexual | 2.52 [2.48, 2.57] |
| | | Gay or Lesbian | Heterosexual | 2.45 [2.38, 2.52] |
| | | Bisexual | Heterosexual | 2.78 [2.71, 2.85] |
| | | Other | Heterosexual | 2.23 [2.12, 2.34] |
| | Living in a communal establishment | LGB+ | Heterosexual | 2.25 [1.90, 2.63] |
| | | Gay or Lesbian | Heterosexual | 1.78 [1.38, 2.21] |
| | | Bisexual | Heterosexual | 3.40 [2.61, 4.23] |
| | | Other | Heterosexual | 0.98 [0.62, 1.34] |

Notes: Rate ratios were calculated using age-specific rates for the age on Census Day variable, and age-standardised rates for all other variables. Cells with the value 'c' have been suppressed for the purposes of statistical disclosure control. All calculations are based on estimates rounded to the nearest multiple of five for disclosure control reasons. CI: confidence interval.
